# Rapid incidence estimation from SARS-CoV-2 genomes reveals decreased case detection in Europe during summer 2020

**DOI:** 10.1101/2021.05.14.21257234

**Authors:** Maureen Rebecca Smith, Maria Trofimova, Ariane Weber, Yannick Duport, Denise Kühnert, Max von Kleist

## Abstract

In May 2021, over 160 million SARS-CoV-2 infections have been reported worldwide. Yet, the true amount of infections is unknown and believed to exceed the reported numbers by several fold, depending on national testing policies that can strongly affect the proportion of undetected cases. To overcome this testing bias and better assess SARS-CoV-2 transmission dynamics, we propose a genome-based computational pipeline, GInPipe, to reconstruct the SARS-CoV-2 incidence dynamics through time. After validating GInPipe against *in silico* generated outbreak data, as well as more complex phylodynamic analyses, we use the pipeline to reconstruct incidence histories in Denmark, Scotland, Switzerland, and Victoria (Australia) solely from viral sequence data.

The proposed method robustly reconstructs the different pandemic waves in the investigated countries and regions, does not require phylodynamic reconstruction, and can be directly applied to publicly deposited SARS-CoV-2 sequencing data sets. We observe differences in the relative magnitude of reconstructed versus reported incidences during times with sparse availability of diagnostic tests. Using the reconstructed incidence dynamics, we assess how testing policies may have affected the probability to diagnose and report infected individuals. We find that under-reporting was highest in mid 2020 in all analysed countries, coinciding with liberal testing policies at times of low test capacities.

Due to the increased use of real-time sequencing, it is envisaged that GInPipe can complement established surveillance tools to monitor the SARS-CoV-2 pandemic and evaluate testing policies. The method executes within minutes on very large data sets and is freely available as a fully automated pipeline from https://github.com/KleistLab/GInPipe.

## Introduction

As of May 2021, the global SARS-CoV-2 pandemic is still ongoing in most parts of the world, with 160 million reported cases worldwide. Novel vaccines of high efficacy have been developed within a year of the outbreak [2, 45]. At the time of writing, approximately 8.2% of the worlds population had already received at least one vaccination. However, distribution of vaccines is uneven and achieving global herd immunity may pose an extremely difficult, long-term task [60, 36]. At the same time, novel variants of concern (VOC) have emerged in high prevalence regions [6, 34], which may be able to reinfect individuals [21, 37] and escape vaccine elicited immune responses [33, 63, 44]. For example, Manaus, Brazil, witnessed a massive second wave of infections [49], despite the fact that approx. 80% had already experienced an infection at the onset of the second wave [6]. Because of the evolutionary versatility of SARS-CoV-2 and difficulties in global vaccine distribution, some experts expect that the virus may not be eliminated globally [43]. Even without adaptation to vaccines in the future, it has been postulated that SARS-CoV-2 may resurge [24, 48] and surveillance may have to be maintained into the mid 2020s to monitor virus spread and evolution [24].

Currently, the gold standard of SARS-CoV-2 surveillance is diagnostic testing via polymerase chain reaction (PCR) or antigen-based rapid diagnostic testing (RDT). Diagnostic test results currently define infection case reports, which are used to survey epidemiological dynamics and to define thresholds for travel bans and non-pharmaceutical measures. Inevitably, case reporting data is affected by test coverage, which changes when testing policies are adapted. While RDT enables point-of-care diagnosis and is less costly than PCR testing [13, 12], gathering and reporting of test results still requires a sophisticated infrastructure, which is difficult to establish and maintain in many developing countries [35]. Independent and complimentary sources of information, such as social media reports [31, 51] or waste water analysis [9, 42] have been used early on to complement our knowledge of the pandemic dynamics. In addition, many regions of the world sequence SARS-CoV-2 genomes to track virus evolution and the emergence of variants of concern. The gathered viral sequences are regularly provided to public databases, such as GISAID [14, 52]. We hypothesize that the genetic data alone holds information about the pandemic trajectory. More specifically, we presume that the speed at which SARS-CoV-2 evolves on the population level contains information about the number of individuals who are actively infected.

In the vast majority of cases, SARS-CoV-2 is transmitted within a very short period, only days after infection [30, 17]. The consequence is a well-defined duration of intra-patient evolutionary time before transmission. Thus, the number of infected individuals is correlated to the rate of divergence of the viral population, implicating an ‘evolutionary signal’.

In this article, we introduce the computational pipeline GInPipe, which only uses time-stamped sequencing data, extracts the ‘evolutionary signal’ and reconstructs SARS-CoV-2 incidence histories. The approach builds on recent work by Khatri and Burt [23], who derived a simple function that relates the mean number of mutant origins to the current allele frequency and the mutational input, which is proportional to the effective population size. Herein, due to the short window of transmission, we anticipate that the effective population size may strongly correlate with the incidence of SARS-CoV-2. We adapt the function derived in [23] and embed it into an automatic computational pipeline (GInPipe) that reconstructs the time course of an incidence correlate *ϕ* merely from SARS-CoV-2 genetic data. GInPipe is validated threefold and performs robustly: (i) against *in silico* generated outbreak data, (ii) against phylodynamic analysis and (iii) in comparison with case reporting data. We applied the method to SARS-CoV-2 sequencing data from Denmark, Scotland, Switzerland, and the Australian state Victoria to reconstruct their respective incidence histories. Lastly, we utilize the inferred epidemic trajectories to compute changes in the probability that an infected individual is reported and highlight how this probability is affected by changes in testing policies.

## Results

### Incidence reconstruction

An outline of GInPipe for SARS-CoV-2 incidence reconstruction is shown in Figure 1A-C. After compiling a set of time-stamped, full-length SARS-CoV-2 genomes, the sequences are placed into temporal bins *b* (Fig. 1A). For each bin, we compute the number of mutant sequences *m*_*b*_, as well as the number of haplotypes *h*_*b*_. These two inputs are used to infer the incidence correlate *ϕ*_*b*_ (Fig. 1B). We then smooth over all *ϕ*_*b*_ point estimates and derive a reconstructed incidence history along the time axis (Fig. 1C). The reconstructed incidence histories can then be used as a basis to estimate the effective reproduction number *R*_*e*_, as well as the relative case detection rate as outlined below.

**Figure 1.**
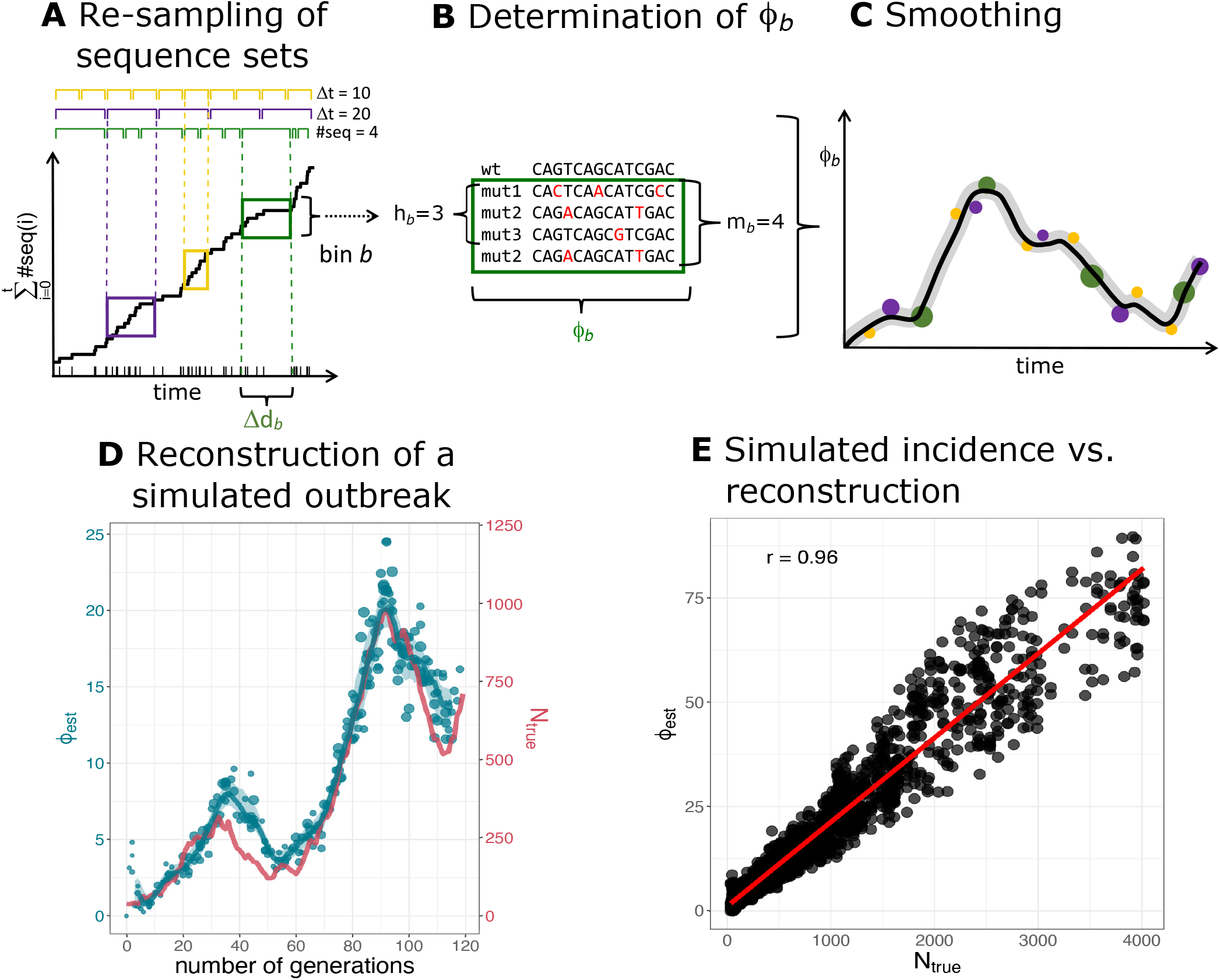
Reconstruction of incidence histories using the proposed method. **A–C** Schematic of the incidence reconstruction method. **A** The sequences are chronologically ordered by collection date. The line shows the cumulative sum of sequences over time. The sequences are allocated into temporal bins, spanning either the same time frame Δ*d*_*b*_ (yellow and purple bins) or containing the same amount of sequences (green bins). **B** For each bin, the number of distinct variants *h*_*b*_, as well as the total amount of mutant sequences *m*_*b*_ are used to infer the incidence correlate *ϕ*_*b*_. **C** The point estimates for all bins *ϕ*_*b*_ (dots) are smoothed with a convolution filter. For uncertainty estimation, the point estimates are sub-sampled and interpolated. **D–E** Reconstruction of a simulated outbreak with GInPipe. **D** *ϕ* estimates resemble the underlying population dynamics over time. The blue line shows the smoothed median of the sub-sampled *ϕ* estimates (dots) for a simulated outbreak. The red line indicates true incidence per generation. **E**. Dotplot showing the true outbreak size from the simulation *N*_true_ versus the *ϕ*_*b*_ point estimates for 10 stochastic simulations. The red line depicts the linear fit.

### Method validation: *in silico* experiment

To test whether GInPipe correctly reconstructs incidence histories, we first performed an *in silico* experiment. We considered a population of *N*(*t*) infected individuals at time *t* that stochastically generate *N*(*t* + 1) infected individuals in the next time step *t* + 1. Each individual is associated with a virus sequence, which can mutate randomly. Individuals can be removed (the associated sequence is removed), or they transmit their virus (the associated virus is copied over). We record the number of infected individuals per generation, as well as all sequences of the currently circulating viruses. We then use the simulated viral sequences to infer *ϕ*(*t*) and reconstruct the incidence history, as presented in Figure 1D-E.

In Figure 1D, we compare one trajectory of simulated population sizes with the reconstructed incidence histories. The simulated outbreak (red line, right axis) consists of two waves of increasing magnitude. GInPipe reconstructs these dynamics (blue lines and dots, left axis) quite accurately, although the incidence correlate *ϕ*(*t*) is on a different scale, implying a linear correlation to the number of infected individuals. To assess this correlation, we performed 10 stochastic simulations and compared the *ϕ*(*t*) point estimates with the corresponding number of infected individuals (Fig. 1E). We observed a strong (*r* = 0.96) and highly significant (*p* < 10^−16^) linear relationship between the number of infected individuals *N*(*t*) and the method’s incidence correlate *ϕ*(*t*).

While these simulations represent idealized scenarios, we evaluated the robustness of GInPipe with regards to incomplete, and sparse data sets, thoroughly elaborated in Supplementary Note 1.

Our analyses showed, that the method can still accurately reconstruct incidence histories over time, when data is missing or when data sampling is unbalanced. In scenarios of extreme under-sampling, the *ϕ* point estimates are prone to slight underestimation. However, through the smoothing step the reconstructed incidence trajectories still follow the overall population dynamics (Suppl. Note 1, section SN.1.7). Finally, we evaluated whether introductions of foreign sequences affect the reconstruction of incidence histories. Even for extreme and unrealistic cases, a stable reconstruction of the underlying dynamic is possible, but we do observe a slight tendency of overestimation in these extreme cases (Suppl. Note 1, section SN.1.8).

### Method validation: phylodynamics

Phylodynamic methods combine phylogeny reconstruction with epidemic models. For example, the piecewise constant birth-death sampling process [53] implemented in BEAST2 [5], allows the reconstruction of the effective reproduction numbers *R*_*e*_(*τ*) for given time periods *τ*. However, these methods are computationally expensive, so that only moderately sized sequence sets can be used, and advanced knowledge is required to apply them properly to larger data sets.

We conducted phylodynamic analyses of SARS-CoV-2 sequence data from Denmark, Scotland, Switzerland, and the Australian state Victoria. In analyzing the data we assumed that 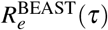 was piecewise constant in between major changes in SARS-CoV-2 non-pharmaceutical interventions (intervals stated in Supplementary Note 2). We then used BEAST2 to estimate 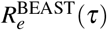 alongside the tree reconstructions.

In parallel, we estimated corresponding effective reproduction numbers 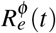 by applying the Wallinga-Teunis method [58] to incidence correlates *ϕ* derived by GInPipe. For both methods, we used publicly available full length SARS-CoV-2 sequencing data from GISAID [14, 52](Supplementary Note 4).

Results of both methods are shown in Figure 2. Overall, both methods show congruent trends for the analyzed countries, when comparing the piecewise constant 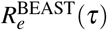 from phylodynamic analysis with the *median* daily 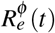 for the same interval. Noteworthy, GInPipe allows for a much finer time-resolution (daily *R*_*e*_ estimates) compared to the piecewise constant *R*_*e*_ estimates on pre-defined intervals, obtained from the phylodynamic analysis.

**Figure 2.**
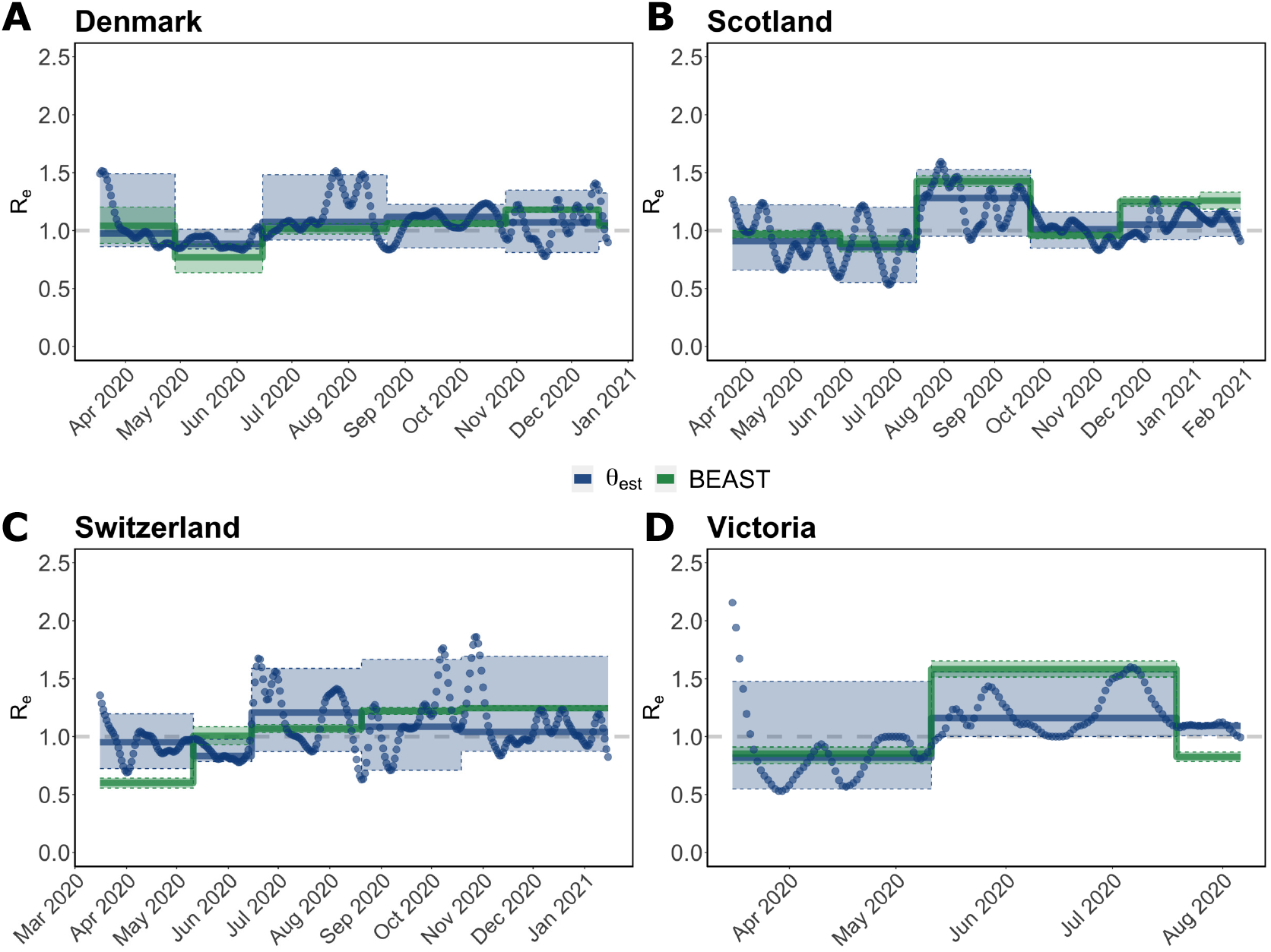
Effective reproduction number *R*_*e*_ estimates using the proposed method (*ϕ*) and phylodynamics (BEAST2). Piecewise constant 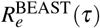 estimates (green solid lines) where calculated using the BDSKY model for the indicated intervals, as described in the Methods section. Daily estimates 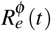 (blue dots) were directly calculated from the incidence correlates *ϕ* using the Wallinga-Teunis method [58]. The median of these values for the indicated intervals 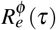 is shown as solid blue lines. The 95% confidence interval is specified by the shaded areas. Justifications of the intervals are found in Supplementary Note 2.

For Denmark, the first interval spans both sides of the peak number of infections during the first wave. Here, we see a median *R*_*e*_ (*τ*) ≈ 1 using both methods. The daily estimates from GInPipe (blue dots), additionally show the transition from 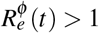 to 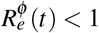 during this interval. After this, between May and mid June 2020 we have *R*_*e*_ (*τ*) < 1. For the next intervals, *R*_*e*_ (*τ*) is predicted to be slightly larger than one. However, GInPipe reconstructs a number of peaks in the daily 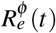 estimates, most pronounced in August, coinciding with the summer holidays in Europe. In the interval from November to mid December the estimates deviate slightly, with a larger median estimate from BEAST2, however, both interval estimates are predicted to be *R*_*e*_(*t*) > 1 and the confidence intervals overlap entirely.

The *R*_*e*_(*τ*) estimates for Scotland agree almost exactly, where GInPipe again allows for a much finer time-resolution. Once again, we see a peak in the summer (August-September 2020), coinciding with the summer holidays in Europe. For the last interval (from December 2020) both methods show a median *R*_*e*_(*t*) > 1, again with a slightly higher median BEAST2 estimate, coinciding with the second wave of infections.

For Switzerland, the estimates disagree slightly, particularly in the first interval (mid March to mid May), which spans both sides of the peak number of infections during the first wave. Although both methods predict a median *R*_*e*_(*τ*) < 1, the absolute value differs in magnitude between the two methods, with BEAST2 estimating a much lower value. The lower estimate from the BEAST2-analysis in the first interval may be explained by the approximation of transmission clusters, which results in the reconstruction of a relatively high number of transmission events many of which may have occurred outside Switzerland (Supplementary Note 2, Figure SN.12 therein, tree B.1). In the daily estimates, we see a transition from 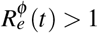 to 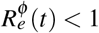, which may explain why the *median* prediction with GInPipe is close to one for the entire interval. The estimates are qualitatively different for the second interval (mid May – mid June), where GInPipe estimates 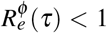, while BEAST2 estimates 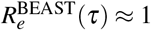. Again, GInPipe estimates a peak in summer (mid June-mid August *R*_*e*_*ϕ*(*τ*) > 1). While BEAST2 predicts the onset of transmission in the second wave to already start in mid August (*R*_*e*_(*τ*) > 1), GInPipe estimates the first major rise in infections at the end of September.

For Victoria we observe an 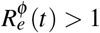 until mid March in the daily estimates. Overall, *R*_*e*_ is less than 1 for the first interval between mid March and May, versus *R*_*e*_ > 1 between June and August. Again, we see various peaks around June and July in the daily *R*_*e*_ estimates with the proposed method. For the final interval, both methods slightly disagree, with 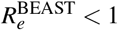 and 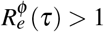, though the daily 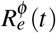 are decreasing towards the end of the final interval.

In terms of computational time, the entire GInPipe analysis pipeline runs in 20 minutes on the full Denmark data set (n = 40.575 sequences) and in 7 minutes on the Victoria data set (n = 10.710 sequences) on a single notebook (2,3 Ghz, 2 cores). Furthermore, GInPipe does not require to pre-assign any intervals, to exclude particular strains, construct a phylogenetic tree, or cluster sequences based on a their phylogenetic relationship. The BEAST2 analysis alone required about 10 hours on an Intel Xeon E5-2687W (3.1 Ghz, 2 × 12 cores) on a sub-sampled data set (*n* ≈ 2500 sequences) with additional computation time needed to construct a multiple sequence alignment and approximate transmission clusters.

### Reconstructed incidence histories

We used GInPipe to reconstruct complete incidence histories for Denmark, Scotland, Switzerland, and Victoria (Australia) from publicly available full length SARS-CoV-2 sequencing data provided through GISAID [14, 52] (Supplementary Note 4). In Figure 3, we compare the reconstructed incidence histories (blue lines and dots, left axis) to the 7-day rolling average of officially reported new cases (red line, right axis). Overall, the reconstructed incidence estimates reflect the different pandemic waves deduced from the reporting data, although there are quantitative differences between the reconstructed and reported incidence trajectories over time. In particular, during the first wave in Scotland, and Victoria (Fig. 3B,D) our method estimates higher incidences than reported, whereas the curves align at later points for the second and third wave. It is worth mentioning that testing capacities were particularly low in Scotland in April (during the first wave), suggesting extensive under-reporting in the initial phase of the pandemic. This is also supported by test positive rates of almost 40% during April 2020 in Scotland (Supplementary Fig. 1). In Victoria, sufficient testing capacities were not available until May, but test positive rates were already declining from April to May (Supplementary Fig. 1). This indicates that the first wave may have been under-reported in magnitude, but had vanished by May.

**Figure 3.**
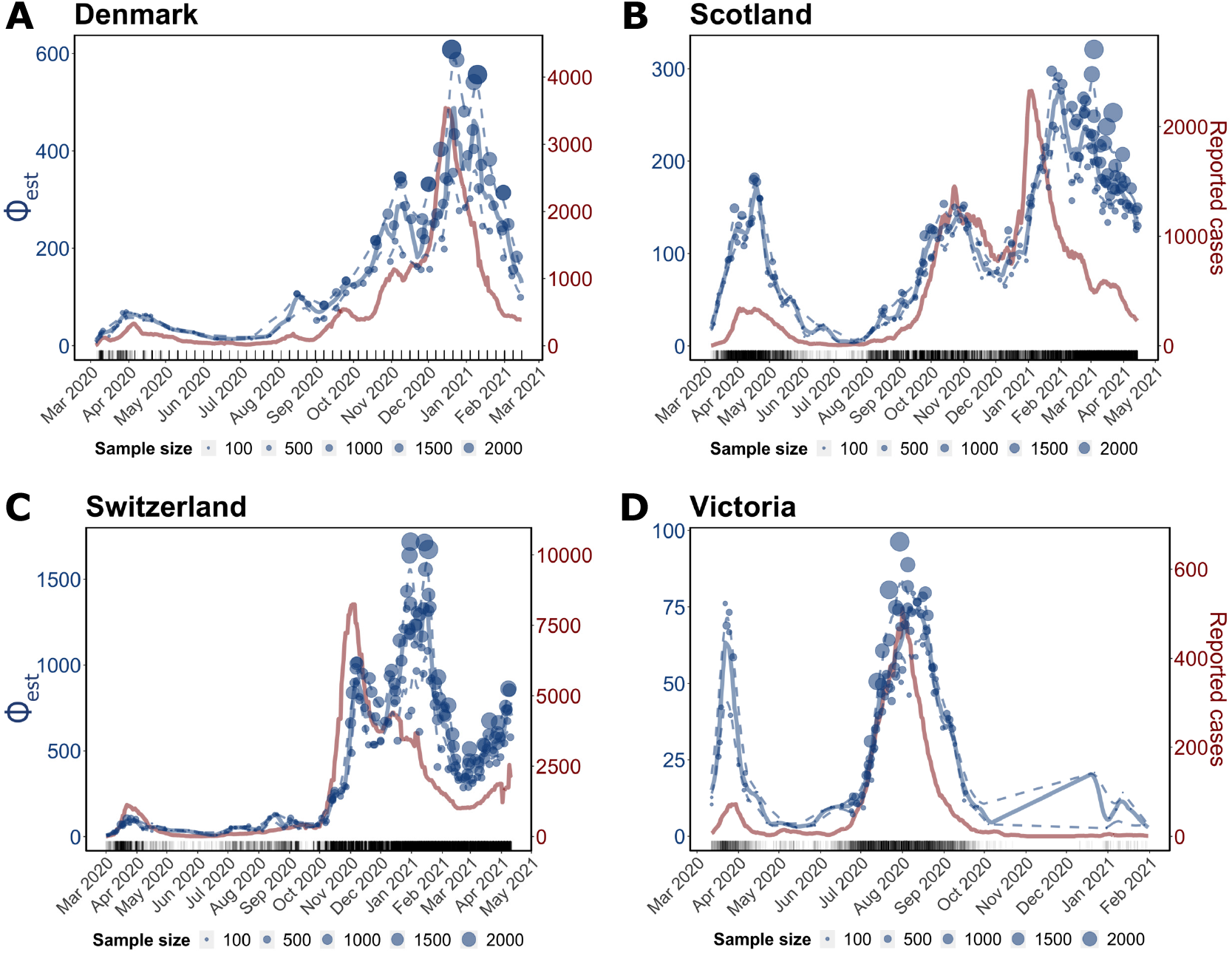
Incidence reconstruction based on sequencing data. The graphic depicts the genome-based incidence reconstruction (in blue) using the proposed method (left axis) vs. the 7 days rolling average of newly reported cases in red (right axis). Blue dots depict *ϕ*_*b*_ point estimates of the incidence correlate, where the size of the dot is related to the number of sequences used to infer *ϕ*_*b*_. The solid and dashed blue lines denote the median smoothed trajectories and their 5th and 95th percentiles. The black markers on the x-axis depict the collected sequences at the given dates. **A** Denmark (n = 40.575 sequences) **B** Scotland (n = 30.258 sequences) **C** Switzerland (n = 25.779 sequences) **D** Victoria (n = 10.710 sequences)

Interestingly, the proposed incidence reconstruction method predicts small summer waves in August in the three European countries (Fig. 3A–C) that are not visible in the reporting data. In the incidence reconstruction method these ‘summer waves’ are immediately followed by the second SARS-CoV-2 wave. For the second wave, reconstructed incidence histories correspond to the reported cases, particularly in Denmark, Scotland, and Victoria. (Fig. 3A-B & D). For Scotland, our method predicts a more long-lasting third wave with rising incidence rates until February 2021 and a moderate decline with several smaller peaks until May, whereas the reporting data indicates a peak in January 2021 with a subsequent fast regression. The argument, that ongoing vaccination in Great Britain could explain the immediate decline of reported infected cases, can be objected with the fact, that by March 2021 (end of the prediction horizon) only about 2% of the Scottish population were fully vaccinated.

For Switzerland, we predict a larger wave around January-February 2021 (third wave) that is not reflected in the reporting data. Towards the end of the prediction horizon, from March 2021 onwards, the reported cases and the incidence estimation both indicate a rise in numbers (fourth wave).

### Relative case detection rate

We investigated whether the proposed incidence reconstruction method may be used to learn about the proportion of infected cases that are actually tested, detected and reported, *P*_*t*_ (tested|infected).

The proportion of SARS-CoV-2 infected who are actually reported can be calculated using Bayes’ formula (see *Methods* section). In order to perform the calculation, the proportion of actively infected individuals in the population *P*_*t*_ (infected) needs to be known. We have shown that the incidence correlates *ϕ* from our method are proportional to the number of infected individuals, *c · ϕ*_*t*_ = *N*_eff_ (Fig. 1D–E, Fig. 3), and hence to the probability of being infected *P*_*t*_ (infected). Consequently, we may use the reconstructed incidence profiles, together with the test sensitivity and specificity, the respective information about the proportion of positive tests, as well as the testing capacities for each country or region to calculate changes in the case detection rate, scaled by unknown factor *c*.

In Figure 4, we show the log_2_ scaled detection probabilities for Denmark, Scotland, Switzerland, and Victoria (Australia). The log scaling allows us to easily gauge the relative change in (under-)detection of the infected population over time (e.g. 2-fold, 4-fold increase or decrease in case detection rate). The dashed vertical lines in the graphics indicate major changes in testing policies in the respective countries. Individual parameters used in the inference procedure, *P*(tested), *P*(inf |tested), and *c·P*(infected) are shown in Supplementary Figure 1.

**Figure 4.**
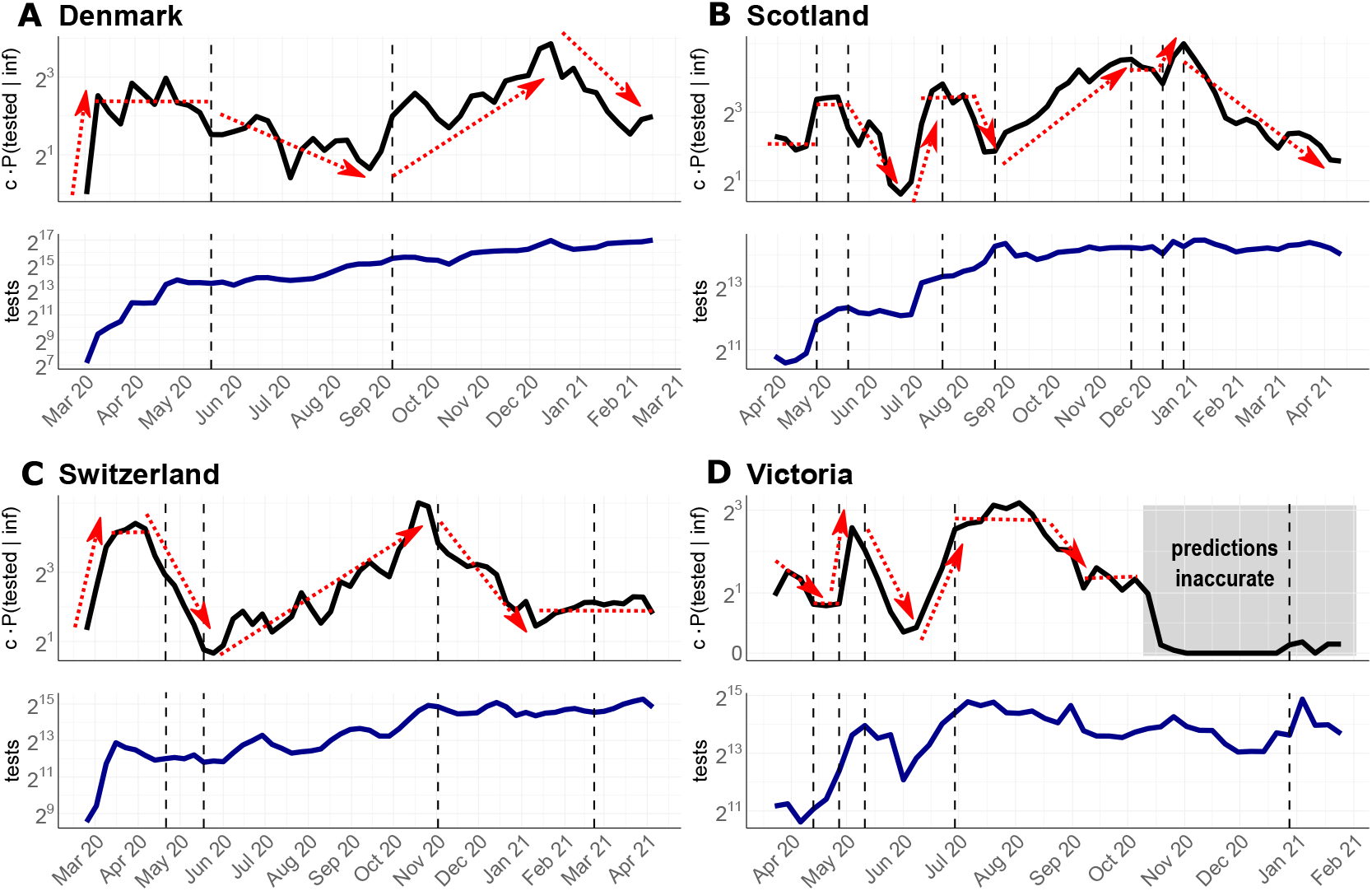
Relative case detection rate. Black line in upper graphics: Estimated and scaled probability of detecting SARS-CoV-2 infected individuals *c·P*(tested |inf). Blue line in the lower graphics: Number of conducted tests per calendar week. Dashed vertical lines indicate major changes in the testing strategies in the respective location. The sources for testing data and strategies are given in Supplementary Note 3. **A** Denmark. Policy changes: 18 May 20: testing for everyone; 9 September 20: increasing testing available **B** Scotland. Policy changes: 1 May 20: expanded testing strategy including enhanced outbreak investigation; 18 May 20: testing for everyone with symptoms; 22 July 20: including young children for testing; 25 August 20: increasing capacity and accessibility of testing; 25 November 20: expansion of testing in health care; 15 December 20: increase of testing capacity; 1 January 21: community testing in areas with high coronavirus prevalence. **C** Switzerland. Policy changes: 18 May 20: priority testing; 2 November 20: rapid antigen tests are included in the testing strategy; 27 February 21: recommended preventative and repeated testing as part of precautionary measures. **D** Victoria (Australia). Policy changes: 14 April 20: anyone having symptoms can be tested; 30 April 20: start of 2 weeks ’testing blitz’; 11 May 20: increased surveillance with testing of sewerage; 1 July 20: expanded ‘testing blitzes’ in outbreak regions; 30 December 21: urging to be tested after re-emergence of positive cases.

For Denmark, we observe an initial period of massive SARS-CoV-2 under-detection in the beginning of March 2020, Fig. 4A (upper panel), which coincides with very low testing capacities at the beginning of the pandemic (Fig. 4A, lower panel). From mid March, case detection stabilizes at a 6-fold higher level, compared to the first week of March. The second interval begins around mid May with an important policy change, allowing every citizen to get tested without medical referral. Interestingly, compared to the fairly stable case detection levels from mid March to mid May, this policy change leads to a 2-3 fold drop in case detection in the summer months from July-September. Of note, while everybody is granted the possibility to test for SARS-CoV-2, testing capacities remained fairly unchanged (Fig. 4A, lower panel). According to our calculations, the largest proportion of infections remained undetected in July. From end of August, testing capacities were steadily increased in Denmark (Fig. 4A, lower panel), particularly in Copenhagen and at the airports, followed by prioritized testing. From September on, this leads to a nearly 8-fold increase of the case detection rate, with a peak in December. From end of December the detection rate drops more than 4-fold, despite continuous testing.

For Scotland (Fig. 4B), the earliest test data is available only from the end of March. Therefore, the data captures only the second part of the first wave, compare Fig. 3B. In the beginning of May, testing capacities were more than doubled (Fig. 3B, lower panel) and outbreak investigation intensified. This led to a doubling of the relative case detection rate from May, compared to the first phase. On 18 May, SARS-CoV-2 testing was opened for everyone with symptoms. However, only in July testing capacities were increased. This may have led to a drop in case detection from mid May to July, after which case detection increased and remained during August at roughly the levels achieved in May. After 25 August, testing capacities and accessibility of testing steadily increased. Accordingly, case detection increased about 6-fold until winter 20/21. From 25 November, testing capacities were further expanded, especially in the health sector, including hospital patients, health and social care staff, with fairly stable case detection rates. Further increase of testing capacities in the end of December allowed to double the probability to detect infected individuals. From beginning of the year 2021, the Scottish government pushed community testing in areas with high SARS-CoV-2 prevalence. At the same time, the proportion of positive tests start to decline (Suppl. Fig. 1), and consequently the case detection rate collapses until April by 9-fold.

Similar to Denmark, Switzerland shows an initial period of massive SARS-CoV-2 under-detection in the beginning of March 2020 (Fig. 4C, upper panel), which coincides with very low testing capacities at the beginning of the pandemic (Fig. 4C, lower panel). When testing capacities increase by mid March, case detection rates grow 8-fold. However, from beginning of April, we observe drop in the probability to detect infections that lasts until mid May (overall 10-fold drop). This trend coincides with a drop of positivity rates (Supplementary Figure 1), as well as the extension of testing criteria on 22nd April: From this date, anybody with symptoms was allowed to get tested, despite the fact that the availability of tests was not increased (Fig. 4C, lower panel). From 18th May, tests were partly prioritized for hospitalized and vulnerable individuals. At the same time, testing capacities steadily increased and incidences dropped. As a net effect, the probability of detecting infected people increases steadily to a maximum at the end of October with a relative difference of nearly 20-fold compared to the low point in mid May. On 2 November, Switzerland begins to supply antigen-based rapid diagnostic tests (RDT) for self-testing as part of their COVID containment strategy. Interestingly, our model predicts that this led to a sharp decline in case detection, again corresponding with the decline in positivity rates (Supplementary Figure 1). From 21st February 2021, further precautionary actions were taken, and the government recommended repeated testing. This is associated with a stable, but relatively low detection rate for infected people until end of April 2021.

For the Australian state Victoria, the earliest data were available from end of March 2020, Fig. 4D, capturing the second part of the first SARS-CoV-2 wave. Detection probabilities in the first interval, until 14th April were changed proportionally to the test capacities during that interval Fig. 4D (upper and lower panel). On 14th April 2020, the testing criteria were expanded, allowing anyone with COVID-like symptoms to be tested. Unlike the situation in Switzerland, where we observed a downward trend in case detection after expanding the testing criteria (Fig. 4C), the detection probability in Victoria remains stable until end of April. In contrast to Switzerland, testing capacities were increased when testing criteria were expanded. On 30th April, the government initiated a two-week ‘testing blitz’, a large, coordinated testing campaign to locate viral spread. The ‘testing blitz’ was accompanied by mass sewerage testing and matched with a massive increase of testing capacities, which led, according to our simulations, to a 4-fold increase in the probability to detect infected individuals. At the end of the ‘testing blitz’, testing capacities steadily decreased and the proportion of detected infections decreased drastically (by roughly 9-fold). At the beginning of June, testing capacities rose again, matched by a rise in the proportion of detected cases. From 1st July onwards, several ‘testing blitzes’ were conducted in outbreak regions, which seemed to have stabilized case detection rates during the second wave of infections. After the second wave (end of August–September, Fig. 3D), case detection rates drop. From October 2020 onwards, our predictions become highly unreliable, as the incidence estimates credibility interval includes zero (compare Fig. 3D), which concludes that the case detection rate cannot be determined anymore.

In general, we make two striking observations: Firstly, and quite intuitively, whenever more tests were conducted, the proportion of detected SARS-CoV-2 cases increases. Secondly, and unexpectedly, whenever testing criteria were relaxed, this led to a drop in the probability of case detection. We see this drop in mid May in Denmark and Scotland and in mid April in Switzerland. Importantly, the expansions of testing criteria were not-, or insufficiently matched by increased testing capacities. Quite surprisingly, our simulations for Switzerland suggested a drop in case detection when antigen-based RDT self-testing became part of the national diagnostic strategies.

## Discussion

SARS-CoV-2 continues to spread around the world, making epidemiological and molecular surveillance indispensable for the evaluation and guidance of public health interventions.

Many national and international sequencing efforts are underway that closely monitor the dynamics and evolution of the virus. In the global fight against SARS-CoV-2, the vast majority of reconstructed sequence data has been made broadly available through public databases, such as GISAID [14, 52] and the COVID data portal. In this work, we introduce GInPipe, a pipeline that utilizes this data to reconstruct SARS-CoV-2 incidence histories.

Viral infections are often characterized by a transmission bottleneck [32], where only a very small number of viruses initiate the infection and subsequently replicate within the host. A sufficient number of viruses (viral load) is required for further transmission. Hence, the temporal window of infectiousness begins with the intra-host viral population reaching a sufficiently large abundance and ends with the virus becoming eliminated by the immune system (or drugs). In contrast to HIV or HCV, SARS-CoV-2 is almost always transmitted within days after infection [30, 17]. If neutral or favourable mutations occur during this time, they may become abundant enough to be passed on to other hosts [32]. The consequence is a well-defined duration of intra-patient evolutionary time in which the virus can randomly mutate and become transmitted subsequently. In SARS-CoV-2, this intra-patient evolutionary time appears to be short and the analysis of outbreak clusters indicates that the virus genomes from linked cases were separated by either none, or very few mutations [4, 18, 50]. Taken together, these lines of evidence suggest that evolutionary change of SARS-CoV-2, the effective viral population size, and the number of infected people are correlated.

In the past, numerous approaches have been published, with the aim to estimate the effective population size from genetic properties (reviewed in [59, 38]). A variety of methods utilize the information of temporal changes in allele frequency (reviewed in [59]), while others build on population genetic theory and phylodynamic reconstruction [16, 56, 27]. GInPipe is rather related to the first class of methods as it adapts recent works of Khatri and Burt, 2019 [23]. Essentially, GInPipe considers snap-shots of inter-patient evolution to estimate a mutational input parameter *ϕ*(*t*). The latter is proportional to the effective population size, which correlates with incidence. Taken together, GInPipe uses time-stamped SARS-CoV-2 sequences and divides them into bins of inter-patient virus evolution to estimate time-dependent incidence correlates *ϕ*_*b*_. From the set of *ϕ*_*b*_ estimates, the entire incidence history *ϕ*(*t*) can be reconstructed.

We assessed the suitability of GInPipe using *in silico* simulated outbreaks, in comparison with phylodynamics and by comparing to reported case statistics. Using simulated data, the method accurately reconstructed incidence histories (Fig. 1). It also performed robustly with incomplete data, and when foreign sequence variants were introduced (Supplementary Note 1). The method even worked when the introduced variants made up a considerable fraction of the population and did not contribute to the mutational input of the outbreak.

We also compared the method with epidemiological estimates from phylodynamic reconstruction using BDSKY [53] in BEAST2 [5], shown in Figure 2. Bayesian phylodynamic methods use Monte Carlo Markov Chain (MCMC) or similar techniques to allow for a Bayesian estimation of phylogenetic relatedness of genomes, by both estimating evolutionary parameters, as well as parameters governing an underlying epidemiological model [61, 57]. The MCMC sampling procedure makes phylodynamic inference computationally demanding and often requires to ‘down-sample’ data sets.

When the epidemiological model entails time-varying parameters, changes in the effective reproduction numbers *R*_*e*_(*τ*) can be computed. However, to enable their estimation (practical parameter identifiability), parameters of the underlying epidemiological model are typically considered to be piecewise constant or to change smoothly. In Figure 2, we show the phylodynamic estimates of the effective reproduction numbers 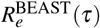. Corresponding reproductive numbers 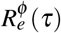 were computed with GInPipe by applying the method of Wallinga-Teunis [58] to the estimated incidence correlates *ϕ*. We compared the medians over the temporal windows used in the phylodynamic analysis. Overall, this methodological comparison yielded highly congruent predictions, with the exception of Switzerland in the first-(mid March – May 2020) and final intervals (mid September 2020 – January 2021). The ETH Zurich provides a visualization^1^ for the daily *R*_*e*_ estimates, based on reporting data. The ETH data, similarly to our daily 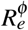 estimates with GInPipe, shows a peak, followed by a decline in the daily *R*_*e*_ for the first interval. This could explain why the *median* 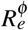 is only slightly smaller than 1 in this first interval, unlike the BEAST2 estimate, which is ≈ 0.6. For the final intervals (mid September 2020 – January 2021) 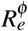 estimates fluctuate around- or slightly above *R*_*e*_(*t*) = 1, in line with the predictions of the ETH, and slightly below the BEAST2 estimate that resulted in a median *R*_*e*_ around 1.2. For the sake of this comparison, a relatively crude transmission cluster detection method was employed for the phylodynamic analyses, which may be causing a slight bias in the estimated effective reproduction numbers.

Overall, it appears that both methods yield similar results with respect to inferring the pandemic trajectories in the majority of cases. The power of GInPipe lies in the swift reconstruction of incidence histories with a fine temporal resolution, without requiring phylodynamic inference, construction of a multiple sequence alignment, down-sampling, clustering by e.g. lineages, or masking of problematic sites in the virus genomes. Moreover, GInPipe performs robustly, even in case of large proportions of introduced variants, which would also include lab-specific errors (Supplementary Note 1). However, *R*_*e*_ estimation is obviously only a side-product of phylodynamic inference, which has many more applications such as the identification and analysis of transmission clusters [19, 46], which GInPipe is not suited for. Hence, the two approaches could complement one another and GInPipe may be used to inform parameter choices for full phylodynamic inference.

To simplify the use of GInPipe, we provide an automatic workflow that can be directly applied to data downloads from GISAID or the COVID Data Portal. The execution time appears to scale linearly with the number of sequences to be analyzed (≈ 1,500 sequences per minute on a 2,3 Ghz computer with 2 cores).

When we applied GInPipe to available GISAID data from Denmark, Scotland, Switzerland, and Victoria (Australia), we observed that the reconstructed incidence histories agree well with the daily numbers of new reported infections (Fig. 3). Particularly for Denmark, reconstructed incidence histories match the reporting data quite well. Of the analyzed countries, Denmark conducted the largest number of SARS-CoV-2 tests per capita (see also *P*(tested) in Supplementary Figure 1). This could imply that the pandemic was relatively well tracked, as also suggested by relatively small changes in the diagnostic rate (Fig. 4). Moreover, a large fraction of the diagnosed cases were sequenced, providing a comprehensive genomic profile of the virus population.

For the first wave in Scotland and Victoria, we determined a much higher incidence than reported. Notably, the number of SARS-CoV-2 tests per capita was very low in Scotland, as well as in Victoria until May 2020 (*P*(tested) in Supplementary Figure 1). Thus, a large proportion of infected individuals may not have been diagnosed during this time. In Victoria and Scotland, testing capacities were increased in May, i.e. *after* the peak of the first wave.

Another striking difference of our predictions in comparison to the reported cases is that GInPipe indicates a rise of infections in August 2020 in all European countries. Notably, this increase in infections coincides with the introduction and community spread of B.1.177 (the ‘Spanish’ variant, 20E (EU1)) in most Western European countries as suggested by phylodynamic analyses [28, 20]. Our results, when compared with the reported cases, therefore imply an under-reporting of cases during the onset of community transmission of B.1.177.

Quantifying case detection is usually not feasible without knowing, or approximating the proportion of infected individuals (compare Eq. (2)). In order to do so, others have used mathematical models to predict the proportion of infected individuals [6, 1] and with this, to estimate the level of under-reporting of SARS-CoV-2. However, these mathematical models cannot be fitted to the reported cases under the presumption of an unknown trajectory of under-reporting. It therefore remains extremely difficult to parameterize suitable models for the task of assessing under-reporting, in particular for non-monotonic pandemic trajectories.

Random testing may inform the number of incidents, as well as asymptomatic infections [41]. Yet, usually only snap-shots of the incidence may be derived, which are insufficient to parameterize the aforementioned models. Moreover, it is not clear, whether the samples in the random testing scheme were representative. Sero-prevalence studies remain the gold-standard to estimate the cumulative number of infections [6, 1], as well as cumulative under-detection. Nevertheless, these studies only provide very coarse time resolution (if any) and require large sample sizes for robust analysis.

A methodologically related approach uses a semi-Bayesian approach to assess under-detection in the US [62]. To enable estimation, the probability of case detection is constrained by the assumption of particular prior distributions.

With regards to the aforementioned approaches, our method to quantify case detecting profiles has the advantage that no complex mathematical modelling is needed, and no constraints are necessary. Instead, we use information about the conducted tests and the test positive rate, in combination with the incidence correlate *ϕ*. This makes the proposed approach simple, interpretable and independent of additional assumptions.

Using this method, we observed that broad testing with little, or no suspicion of SARS-CoV-2 infection coincides with apparent under-reporting of infections from the second quarter of 2020. This coincides with a drastic decrease in the proportion of positive test results. From the latter, it is possible to compute the conditional probability that a tested person is actually infected (*P*(inf | tested), Supplementary Figure 1). A drop in *P*(inf | tested) coinciding with a steady amount of tests can negatively affect the probability to detect infected individuals *P*(tested | inf), which may have happened in the European summer of 2020. In other words, the scarce testing resources available during that time, may not have been employed in the most effective way. This suggests that it may be advisable to focus on testing symptomatic individuals when testing capacity is low.

Nevertheless, the apparent under-reporting was overcome relatively quickly by either increasing testing capacities (Denmark, Scotland, Victoria) or re-focusing capacities or both (Switzerland), Fig. 4. Interestingly, our method predicts a decline in case detection in Switzerland after the broad introduction of antigen self-testing in November 2020. A potential explanation for this observation is that only a fraction of positive antigen self-tests is confirmed by PCR and hence enters the Swiss reporting system. At the time of writing, the final interpretation of this observation is still unclear and will require further analysis.

In summary, we have developed a method that allows to reconstruct incidence histories solely based on time-stamped genetic sequences of SARS-CoV-2. We implemented the method in a fully automated workflow that can be applied to publicly available data. Moreover, this method can be used to assess the impact of testing strategies on case reporting. Finally, we envision that the method will be particularly useful to estimate the extent of the SARS-CoV-2 pandemic in regions where diagnostic surveillance is insufficient for monitoring, but may still yield a few samples for sequencing. In some of these regions pandemic control may be impossible or cause more harm than benefit and hence these regions may constitute reservoirs for the emergence of novel SARS-CoV-2 variants. Gaining insight in the pandemic dynamics in these regions through alternative methods, such as GInPipe, could yield valuable information that helps to direct global SARS-CoV-2 control efforts.

## Methods

### Data and data pre-processing

Sequences and meta data for Denmark, Scotland, Switzerland, and Victoria (Australia) were downloaded from the GISAID EpiCoV database [14, 52] (Supplementary Note 4). Sequences, where only the year of collection was provided were omitted. If year and month are specified, the 15th day of the month was added to the meta data.

The retained sequences were individually mapped to the reference (NCBI Wuhan Reference Sequence: NC_045512.2 [39]) with minimap2 version 2.17 (r941) [29]. From the mapping files (SAM), we deduced the nucleotide substitutions for each sequence. Point mutations appearing less than three times in the whole data set were filtered out, as they may occur due to sequencing errors [55].

### Construction of temporal sequence bins

SARS-CoV-2 sequences were sorted chronologically by collection date and assigned to temporal bins *b* in a redundant manner. We subdivided the sequence set into bins of

- equal size (proportions of 2%, 5%, 7% of all samples)
- spanning an equal amount of days (10, 15, and 20, and one calendar week).

Bins that contain a proportion of sequences should however span at least 3 days and maximally 21 days, and bins that span a predefined time period should contain at least 15 sequences. The date assigned to a bin is the mean collection date of the comprised sequences.

The redundant binning (‘re-sampling’) allows to evaluate cases where there is insufficient data along the time line (Figure 1A), and makes the proposed method statistically more robust to outliers.

### Incidence correlate *ϕ*_*b*_

The proposed method is inspired by the work of Khatri and Burt, 2019 [23], who derived a simple relation between the mean number of independent origins of soft selective sweeps in a population sample 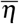, the current number of an allele *m* and mutational input, i.e. the scaled (haploid) effective population size *θ* = 2*N*_eff_*µ*, with *µ* denoting the mutation rate: 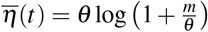.

Unlike Khatri and Burt, who aim at estimating the recent effective population size utilizing the recurrent mutations which have been fixated in the population, we seek to reconstruct the history of incidences of a population over time. We adapted the equation accordingly, also under the presumption that the *de novo* occurrence of mutations is driven by random chance events, whose likelihood may increase with the number of infected individuals [25, 10]. Seeking to estimate the incidence correlate *ϕ* = *c·N*_eff_, with the incidence being equivalent to the effective population size *N*_eff_, scaled by a constant factor *c*, we parameterize the equation as follows: For each temporal bin *b* we estimate incidence correlate *ϕ*_*b*_ at time *t*_*b*_. From the sequences comprised in bin *b*, i.e. dated within a certain time frame Δ*d*_*b*_ (Fig 1A), we infer the number of haplotypes *h*_*b*_ and the total number of mutant sequences *m*_*b*_ in the bin (Fig 1B). The mutations are determined with respect to a given reference sequence. In the original equation, we replace the mean number of origins 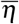 with the number of distinct variants *h*_*b*_. In each temporal bin, however, haplotypes and mutants are accumulated over the time span Δ*d*_*b*_. To correct for biases that result from this accumulation, especially for large time spans, we normalize the inputs *h*_*b*_ and *m*_*b*_ using a logistic function 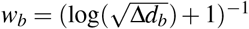.

The parameter *ϕ*_*b*_ is derived by numerically solving

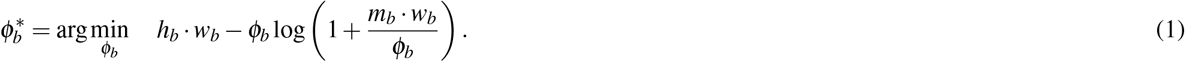

### Reconstructing the incidence history

Incidence point estimates *ϕ*_*b*_ are assigned to the mean collection date *t*_*b*_ of the sequences contained in the bin. We applied a convolution filter with window size 7 days to derive a continuous, smoothed trajectory (Fig. 1C). For uncertainty estimation, we sub-sampled *ϕ* trajectories 1000 times, by randomly leaving out 50% of the point estimates and reconstructed the trajectory by smoothing and linear interpolation between the remaining point estimates.

### Implementation and availability

All methods were implemented in Python version 3.9 and R version 4.0. A fully automated workflow has been generated using Snakemake [26] and is available from https://github.com/KleistLab/GInPipe.

### Simulation study

To test the proposed incidence reconstruction method, we stochastically simulated the evolutionary dynamics of a viral outbreak using a Poisson process formalism. We started with *N*(*t*_0_) = 50 copies of a random sequence of length *L* = 200 nt, that evolved in 120 discrete time steps, depending on a population dynamic. A succeeding generation was modelled to consist of *N*(*t* + 1) ∼*Poiss* (*N*(*t*) ·*ρ*(*t*)) sequences (= effective population size), where we chose a sinodial rate 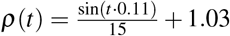. Thus, *N*(*t* + 1) sequences from the actual generation were randomly chosen with replacement and copied over to the next generation. We then introduced *n*_mut_ ∼*Poiss* (*µ·N*(*t* + 1) ·*L*) random mutations into these sequences with per site mutation rate *µ* = 0.0001.

For each generation, a fasta file with all sequences was stored and used as input for the incidence reconstruction pipeline. We ran 10 stochastic simulations with the settings stated above to compare the ground truth effective population sizes *N*(*t*) from our simulations with the corresponding inferred incidence trajectories *ϕ*.

In Supplementary Note 1, we evaluated scenarios where only a fraction of the sequences were sampled (10-90%) or, to rule out sampling biases, we sub-sampled equal amounts of sequences at each time point, independent of *N*(*t*). Moreover, we assessed whether our predictions were affected by the introduction of unrelated sequence variants into the population.

### Effective reproduction number *R*_*e*_

Based on the reconstructed incidence histories, the effective reproduction number *R*_*e*_(*t*) was computed using the established method by Wallinga and Teunis [58] (R package R0 [40]). Daily estimates of *ϕ* were assigned a pseudocount of one and rounded to the nearest integer. For the generation time distribution *g*(*τ*) of SARS-CoV-2, we chose the Gamma distribution with a mean of 5 days and a standard deviation of 1 day [15, 7].

### Phylodynamic analyses

Phylodynamic analyses were performed on subsampled sets of the data described above (*Data and data pre-processing*) using a birth-death-sampling process as implemented in the BDSKY [53] model in BEAST2 [5]. Here the precise collection day of sequence samples with only information on year and month was inferred during the analysis and not *a priori* set to the 15th. The full data sets were first grouped by Pango lineage [47, 8] and then subsampled by randomly selecting a specific percentage of sequences per week (Victoria: 10% for lineage D.2, 50% for other lineages; Switzerland: 50% for all lineages; Scotland: 20% for all lineages; Denmark: 5% for all lineages). In addition, sequences were excluded if they belonged to a lineage with less than two representatives in the analyzed set and lineages with periods longer than 75 days without any sample were split into parts. Retained sequences were aligned to the reference genome (Genbank-ID MN908947.3 [3]) in MAFFT [22] using the *–keeplength* option and problematic sites were masked by replacing the them with ‘*N*’ in the alignment [11].

For each remaining approximate cluster a separate phylogeny was reconstructed. A strict clock model with a fixed rate of 8·10^−4^ substitutions per site per year and an HKY substitution model were used. In the embedded transmission model, transmission (*λ*), recovery (*µ*) and sampling (*Ψ*) rates were assumed to be piecewise constant with changes allowed either when intervention measures changed, or in a uniform manner (Supplementary Note 2). The reproductive number *R*_*e*_(*t*) = *λ* (*t*)*/*(*µ*(*t*) + *Ψ*(*t*)) was drawn from a log-normal distribution *R*_*e*_(*t*) ∼ log *𝒩* (0, 4), the rate to become non-infectious *δ* (*t*) = *µ*(*t*) + *Ψ*(*t*) from a narrow normal distribution with *δ* (*t*) ∼ *𝒩* (27.11, 1) which is changed to *𝒩* (48.8, 1) after first control measures are implemented in the respective area. The sampling proportion *s*(*t*) = *Ψ*(*t*)*/*(*Ψ*(*t*) + *µ*(*t*)) was *a priori* assumed to arise from a uniform distribution with a lower limit of zero and the upper limit determined by the ratio of analyzed sequences over diagnosed cases *s* ∼ *U* (0, *q*_*i*_*/d*_*i*_) where *d*_*i*_ is the number of diagnoses and *q*_*i*_ the number of sequences included in the analysis in interval *i*. To account for the lineage specific subsampling, a separate sampling proportion for lineage D.2, *s*_*D*.2_, was modelled in the analysis of the Victoria data. A uniform distribution with an upper limit corresponding to the subsampling percentage was thus used as prior distribution of the D.2 specific-, as well as general sampling proportion *s*_*g*_, i.e. *s*_*D*.2_ ∼ *U* (0, 0.1) and *s*_*g*_ ∼ *U* (0, 0.5). Setup files for all four analyses can be found in the Supplementary Note 2.

MCMC chains were run until all parameters converged, i.e. showed an effective sample size greater than 200. On an Intel Xeon CPU E5-2687W (3.1 Ghz; 2 × 12 cores), this corresponded to about 10 hours to run one analysis for at least 150 million MCMC steps after burn-in (about 4min/Msample). Log files are assessed using Tracer and TreeAnnotator was used to summarize the posterior sample of phylogenetic trees to a maximum clade credibility tree using median node heights. Lineage through time plots of all summary trees were calculated using the R package ape and are shown in Supplementary Note 2.

### Relative case detection rate

We used GInPipe to detect changes in SARS-CoV-2 case detection. Let us denote by *P*_*t*_(tested |infected) the proportion of infected individuals that are actually diagnosed with the virus in week *t*. According to Bayes’ theorem we have

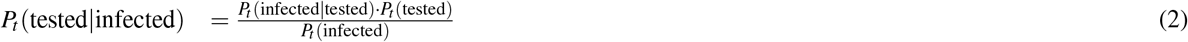

where *P*_*t*_ (infected|tested) denotes the proportion of tested individuals that are infected, *P*_*t*_ (tested) the proportion of individuals that are tested and *P*_*t*_ (infected) the proportion currently infected in week *t*. We calculate 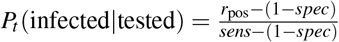 from the positivity rate *r*_pos_ of the conducted tests, corrected for the clinical sensitivity *sens*= 0.7 and specificity *spec*= 0.999 of the diagnostic tests [54]. For calculating the probability of being tested *P* (tested), we considered linear-, Poisson- and Binomial models, all of which yielded identical results. For all illustrations herein, we used the latter, yielding 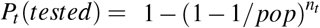, with *pop* denoting the population size in the respective regions or country and *n*_*t*_ denoting the number of tests conducted in the respective week.

The probability of currently being infected 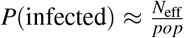 is unknown. However, since we know that *N*_eff_ is linearly correlated with the incidence estimate *ϕ*, we have 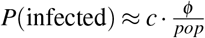. Putting everything together we can estimate the relative case detection rate:

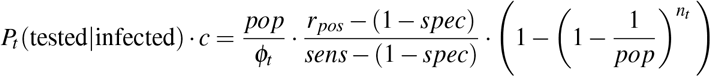

Sources for the weekly number of performed tests, as well as test positive rates are stated in Supplementary Note 3.

## Author Contributions

Conceptualization, M.R.S., M.T. and M.v.K.; Methodology, M.R.S., M.T., A.W., D.K. and M.v.K.; Investigation, M.R.S., M.T., A.W., Y.D. Writing - Original Draft, M.R.S., M.T., A.W. and M.v.K.; Writing-Review and Editing, M.R.S., M.T., A.W., D.K. and M.v.K.; Funding Acquisition, A.W., D.K. and M.v.K.; Supervision, D.K. and M.v.K.;

## Supporting information

Combined Supplementary Materials (Figures and Notes 1-4)

## Data Availability

All utilized data is publically available

## Acknowledgements

The authors acknowledge all labs contributing SARS-CoV-2 sequences to the GISAID EpiCoV database as stated in Supplementary Note 4.

## Footnotes

1 https://ibz-shiny.ethz.ch/covid-19-re-international/

