## Supplementary material for "Rapid incidence estimation from SARS-CoV-2 genomes reveals decreased case detection in Europe during summer 2020": Combined Supplementary Materials (Figures and Notes 1-4)

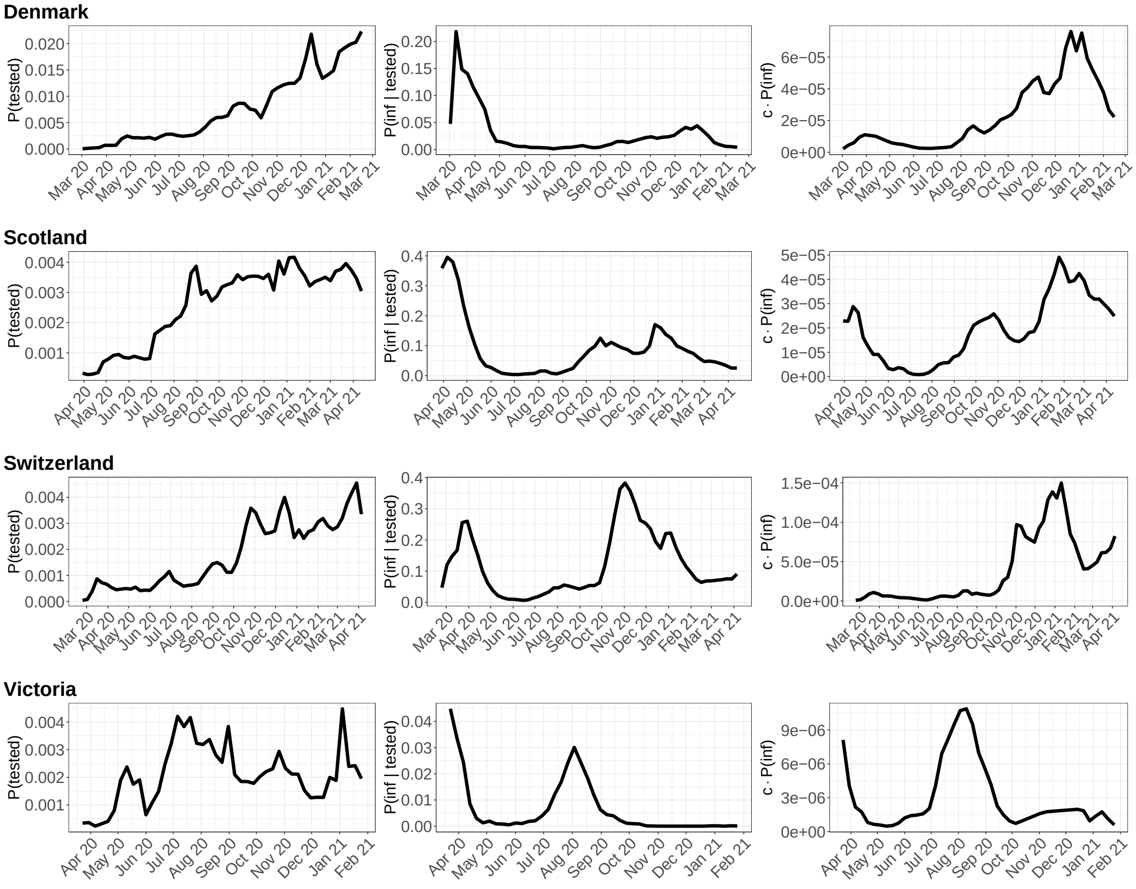


**Supplementary Figure 1:** From left to right: Probability of being tested, conditional probability that a tested individual is infected, and probability of being infected over time. From top to bottom: Denmark, Scotland, Switzerland, and Victoria (Australia).

### Supplementary Note 1

The aim of Supplementary Note 1 is to evaluate, based on in silico simulated outbreaks, whether GInPipe can infer the number of infected individuals solely from viral genetic data. Moreover, we want to study the robustness of the method to missing data, unbalanced sampling and introduction of unrelated sequence variants.

#### SN.1 *In Silico* evaluation of the proposed Method

As a proof of concept, we set up a simulation of a virtual outbreak: We formulate a stochastic epidemiological process by which a population of infectious agents progress with a time-dependent replication rate. This generates the number of infected individuals at each time step ('ground truth'). In our simulations, each infected individual can contribute to the pandemic and is associated with one viral genome. These genomes can be transmitted and randomly mutated in every time step. The set of genomes constitutes the data source for GInPipe. More precisely, we use the *in silico* generated viral genomes to compute the incidence correlate $\phi(t)$ using GInPipe. The derived $\phi(t)$ estimates are then compared with the number of infected individuals $N(t)$ at each time step.

##### SN.1.1 Summary and Limitations

We thoroughly tested the performance of our method and probed its weaknesses based on the *in silico* simulations. We found that the method's $\phi$ estimates linearly correlate with the infectious population size $N_{\text{true}}$. Different binning strategies, which are used to reconstruct the temporal incidence profiles yield complementary results. Too large temporal bins may overestimate the number of infected individuals. On the other hand, $\phi$ estimates can become inaccurate when the population of infectious individuals is small. These 'outliers' are however eliminated through application of a convolutional filter (smoothing). The $\phi$ estimates are unbiased in the case of missing data. However, the method becomes more inaccurate when too little viral sequences are available. Importation of variants (introductions) that subsequently are community transmitted, does not affect the performance of the method. The same applies to introductions which are not community transmitted. In the extreme case of persistently large frequencies of non-transmitted introductions, however, the slope of the linear correlation function may change. Yet, a linear relationship between the incidence estimate $\phi$ and the infectious population size $N_{\text{true}}$ is retained and the reconstruction of the pandemic trajectory is still possible.

##### SN.1.2 Outbreak simulation

For the evaluation of the method, we simulate the evolutionary dynamics of a viral outbreak. We simulate a time-discrete (generation-wise) population growth, where each simulation starts at time $t_0$ and ends at $t_{\text{final}} = 120$. At $t_0$, the outbreak starts with $N_{\text{init}} = 50$ identical sequences of length $L = 200$.

The nucleotides of the initial sequence are chosen randomly. In each of the $t_{\text{final}}$ generations, the current population of size $N(t)$ replicates with replication rate $\rho$ and mutates with mutation rate

$\mu$. To simulate non-monotonic outbreak trajectories, we chose a sinodial rate, depending on time $t$:

$$\rho(t) = \frac{\sin(t * 0.11)}{15} + 1.03 \tag{SN.1}$$

The number of sequences in the new generation $t + 1$ is sampled from the Poisson distribution with

$$N(t+1) \sim \mathcal{P}\big(N(t) \cdot \rho(t)\big) \tag{SN.2}$$

The sequences for the next generation are chosen randomly with replacement.

The sampled sequences of the new generation are mutated with mutation rate $\mu = 0.0001$. Analogous to the previous step, the total number of positions to be mutated is sampled from a Poisson distribution:

$$n_{\text{mut}} \sim \mathcal{P}\big(\mu \cdot N(t+1) \cdot L\big) \tag{SN.3}$$

These mutations are randomly chosen across all sequences and positions without replacement. Each nucleotide is mutated according to the Kimura substitution type model [1] with fixed rates for transitions ($A \leftrightarrow G$, $C \leftrightarrow T$ with $\alpha = 0.6$) and both types of transversions ($A \leftrightarrow C$, $G \leftrightarrow T$ with $\beta = 0.2$, as well as $A \leftrightarrow T$, $C \leftrightarrow G$ with $\gamma = 0.2$). During the course of simulation, each position could in principle be mutated several times.

This procedure results in a sequence set for each generation, which is written into a fasta file. The header of each sequence contains an ID and a date (arbitrary start date + $t$ days) to conform with the format required for the computational pipeline (`https://github.com/KleistLab/GInPipe`). We ran 10 simulations with the settings stated above. The trajectory of the population dynamics are depicted in Figure SN.2.

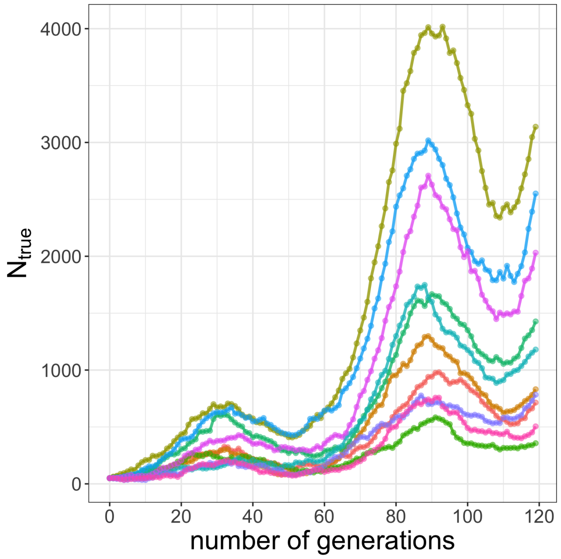


**Figure SN.2:** Trajectories for 10 simulations of population growth showing $N_{\text{true}}(t)$.

Moreover, we subsampled the the generation-wise dataset in different scenarios (section SN.1.7) and created fasta files for the respective scenarios.

#### SN.1.3 Execution of the method

For each of the generated (sub-)samples we ran the whole pipeline as described in the *Methods* section (main manuscript). 'Temporal bins' were spanning 2, 4, 6, 8, and 10 days (here days correspond to generations) and 'equal size bins' were created comprising 2, 5, and 7 percent of all sequences. We evaluated the normalised $\phi_b$ estimates for each bin $b$ according to Eq. (1) in the main manuscript.

##### SN.1.4 Incidence estimate $\phi_b$ correlates linearly with infectious population size $N_{\mathbf{true}}$

As depicted in Figure SN.3 (left), the bin-wise $\phi_b$ estimates show a strong linear relationship to the ground truth $N_{\text{true}}$ with high significance (Pearson correlation coefficient $r = 0.96$, $p < 10^{-16}$). More specifically, we observe a linear scaling $c$ of the incidence estimate $\phi$ with $N_{\text{true}}$ of the form

$$N_{\text{true}}(t) = c \cdot \phi(t) + \text{error}. \tag{SN.4}$$

The deviation grows for larger values, because the considered evolutionary model is a Poisson process with error $= \sqrt{\phi(t)} \cdot \epsilon$, where $\epsilon$ is a zero-centered, symmetric, additive error.

In Figure SN.3 (right), we compare the percent deviation from the linear regression $\hat{\phi}(t)$:

$$\%\text{deviation} = \frac{\phi_b(t_b) - \hat{\phi}(t_b)}{\hat{\phi}(t_b)} \cdot 100, \tag{SN.5}$$

with $t_b$ denoting the *mean* date of the all sequences in bin $b$. The deviation measure allows to center the approximation errors and henceforth analyse how distinct parameters may affect the method's performance.

We can see that the majority of the point estimates are quite accurately approximated with low

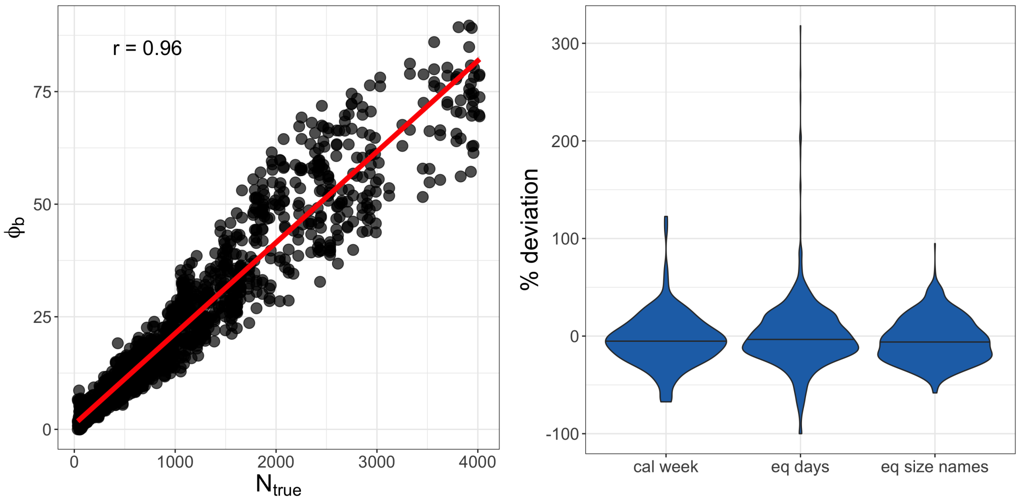


**Figure SN.3: Linear relation between $N_{\text{true}}$ and $\phi$. Left:** Dotplot showing $N_{\text{true}}$ versus the estimated point estimates $\phi_b$ per bin (full data set). The red line shows the linear fit. **Right:** Violin plot of the percent deviation of $\phi_b$ from the linear fit for the different binning types (i) calendar week, (ii) equal amount of days and (iii) equal number of sequences per bin.

percent deviation. Moreover, the estimation seems unbiased (symmetrically dispersed around 0% deviation) and large deviations are rare. Outliers are most prominent when we bin sequences by equal amounts of days (which includes calendar week), scattering in both directions, hence under- and overestimating. Creating bins of equal size, however, is slightly more prone to underestimating of $c \cdot N_{\text{true}}$.

##### SN.1.5 Effect of binning strategies

Next, we investigated the sources of outliers. In Figure SN.4, we show violin plots that colour-code the spanned days per bin (left) and the number of sequences per bin (right). Large deviations

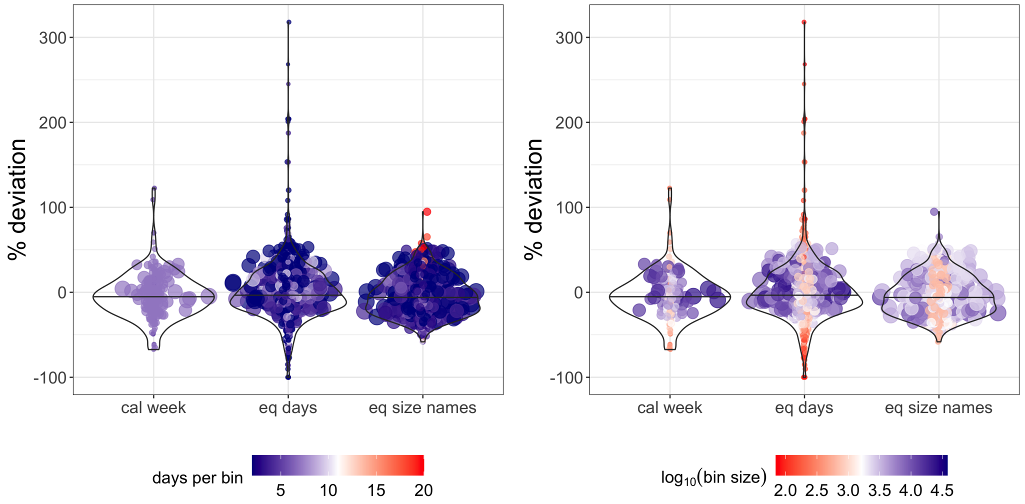


**Figure SN.4: Deviation of $\phi$ from the linear correlation to $N_{\mathbf{true}}$.** Incidence correlate $\phi$ was calculated for the different binning types, i.e. by calendar week, equal days and equal bin size. The size and dispersion of data points indicate the value of $N_{\text{true}}$. **Left:** Colour and transparency indicate the days spanned per bin, with blue and transparent for low values, red and solid for large values. **Right:** Colour and transparency indicate the number of sequences per bin (in $\log_{10}$), with blue and transparent for large bin sizes, red and solid for small bin sizes.

from the linear fit are particularly occurring for very small infectious population sizes $N_{\text{true}}$, which is indicated by the smaller markers in Fig. SN.4.

Moreover, overestimation for the 'equal size' binning is caused by bins spanning large temporal windows, around 20 days (see red dots in SN.4 (left), third violin plot). The outliers that occur for binning with equal amounts of days, to the contrary, are mainly caused by bins containing only very few sequences (red dots in the central violin plot in Figure SN.4 (left)). In these cases, the true population size is already small (small dots) and hence the amount of sequences used to infer the parameter are also low. Therefore, it may happen that the time until the next mutational event lies outside the particular bin, and not enough information is given to infer the (very low) population size.

We can conclude, that the choice of the binning strategy is important for accurate results. On the one hand, the bin has to be large enough in order to contain mutational information, but on the other hand not too large such that 'valleys' within the population dynamic can still be captured. Moreover, prediction accuracy declines with decreasing population size $N_{\text{true}}$.

##### SN.1.6 Smoothing corrects for outliers

The point estimates are smoothed using a convolution filter of a window size of 7 (days). To infer statistical uncertainty in $\phi$ profiles, we conducted a sub-sampling procedure of the $\phi$ point estimates (details in the *Methods* section, main article).

In Figure SN.5 (left), we show the smoothed reconstruction $\phi_{\text{est}}$ of the simulated dynamics of $N_{\text{true}}$, which we saw in Figure SN.2. Figure SN.5(right) also shows, in a representative simulation, that the incidence correlates $\phi$ (blue dots and line) yield a proportional estimate of $N_{\text{true}}$ (red line) and reflects the overall epidemiological dynamic quite accurately.

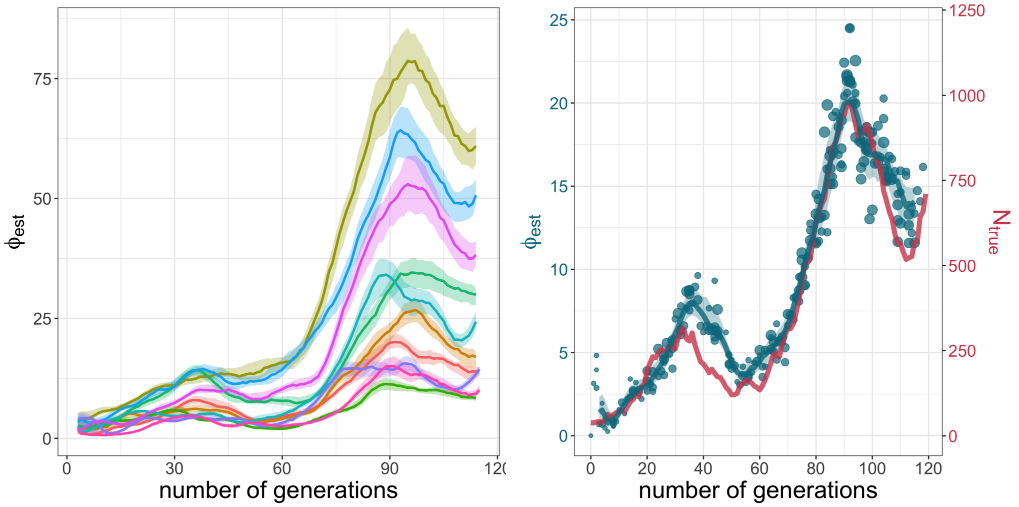


**Figure SN.5: Reconstruction of outbreak dynamics. Top:** Smoothed median of the $\phi$ estimates for the 10 stochastic simulations of population growth. The coloured shades present the area between the 5th and 95th percentile derived from sub-sampling over $\phi_b$ point estimates. **Bottom:** The blue line shows the smoothed median of the sub-sampled $\phi_b$ estimates (points) for one of the simulations. The red line shows true incidence per generation $N_{\text{true}}$.

#### SN.1.7 Missing data does not affect incidence reconstructions

In the evaluation above, we have used all sequences which emerge in the evolutionary process. For a more realistic setting, we evaluated the estimation of $\phi$ by taking sub-samples of the total sequence set: We sub-sampled a certain fraction of all sequences (WSREL = proportional sub-sampling), and drew the same amount of sequences per generation (WSABS = unbalanced sub-sampling). For the sub-sampling schemes, we tested the following parameters:

- WSREL: fraction of sequences used for inference $r_{\text{sub}} \in \{0.9, 0.7, 0.5, 0.3, 0.1\}$
- WSABS: fixed amount of sequences at each generation (up to) $a_{\text{sub}} \in \{500, 350, 150, 50\}$

The sequences are randomly chosen without replacement at every time point.

In Figure SN.6, we show violin plots of the percent deviation, akin to Eq. (SN.5) for the raw point estimates (yellow) and the smoothed estimates (green) for different sub-sampling schemes. We can generally observe that $\phi$ is strongly correlated with the population size $N_{\text{true}}$ ($r > 0.87$), even in scenarios with significant under-sampling. Moreover, $\phi$ is an unbiased predictor of the incidence (median deviation centered around 0). The deviation from the linear correlation line slightly increases with more severe under-sampling, i.e. the $\phi$ estimates are still linearly correlated with $N_{\text{true}}$, but the $\phi$ estimates become less accurate.

#### SN.1.8 Introductions of new sequences do not affect prediction accuracy

Above, we evaluated an isolated outbreak, where the diversity of mutants is emerging solely through evolution, i.e. reproduction and mutation. However, in case of a widespread epidemic, several outbreaks at different locations could lead to populations which evolve in parallel, causing different strains to emerge. Foreign strains may then be introduced into a local epidemic at any point in time.
To test whether the introduction of foreign strains affects the proposed method, we added $x_i \in \{5, 10, 50, 100, 500\}$ 'introduction sequences' at random time points to the initial outbreak. In all

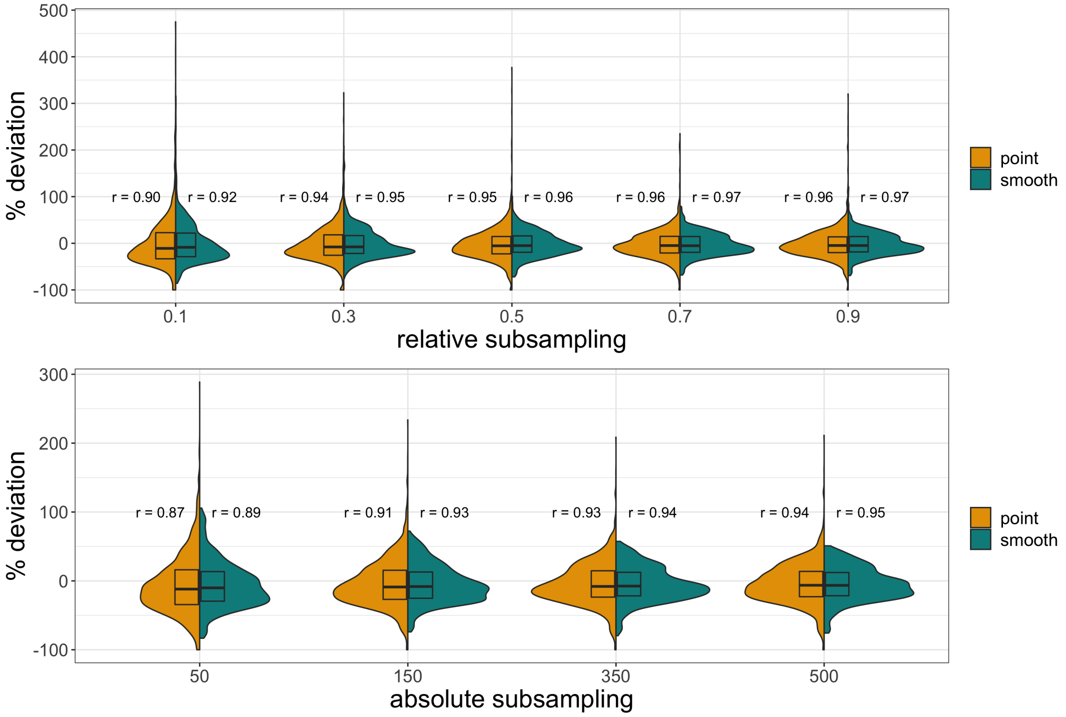


**Figure SN.6: Split violin plot showing the percent deviation of the linear fit for $\phi$ estimates for respective levels of subsampling.** The box indicates the IQR with the median. The left, yellow part shows the deviation for the point estimates, the right, grey part depicts the deviation for the smoothed medians. On each side of the violin plot, the $r$ value indicates the respective level of linear relationship of $\phi$ and $N_{\text{true}}$. **Top:** Proportional sub-sampling. **Bottom:** Absolute sub-sampling.

simulations, the 'introduction sequences' have 10% of the genome randomly mutated with regards to the founder sequence of the local outbreak. We could also have taken completely random sequences, which would presumably result in the same outcome, but we wanted to keep the framework more realistic, using slightly modified strains which have independently emerged.
We performed 5 simulations that began with 1–10 start sequences. All sequences were considered to evolve with the same replication and mutation rates.
For a representative simulation, Figure SN.7 (left) shows the simulated epidemiological dynamics for different numbers of introductions.
In Figure SN.7 (right), we see the corresponding correlations of the incidence estimate $\phi$ with the true population size $N_{\text{true}}$. The inferred $\phi$ estimates show a strong linear correlation with $N_{\text{true}}$ across all simulated scenarios ($r = 0.96$, $p < 10^{-16}$).
In Figure SN.8 we evaluate how the relative amount of introductions affects the percent deviation, akin to Eq. (SN.5). We can see that for the majority of the simulations the deviation is around zero with no apparent trend of increasing dispersion (prediction inaccuracy), when large numbers of foreign variants become introduced.
In summary, the predictions are not perturbed by introductions of foreign sequences. Potential biases from single introductions are compensated by ongoing local evolution (community transmission) of the imported sequences.

To investigate the effect of introductions without the compensating effect of community transmissions, we simulated an extreme scenario, where the introduced sequences are only added to the local outbreak and do not replicate or mutate. They simply occur as additional haplotypes in the population. In reality this would be the case, if infected people from a different region get tested, their virus sequenced and subsequent quarantine would completely prevent further transmission of imported variants.

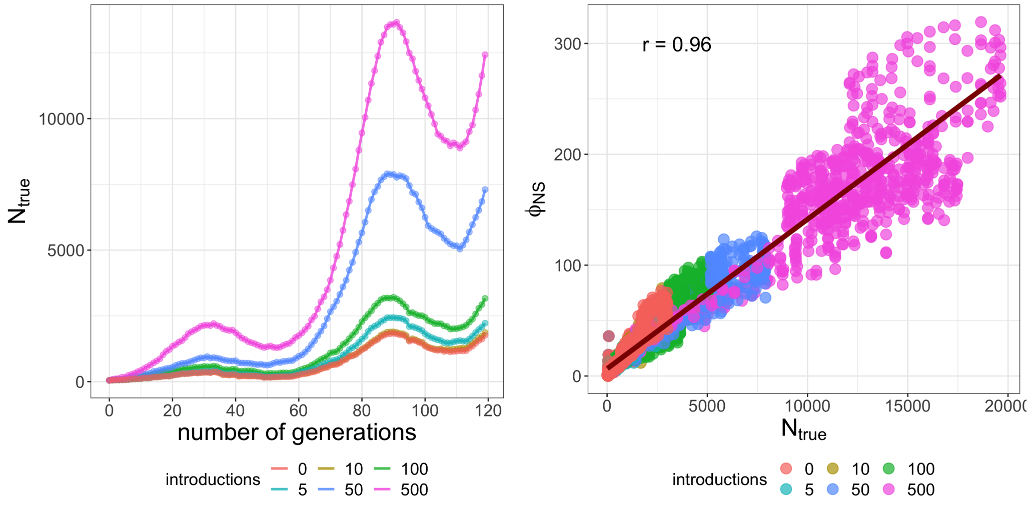


**Figure SN.7: Simulations with foreign sequence introductions**. **Left:** True population dynamics for the original outbreak (0 introductions) and after introduction of new sequences, each in a representative simulation. The total amount of introduced sequences at random time points is colour-coded. **Right:** Dotplot showing $N_{\text{true}}$ versus the point estimates $\phi$ using 5 simulations per 'number of introductions' scenario. The colours indicate the number of introduced foreign sequences to the original outbreak. The black line shows the linear fit.

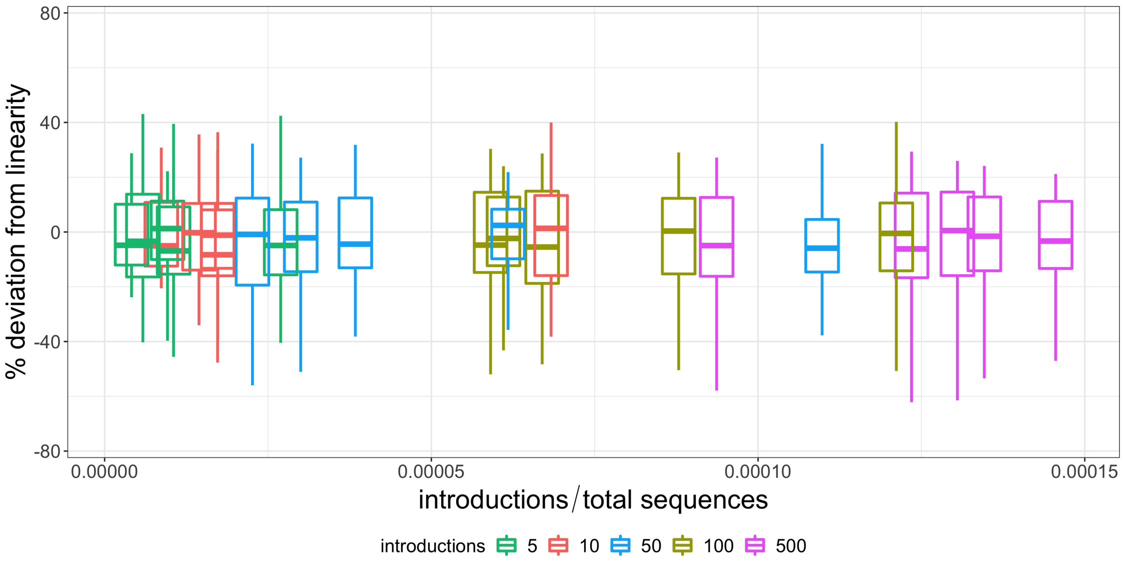


**Figure SN.8: Deviation from linear correlation for smoothed $\phi$ with introduced sequences.** Boxplots showing for each simulation the percent deviation of smoothed $\phi$ from the linear fit to $N_{\text{true}}$ in comparison to the ratio of introductions and total amount of emerging sequences in the evolutionary process. The box includes the IQR with median and whiskers show 1.5 IQR. The colour indicates the number of introductions.

For illustration, Figure SN.9 (top left) shows a simulation where introduced variants are not onwards transmitted. Unlike in Fig. SN.7 (left) where imported variants spread, we can see that the outbreak dynamics are not affected by the non-replicating introductions.

The corresponding $\phi$ estimates in Figure SN.9 (top right) show the following: In simulations, where 10–100 variants where introduced, the corresponding $\phi$ estimate remains quantitatively unchanged in comparison to a 'zero-introduction' simulation. In the extreme case, where 500 variants were introduced, (pink line in Figure SN.9, top right), the dynamical profile of the number of infected individuals is still very accurately captured (highlighting the 2.5 waves of infection). However, quantitatively the $\phi$ estimates are slightly increased in comparison to the 'zero-introduction' simulation. Thus, if one would like to compare a setting, where there are many non-replicating introductions, to a setting with replicating- or 'zero-' introductions, the quantitative interpretation of the $\phi$ estimates with regards to the true number of infected individuals $N_{\text{true}}$ might slightly differ. There may be a slightly different slope for the linear correlation function of $\phi$ vs. $N_{\text{true}}$.

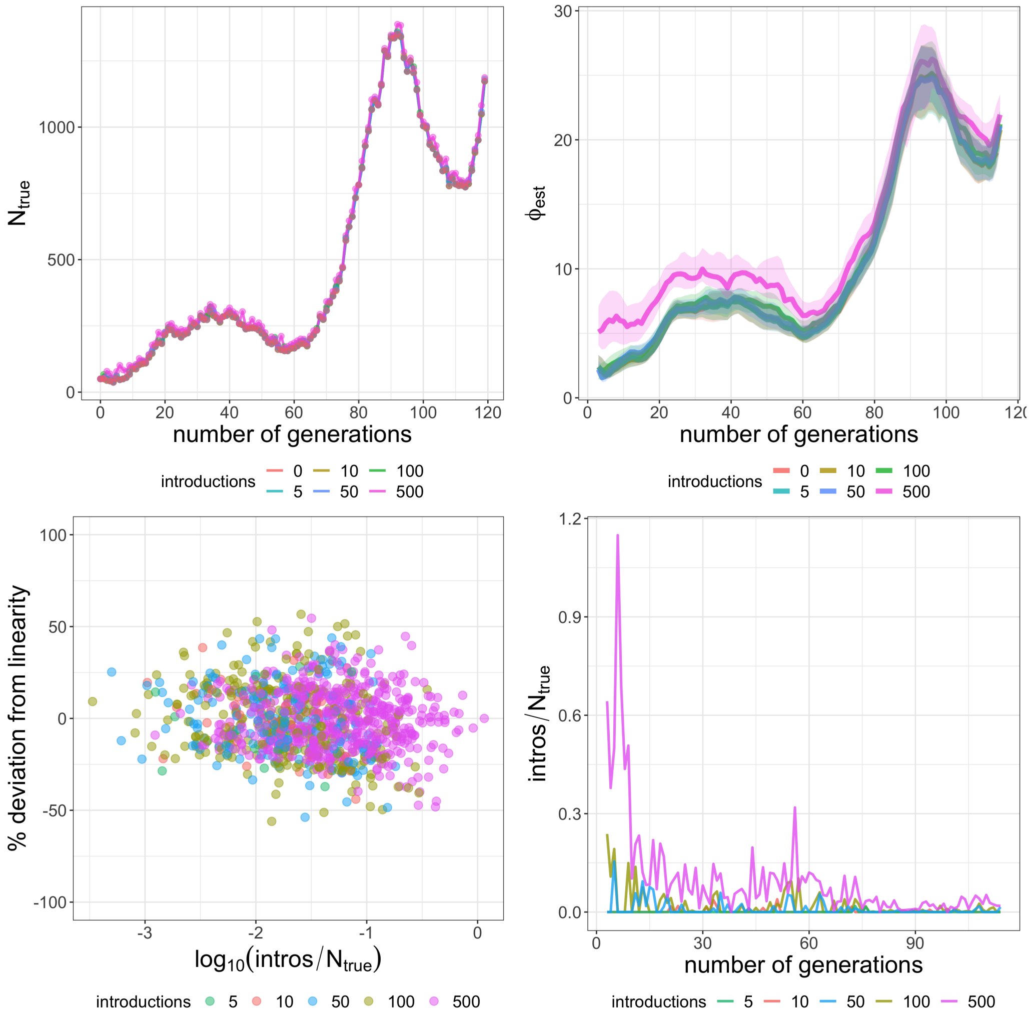


**Figure SN.9: Simulations for introduced variants that do not spread in the population.** The total amount of introduced sequences at random time points is colour coded. **Top left:** True population size over time for the original outbreak (0 introductions) and after introduction of new sequences for a representative simulation. **Top right:** Smoothed median of the $\phi$ estimates for one stochastic simulation with different amounts of introductions. The shaded areas denote the 90% confidence interval. **Bottom left:** Dotplot showing the percent deviation of the smoothed $\phi$ estimates from the respective linear fit to $N_{\text{true}}$ in relation to the relative frequency ($\log_{10}$) of introductions per time point. **Bottom right:** Relative amount of new introductions over time, corresponding to the simulated trajectories in the top right figure.

In Figure SN.9 (bottom left), we systematically assess whether large relative amounts of non-replicating introductions decrease prediction accuracy, i.e. increase deviation from the linear regression function for the specific simulation. We do not discover a systematic trend, even if there were more non-replicating introductions than regionally transmitting viruses. We monitor the relative amount of introductions over time for the corresponding smoothed $\phi$ estimates in the panel above (Fig. SN.9, bottom right). Looking at the ratios of introductions vs the effective population size per generation for the case of 500 introductions (pink lines), we observe that the method is still able to reconstruct the dynamic and attain a linear relationship, despite having consecutively more than 15% introductions during the first wave and partly even exceeding the number of evolving sequences.

### Supplementary Note 2

In this Supplementary Note all parameters used in the phylodynamic inference are stated, such as (i) change dates where major non-pharmaceutical measures were introduced or lifted (these intervals are used to calculate the piece-wise constant $R_e(\tau)$ values); (ii) All final parameter estimates, as well as (iii) 'lineage-through-time' plots for phylodynamic reconstruction.

#### Birth-death-sampling rate change dates

| **Date** | **Non-pharmaceutical interventions** |
|---|---|
| 18 March 2020 | Implementation of strict lockdown measures: private gathering restrictions, closure of e.g. restaurants and hairdressers |
| 28 April 2020 | Start of easing of restrictions: Opening of primary schools, later public places |
| 15 June 2020 | Further easing of restrictions: bigger private and mass gatherings allowed, homeoffice partially recommended |
| 22 August 2020 | Protective masks: Face masks mandatory in closed public spaces |
| 26 October 2020 | Reimplementation of light restrictions: private gatherings |
| 26 December 2020 | Start of lockdown: closure of schools, restaurants and entertainment venues |

**Table SN.1:** Denmark: Intervention measures used to set the birth-death-sampling rate change dates

| Date | Non-pharmaceutical interventions |
|---|---|
| 24 March 2020 | Start of national lockdown: Closure of shops, restaurants etc, stay-at-home order |
| 29 May 2020 | First lifting of restrictions: Phase 1 of route map out of lockdown |
| 15 July 2020 | Phase 3 of route map out of lockdown: Opening restaurants, hairdressers etc. |
| 23 September 2020 | Reimplementation of restrictive measures: Restrictions on household visits and national curfew |
| 17 November 2020 | Further restrictions: Update on COVID19 protection levels and travel regulations |
| 05 January 2021 | Start of second lockdown for mainland Scotland: stay-at-home order |

**Table SN.2:** Scotland: Intervention measures used to set the birth-death-sampling rate change dates

| Date | Non-pharmaceutical interventions |
|---|---|
| 16 March 2020 | Start of national lockdown: Closure of shops, museums, restaurants, other gathering places and borders |
| 11 May 2020 | Lifting of stricter lockdown measures: Opening of i.a. schools, shops and museums |
| 15 June 2020 | Second step in reopening the borders |
| 20 August 2020 | Unspecified |
| 19 October 2020 | Reimplementation of restrictions: home office recommendation, restrictions on private gatherings and for restaurants |

**Table SN.3:** Switzerland: Intervention measures used to set the birth-death-sampling rate change dates

| Date | Non-pharmaceutical interventions |
|---|---|
| 16 March 2020 | State of Emergency and first restrictions |
| 11 May 2020 | First easing of restrictions |
| 19 July 2020 | Reimplementation of restrictions, especially around Melbourne |
| 13 September 2020 | First step in roadmap for reopening |
| 26 October 2020 | Zero new cases recorded |

**Table SN.4:** Victoria, Australia: Intervention measures used to set the birth-death-sampling rate change dates

#### Phylogenetic and epidemiological parameter estimates

| | median | 95% HPDI |
|---|---|---|
| become non−infectious rate $\delta_1$ | 2.6501e+01 | [2.4488e+01, 2.8412e+01] |
| become non−infectious rate $\delta_2$ | 4.8708e+01 | [4.6744e+01, 5.0617e+01] |
| become non−infectious rate $\delta_3$ | 4.8740e+01 | [4.6804e+01, 5.0663e+01] |
| become non−infectious rate $\delta_4$ | 4.8952e+01 | [4.7046e+01, 5.0941e+01] |
| become non−infectious rate $\delta_5$ | 5.1244e+01 | [4.9410e+01, 5.2993e+01] |
| become non−infectious rate $\delta_6$ | 5.9074e+01 | [5.7414e+01, 6.0797e+01] |
| become non−infectious rate $\delta_7$ | 5.7028e+01 | [5.5238e+01, 5.8826e+01] |
| reproductive number $R_{e,1}$ | 3.9112e+00 | [2.8245e+00, 5.0122e+00] |
| reproductive number $R_{e,2}$ | 1.0472e+00 | [8.7220e−01, 1.2374e+00] |
| reproductive number $R_{e,3}$ | 7.7224e−01 | [6.2073e−01, 9.1495e−01] |
| reproductive number $R_{e,4}$ | 1.0155e+00 | [9.6244e−01, 1.0693e+00] |
| reproductive number $R_{e,5}$ | 1.0607e+00 | [1.0273e+00, 1.0941e+00] |
| reproductive number $R_{e,6}$ | 1.1818e+00 | [1.1542e+00, 1.2094e+00] |
| reproductive number $R_{e,7}$ | 1.0427e+00 | [1.0175e+00, 1.0676e+00] |
| sampling proportion $s_1$ | 0.0000e+00 | [0.0000e+00, 0.0000e+00] |
| sampling proportion $s_2$ | 1.4238e−02 | [4.6416e−03, 2.7545e−02] |
| sampling proportion $s_3$ | 1.1721e−03 | [3.5208e−04, 2.2823e−03] |
| sampling proportion $s_4$ | 9.1423e−04 | [4.0777e−04, 1.5568e−03] |
| sampling proportion $s_5$ | 2.8559e−03 | [1.9751e−03, 3.8874e−03] |
| sampling proportion $s_6$ | 1.0887e−02 | [1.0523e−02, 1.1000e−02] |
| sampling proportion $s_7$ | 9.3824e−03 | [9.3254e−03, 9.4000e−03] |
| sampling proportion $s_8$ | 1.3949e−02 | [1.3784e−02, 1.4000e−02] |
| frequency parameter $\pi_A$ | 2.9851e−01 | [2.9794e−01, 2.9917e−01] |
| frequency parameter $\pi_C$ | 1.8342e−01 | [1.8294e−01, 1.8396e−01] |
| frequency parameter $\pi_G$ | 1.9591e−01 | [1.9539e−01, 1.9646e−01] |
| frequency parameter $\pi_T$ | 3.2216e−01 | [3.2151e−01, 3.2278e−01] |
| transition/transversion bias | 5.0636e+00 | [4.7320e+00, 5.4283e+00] |
| gamma shape parameter | 6.8115e−01 | [1.0269e−03, 2.9279e+00] |
| clock rate | 8.0000e−04 | [8.0000e−04, 8.0000e−04] |

**Table SN.5:** Denmark: Median and 95% highest posterior density interval (HPDI) of the posterior sample of the model-specific phylogenetic and epidemiological parameters. Subscript of the latter indicates the interval for which it is inferred. Intervals are labelled numerically from the date of the first to the date of the last sample with boundaries as specified in Table SN.1

| | median | 95% HPDI |
|---|---|---|
| become non−infectious rate $\delta_1$ | 2.6011e+01 | [2.4032e+01, 2.8011e+01] |
| become non−infectious rate $\delta_2$ | 4.9913e+01 | [4.8009e+01, 5.1799e+01] |
| become non−infectious rate $\delta_3$ | 4.8026e+01 | [4.6081e+01, 4.9913e+01] |
| become non−infectious rate $\delta_4$ | 5.0591e+01 | [4.8654e+01, 5.2602e+01] |
| become non−infectious rate $\delta_5$ | 5.7396e+01 | [5.5629e+01, 5.9142e+01] |
| become non−infectious rate $\delta_6$ | 5.4126e+01 | [5.2255e+01, 5.5933e+01] |
| become non−infectious rate $\delta_7$ | 4.9603e+01 | [4.7673e+01, 5.1513e+01] |
| reproductive number $R_{e,1}$ | 2.3930e+00 | [2.1652e+00, 2.6366e+00] |
| reproductive number $R_{e,2}$ | 9.6874e−01 | [9.3089e−01, 1.0066e+00] |
| reproductive number $R_{e,3}$ | 8.8540e−01 | [8.0281e−01, 9.6845e−01] |
| reproductive number $R_{e,4}$ | 1.4266e+00 | [1.3718e+00, 1.4830e+00] |
| reproductive number $R_{e,5}$ | 9.5870e−01 | [9.2372e−01, 9.9171e−01] |
| reproductive number $R_{e,6}$ | 1.2504e+00 | [1.2047e+00, 1.2994e+00] |
| reproductive number $R_{e,7}$ | 1.2587e+00 | [1.1726e+00, 1.3460e+00] |
| sampling proportion $s_1$ | 0.0000e+00 | [0.0000e+00, 0.0000e+00] |
| sampling proportion $s_2$ | 4.4042e−02 | [3.1986e−02, 5.7453e−02] |
| sampling proportion $s_3$ | 4.3064e−02 | [3.6441e−02, 4.9981e−02] |
| sampling proportion $s_4$ | 7.2413e−03 | [5.6073e−03, 8.0000e−03] |
| sampling proportion $s_5$ | 3.9766e−02 | [3.3262e−02, 4.6574e−02] |
| sampling proportion $s_6$ | 9.9783e−03 | [9.9043e−03, 1.0000e−02] |
| sampling proportion $s_7$ | 5.4743e−03 | [5.3902e−03, 5.5000e−03] |
| sampling proportion $s_8$ | 7.3377e−03 | [6.6098e−03, 7.5999e−03] |
| frequency parameter $\pi_A$ | 2.9857e−01 | [2.9806e−01, 2.9918e−01] |
| frequency parameter $\pi_C$ | 1.8351e−01 | [1.8304e−01, 1.8396e−01] |
| frequency parameter $\pi_G$ | 1.9605e−01 | [1.9556e−01, 1.9653e−01] |
| frequency parameter $\pi_T$ | 3.2186e−01 | [3.2127e−01, 3.2241e−01] |
| transition/transversion bias | 6.0910e+00 | [5.5833e+00, 6.6160e+00] |
| gamma shape parameter | 6.6908e−01 | [1.0983e−03, 3.1089e+00] |
| clock rate | 8.0000e−04 | [8.0000e−04, 8.0000e−04] |

**Table SN.6:** Scotland: Median and 95% highest posterior density interval (HPDI) of the posterior sample of the model-specific phylogenetic and epidemiological parameters. Subscript of the latter indicates the interval for which it is inferred. Intervals are labelled numerically from the date of the first to the date of the last sample with boundaries as specified in Table SN.2

| | median | 95% HPDI |
|---|---|---|
| become non−infectious rate $\delta_1$ | 2.6636e+01 | [2.4662e+01, 2.8596e+01] |
| become non−infectious rate $\delta_2$ | 4.9531e+01 | [4.7653e+01, 5.1450e+01] |
| become non−infectious rate $\delta_3$ | 4.8525e+01 | [4.6562e+01, 5.0471e+01] |
| become non−infectious rate $\delta_4$ | 4.9118e+01 | [4.7172e+01, 5.1068e+01] |
| become non−infectious rate $\delta_5$ | 5.4165e+01 | [5.2425e+01, 5.5999e+01] |
| become non−infectious rate $\delta_6$ | 5.3494e+01 | [5.1588e+01, 5.5354e+01] |
| reproductive number $R_{e,1}$ | 3.7785e+00 | [3.4824e+00, 4.0843e+00] |
| reproductive number $R_{e,2}$ | 5.9984e−01 | [5.4907e−01, 6.5164e−01] |
| reproductive number $R_{e,3}$ | 1.0081e+00 | [9.1422e−01, 1.0989e+00] |
| reproductive number $R_{e,4}$ | 1.0692e+00 | [1.0286e+00, 1.1117e+00] |
| reproductive number $R_{e,5}$ | 1.2212e+00 | [1.1854e+00, 1.2577e+00] |
| reproductive number $R_{e,6}$ | 1.2447e+00 | [1.2197e+00, 1.2700e+00] |
| sampling proportion $s_1$ | 0.0000e+00 | [0.0000e+00, 0.0000e+00] |
| sampling proportion $s_2$ | 2.9001e−02 | [2.2124e−02, 3.6373e−02] |
| sampling proportion $s_3$ | 1.2163e−02 | [1.0312e−02, 1.3000e−02] |
| sampling proportion $s_4$ | 5.6867e−03 | [3.3805e−03, 8.4220e−03] |
| sampling proportion $s_5$ | 2.1780e−02 | [1.7993e−02, 2.6085e−02] |
| sampling proportion $s_6$ | 1.0947e−02 | [1.0776e−02, 1.1000e−02] |
| sampling proportion $s_7$ | 1.9903e−03 | [1.9587e−03, 2.0000e−03] |
| frequency parameter $\pi_A$ | 2.9852e−01 | [2.9794e−01, 2.9914e−01] |
| frequency parameter $\pi_C$ | 1.8346e−01 | [1.8293e−01, 1.8395e−01] |
| frequency parameter $\pi_G$ | 1.9598e−01 | [1.9548e−01, 1.9653e−01] |
| frequency parameter $\pi_T$ | 3.2205e−01 | [3.2145e−01, 3.2266e−01] |
| transition/transversion bias | 5.1965e+00 | [4.7981e+00, 5.5953e+00] |
| gamma shape parameter | 7.1509e−01 | [1.2162e−03, 3.0187e+00] |
| clock rate | 8.0000e−04 | [8.0000e−04, 8.0000e−04] |

**Table SN.7:** Switzerland: Median and 95% highest posterior density interval (HPDI) of the posterior sample of the model-specific phylogenetic and epidemiological parameters. Subscript of the latter indicates the interval for which it is inferred. Intervals are labelled numerically from the date of the first to the date of the last sample with boundaries as specified in Table SN.3

| | median | 95% HPDI |
|---|---|---|
| become non−infectious rate $\delta_1$ | 2.6176e+01 | [2.4184e+01, 2.8121e+01] |
| become non−infectious rate $\delta_2$ | 4.9619e+01 | [4.7642e+01, 5.1533e+01] |
| become non−infectious rate $\delta_3$ | 5.1129e+01 | [4.9253e+01, 5.3030e+01] |
| become non−infectious rate $\delta_4$ | 5.3906e+01 | [5.2126e+01, 5.5761e+01] |
| become non−infectious rate $\delta_5$ | 4.8738e+01 | [4.6758e+01, 5.0695e+01] |
| become non−infectious rate $\delta_6$ | 4.8760e+01 | [4.6777e+01, 5.0721e+01] |
| reproductive number $R_{e,1}$ | 1.4979e+00 | [1.3556e+00, 1.6498e+00] |
| reproductive number $R_{e,2}$ | 8.5338e−01 | [7.5441e−01, 9.2525e−01] |
| reproductive number $R_{e,3}$ | 1.5827e+00 | [1.5006e+00, 1.6670e+00] |
| reproductive number $R_{e,4}$ | 8.2540e−01 | [7.7799e−01, 8.7474e−01] |
| reproductive number $R_{e,5}$ | 2.3492e−01 | [2.4743e−07, 5.6690e−01] |
| reproductive number $R_{e,6}$ | 2.0812e+00 | [1.3322e+00, 2.9222e+00] |
| sampling proportion $s_{g,1}$ | 0.0000e+00 | [0.0000e+00, 0.0000e+00] |
| sampling proportion $s_{g,2}$ | 2.9672e−02 | [1.9741e−02, 4.1761e−02] |
| sampling proportion $s_{g,3}$ | 1.7338e−01 | [1.3296e−01, 2.1817e−01] |
| sampling proportion $s_{g,4}$ | 6.4884e−02 | [4.2755e−02, 9.0529e−02] |
| sampling proportion $s_{g,5}$ | 2.2576e−01 | [8.9750e−02, 4.1722e−01] |
| sampling proportion $s_{g,6}$ | 1.6813e−01 | [6.2391e−03, 4.3783e−01] |
| sampling proportion $s_{g,7}$ | 1.7311e−02 | [6.2422e−10, 1.0029e−01] |
| sampling proportion $s_{D.2,1}$ | 0.0000e+00 | [0.0000e+00, 0.0000e+00] |
| sampling proportion $s_{D.2,2}$ | 4.9214e−02 | [3.1247e−04, 9.5247e−02] |
| sampling proportion $s_{D.2,3}$ | 2.5908e−02 | [1.6407e−06, 8.5224e−02] |
| sampling proportion $s_{D.2,4}$ | 9.9072e−02 | [9.6042e−02, 1.0000e−01] |
| sampling proportion $s_{D.2,5}$ | 9.9457e−02 | [9.7727e−02, 1.0000e−01] |
| sampling proportion $s_{D.2,6}$ | 8.4776e−02 | [5.6447e−02, 1.0000e−01] |
| sampling proportion $s_{D.2,7}$ | 5.0036e−02 | [5.0101e−03, 9.9902e−02] |
| frequency parameter $\pi_A$ | 2.9860e−01 | [2.9780e−01, 2.9945e−01] |
| frequency parameter $\pi_C$ | 1.8358e−01 | [1.8287e−01, 1.8427e−01] |
| frequency parameter $\pi_G$ | 1.9610e−01 | [1.9536e−01, 1.9679e−01] |
| frequency parameter $\pi_T$ | 3.2173e−01 | [3.2089e−01, 3.2257e−01] |
| transition/transversion bias | 6.3126e+00 | [5.5129e+00, 7.2442e+00] |
| gamma shape parameter | 6.8480e−01 | [1.0102e−03, 3.0228e+00] |
| clock rate | 8.0000e−04 | [8.0000e−04, 8.0000e−04] |

**Table SN.8:** Victoria: Median and 95% highest posterior density interval (HPDI) of the posterior sample of the model-specific phylogenetic and epidemiological parameters. Subscript of the latter indicates the interval for which it is inferred. Intervals are labelled numerically from the date of the first to the date of the last sample with boundaries as specified in Table SN.4

#### Identified Pango lineages and tree reconstructions

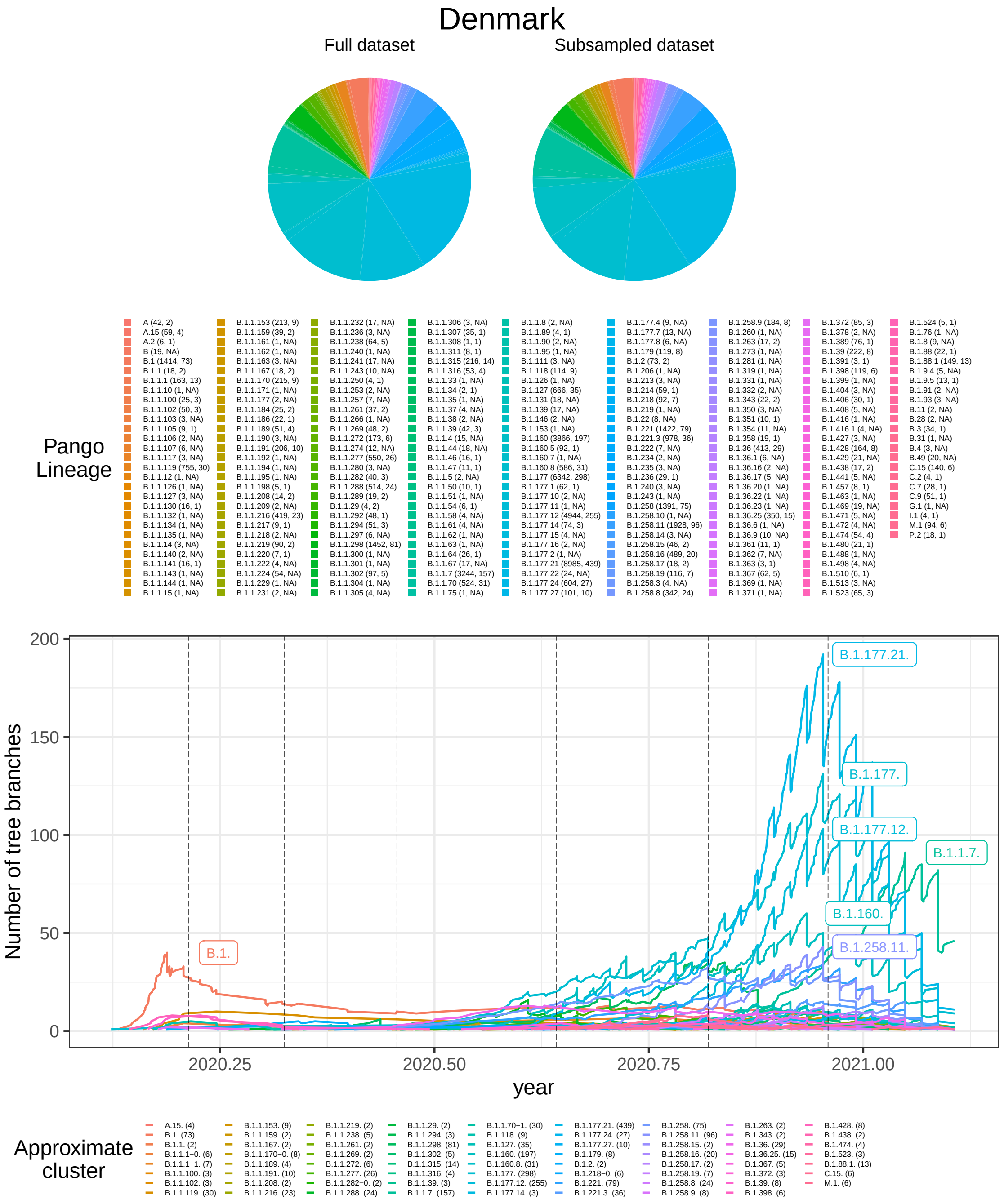


**Figure SN.10: Denmark:** Top: Pango lineages identified in the full (left) and subsampled (right) data set with the coloured area being proportional to the size of the specific lineage. Values in brackets next to the lineage name in the legend represent the number of sequences identified in the full (left) and subsampled (right) data set. Bottom: Lineage-through-time plot, counting the number of phylogenetic tree branches over time, of maximum clade credibility tree for all identified approximate clusters. Pango lineages that were split due to a longer period without any sample are denoted numerically using a dash separator, e.g. B.1.1.1-0 and B.1.1.1-1. Vertical dotted lines represent the dates at which birth, death and sampling rate were allowed to change. The number of sequences included for each approximate cluster in the analysis are given in brackets next to the cluster name in the legend.

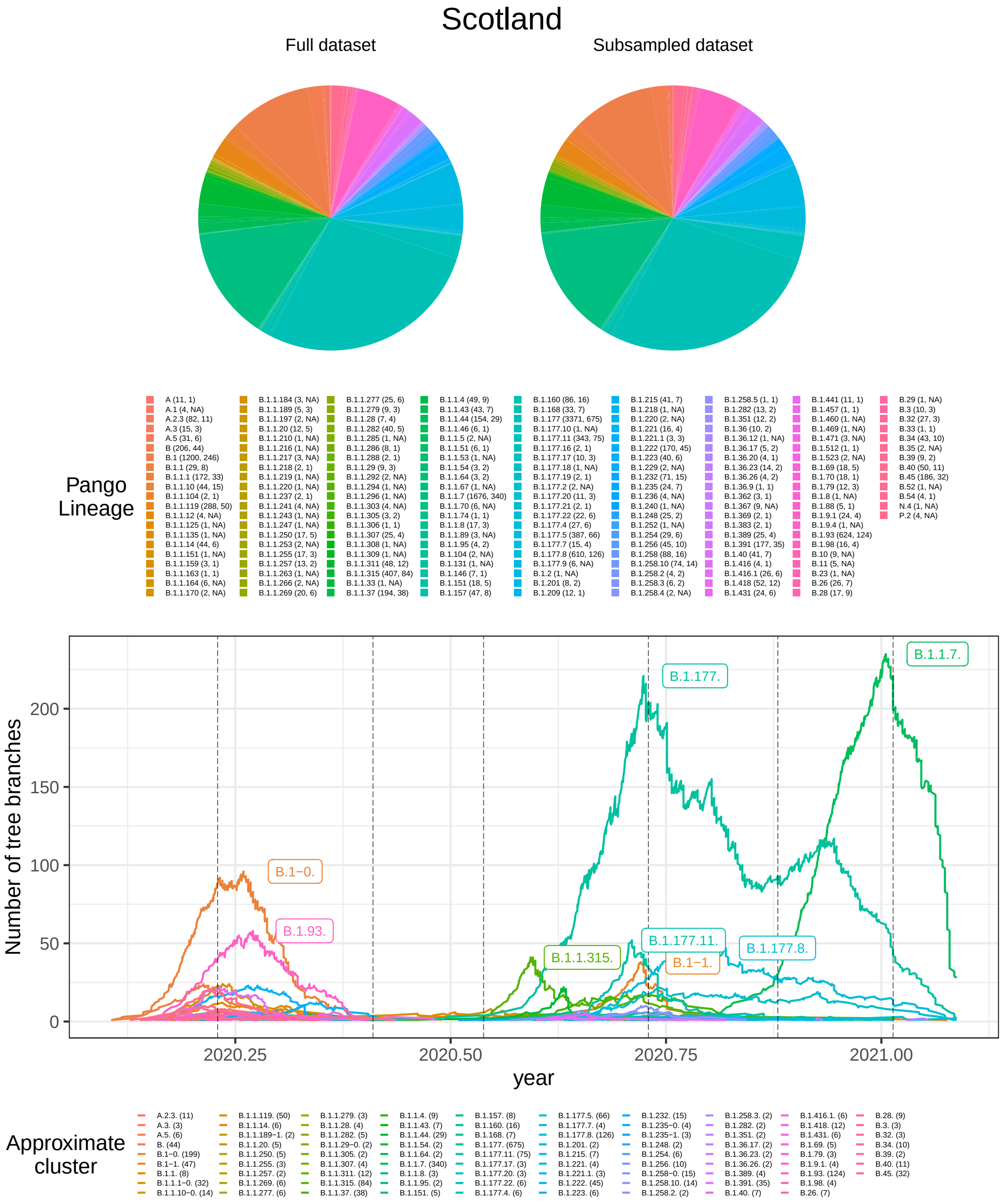


**Figure SN.11: Scotland:** Top: Pango lineages identified in the full (left) and subsampled (right) data set with the coloured area being proportional to the size of the specific lineage. Values in brackets next to the lineage name in the legend represent the number of sequences identified in the full (left) and subsampled (right) data set. Bottom: Lineage-through-time plot, counting the number of phylogenetic tree branches over time, of maximum clade credibility tree for all identified approximate clusters. Pango lineages that were split due to a longer period without any sample are denoted numerically using a dash separator, e.g. B.1-0 and B.1-1. Vertical dotted lines represent the dates at which birth, death and sampling rate were allowed to change. The number of sequences included for each approximate cluster in the analysis are given in brackets next to the cluster name in the legend.

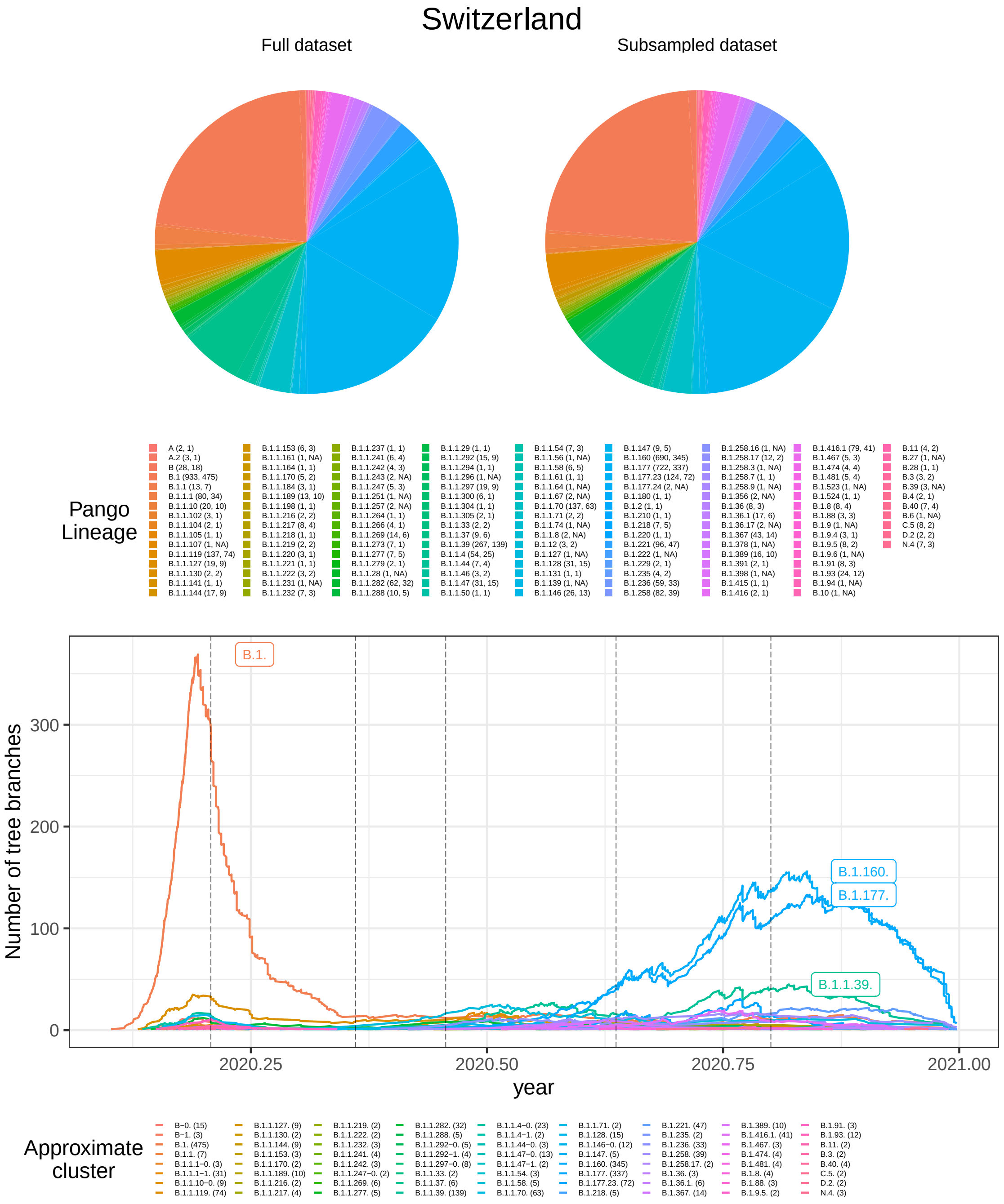


**Figure SN.12: Switzerland:** Top: Pango lineages identified in the full (left) and subsampled (right) data set with the coloured area being proportional to the size of the specific lineage. Values in brackets next to the lineage name in the legend represent the number of sequences identified in the full (left) and subsampled (right) data set. Bottom: Lineage-through-time plot, counting the number of phylogenetic tree branches over time, of maximum clade credibility tree for all identified approximate clusters. Pango lineages that were split due to a longer period without any sample are denoted numerically using a dash separator, e.g. B-0 and B-1. Vertical dotted lines represent the dates at which birth, death and sampling rate were allowed to change. The number of sequences included for each approximate cluster in the analysis are given in brackets next to the cluster name in the legend.

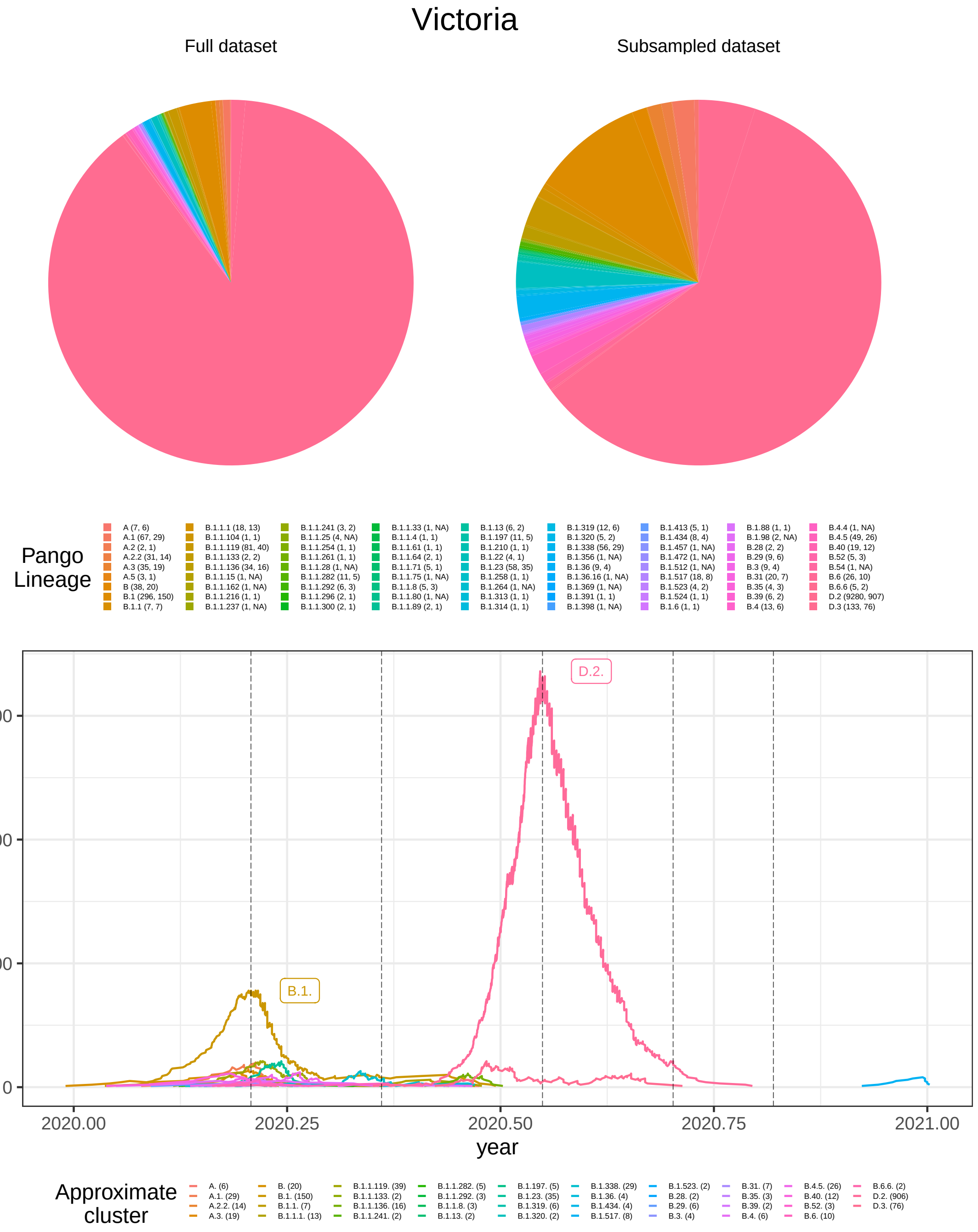


**Figure SN.13: Victoria, Australia:** Top: Pango lineages identified in the full (left) and subsampled (right) data set with the coloured area being proportional to the size of the specific lineage. Values in brackets next to the lineage name in the legend represent the number of sequences identified in the full (left) and subsampled (right) data set. Due to the lineage-specific subsampling procedure, the relative lineage compostion is different between the full and the subsampled dataset. Bottom: Lineage-through-time plot, counting the number of phylogenetic tree branches over time, of maximum clade credibility tree for all identified approximate clusters. Vertical dotted lines represent the dates at which birth, death and sampling rate were allowed to change. The number of sequences included for each approximate cluster in the analysis are given in brackets next to the cluster name in the legend.

### Supplementary Note 3

This Supplementary Note contains all weblinks that point to the primary data depicted or discussed in the main manuscript. For example, the data related to (i) changes in non-pharmaceutical interventions, which are used in BEAST2; (ii) case reporting data which is depicted in Fig. 3 (main article); (iii) the number of tests performed and the proportion of positive tests, which are used to calculate the proportion of detected cases; and (iv) changes in testing strategies, which are depicted in Fig. 4 (main article).

#### Supplementary Links

##### Changes in non-pharmaceutical interventions

The data sources stated herein are used to determine change dates for phylodynamic reconstruction. The latter is used to compute $R_e$ values that are depicted in Fig. 2 (main manuscript).
**Denmark and Switzerland (Phylodynamics).** WHO Coronavirus (COVID-19) Dashboard. Link: `https://covid19.who.int/table`, (accessed: 2021-03-10)
**Victoria (Phylodynamics).** Victorian coronavirus (COVID-19) data. Link: `https://www.dhhs.vic.gov.au/victorian-coronavirus-covid-19-data`, accessed: 2021-03-10)
**Scotland (Phylodynamics).** Coronavirus (COVID-19): trends in daily data. Link: `https://www.gov.scot/publications/coronavirus-covid-19-trends-in-daily-data`, (accessed: 2021-03-10)

##### Case reports

Reported SARS-CoV-2 infections are depicted in Fig. 3 (main manuscript) alongside the reconstructed incidence histories using GInPipe.
**Denmark and Switzerland.** Daily new reported cases. Link: `https://github.com/owid/covid-19-data/tree/master/public/data`, (accessed: 2021-04-27)
**Scotland.** New reported cases by date. Link: `https://coronavirus.data.gov.uk/details/download`, (accessed: 2021-04-27)
**Victoria (AU).** Daily new reported cases by state. Link: `https://public.tableau.com/profile/vicdhhs#!/vizhome/Cases_15982342702770/DashCasesGSG`, (accessed: 2021-04-27)

##### Number of tests performed and the test positivity

This data is used in Fig. 4 (main manuscript) to compute the case detection rate as outlined in the *Methods* section.
**Denmark.** A cross-country database of COVID-19 testing. Link: `https://covid.ourworldindata.org/data/owid-covid-data.csv`, (accessed: 2021-03-11)
**Scotland.** Coronavirus (COVID-19): trends in daily data. Link: `https://www.gov.scot/publications/coronavirus-covid-19-trends-in-daily-data`, (accessed: 2021-03-11)
**Switzerland.** A cross-country database of COVID-19 testing . Link: `https://covid.ourworldindata.org/data/owid-covid-data.csv`, (accessed: 2021-03-11)

**Victoria.** Testing for COVID-19. Link: https://www.covid19data.com.au/testing, (accessed: 2021-03-19)

#### Testing Strategy

Data on major changes in testing strategies are depicted in Fig. 4 as vertical dashed lines.
**Denmark.** Policy responses for Denmark. Link: https://www.covid19healthsystem.org/countries/denmark/livinghit.aspx?Section=1.5%20Testing&Type=Section, (accessed: 2021-03-11)
**Scotland.** Timeline of Coronavirus (COVID-19) in Scotland. Link: https://spice-spotlight.scot/2021/03/12/timeline-of-coronavirus-covid-19-in-scotland, (accessed: 2021-03-11)
**Switzerland.** Policy responses for Switzerland (Covid-19 Health System Response Monitor). Link: https://www.covid19healthsystem.org/countries/switzerland/livinghit.aspx?Section=1.5%20Testing&Type=Section, (accessed: 2021-03-18)
**Victoria.** Updates about the outbreak of the coronavirus disease (COVID-19). Link: https://www.dhhs.vic.gov.au/coronavirus/updates, (accessed: 2021-03-11)

### Supplementary Note 4

#### GISAID acknowledgement tables

##### Denmark

We gratefully acknowledge the following Authors from the Originating laboratories responsible for obtaining the specimens, as well as the Submitting laboratories where the genome data were generated and shared via GISAID, on which this research is based.

All Submitters of data may be contacted directly via www.gisaid.org

Authors are sorted alphabetically.

| Accession ID | Originating Laboratory | Submitting Laboratory | Authors |
|---|---|---|---|
| EPI_ISL_1065158, EPI_ISL_1065476, EPI_ISL_1065490, EPI_ISL_1065547, EPI_ISL_1065586, EPI_ISL_1065725, EPI_ISL_1065843, EPI_ISL_1065879, EPI_ISL_1066055, EPI_ISL_1066329, EPI_ISL_1066354, EPI_ISL_1066434, EPI_ISL_1066442, EPI_ISL_1066894, EPI_ISL_1066900, EPI_ISL_1066966, EPI_ISL_1067042, EPI_ISL_1067536 | | | |
| see above | Department of Virus and Microbiological Special Diagnostics, Statens Serum Institut, Copenhagen, Denmark | Aalborg University | Danish Covid-19 Genome Consortium |
| EPI_ISL_415646 | Department of Virus and Microbiological Special diagnostics, Statens Serum Institut, Copenhagen, Denmark. | ViFU | Morten Rasmussen, Maiken Worsoe Rosenstierne , Anders Fomsgaard |
| EPI_ISL_415647 | Department of Virus and Microbiological Special diagnostics, Statens Serum Institut, Copenhagen, Denmark. | Statens Serum Institute | Morten Rasmussen, Maiken Worsoe Rosenstierne , Anders Fomsgaard |
| EPI_ISL_415648 | Department of Virus and Microbiological Special diagnostics, Statens Serum Institut, Copenhagen, Denmark. | ViFU | Morten Rasmussen, Maiken Worsoe Rosenstierne , Anders Fomsgaard |
| EPI_ISL_416140, EPI_ISL_416141, EPI_ISL_416142 | Department of Virus and Microbiological Special diagnostics, Statens Serum Institut, Copenhagen, Denmark. | Statens Serum Institute | Morten Rasmussen, Maiken Worsoe Rosenstierne , Anders Fomsgaard |
| EPI_ISL_416143, EPI_ISL_416144, EPI_ISL_416153 | Department of Virus and Microbiological Special diagnostics, Statens Serum Institut, Copenhagen, Denmark. | ViFU | Morten Rasmussen, Maiken Worsoe Rosenstierne , Anders Fomsgaard |
| EPI_ISL_429262, EPI_ISL_429263, EPI_ISL_429264, EPI_ISL_429265, EPI_ISL_429266, EPI_ISL_429267, EPI_ISL_429268, EPI_ISL_429269, EPI_ISL_429270, EPI_ISL_429271, EPI_ISL_429272, EPI_ISL_429273, EPI_ISL_429274, EPI_ISL_429275, EPI_ISL_429276, EPI_ISL_429277, EPI_ISL_429278, EPI_ISL_429279, EPI_ISL_429280, EPI_ISL_429281, EPI_ISL_429282, EPI_ISL_429283, EPI_ISL_429284, EPI_ISL_429285, EPI_ISL_429286, EPI_ISL_429287, EPI_ISL_429288, EPI_ISL_429289, EPI_ISL_429290, EPI_ISL_429291, EPI_ISL_429292, EPI_ISL_429293, EPI_ISL_429294, EPI_ISL_429295, EPI_ISL_429296, EPI_ISL_429297, EPI_ISL_429298, EPI_ISL_429299, EPI_ISL_429300, EPI_ISL_429301, EPI_ISL_429302, EPI_ISL_429303, EPI_ISL_429304, EPI_ISL_429305, EPI_ISL_429306, EPI_ISL_429307, EPI_ISL_429308, EPI_ISL_429309, EPI_ISL_429310, EPI_ISL_429311, EPI_ISL_429312, EPI_ISL_429313, EPI_ISL_429314, EPI_ISL_429315, EPI_ISL_429316, EPI_ISL_429317, EPI_ISL_429318, EPI_ISL_429320, EPI_ISL_429321, EPI_ISL_429322, EPI_ISL_429323, EPI_ISL_429324, EPI_ISL_429325, EPI_ISL_429326, EPI_ISL_429327, EPI_ISL_429328, EPI_ISL_429329, EPI_ISL_429330, EPI_ISL_429331, EPI_ISL_429332 | | | |
| see above | Department of Clinical Microbiology, Copenhagen University Hospital, Hvidovre, Kettegaard Alle 30, 2650 Hvidovre. | Albertsen lab, Department of Chemistry and Bioscience, Aalborg University, Denmark | Rasmus Kirkegaard |
| EPI_ISL_429333, EPI_ISL_429334, EPI_ISL_429335, EPI_ISL_429336, EPI_ISL_429337, EPI_ISL_429338, EPI_ISL_429339, EPI_ISL_429340, EPI_ISL_429341, EPI_ISL_429342, EPI_ISL_429343, EPI_ISL_429344, EPI_ISL_429345, EPI_ISL_429346, EPI_ISL_429347, EPI_ISL_429348, EPI_ISL_429349, EPI_ISL_429350, EPI_ISL_429351, EPI_ISL_429352, EPI_ISL_429353, EPI_ISL_429354, EPI_ISL_429355, EPI_ISL_429356, EPI_ISL_429357, EPI_ISL_429358, EPI_ISL_429359, EPI_ISL_429360, EPI_ISL_429361, EPI_ISL_429362, EPI_ISL_429363, EPI_ISL_429364, EPI_ISL_429365, EPI_ISL_429366, EPI_ISL_429367, EPI_ISL_429368, EPI_ISL_429369, EPI_ISL_429370, EPI_ISL_429371, EPI_ISL_429372, EPI_ISL_429373, EPI_ISL_429374, EPI_ISL_429375, EPI_ISL_429376, EPI_ISL_429377, EPI_ISL_429378, EPI_ISL_429379, EPI_ISL_429380, EPI_ISL_429381, EPI_ISL_429382, EPI_ISL_429383, EPI_ISL_429384, EPI_ISL_429385, EPI_ISL_429386, EPI_ISL_429387, EPI_ISL_429388, EPI_ISL_429389, EPI_ISL_429390, EPI_ISL_429391, EPI_ISL_429392, EPI_ISL_429393, EPI_ISL_429394, EPI_ISL_429395, EPI_ISL_429396, EPI_ISL_429397, EPI_ISL_429398, EPI_ISL_429399, EPI_ISL_429400, EPI_ISL_429401, EPI_ISL_429402, EPI_ISL_429403, EPI_ISL_429404, EPI_ISL_429405, EPI_ISL_429406, EPI_ISL_429407, EPI_ISL_429408, EPI_ISL_429409, EPI_ISL_429410, EPI_ISL_429411, EPI_ISL_429412, EPI_ISL_429413, EPI_ISL_429414, EPI_ISL_429415, EPI_ISL_429416, EPI_ISL_429417, EPI_ISL_429418, EPI_ISL_429419, EPI_ISL_429420, EPI_ISL_429421, EPI_ISL_429422, EPI_ISL_429423, EPI_ISL_429424, EPI_ISL_429425, EPI_ISL_429426, EPI_ISL_429427, EPI_ISL_429428, EPI_ISL_429429, EPI_ISL_429430, EPI_ISL_429431, EPI_ISL_429432, EPI_ISL_429433, EPI_ISL_429434, EPI_ISL_429435, EPI_ISL_429436, EPI_ISL_429437, EPI_ISL_429438, EPI_ISL_429439, EPI_ISL_429440, EPI_ISL_429441, EPI_ISL_429442, EPI_ISL_429443, EPI_ISL_429444, EPI_ISL_429445, EPI_ISL_429446, EPI_ISL_429447, EPI_ISL_429448, EPI_ISL_429449, EPI_ISL_429450, EPI_ISL_429451, EPI_ISL_429452, EPI_ISL_429453, EPI_ISL_429454, EPI_ISL_429455, EPI_ISL_429456, EPI_ISL_429457, EPI_ISL_429458, EPI_ISL_429459, EPI_ISL_429460, EPI_ISL_429461, EPI_ISL_429462, EPI_ISL_429463, EPI_ISL_429464, EPI_ISL_429465, EPI_ISL_429466, EPI_ISL_429467, EPI_ISL_429468, EPI_ISL_429469, EPI_ISL_429470, EPI_ISL_429471, EPI_ISL_429472, EPI_ISL_429473, EPI_ISL_429474, EPI_ISL_429475, EPI_ISL_429476, EPI_ISL_429477, EPI_ISL_429478, EPI_ISL_429479, EPI_ISL_429480, EPI_ISL_429481, EPI_ISL_429482, EPI_ISL_429483, EPI_ISL_429484, EPI_ISL_429485, EPI_ISL_429486, EPI_ISL_429487, EPI_ISL_429488, EPI_ISL_429489, EPI_ISL_429490, EPI_ISL_429491, EPI_ISL_429492, EPI_ISL_429493, EPI_ISL_429494, EPI_ISL_429495, EPI_ISL_429496, EPI_ISL_429497, EPI_ISL_429498, EPI_ISL_429499, EPI_ISL_429500, EPI_ISL_429501, EPI_ISL_429502, EPI_ISL_429503, EPI_ISL_429504, EPI_ISL_429505, EPI_ISL_429506, EPI_ISL_429507, EPI_ISL_429508, EPI_ISL_429509, EPI_ISL_429510, EPI_ISL_429511, EPI_ISL_429512, EPI_ISL_429513, EPI_ISL_429514, EPI_ISL_429515, EPI_ISL_429516, EPI_ISL_429517, EPI_ISL_429518, EPI_ISL_429519, EPI_ISL_429520, EPI_ISL_429521, EPI_ISL_429522, EPI_ISL_429523, EPI_ISL_429524, EPI_ISL_429525, EPI_ISL_429526, EPI_ISL_429527, EPI_ISL_429528, EPI_ISL_429529, EPI_ISL_429530, EPI_ISL_429531, EPI_ISL_429532, EPI_ISL_429533, EPI_ISL_429534, EPI_ISL_429535, EPI_ISL_429536, EPI_ISL_429537, EPI_ISL_429538, EPI_ISL_429539, EPI_ISL_429540, EPI_ISL_429541, EPI_ISL_429542, EPI_ISL_429543, EPI_ISL_429544, EPI_ISL_429545, EPI_ISL_429546, EPI_ISL_429547, EPI_ISL_429548, EPI_ISL_429549, EPI_ISL_429550, EPI_ISL_429551, EPI_ISL_429552, EPI_ISL_429553, EPI_ISL_429554, EPI_ISL_429555, EPI_ISL_429556, EPI_ISL_429557, EPI_ISL_429558, EPI_ISL_429559, EPI_ISL_429560, EPI_ISL_429561, EPI_ISL_429562, EPI_ISL_429563, EPI_ISL_429564, EPI_ISL_429565, EPI_ISL_429566, EPI_ISL_429567, EPI_ISL_429568, EPI_ISL_429569, EPI_ISL_429570, EPI_ISL_429571, EPI_ISL_429572, EPI_ISL_429573, EPI_ISL_429574, EPI_ISL_429575, EPI_ISL_429576, EPI_ISL_429577, EPI_ISL_429578, EPI_ISL_429579, EPI_ISL_429580, EPI_ISL_429581, EPI_ISL_429582, EPI_ISL_429583, EPI_ISL_429584, EPI_ISL_429585, EPI_ISL_429586, EPI_ISL_429587, EPI_ISL_429588, EPI_ISL_429589, EPI_ISL_429590, EPI_ISL_436962, EPI_ISL_436963, EPI_ISL_436964, EPI_ISL_436965, EPI_ISL_436966, EPI_ISL_436967, EPI_ISL_436968, EPI_ISL_436969, EPI_ISL_436970, EPI_ISL_436971, EPI_ISL_436972, EPI_ISL_436973, EPI_ISL_436974, EPI_ISL_436975, EPI_ISL_436976, EPI_ISL_436977, EPI_ISL_436978, EPI_ISL_436979, EPI_ISL_436980, EPI_ISL_436981, EPI_ISL_436982, EPI_ISL_436983, EPI_ISL_436984, EPI_ISL_436985, EPI_ISL_436986, EPI_ISL_436987, EPI_ISL_436988, EPI_ISL_436989, EPI_ISL_436990, EPI_ISL_436991, EPI_ISL_436992, EPI_ISL_436993, EPI_ISL_436994, EPI_ISL_436995, EPI_ISL_436996, EPI_ISL_436997, EPI_ISL_436998, EPI_ISL_436999, EPI_ISL_437000, EPI_ISL_437001, EPI_ISL_437002, EPI_ISL_437003, EPI_ISL_437004, EPI_ISL_437005, EPI_ISL_437006, EPI_ISL_437007, EPI_ISL_437008, EPI_ISL_437009, EPI_ISL_437010, EPI_ISL_437011, EPI_ISL_437012, EPI_ISL_437013, EPI_ISL_437014, EPI_ISL_437015, EPI_ISL_437016, EPI_ISL_437017, EPI_ISL_437018, EPI_ISL_437019, EPI_ISL_437020, EPI_ISL_437021, EPI_ISL_437022, EPI_ISL_437023, EPI_ISL_437024, EPI_ISL_437025, EPI_ISL_437026, EPI_ISL_437027, EPI_ISL_437028, EPI_ISL_437029, EPI_ISL_437030, EPI_ISL_437031, EPI_ISL_437032, EPI_ISL_437033, EPI_ISL_437034, EPI_ISL_437035, EPI_ISL_437036, EPI_ISL_437037, EPI_ISL_437038, EPI_ISL_437039, EPI_ISL_437040, EPI_ISL_437041, EPI_ISL_437042, EPI_ISL_437627, EPI_ISL_437628, EPI_ISL_437629, EPI_ISL_437630, EPI_ISL_437631, EPI_ISL_437632, EPI_ISL_437633, EPI_ISL_437634, EPI_ISL_437635, EPI_ISL_437636, EPI_ISL_437637, EPI_ISL_437638, EPI_ISL_437639, EPI_ISL_437640, EPI_ISL_437641, EPI_ISL_437642, EPI_ISL_437643, EPI_ISL_437644, EPI_ISL_437645, EPI_ISL_437646, EPI_ISL_437647, EPI_ISL_437648, EPI_ISL_437649, EPI_ISL_437650, EPI_ISL_437651, EPI_ISL_437652, EPI_ISL_437653, EPI_ISL_437654, EPI_ISL_437655, EPI_ISL_437656, EPI_ISL_437657, EPI_ISL_437658, EPI_ISL_437659, EPI_ISL_437660, EPI_ISL_437661, EPI_ISL_437662, EPI_ISL_437663, EPI_ISL_437664, EPI_ISL_437665, EPI_ISL_437666, EPI_ISL_437667, EPI_ISL_437668, EPI_ISL_437669, EPI_ISL_437670, EPI_ISL_437671, EPI_ISL_437672, EPI_ISL_437673, EPI_ISL_437674, EPI_ISL_437675, EPI_ISL_437676, EPI_ISL_437677, EPI_ISL_437678, EPI_ISL_437679, EPI_ISL_437680, EPI_ISL_437681, EPI_ISL_437682, EPI_ISL_437683, EPI_ISL_444817, EPI_ISL_444818, EPI_ISL_444819, EPI_ISL_444820, EPI_ISL_444821, EPI_ISL_444822, EPI_ISL_444823, EPI_ISL_444824, EPI_ISL_444825, EPI_ISL_444826, EPI_ISL_444827, EPI_ISL_444828, EPI_ISL_444829, EPI_ISL_444830, EPI_ISL_444831, EPI_ISL_444832, EPI_ISL_444833, EPI_ISL_444834, EPI_ISL_444835, EPI_ISL_444836, EPI_ISL_444837, EPI_ISL_444838, EPI_ISL_444839, EPI_ISL_444840, EPI_ISL_444841, EPI_ISL_444842, EPI_ISL_444843, EPI_ISL_444844, EPI_ISL_444845, EPI_ISL_444846, EPI_ISL_444847, EPI_ISL_444848, EPI_ISL_444849, EPI_ISL_444850, EPI_ISL_444851, EPI_ISL_444852, EPI_ISL_444853, EPI_ISL_444854, EPI_ISL_444855, EPI_ISL_444856, EPI_ISL_444857, EPI_ISL_444858, EPI_ISL_444859, EPI_ISL_444860, EPI_ISL_444861, EPI_ISL_444862, EPI_ISL_444863, EPI_ISL_444864, EPI_ISL_444865, EPI_ISL_444866, EPI_ISL_444867, EPI_ISL_444868, EPI_ISL_444869, EPI_ISL_444870, EPI_ISL_444871, EPI_ISL_444872, EPI_ISL_444873, EPI_ISL_444874, EPI_ISL_444875, EPI_ISL_444876, EPI_ISL_444877, EPI_ISL_444878, EPI_ISL_444879, EPI_ISL_444880, EPI_ISL_444881, EPI_ISL_444882, EPI_ISL_444883, EPI_ISL_444884, EPI_ISL_444885, EPI_ISL_444886, EPI_ISL_444887, EPI_ISL_444888, EPI_ISL_444889, EPI_ISL_444890, EPI_ISL_444891, EPI_ISL_444892, EPI_ISL_444893, EPI_ISL_444894, EPI_ISL_444895, EPI_ISL_444896, EPI_ISL_444897, EPI_ISL_444898, EPI_ISL_444899, EPI_ISL_444900, EPI_ISL_444901, EPI_ISL_444902, EPI_ISL_444903, EPI_ISL_444904, EPI_ISL_444905, EPI_ISL_444906, EPI_ISL_444907, EPI_ISL_444908, EPI_ISL_444909, EPI_ISL_444910, EPI_ISL_444911, EPI_ISL_444912, EPI_ISL_444913, EPI_ISL_444914, EPI_ISL_444915, EPI_ISL_444916, EPI_ISL_444917, EPI_ISL_444918, EPI_ISL_444919, EPI_ISL_444920, EPI_ISL_444921, EPI_ISL_444922, EPI_ISL_444923, EPI_ISL_444924, EPI_ISL_444925, EPI_ISL_444926, EPI_ISL_444927, EPI_ISL_444928, EPI_ISL_444929, EPI_ISL_444930, EPI_ISL_444931, EPI_ISL_444932, EPI_ISL_444933, EPI_ISL_444934, EPI_ISL_444935, EPI_ISL_444936, EPI_ISL_444937, EPI_ISL_444938, EPI_ISL_444939, EPI_ISL_444940, EPI_ISL_444941, EPI_ISL_444942, EPI_ISL_444943, EPI_ISL_444944, EPI_ISL_444945, EPI_ISL_444946, EPI_ISL_444947, EPI_ISL_444948, EPI_ISL_444949, EPI_ISL_444950, EPI_ISL_444951, EPI_ISL_444952, EPI_ISL_444953, EPI_ISL_444954, EPI_ISL_444955, EPI_ISL_444956, EPI_ISL_444957, EPI_ISL_444958, EPI_ISL_444959, EPI_ISL_444960, EPI_ISL_444961, EPI_ISL_444962, EPI_ISL_444963, EPI_ISL_444964, EPI_ISL_444965, EPI_ISL_444966, EPI_ISL_444967, EPI_ISL_444968 | | | |
| see above | Department of Virus and Microbiological Special Diagnostics, Statens Serum Institut, Copenhagen, Denmark, Artillerivej 5, 2300 Copenahgen S | Albertsen lab, Department of Chemistry and Bioscience, Aalborg University, Denmark | Rasmus Kirkegaard |
| EPI_ISL_451988, EPI_ISL_451989, EPI_ISL_451990, EPI_ISL_451991, EPI_ISL_451992, EPI_ISL_451993, EPI_ISL_451994, EPI_ISL_451995, EPI_ISL_451996, EPI_ISL_451997, EPI_ISL_451998, EPI_ISL_451999, EPI_ISL_452000, EPI_ISL_452001, EPI_ISL_452002, EPI_ISL_452003, EPI_ISL_452004, EPI_ISL_452005, EPI_ISL_452006, EPI_ISL_452007, EPI_ISL_452008, EPI_ISL_452009, EPI_ISL_452010, EPI_ISL_452011, EPI_ISL_452012, EPI_ISL_452013, EPI_ISL_452014, EPI_ISL_452015, EPI_ISL_452016, EPI_ISL_452017, EPI_ISL_452018, EPI_ISL_452019, EPI_ISL_452020, EPI_ISL_452021, EPI_ISL_452022, EPI_ISL_452023, EPI_ISL_452024, EPI_ISL_452025, EPI_ISL_452026, EPI_ISL_452027, EPI_ISL_452028, EPI_ISL_452029, EPI_ISL_452030, EPI_ISL_452031, EPI_ISL_452032, EPI_ISL_452033, EPI_ISL_452034, EPI_ISL_452035, EPI_ISL_452036, EPI_ISL_452037, EPI_ISL_452038, EPI_ISL_452039, EPI_ISL_452040, EPI_ISL_452041, EPI_ISL_452042, EPI_ISL_452043, EPI_ISL_452044, EPI_ISL_452045, EPI_ISL_452046, EPI_ISL_452047, EPI_ISL_452048, EPI_ISL_452049, EPI_ISL_452050, EPI_ISL_452051, EPI_ISL_452052, EPI_ISL_452053, EPI_ISL_452054, EPI_ISL_452055, EPI_ISL_452056, EPI_ISL_452057, EPI_ISL_452058, EPI_ISL_452059, EPI_ISL_452060, EPI_ISL_452061, EPI_ISL_452062, EPI_ISL_452063, EPI_ISL_452064, EPI_ISL_452065, EPI_ISL_452066, EPI_ISL_452067, EPI_ISL_452068, EPI_ISL_452069, EPI_ISL_452070, EPI_ISL_452071, EPI_ISL_452072, EPI_ISL_452073, EPI_ISL_452074, EPI_ISL_452075, EPI_ISL_452076, EPI_ISL_452077, EPI_ISL_452078, EPI_ISL_452079, EPI_ISL_452080, EPI_ISL_452081, EPI_ISL_452082, EPI_ISL_452083, EPI_ISL_452084, EPI_ISL_452085, EPI_ISL_452086, EPI_ISL_452087, EPI_ISL_452088, EPI_ISL_452089, EPI_ISL_452090, EPI_ISL_452091, EPI_ISL_452092, EPI_ISL_452093, EPI_ISL_452094, EPI_ISL_452095, EPI_ISL_452096, EPI_ISL_452097, EPI_ISL_452098, EPI_ISL_452099, EPI_ISL_452100 | | | |
| see above | Department of Clinical Microbiology, Copenhagen University Hospital, Hvidovre, Kettegaard Alle 30, 2650 Hvidovre. | Albertsen lab, Department of Chemistry and Bioscience, Aalborg University, Denmark | Rasmus Kirkegaard |
| EPI_ISL_452101, EPI_ISL_452102 | Department of Virus and Microbiological Special Diagnostics, Statens Serum Institut, Copenhagen, Denmark, Artillerivej 5, 2300 Copenahgen S | Albertsen lab, Department of Chemistry and Bioscience, Aalborg University, Denmark | Rasmus Kirkegaard |
| EPI_ISL_515974 | Department of Virus and Microbiological Special Diagnostics, Statens Serum Institut | Albertsen lab, Department of Chemistry and Bioscience, Aalborg University, Denmark | Danish Corona Danica Consortia |

We gratefully acknowledge the following Authors from the Originating laboratories responsible for obtaining the specimens, as well as the Submitting laboratories where the genome data were generated and shared via GISAID, on which this research is based.

All Submitters of data may be contacted directly via www.gisaid.org

Authors are sorted alphabetically.

| Accession ID | Originating Laboratory | Submitting Laboratory | Authors |
|---|---|---|---|
| EPI_ISL_1025908, EPI_ISL_1025909, EPI_ISL_1025910, EPI_ISL_1025911, EPI_ISL_1025912, EPI_ISL_1025913, EPI_ISL_1025914, EPI_ISL_1025915, EPI_ISL_1025916, EPI_ISL_1025917, EPI_ISL_1025918, EPI_ISL_1025919, EPI_ISL_1025920, EPI_ISL_1025921, EPI_ISL_1025922, EPI_ISL_1025923, EPI_ISL_1025924, EPI_ISL_1025925, EPI_ISL_1025926, EPI_ISL_1025927, EPI_ISL_1025928, EPI_ISL_1025929, EPI_ISL_1025930, EPI_ISL_1025931, EPI_ISL_1025932, EPI_ISL_1025933, EPI_ISL_1025934, EPI_ISL_1025935, EPI_ISL_1025936, EPI_ISL_1025937, EPI_ISL_1025938, EPI_ISL_1025939, EPI_ISL_1025940, EPI_ISL_1025941, EPI_ISL_1025942, EPI_ISL_1025943, EPI_ISL_1025944, EPI_ISL_1025945, EPI_ISL_1025946, EPI_ISL_1025947, EPI_ISL_1025948, EPI_ISL_1025949, EPI_ISL_1025950, EPI_ISL_1025951, EPI_ISL_1025952, EPI_ISL_1025953, EPI_ISL_1025954, EPI_ISL_1025955, EPI_ISL_1025956, EPI_ISL_1025957, EPI_ISL_1025958, EPI_ISL_1025959, EPI_ISL_1025960, EPI_ISL_1025961, EPI_ISL_1025962, EPI_ISL_1065096, EPI_ISL_1065097, EPI_ISL_1065099, EPI_ISL_1065103, EPI_ISL_1065189, EPI_ISL_1065275, EPI_ISL_1065294, EPI_ISL_1065312, EPI_ISL_1065370, EPI_ISL_1065437, EPI_ISL_1065587, EPI_ISL_1065603, EPI_ISL_1065614, EPI_ISL_1065662, EPI_ISL_1065709, EPI_ISL_1065716, EPI_ISL_1065721, EPI_ISL_1065764, EPI_ISL_1065786, EPI_ISL_1065801, EPI_ISL_1065886, EPI_ISL_1065939, EPI_ISL_1066047, EPI_ISL_1066147, EPI_ISL_1066162, EPI_ISL_1066181, EPI_ISL_1066231, EPI_ISL_1066241, EPI_ISL_1066255, EPI_ISL_1066265, EPI_ISL_1066295, EPI_ISL_1066303, EPI_ISL_1066413, EPI_ISL_1066427, EPI_ISL_1066450, EPI_ISL_1066454, EPI_ISL_1066504, EPI_ISL_1066541, EPI_ISL_1066560, EPI_ISL_1066575, EPI_ISL_1066577, EPI_ISL_1066678, EPI_ISL_1066715, EPI_ISL_1066717, EPI_ISL_1066836, EPI_ISL_1066879, EPI_ISL_1066880, EPI_ISL_1066881, EPI_ISL_1066882, EPI_ISL_1066933, EPI_ISL_1066977, EPI_ISL_1067026, EPI_ISL_1067083, EPI_ISL_1067087, EPI_ISL_1067111, EPI_ISL_1067180, EPI_ISL_1067182, EPI_ISL_1067191, EPI_ISL_1067206, EPI_ISL_1067215, EPI_ISL_1067222, EPI_ISL_1067288, EPI_ISL_1067293, EPI_ISL_1067316, EPI_ISL_1067320, EPI_ISL_1067364, EPI_ISL_1067402, EPI_ISL_1067409, EPI_ISL_1067414, EPI_ISL_1067433, EPI_ISL_1067448, EPI_ISL_1067487, EPI_ISL_1067489, EPI_ISL_1067504, EPI_ISL_1067518, EPI_ISL_1067524, EPI_ISL_1067535, EPI_ISL_1067571 | | | |
| see above | Department of Virus and Microbiological Special Diagnostics, Statens Serum Institut, Copenhagen, Denmark | Aalborg University | Danish Covid-19 Genome Consortium |
| EPI_ISL_625695, EPI_ISL_625698, EPI_ISL_625706, EPI_ISL_625709, EPI_ISL_625742, EPI_ISL_625822, EPI_ISL_625823, EPI_ISL_625864, EPI_ISL_625883, EPI_ISL_625890, EPI_ISL_626124, EPI_ISL_626127, EPI_ISL_626129, EPI_ISL_626132, EPI_ISL_626133, EPI_ISL_626134, EPI_ISL_626137, EPI_ISL_626138, EPI_ISL_626139, EPI_ISL_626144, EPI_ISL_626146, EPI_ISL_626148, EPI_ISL_626149, EPI_ISL_626157, EPI_ISL_626162, EPI_ISL_626164 | | | |
| see above | Department of Virus and Microbiological Special Diagnostics, Statens Serum Institut, Denmark | Albertsen lab, Department of Chemistry and Bioscience, Aalborg University, Denmark | Danish Covid-19 Genome Consortia |
| EPI_ISL_641514, EPI_ISL_641515, EPI_ISL_641516 | Department of Virus and Microbiological Special Diagnostics, Statens Serum Institut, Copenhagen, Denmark | Albertsen lab, Department of Chemistry and Bioscience, Aalborg University, Denmark | Thomas Bruun Rasmussen, Jannik Fonager, Morten Rasmussen |
| EPI_ISL_668458, EPI_ISL_668459, EPI_ISL_668460, EPI_ISL_668461, EPI_ISL_668462, EPI_ISL_668463, EPI_ISL_668464, EPI_ISL_668465, EPI_ISL_668466, EPI_ISL_668467, EPI_ISL_668468, EPI_ISL_668469, EPI_ISL_668470, EPI_ISL_668471, EPI_ISL_668472, EPI_ISL_668473, EPI_ISL_668474, EPI_ISL_668475, EPI_ISL_668476, EPI_ISL_668477, EPI_ISL_668478, EPI_ISL_668479, EPI_ISL_668480, EPI_ISL_668481, EPI_ISL_668482, EPI_ISL_668483, EPI_ISL_668484, EPI_ISL_668485, EPI_ISL_668486, EPI_ISL_668487, EPI_ISL_668488, EPI_ISL_668489, EPI_ISL_668490, EPI_ISL_668491, EPI_ISL_668492, EPI_ISL_668493, EPI_ISL_668494, EPI_ISL_668495, EPI_ISL_668496, EPI_ISL_668497, EPI_ISL_668498, EPI_ISL_668499, EPI_ISL_668500, EPI_ISL_668501, EPI_ISL_668502, EPI_ISL_668503, EPI_ISL_668504, EPI_ISL_668505, EPI_ISL_668506, EPI_ISL_668507, EPI_ISL_668508, EPI_ISL_668509, EPI_ISL_668510, EPI_ISL_668511, EPI_ISL_668512, EPI_ISL_668513, EPI_ISL_668514, EPI_ISL_668515, EPI_ISL_668516, EPI_ISL_668517, EPI_ISL_668518, EPI_ISL_668519, EPI_ISL_668520, EPI_ISL_668521, EPI_ISL_668522, EPI_ISL_668523, EPI_ISL_668524, EPI_ISL_668525, EPI_ISL_668526, EPI_ISL_668527, EPI_ISL_668528, EPI_ISL_668529, EPI_ISL_668530, EPI_ISL_668531, EPI_ISL_668532, EPI_ISL_668533, EPI_ISL_668534, EPI_ISL_668535, EPI_ISL_668536, EPI_ISL_668537, EPI_ISL_668538, EPI_ISL_668539, EPI_ISL_668540, EPI_ISL_668541, EPI_ISL_668542, EPI_ISL_668543, EPI_ISL_668544, EPI_ISL_668545, EPI_ISL_668546, EPI_ISL_668547, EPI_ISL_668548, EPI_ISL_668549, EPI_ISL_668550, EPI_ISL_668551, EPI_ISL_668552, EPI_ISL_668553, EPI_ISL_668554, EPI_ISL_668555, EPI_ISL_668556, EPI_ISL_668557, EPI_ISL_668558, EPI_ISL_668559, EPI_ISL_668560, EPI_ISL_668561, EPI_ISL_668562, EPI_ISL_668563, EPI_ISL_668564, EPI_ISL_668565, EPI_ISL_668566, EPI_ISL_668567, EPI_ISL_668568, EPI_ISL_668569, EPI_ISL_668570, EPI_ISL_668571, EPI_ISL_668572, EPI_ISL_668573, EPI_ISL_668574, EPI_ISL_668575, EPI_ISL_668576, EPI_ISL_668577, EPI_ISL_668578, EPI_ISL_668579, EPI_ISL_668580, EPI_ISL_668581, EPI_ISL_668582, EPI_ISL_668583, EPI_ISL_668584, EPI_ISL_668585, EPI_ISL_668586, EPI_ISL_668587, EPI_ISL_668588, EPI_ISL_668589, EPI_ISL_668590, EPI_ISL_668591, EPI_ISL_668592, EPI_ISL_668593, EPI_ISL_668594, EPI_ISL_668595, EPI_ISL_668596, EPI_ISL_668597, EPI_ISL_668598, EPI_ISL_668599, EPI_ISL_668600, EPI_ISL_668601, EPI_ISL_668602, EPI_ISL_668603, EPI_ISL_668604, EPI_ISL_668605, EPI_ISL_668606, EPI_ISL_668607, EPI_ISL_668608, EPI_ISL_668609, EPI_ISL_668610, EPI_ISL_668611, EPI_ISL_668612, EPI_ISL_668613, EPI_ISL_668614, EPI_ISL_668615, EPI_ISL_668616, EPI_ISL_668617, EPI_ISL_668618, EPI_ISL_668619, EPI_ISL_668620, EPI_ISL_668621, EPI_ISL_668622, EPI_ISL_668623, EPI_ISL_668624, EPI_ISL_668625, EPI_ISL_668626, EPI_ISL_668627, EPI_ISL_668628, EPI_ISL_668629, EPI_ISL_668630, EPI_ISL_668631, EPI_ISL_668632, EPI_ISL_668633, EPI_ISL_668634, EPI_ISL_668635, EPI_ISL_668636, EPI_ISL_668637, EPI_ISL_668638, EPI_ISL_668639, EPI_ISL_668640, EPI_ISL_668641, EPI_ISL_668642, EPI_ISL_668643, EPI_ISL_668644, EPI_ISL_668645, EPI_ISL_668646, EPI_ISL_668647, EPI_ISL_668648, EPI_ISL_668649, EPI_ISL_668650, EPI_ISL_668651, EPI_ISL_668652, EPI_ISL_668653, EPI_ISL_668654, EPI_ISL_668655, EPI_ISL_668656, EPI_ISL_668657, EPI_ISL_668658, EPI_ISL_668659, EPI_ISL_668660, EPI_ISL_668661, EPI_ISL_668662, EPI_ISL_668663, EPI_ISL_668664, EPI_ISL_668665, EPI_ISL_668666, EPI_ISL_668667, EPI_ISL_668668, EPI_ISL_668669, EPI_ISL_668670, EPI_ISL_668671, EPI_ISL_668672, EPI_ISL_668673, EPI_ISL_668674, EPI_ISL_668675, EPI_ISL_668676, EPI_ISL_668677, EPI_ISL_668678, EPI_ISL_668679, EPI_ISL_668680, EPI_ISL_668681, EPI_ISL_668682, EPI_ISL_668683, EPI_ISL_668684, EPI_ISL_668685, EPI_ISL_668686, EPI_ISL_668687, EPI_ISL_668688, EPI_ISL_668689, EPI_ISL_668690, EPI_ISL_668691, EPI_ISL_668692, EPI_ISL_668693, EPI_ISL_668694, EPI_ISL_668695, EPI_ISL_668696, EPI_ISL_668697, EPI_ISL_668698, EPI_ISL_668699, EPI_ISL_668700, EPI_ISL_668701, EPI_ISL_668702, EPI_ISL_668703, EPI_ISL_668704, EPI_ISL_668705, EPI_ISL_668706, EPI_ISL_668707, EPI_ISL_668708, EPI_ISL_668709, EPI_ISL_668710, EPI_ISL_668711, EPI_ISL_668712, EPI_ISL_668713, EPI_ISL_668714, EPI_ISL_668715, EPI_ISL_668716, EPI_ISL_668717, EPI_ISL_668718, EPI_ISL_668719, EPI_ISL_668720, EPI_ISL_668721, EPI_ISL_668722, EPI_ISL_668723, EPI_ISL_668724, EPI_ISL_668725, EPI_ISL_668726, EPI_ISL_668727, EPI_ISL_668728, EPI_ISL_668729, EPI_ISL_668730, EPI_ISL_668731, EPI_ISL_668732, EPI_ISL_668733, EPI_ISL_668734, EPI_ISL_668735, EPI_ISL_668736, EPI_ISL_668737, EPI_ISL_668738, EPI_ISL_668739, EPI_ISL_668740, EPI_ISL_668741, EPI_ISL_668742, EPI_ISL_668743, EPI_ISL_668744, EPI_ISL_668745, EPI_ISL_668746, EPI_ISL_668747, EPI_ISL_668748, EPI_ISL_668749, EPI_ISL_668750, EPI_ISL_668751, EPI_ISL_668752, EPI_ISL_668753, EPI_ISL_668754, EPI_ISL_668755, EPI_ISL_668756, EPI_ISL_668757, EPI_ISL_668758, EPI_ISL_668759, EPI_ISL_668760, EPI_ISL_668761, EPI_ISL_668762, EPI_ISL_668763, EPI_ISL_668764, EPI_ISL_668765, EPI_ISL_668766, EPI_ISL_668767, EPI_ISL_668768, EPI_ISL_668769, EPI_ISL_668770, EPI_ISL_668771, EPI_ISL_668772, EPI_ISL_668773, EPI_ISL_668774, EPI_ISL_668775, EPI_ISL_668776, EPI_ISL_668777, EPI_ISL_668778, EPI_ISL_668779, EPI_ISL_668780, EPI_ISL_668781, EPI_ISL_668782, EPI_ISL_668783, EPI_ISL_668784, EPI_ISL_668785, EPI_ISL_668786, EPI_ISL_668787, EPI_ISL_668788, EPI_ISL_668789, EPI_ISL_668790, EPI_ISL_668791, EPI_ISL_668792, EPI_ISL_668793, EPI_ISL_668794, EPI_ISL_668795, EPI_ISL_668796, EPI_ISL_668797, EPI_ISL_668798, EPI_ISL_668799, EPI_ISL_668800, EPI_ISL_668801, EPI_ISL_668802, EPI_ISL_668803, EPI_ISL_668804, EPI_ISL_668805, EPI_ISL_668806, EPI_ISL_668807, EPI_ISL_668808, EPI_ISL_668809, EPI_ISL_668810, EPI_ISL_668811, EPI_ISL_668812, EPI_ISL_668813, EPI_ISL_668814, EPI_ISL_668815, EPI_ISL_668816, EPI_ISL_668817, EPI_ISL_668818, EPI_ISL_668819, EPI_ISL_668820, EPI_ISL_668821, EPI_ISL_668822, EPI_ISL_668823, EPI_ISL_668824, EPI_ISL_668825, EPI_ISL_668826, EPI_ISL_668827, EPI_ISL_668828, EPI_ISL_668829, EPI_ISL_668830, EPI_ISL_668831, EPI_ISL_668832, EPI_ISL_668833, EPI_ISL_668834, EPI_ISL_668835, EPI_ISL_668836, EPI_ISL_668837, EPI_ISL_668838, EPI_ISL_668839, EPI_ISL_668840, EPI_ISL_668841, EPI_ISL_668842, EPI_ISL_668843, EPI_ISL_668844, EPI_ISL_668845, EPI_ISL_668846, EPI_ISL_668847, EPI_ISL_668848, EPI_ISL_668849, EPI_ISL_668850, EPI_ISL_668851, EPI_ISL_668852, EPI_ISL_668853, EPI_ISL_668854, EPI_ISL_668855, EPI_ISL_668856, EPI_ISL_668857, EPI_ISL_668858, EPI_ISL_668859, EPI_ISL_668860, EPI_ISL_668861, EPI_ISL_668862, EPI_ISL_668863, EPI_ISL_668864, EPI_ISL_668865, EPI_ISL_668866, EPI_ISL_668867, EPI_ISL_668868, EPI_ISL_668869, EPI_ISL_668870, EPI_ISL_668871, EPI_ISL_668872, EPI_ISL_668873, EPI_ISL_668874, EPI_ISL_668875, EPI_ISL_668876, EPI_ISL_668877, EPI_ISL_668878, EPI_ISL_668879, EPI_ISL_668880, EPI_ISL_668881, EPI_ISL_668882, EPI_ISL_668883, EPI_ISL_668884, EPI_ISL_668885, EPI_ISL_668886, EPI_ISL_668887, EPI_ISL_668888, EPI_ISL_668889, EPI_ISL_668890, EPI_ISL_668891, EPI_ISL_668892, EPI_ISL_668893, EPI_ISL_668894, EPI_ISL_668895, EPI_ISL_668896, EPI_ISL_668897, EPI_ISL_668898, EPI_ISL_668899, EPI_ISL_668900, EPI_ISL_668901, EPI_ISL_668902, EPI_ISL_668903, EPI_ISL_668904, EPI_ISL_668905, EPI_ISL_668906, EPI_ISL_668907, EPI_ISL_668908, EPI_ISL_668909, EPI_ISL_668910, EPI_ISL_668911, EPI_ISL_668912, EPI_ISL_668913, EPI_ISL_668914, EPI_ISL_668915, EPI_ISL_668916, EPI_ISL_668917, EPI_ISL_668918, EPI_ISL_668919, EPI_ISL_668920, EPI_ISL_668921, EPI_ISL_668922, EPI_ISL_668923, EPI_ISL_668924, EPI_ISL_668925, EPI_ISL_668926, EPI_ISL_668927, EPI_ISL_668928, EPI_ISL_668929, EPI_ISL_668930, EPI_ISL_668931, EPI_ISL_668932, EPI_ISL_668933, EPI_ISL_668934, EPI_ISL_668935, EPI_ISL_668936, EPI_ISL_668937, EPI_ISL_668938, EPI_ISL_668939, EPI_ISL_668940, EPI_ISL_668941, EPI_ISL_668942, EPI_ISL_668943, EPI_ISL_668944, EPI_ISL_668945, EPI_ISL_668946, EPI_ISL_668947, EPI_ISL_668948, EPI_ISL_668949, EPI_ISL_668950, EPI_ISL_668951, EPI_ISL_668952, EPI_ISL_668953, EPI_ISL_668954, EPI_ISL_668955, EPI_ISL_668956, EPI_ISL_668957, EPI_ISL_668958, EPI_ISL_668959, EPI_ISL_668960, EPI_ISL_668961, EPI_ISL_668962, EPI_ISL_668963, EPI_ISL_668964, EPI_ISL_668965, EPI_ISL_668966, EPI_ISL_668967, EPI_ISL_668968, EPI_ISL_668969, EPI_ISL_668970, EPI_ISL_668971, EPI_ISL_668972, EPI_ISL_668973, EPI_ISL_668974, EPI_ISL_668975, EPI_ISL_668976, EPI_ISL_668977, EPI_ISL_668978, EPI_ISL_668979, EPI_ISL_668980, EPI_ISL_668981, EPI_ISL_668982, EPI_ISL_668983, EPI_ISL_668984, EPI_ISL_668985, EPI_ISL_668986, EPI_ISL_668987, EPI_ISL_668988, EPI_ISL_668989, EPI_ISL_668990, EPI_ISL_668991, EPI_ISL_668992, EPI_ISL_668993, EPI_ISL_668994, EPI_ISL_668995, EPI_ISL_668996, EPI_ISL_668997, EPI_ISL_668998, EPI_ISL_668999, EPI_ISL_669000, EPI_ISL_669001, EPI_ISL_669002, EPI_ISL_669003, EPI_ISL_669004, EPI_ISL_669005, EPI_ISL_669006, EPI_ISL_669007, EPI_ISL_669008, EPI_ISL_669009, EPI_ISL_669010, EPI_ISL_669011, EPI_ISL_669012, EPI_ISL_669013, EPI_ISL_669014, EPI_ISL_669015, EPI_ISL_669016, EPI_ISL_669017, EPI_ISL_669018, EPI_ISL_669019, EPI_ISL_669020, EPI_ISL_669021, EPI_ISL_669022, EPI_ISL_669023, EPI_ISL_669024, EPI_ISL_669025, EPI_ISL_669026, EPI_ISL_669027, EPI_ISL_669028, EPI_ISL_669029, EPI_ISL_669030, EPI_ISL_669031, EPI_ISL_669032, EPI_ISL_669033, EPI_ISL_669034, EPI_ISL_669035, EPI_ISL_669036, EPI_ISL_669037, EPI_ISL_669038, EPI_ISL_669039, EPI_ISL_669040, EPI_ISL_669041, EPI_ISL_669042, EPI_ISL_669043, EPI_ISL_669044, EPI_ISL_669045, EPI_ISL_669046, EPI_ISL_669047, EPI_ISL_669048, EPI_ISL_669049, EPI_ISL_669050, EPI_ISL_669051, EPI_ISL_669052, EPI_ISL_669053, EPI_ISL_669054, EPI_ISL_669055, EPI_ISL_669056, EPI_ISL_669057, EPI_ISL_669058, EPI_ISL_669059, EPI_ISL_669060, EPI_ISL_669061, EPI_ISL_669062, EPI_ISL_669063, EPI_ISL_669064, EPI_ISL_669065, EPI_ISL_669066, EPI_ISL_669067, EPI_ISL_669068, EPI_ISL_669069, EPI_ISL_669070, EPI_ISL_669071, EPI_ISL_669072, EPI_ISL_669073, EPI_ISL_669074, EPI_ISL_669075, EPI_ISL_669076, EPI_ISL_669077, EPI_ISL_669078, EPI_ISL_669079, EPI_ISL_669080, EPI_ISL_669081, EPI_ISL_669082, EPI_ISL_669083, EPI_ISL_669084, EPI_ISL_669085, EPI_ISL_669086, EPI_ISL_669087, EPI_ISL_669088, EPI_ISL_669089, EPI_ISL_669090, EPI_ISL_669091, EPI_ISL_669092, EPI_ISL_669093, EPI_ISL_669094, EPI_ISL_669095, EPI_ISL_669096, EPI_ISL_669097, EPI_ISL_669098, EPI_ISL_669099, EPI_ISL_669100, EPI_ISL_669101, EPI_ISL_669102, EPI_ISL_669103, EPI_ISL_669104, EPI_ISL_669105, EPI_ISL_669106, EPI_ISL_669107, EPI_ISL_669108, EPI_ISL_669109, EPI_ISL_669110, EPI_ISL_669111, EPI_ISL_669112, EPI_ISL_669113, EPI_ISL_669114, EPI_ISL_669115, EPI_ISL_669116, EPI_ISL_669117, EPI_ISL_669118, EPI_ISL_669119, EPI_ISL_669120, EPI_ISL_669121, EPI_ISL_669122, EPI_ISL_669123, EPI_ISL_669124, EPI_ISL_669125, EPI_ISL_669126, EPI_ISL_669127, EPI_ISL_669128, EPI_ISL_669129, EPI_ISL_669130, EPI_ISL_669131, EPI_ISL_669132, EPI_ISL_669133, EPI_ISL_669134, EPI_ISL_669135, EPI_ISL_669136, EPI_ISL_669137, EPI_ISL_669138, EPI_ISL_669139, EPI_ISL_669140, EPI_ISL_669141, EPI_ISL_669142, EPI_ISL_669143, EPI_ISL_669144, EPI_ISL_669145, EPI_ISL_669146, EPI_ISL_669147, EPI_ISL_669148, EPI_ISL_669149, EPI_ISL_669150, EPI_ISL_669151, EPI_ISL_669152, EPI_ISL_669153, EPI_ISL_669154, EPI_ISL_669155, EPI_ISL_669156, EPI_ISL_669157, EPI_ISL_669158, EPI_ISL_669159, EPI_ISL_669160, EPI_ISL_669161, EPI_ISL_669162, EPI_ISL_669163, EPI_ISL_669164, EPI_ISL_669165, EPI_ISL_669166, EPI_ISL_669167, EPI_ISL_669168, EPI_ISL_669169, EPI_ISL_669170, EPI_ISL_669171, EPI_ISL_669172, EPI_ISL_669173, EPI_ISL_669174, EPI_ISL_669175, EPI_ISL_669176, EPI_ISL_669177, EPI_ISL_669178, EPI_ISL_669179, EPI_ISL_669180, EPI_ISL_669181, EPI_ISL_669182, EPI_ISL_669183, EPI_ISL_669184, EPI_ISL_669185, EPI_ISL_669186, EPI_ISL_669187, EPI_ISL_669188, EPI_ISL_669189, EPI_ISL_669190, EPI_ISL_669191, EPI_ISL_669192, EPI_ISL_669193, EPI_ISL_669194, EPI_ISL_669195, EPI_ISL_669196, EPI_ISL_669197, EPI_ISL_669198, EPI_ISL_669199, EPI_ISL_669200, EPI_ISL_669201, EPI_ISL_669202, EPI_ISL_669203, EPI_ISL_669204, EPI_ISL_669205, EPI_ISL_669206, EPI_ISL_669207, EPI_ISL_669208, EPI_ISL_669209, EPI_ISL_669210, EPI_ISL_669211, EPI_ISL_669212, EPI_ISL_669213, EPI_ISL_669214, EPI_ISL_669215, EPI_ISL_669216, EPI_ISL_669217, EPI_ISL_669218, EPI_ISL_669219, EPI_ISL_669220, EPI_ISL_669221, EPI_ISL_669222, EPI_ISL_669223, EPI_ISL_669224, EPI_ISL_669225, EPI_ISL_669226, EPI_ISL_669227, EPI_ISL_669228, EPI_ISL_669229, EPI_ISL_669230, EPI_ISL_669231, EPI_ISL_669232, EPI_ISL_669233, EPI_ISL_669234, EPI_ISL_669235, EPI_ISL_669236, EPI_ISL_669237, EPI_ISL_669238, EPI_ISL_669239, EPI_ISL_669240, EPI_ISL_669241, EPI_ISL_669242, EPI_ISL_669243, EPI_ISL_669244, EPI_ISL_669245, EPI_ISL_669246, EPI_ISL_669247, EPI_ISL_669248, EPI_ISL_669249, EPI_ISL_669250, EPI_ISL_669251, EPI_ISL_669252, EPI_ISL_669253, EPI_ISL_669254, EPI_ISL_669255, EPI_ISL_669256, EPI_ISL_669257, EPI_ISL_669258, EPI_ISL_669259, EPI_ISL_669260, EPI_ISL_669261, EPI_ISL_669262, EPI_ISL_669263, EPI_ISL_669264, EPI_ISL_669265, EPI_ISL_669266, EPI_ISL_669267, EPI_ISL_669268, EPI_ISL_669269, EPI_ISL_669270, EPI_ISL_669271, EPI_ISL_669272, EPI_ISL_669273, EPI_ISL_669274, EPI_ISL_669275, EPI_ISL_669276, EPI_ISL_669277, EPI_ISL_669278, EPI_ISL_669279, EPI_ISL_669280, EPI_ISL_669281, EPI_ISL_669282, EPI_ISL_669283, EPI_ISL_669284, EPI_ISL_669285, EPI_ISL_669286, EPI_ISL_669287, EPI_ISL_669288, EPI_ISL_669289, EPI_ISL_669290, EPI_ISL_669291, EPI_ISL_669292, EPI_ISL_669293, EPI_ISL_669294, EPI_ISL_669295, EPI_ISL_669296, EPI_ISL_669297, EPI_ISL_669298, EPI_ISL_669299, EPI_ISL_669300, EPI_ISL_669301, EPI_ISL_669302, EPI_ISL_669303, EPI_ISL_669304, EPI_ISL_669305, EPI_ISL_669306, EPI_ISL_669307, EPI_ISL_669308, EPI_ISL_669309, EPI_ISL_669310, EPI_ISL_669311, EPI_ISL_669312, EPI_ISL_669313, EPI_ISL_669314, EPI_ISL_669315, EPI_ISL_669316, EPI_ISL_669317, EPI_ISL_669318, EPI_ISL_669319, EPI_ISL_669320, EPI_ISL_669321, EPI_ISL_669322, EPI_ISL_669323, EPI_ISL_669324, EPI_ISL_669325, EPI_ISL_669326, EPI_ISL_669327, EPI_ISL_669328, EPI_ISL_669329, EPI_ISL_669330, EPI_ISL_669331, EPI_ISL_669332, EPI_ISL_669333, EPI_ISL_669334, EPI_ISL_669335, EPI_ISL_669336, EPI_ISL_669337, EPI_ISL_669338, EPI_ISL_669339, EPI_ISL_669340, EPI_ISL_669341, EPI_ISL_669342, EPI_ISL_669343, EPI_ISL_669344, EPI_ISL_669345, EPI_ISL_669346, EPI_ISL_669347, EPI_ISL_669348, EPI_ISL_669349, EPI_ISL_669350, EPI_ISL_669351, EPI_ISL_669352, EPI_ISL_669353, EPI_ISL_669354, EPI_ISL_669355, EPI_ISL_669356, EPI_ISL_669357, EPI_ISL_669358, EPI_ISL_669359, EPI_ISL_669360, EPI_ISL_669361, EPI_ISL_669362, EPI_ISL_669363, EPI_ISL_669364, EPI_ISL_669365, EPI_ISL_669366, EPI_ISL_669367, EPI_ISL_669368, EPI_ISL_669369, EPI_ISL_669370, EPI_ISL_669371, EPI_ISL_669372, EPI_ISL_669373, EPI_ISL_669374, EPI_ISL_669375, EPI_ISL_669376, EPI_ISL_669377, EPI_ISL_669378, EPI_ISL_669379, EPI_ISL_669380, EPI_ISL_669381, EPI_ISL_669382, EPI_ISL_669383, EPI_ISL_669384, EPI_ISL_669385, EPI_ISL_669386, EPI_ISL_669387, EPI_ISL_669388, EPI_ISL_669389, EPI_ISL_669390, EPI_ISL_669391, EPI_ISL_669392, EPI_ISL_669393, EPI_ISL_669394, EPI_ISL_669395, EPI_ISL_669396, EPI_ISL_669397, EPI_ISL_669398, EPI_ISL_669399, EPI_ISL_669400, EPI_ISL_669401, EPI_ISL_669402, EPI_ISL_669403, EPI_ISL_669404, EPI_ISL_669405, EPI_ISL_669406, EPI_ISL_669407, EPI_ISL_669408, EPI_ISL_669409, EPI_ISL_669410, EPI_ISL_669411, EPI_ISL_669412, EPI_ISL_669413, EPI_ISL_669414, EPI_ISL_669415, EPI_ISL_669416, EPI_ISL_669417, EPI_ISL_669418, EPI_ISL_669419, EPI_ISL_669420, EPI_ISL_669421, EPI_ISL_669422, EPI_ISL_669423, EPI_ISL_669424, EPI_ISL_669425, EPI_ISL_669426, EPI_ISL_669427, EPI_ISL_669428, EPI_ISL_669429, EPI_ISL_669430, EPI_ISL_669431, EPI_ISL_669432, EPI_ISL_669433, EPI_ISL_669434, EPI_ISL_669435, EPI_ISL_669436, EPI_ISL_669437, EPI_ISL_669438, EPI_ISL_669439, EPI_ISL_669440, EPI_ISL_669441, EPI_ISL_669442, EPI_ISL_669443, EPI_ISL_669444, EPI_ISL_669445, EPI_ISL_669446, EPI_ISL_669447, EPI_ISL_669448, EPI_ISL_669449, EPI_ISL_669450, EPI_ISL_669451, EPI_ISL_669452, EPI_ISL_669453, EPI_ISL_669454, EPI_ISL_669455, EPI_ISL_669456, EPI_ISL_669457, EPI_ISL_669458, EPI_ISL_669459, EPI_ISL_669460, EPI_ISL_669461, EPI_ISL_669462, EPI_ISL_669463, EPI_ISL_669464, EPI_ISL_669465, EPI_ISL_669466, EPI_ISL_669467, EPI_ISL_669468, EPI_ISL_669469, EPI_ISL_669470, EPI_ISL_669471, EPI_ISL_669472, EPI_ISL_669473, EPI_ISL_669474, EPI_ISL_669475, EPI_ISL_669476, EPI_ISL_669477, EPI_ISL_669478, EPI_ISL_669479, EPI_ISL_669480, EPI_ISL_669481, EPI_ISL_669482, EPI_ISL_669483, | | | |

We gratefully acknowledge the following Authors from the Originating laboratories responsible for obtaining the specimens, as well as the Submitting laboratories where the genome data were generated and shared via GISAID, on which this research is based.

All Submitters of data may be contacted directly via www.gisaid.org

Authors are sorted alphabetically.

| Accession ID | Originating Laboratory | Submitting Laboratory | Authors |
|---|---|---|---|
| EPI_ISL_1025670, EPI_ISL_1025671, EPI_ISL_1025672, EPI_ISL_1025673, EPI_ISL_1025674, EPI_ISL_1025675, EPI_ISL_1025676, EPI_ISL_1025677, EPI_ISL_1025678, EPI_ISL_1025679, EPI_ISL_1025680, EPI_ISL_1025681, EPI_ISL_1025682, EPI_ISL_1025683, EPI_ISL_1025684, EPI_ISL_1025685, EPI_ISL_1025686, EPI_ISL_1025687, EPI_ISL_1025688, EPI_ISL_1025689, EPI_ISL_1025690, EPI_ISL_1025691, EPI_ISL_1025692, EPI_ISL_1025693, EPI_ISL_1025694, EPI_ISL_1025695, EPI_ISL_1025696, EPI_ISL_1025697, EPI_ISL_1025698, EPI_ISL_1025699, EPI_ISL_1025700, EPI_ISL_1025701, EPI_ISL_1025702, EPI_ISL_1025703, EPI_ISL_1025704, EPI_ISL_1025705, EPI_ISL_1025706, EPI_ISL_1025707, EPI_ISL_1025708, EPI_ISL_1025709, EPI_ISL_1025710, EPI_ISL_1025711, EPI_ISL_1025712, EPI_ISL_1025713, EPI_ISL_1025714, EPI_ISL_1025715, EPI_ISL_1025716, EPI_ISL_1025717, EPI_ISL_1025718, EPI_ISL_1025719, EPI_ISL_1025720, EPI_ISL_1025721, EPI_ISL_1025722, EPI_ISL_1025723, EPI_ISL_1025724, EPI_ISL_1025725, EPI_ISL_1025726, EPI_ISL_1025727, EPI_ISL_1025728, EPI_ISL_1025729, EPI_ISL_1025730, EPI_ISL_1025731, EPI_ISL_1025732, EPI_ISL_1025733, EPI_ISL_1025734, EPI_ISL_1025735, EPI_ISL_1025736, EPI_ISL_1025737, EPI_ISL_1025738, EPI_ISL_1025739, EPI_ISL_1025740, EPI_ISL_1025741, EPI_ISL_1025742, EPI_ISL_1025743, EPI_ISL_1025744, EPI_ISL_1025745, EPI_ISL_1025746, EPI_ISL_1025747, EPI_ISL_1025748, EPI_ISL_1025749, EPI_ISL_1025750, EPI_ISL_1025751, EPI_ISL_1025752, EPI_ISL_1025753, EPI_ISL_1025754, EPI_ISL_1025755, EPI_ISL_1025756, EPI_ISL_1025757, EPI_ISL_1025758, EPI_ISL_1025759, EPI_ISL_1025760, EPI_ISL_1025761, EPI_ISL_1025762, EPI_ISL_1025763, EPI_ISL_1025764, EPI_ISL_1025765, EPI_ISL_1025766, EPI_ISL_1025767, EPI_ISL_1025768, EPI_ISL_1025769, EPI_ISL_1025770, EPI_ISL_1025771, EPI_ISL_1025772, EPI_ISL_1025773, EPI_ISL_1025774, EPI_ISL_1025775, EPI_ISL_1025776, EPI_ISL_1025777, EPI_ISL_1025778, EPI_ISL_1025779, EPI_ISL_1025780, EPI_ISL_1025781, EPI_ISL_1025782, EPI_ISL_1025783, EPI_ISL_1025784, EPI_ISL_1025785, EPI_ISL_1025786, EPI_ISL_1025787, EPI_ISL_1025788, EPI_ISL_1025789, EPI_ISL_1025790, EPI_ISL_1025791, EPI_ISL_1025792, EPI_ISL_1025793, EPI_ISL_1025794, EPI_ISL_1025795, EPI_ISL_1025796, EPI_ISL_1025797, EPI_ISL_1025798, EPI_ISL_1025799, EPI_ISL_1025800, EPI_ISL_1025801, EPI_ISL_1025802, EPI_ISL_1025803, EPI_ISL_1025804, EPI_ISL_1025805, EPI_ISL_1025806, EPI_ISL_1025807, EPI_ISL_1025808, EPI_ISL_1025809, EPI_ISL_1025810, EPI_ISL_1025811, EPI_ISL_1025812, EPI_ISL_1025813, EPI_ISL_1025814, EPI_ISL_1025815, EPI_ISL_1025816, EPI_ISL_1025817, EPI_ISL_1025818, EPI_ISL_1025819, EPI_ISL_1025820, EPI_ISL_1025821, EPI_ISL_1025822, EPI_ISL_1025823, EPI_ISL_1025824, EPI_ISL_1025825, EPI_ISL_1025826, EPI_ISL_1025827, EPI_ISL_1025828, EPI_ISL_1025829, EPI_ISL_1025830, EPI_ISL_1025831, EPI_ISL_1025832, EPI_ISL_1025833, EPI_ISL_1025834, EPI_ISL_1025835, EPI_ISL_1025836, EPI_ISL_1025837, EPI_ISL_1025838, EPI_ISL_1025839, EPI_ISL_1025840, EPI_ISL_1025841, EPI_ISL_1025842, EPI_ISL_1025843, EPI_ISL_1025844, EPI_ISL_1025845, EPI_ISL_1025846, EPI_ISL_1025847, EPI_ISL_1025848, EPI_ISL_1025849, EPI_ISL_1025850, EPI_ISL_1025851, EPI_ISL_1025852, EPI_ISL_1025853, EPI_ISL_1025854, EPI_ISL_1025855, EPI_ISL_1025856, EPI_ISL_1025857, EPI_ISL_1025858, EPI_ISL_1025859, EPI_ISL_1025860, EPI_ISL_1025861, EPI_ISL_1025862, EPI_ISL_1025863, EPI_ISL_1025864, EPI_ISL_1025865, EPI_ISL_1025866, EPI_ISL_1025867, EPI_ISL_1025868, EPI_ISL_1025869, EPI_ISL_1025870, EPI_ISL_1025871, EPI_ISL_1025872, EPI_ISL_1025873, EPI_ISL_1025874, EPI_ISL_1025875, EPI_ISL_1025876, EPI_ISL_1025877, EPI_ISL_1025878, EPI_ISL_1025879, EPI_ISL_1025880, EPI_ISL_1025881, EPI_ISL_1025882, EPI_ISL_1025883, EPI_ISL_1025884, EPI_ISL_1025885, EPI_ISL_1025886, EPI_ISL_1025887, EPI_ISL_1025888, EPI_ISL_1025889, EPI_ISL_1025890, EPI_ISL_1025891, EPI_ISL_1025892, EPI_ISL_1025893, EPI_ISL_1025894, EPI_ISL_1025895, EPI_ISL_1025896, EPI_ISL_1025897, EPI_ISL_1025898, EPI_ISL_1025899, EPI_ISL_1025900, EPI_ISL_1025901, EPI_ISL_1025902, EPI_ISL_1025903, EPI_ISL_1025904, EPI_ISL_1025905, EPI_ISL_1025906, EPI_ISL_1025907, EPI_ISL_1065100, EPI_ISL_1065101, EPI_ISL_1065112, EPI_ISL_1065114, EPI_ISL_1065115, EPI_ISL_1065116, EPI_ISL_1065117, EPI_ISL_1065119, EPI_ISL_1065126, EPI_ISL_1065128, EPI_ISL_1065132, EPI_ISL_1065134, EPI_ISL_1065145, EPI_ISL_1065147, EPI_ISL_1065149, EPI_ISL_1065153, EPI_ISL_1065162, EPI_ISL_1065169, EPI_ISL_1065171, EPI_ISL_1065187, EPI_ISL_1065188, EPI_ISL_1065194, EPI_ISL_1065196, EPI_ISL_1065202, EPI_ISL_1065206, EPI_ISL_1065208, EPI_ISL_1065213, EPI_ISL_1065217, EPI_ISL_1065218, EPI_ISL_1065233, EPI_ISL_1065234, EPI_ISL_1065242, EPI_ISL_1065245, EPI_ISL_1065248, EPI_ISL_1065250, EPI_ISL_1065251, EPI_ISL_1065258, EPI_ISL_1065259, EPI_ISL_1065267, EPI_ISL_1065272, EPI_ISL_1065273, EPI_ISL_1065279, EPI_ISL_1065282, EPI_ISL_1065284, EPI_ISL_1065285, EPI_ISL_1065286, EPI_ISL_1065290, EPI_ISL_1065291, EPI_ISL_1065296, EPI_ISL_1065301, EPI_ISL_1065302, EPI_ISL_1065304, EPI_ISL_1065309, EPI_ISL_1065311, EPI_ISL_1065313, EPI_ISL_1065314, EPI_ISL_1065318, EPI_ISL_1065322, EPI_ISL_1065326, EPI_ISL_1065327, EPI_ISL_1065328, EPI_ISL_1065329, EPI_ISL_1065340, EPI_ISL_1065343, EPI_ISL_1065347, EPI_ISL_1065348, EPI_ISL_1065352, EPI_ISL_1065359, EPI_ISL_1065363, EPI_ISL_1065366, EPI_ISL_1065371, EPI_ISL_1065374, EPI_ISL_1065376, EPI_ISL_1065377, EPI_ISL_1065378, EPI_ISL_1065387, EPI_ISL_1065388, EPI_ISL_1065393, EPI_ISL_1065395, EPI_ISL_1065397, EPI_ISL_1065399, EPI_ISL_1065402, EPI_ISL_1065407, EPI_ISL_1065421, EPI_ISL_1065422, EPI_ISL_1065428, EPI_ISL_1065431, EPI_ISL_1065438, EPI_ISL_1065444, EPI_ISL_1065446, EPI_ISL_1065452, EPI_ISL_1065458, EPI_ISL_1065460, EPI_ISL_1065461, EPI_ISL_1065462, EPI_ISL_1065463, EPI_ISL_1065464, EPI_ISL_1065465, EPI_ISL_1065466, EPI_ISL_1065468, EPI_ISL_1065474, EPI_ISL_1065475, EPI_ISL_1065487, EPI_ISL_1065491, EPI_ISL_1065497, EPI_ISL_1065509, EPI_ISL_1065510, EPI_ISL_1065515, EPI_ISL_1065517, EPI_ISL_1065521, EPI_ISL_1065522, EPI_ISL_1065523, EPI_ISL_1065528, EPI_ISL_1065529, EPI_ISL_1065531, EPI_ISL_1065533, EPI_ISL_1065534, EPI_ISL_1065541, EPI_ISL_1065544, EPI_ISL_1065545, EPI_ISL_1065550, EPI_ISL_1065551, EPI_ISL_1065553, EPI_ISL_1065556, EPI_ISL_1065562, EPI_ISL_1065564, EPI_ISL_1065566, EPI_ISL_1065567, EPI_ISL_1065571, EPI_ISL_1065576, EPI_ISL_1065577, EPI_ISL_1065579, EPI_ISL_1065580, EPI_ISL_1065581, EPI_ISL_1065582, EPI_ISL_1065593, EPI_ISL_1065595, EPI_ISL_1065597, EPI_ISL_1065601, EPI_ISL_1065602, EPI_ISL_1065604, EPI_ISL_1065610, EPI_ISL_1065611, EPI_ISL_1065616, EPI_ISL_1065618, EPI_ISL_1065620, EPI_ISL_1065621, EPI_ISL_1065625, EPI_ISL_1065626, EPI_ISL_1065636, EPI_ISL_1065638, EPI_ISL_1065640, EPI_ISL_1065642, EPI_ISL_1065643, EPI_ISL_1065651, EPI_ISL_1065655, EPI_ISL_1065658, EPI_ISL_1065659, EPI_ISL_1065660, EPI_ISL_1065666, EPI_ISL_1065671, EPI_ISL_1065674, EPI_ISL_1065675, EPI_ISL_1065677, EPI_ISL_1065678, EPI_ISL_1065679, EPI_ISL_1065685, EPI_ISL_1065687, EPI_ISL_1065696, EPI_ISL_1065699, EPI_ISL_1065700, EPI_ISL_1065702, EPI_ISL_1065707, EPI_ISL_1065708, EPI_ISL_1065710, EPI_ISL_1065715, EPI_ISL_1065717, EPI_ISL_1065719, EPI_ISL_1065720, EPI_ISL_1065723, EPI_ISL_1065724, EPI_ISL_1065728, EPI_ISL_1065729, EPI_ISL_1065732, EPI_ISL_1065733, EPI_ISL_1065735, EPI_ISL_1065744, EPI_ISL_1065745, EPI_ISL_1065748, EPI_ISL_1065753, EPI_ISL_1065757, EPI_ISL_1065760, EPI_ISL_1065761, EPI_ISL_1065762, EPI_ISL_1065770, EPI_ISL_1065774, EPI_ISL_1065777, EPI_ISL_1065778, EPI_ISL_1065780, EPI_ISL_1065781, EPI_ISL_1065787, EPI_ISL_1065788, EPI_ISL_1065789, EPI_ISL_1065790, EPI_ISL_1065795, EPI_ISL_1065796, EPI_ISL_1065799, EPI_ISL_1065803, EPI_ISL_1065805, EPI_ISL_1065806, EPI_ISL_1065807, EPI_ISL_1065810, EPI_ISL_1065814, EPI_ISL_1065816, EPI_ISL_1065817, EPI_ISL_1065820, EPI_ISL_1065825, EPI_ISL_1065829, EPI_ISL_1065831, EPI_ISL_1065833, EPI_ISL_1065835, EPI_ISL_1065837, EPI_ISL_1065838, EPI_ISL_1065839, EPI_ISL_1065847, EPI_ISL_1065849, EPI_ISL_1065851, EPI_ISL_1065852, EPI_ISL_1065853, EPI_ISL_1065856, EPI_ISL_1065868, EPI_ISL_1065869, EPI_ISL_1065870, EPI_ISL_1065883, EPI_ISL_1065884, EPI_ISL_1065890, EPI_ISL_1065893, EPI_ISL_1065896, EPI_ISL_1065901, EPI_ISL_1065902, EPI_ISL_1065909, EPI_ISL_1065921, EPI_ISL_1065923, EPI_ISL_1065925, EPI_ISL_1065930, EPI_ISL_1065932, EPI_ISL_1065941, EPI_ISL_1065942, EPI_ISL_1065947, EPI_ISL_1065956, EPI_ISL_1065959, EPI_ISL_1065969, EPI_ISL_1065971, EPI_ISL_1065975, EPI_ISL_1065978, EPI_ISL_1065984, EPI_ISL_1065985, EPI_ISL_1065988, EPI_ISL_1065992, EPI_ISL_1065994, EPI_ISL_1065998, EPI_ISL_1066003, EPI_ISL_1066006, EPI_ISL_1066008, EPI_ISL_1066017, EPI_ISL_1066023, EPI_ISL_1066028, EPI_ISL_1066029, EPI_ISL_1066030, EPI_ISL_1066032, EPI_ISL_1066034, EPI_ISL_1066036, EPI_ISL_1066038, EPI_ISL_1066043, EPI_ISL_1066052, EPI_ISL_1066057, EPI_ISL_1066058, EPI_ISL_1066059, EPI_ISL_1066060, EPI_ISL_1066072, EPI_ISL_1066073, EPI_ISL_1066075, EPI_ISL_1066083, EPI_ISL_1066084, EPI_ISL_1066085, EPI_ISL_1066091, EPI_ISL_1066096, EPI_ISL_1066098, EPI_ISL_1066101, EPI_ISL_1066103, EPI_ISL_1066107, EPI_ISL_1066109, EPI_ISL_1066111, EPI_ISL_1066113, EPI_ISL_1066115, EPI_ISL_1066119, EPI_ISL_1066120, EPI_ISL_1066121, EPI_ISL_1066124, EPI_ISL_1066125, EPI_ISL_1066126, EPI_ISL_1066135, EPI_ISL_1066137, EPI_ISL_1066139, EPI_ISL_1066149, EPI_ISL_1066151, EPI_ISL_1066172, EPI_ISL_1066185, EPI_ISL_1066187, EPI_ISL_1066203, EPI_ISL_1066211, EPI_ISL_1066213, EPI_ISL_1066217, EPI_ISL_1066219, EPI_ISL_1066221, EPI_ISL_1066223, EPI_ISL_1066225, EPI_ISL_1066226, EPI_ISL_1066238, EPI_ISL_1066240, EPI_ISL_1066244, EPI_ISL_1066246, EPI_ISL_1066247, EPI_ISL_1066251, EPI_ISL_1066260, EPI_ISL_1066268, EPI_ISL_1066271, EPI_ISL_1066274, EPI_ISL_1066276, EPI_ISL_1066296, EPI_ISL_1066298, EPI_ISL_1066300, EPI_ISL_1066306, EPI_ISL_1066317, EPI_ISL_1066319, EPI_ISL_1066320, EPI_ISL_1066327, EPI_ISL_1066334, EPI_ISL_1066341, EPI_ISL_1066343, EPI_ISL_1066347, EPI_ISL_1066348, EPI_ISL_1066358, EPI_ISL_1066360, EPI_ISL_1066364, EPI_ISL_1066372, EPI_ISL_1066378, EPI_ISL_1066381, EPI_ISL_1066386, EPI_ISL_1066387, EPI_ISL_1066389, EPI_ISL_1066393, EPI_ISL_1066416, EPI_ISL_1066419, EPI_ISL_1066421, EPI_ISL_1066424, EPI_ISL_1066430, EPI_ISL_1066435, EPI_ISL_1066440, EPI_ISL_1066447, EPI_ISL_1066451, EPI_ISL_1066452, EPI_ISL_1066453, EPI_ISL_1066462, EPI_ISL_1066463, EPI_ISL_1066468, EPI_ISL_1066469, EPI_ISL_1066470, EPI_ISL_1066475, EPI_ISL_1066479, EPI_ISL_1066481, EPI_ISL_1066482, EPI_ISL_1066485, EPI_ISL_1066486, EPI_ISL_1066488, EPI_ISL_1066493, EPI_ISL_1066495, EPI_ISL_1066499, EPI_ISL_1066503, EPI_ISL_1066511, EPI_ISL_1066513, EPI_ISL_1066514, EPI_ISL_1066520, EPI_ISL_1066521, EPI_ISL_1066523, EPI_ISL_1066527, EPI_ISL_1066529, EPI_ISL_1066531, EPI_ISL_1066535, EPI_ISL_1066537, EPI_ISL_1066548, EPI_ISL_1066549, EPI_ISL_1066554, EPI_ISL_1066555, EPI_ISL_1066556, EPI_ISL_1066557, EPI_ISL_1066561, EPI_ISL_1066562, EPI_ISL_1066563, EPI_ISL_1066564, EPI_ISL_1066566, EPI_ISL_1066568, EPI_ISL_1066569, EPI_ISL_1066573, EPI_ISL_1066574, EPI_ISL_1066580, EPI_ISL_1066582, EPI_ISL_1066589, EPI_ISL_1066598, EPI_ISL_1066600, EPI_ISL_1066601, EPI_ISL_1066611, EPI_ISL_1066615, EPI_ISL_1066618, EPI_ISL_1066621, EPI_ISL_1066622, EPI_ISL_1066623, EPI_ISL_1066624, EPI_ISL_1066625, EPI_ISL_1066626, EPI_ISL_1066627, EPI_ISL_1066632, EPI_ISL_1066634, EPI_ISL_1066636, EPI_ISL_1066643, EPI_ISL_1066644, EPI_ISL_1066645, EPI_ISL_1066648, EPI_ISL_1066650, EPI_ISL_1066651, EPI_ISL_1066652, EPI_ISL_1066654, EPI_ISL_1066664, EPI_ISL_1066671, EPI_ISL_1066674, EPI_ISL_1066680, EPI_ISL_1066682, EPI_ISL_1066688, EPI_ISL_1066689, EPI_ISL_1066690, EPI_ISL_1066695, EPI_ISL_1066702, EPI_ISL_1066704, EPI_ISL_1066706, EPI_ISL_1066707, EPI_ISL_1066709, EPI_ISL_1066711, EPI_ISL_1066713, EPI_ISL_1066718, EPI_ISL_1066720, EPI_ISL_1066725, EPI_ISL_1066733, EPI_ISL_1066735, EPI_ISL_1066737, EPI_ISL_1066738, EPI_ISL_1066745, EPI_ISL_1066746, EPI_ISL_1066749, EPI_ISL_1066752, EPI_ISL_1066755, EPI_ISL_1066779, EPI_ISL_1066785, EPI_ISL_1066791, EPI_ISL_1066809, EPI_ISL_1066817, EPI_ISL_1066820, EPI_ISL_1066825, EPI_ISL_1066826, EPI_ISL_1066827, EPI_ISL_1066834, EPI_ISL_1066839, EPI_ISL_1066841, EPI_ISL_1066842, EPI_ISL_1066844, EPI_ISL_1066850, EPI_ISL_1066852, EPI_ISL_1066853, EPI_ISL_1066857, EPI_ISL_1066858, EPI_ISL_1066859, EPI_ISL_1066860, EPI_ISL_1066862, EPI_ISL_1066864, EPI_ISL_1066869, EPI_ISL_1066873, EPI_ISL_1066878, EPI_ISL_1066887, EPI_ISL_1066891, EPI_ISL_1066897, EPI_ISL_1066903, EPI_ISL_1066904, EPI_ISL_1066906, EPI_ISL_1066912, EPI_ISL_1066921, EPI_ISL_1066923, EPI_ISL_1066926, EPI_ISL_1066931, EPI_ISL_1066935, EPI_ISL_1066937, EPI_ISL_1066941, EPI_ISL_1066943, EPI_ISL_1066946, EPI_ISL_1066948, EPI_ISL_1066950, EPI_ISL_1066952, EPI_ISL_1066955, EPI_ISL_1066957, EPI_ISL_1066963, EPI_ISL_1066964, EPI_ISL_1066965, EPI_ISL_1066968, EPI_ISL_1066969, EPI_ISL_1066970, EPI_ISL_1066973, EPI_ISL_1066974, EPI_ISL_1066975, EPI_ISL_1066979, EPI_ISL_1066980, EPI_ISL_1066985, EPI_ISL_1066986, EPI_ISL_1066987, EPI_ISL_1066995, EPI_ISL_1066996, EPI_ISL_1066999, EPI_ISL_1067003, EPI_ISL_1067006, EPI_ISL_1067007, EPI_ISL_1067013, EPI_ISL_1067014, EPI_ISL_1067015, EPI_ISL_1067016, EPI_ISL_1067018, EPI_ISL_1067020, EPI_ISL_1067021, EPI_ISL_1067022, EPI_ISL_1067029, EPI_ISL_1067036, EPI_ISL_1067038, EPI_ISL_1067044, EPI_ISL_1067045, EPI_ISL_1067046, EPI_ISL_1067052, EPI_ISL_1067054, EPI_ISL_1067066, EPI_ISL_1067068, EPI_ISL_1067072, EPI_ISL_1067075, EPI_ISL_1067080, EPI_ISL_1067090, EPI_ISL_1067091, EPI_ISL_1067094, EPI_ISL_1067110, EPI_ISL_1067124, EPI_ISL_1067125, EPI_ISL_1067132, EPI_ISL_1067133, EPI_ISL_1067137, EPI_ISL_1067141, EPI_ISL_1067142, EPI_ISL_1067143, EPI_ISL_1067145, EPI_ISL_1067148, EPI_ISL_1067152, EPI_ISL_1067156, EPI_ISL_1067158, EPI_ISL_1067161, EPI_ISL_1067163, EPI_ISL_1067164, EPI_ISL_1067168, EPI_ISL_1067172, EPI_ISL_1067176, EPI_ISL_1067177, EPI_ISL_1067184, EPI_ISL_1067188, EPI_ISL_1067193, EPI_ISL_1067197, EPI_ISL_1067199, EPI_ISL_1067204, EPI_ISL_1067205, EPI_ISL_1067210, EPI_ISL_1067211, EPI_ISL_1067212, EPI_ISL_1067216, EPI_ISL_1067217, EPI_ISL_1067224, EPI_ISL_1067229, EPI_ISL_1067235, EPI_ISL_1067237, EPI_ISL_1067241, EPI_ISL_1067244, EPI_ISL_1067246, EPI_ISL_1067249, EPI_ISL_1067253, EPI_ISL_1067256, EPI_ISL_1067257, EPI_ISL_1067258, EPI_ISL_1067264, EPI_ISL_1067267, EPI_ISL_1067268, EPI_ISL_1067274, EPI_ISL_1067277, EPI_ISL_1067279, EPI_ISL_1067280, EPI_ISL_1067281, EPI_ISL_1067284, EPI_ISL_1067285, EPI_ISL_1067286, EPI_ISL_1067289, EPI_ISL_1067294, EPI_ISL_1067296, EPI_ISL_1067302, EPI_ISL_1067307, EPI_ISL_1067309, EPI_ISL_1067313, EPI_ISL_1067315, EPI_ISL_1067318, EPI_ISL_1067321, EPI_ISL_1067323, EPI_ISL_1067325, EPI_ISL_1067332, EPI_ISL_1067339, EPI_ISL_1067342, EPI_ISL_1067350, EPI_ISL_1067352, EPI_ISL_1067355, EPI_ISL_1067357, EPI_ISL_1067358, EPI_ISL_1067359, EPI_ISL_1067365, EPI_ISL_1067368, EPI_ISL_1067374, EPI_ISL_1067380, EPI_ISL_1067381, EPI_ISL_1067382, EPI_ISL_1067383, EPI_ISL_1067384, EPI_ISL_1067385, EPI_ISL_1067386, EPI_ISL_1067395, EPI_ISL_1067396, EPI_ISL_1067401, EPI_ISL_1067405, EPI_ISL_1067407, EPI_ISL_1067410, EPI_ISL_1067411, EPI_ISL_1067415, EPI_ISL_1067425, EPI_ISL_1067437, EPI_ISL_1067438, EPI_ISL_1067440, EPI_ISL_1067441, EPI_ISL_1067444, EPI_ISL_1067445, EPI_ISL_1067446, EPI_ISL_1067450, EPI_ISL_1067462, EPI_ISL_1067469, EPI_ISL_1067473, EPI_ISL_1067483, EPI_ISL_1067490, EPI_ISL_1067491, EPI_ISL_1067493, EPI_ISL_1067495, EPI_ISL_1067500, EPI_ISL_1067501, EPI_ISL_1067506, EPI_ISL_1067512, EPI_ISL_1067513, EPI_ISL_1067519, EPI_ISL_1067523, EPI_ISL_1067525, EPI_ISL_1067530, EPI_ISL_1067531, EPI_ISL_1067533, EPI_ISL_1067537, EPI_ISL_1067538, EPI_ISL_1067543, EPI_ISL_1067549, EPI_ISL_1067555, EPI_ISL_1067558, EPI_ISL_1067559, EPI_ISL_1067561, EPI_ISL_1067568, EPI_ISL_1067569, EPI_ISL_1067574, EPI_ISL_1067578, EPI_ISL_1067579, EPI_ISL_1067582 | | | |
| see above | Department of Virus and Microbiological Special Diagnostics, Statens Serum Institut, Copenhagen, Denmark | Aalborg University | Danish Covid-19 Genome Consortium |
| EPI_ISL_712099, EPI_ISL_712100, EPI_ISL_712119, EPI_ISL_712120, EPI_ISL_712121, EPI_ISL_712122, EPI_ISL_712123, EPI_ISL_712124, EPI_ISL_712125, EPI_ISL_712126, EPI_ISL_712127, EPI_ISL_712128, EPI_ISL_712250, EPI_ISL_712251, EPI_ISL_712252, EPI_ISL_712253, EPI_ISL_712295, EPI_ISL_712321, EPI_ISL_712340, EPI_ISL_712341, EPI_ISL_712342, EPI_ISL_712343, EPI_ISL_712344, EPI_ISL_712345, EPI_ISL_712346, EPI_ISL_712417, EPI_ISL_712431, EPI_ISL_712439, EPI_ISL_712446, EPI_ISL_712447, EPI_ISL_712448, EPI_ISL_712455, EPI_ISL_712456, EPI_ISL_712457, EPI_ISL_712458, EPI_ISL_712459, EPI_ISL_712460, EPI_ISL_712461, EPI_ISL_712462, EPI_ISL_712463, EPI_ISL_712464, EPI_ISL_712465, EPI_ISL_712466, EPI_ISL_712467, EPI_ISL_712468, EPI_ISL_712469, EPI_ISL_712470, EPI_ISL_712471, EPI_ISL_712472, EPI_ISL_712473, EPI_ISL_712474, EPI_ISL_712475, EPI_ISL_712476, EPI_ISL_712477, EPI_ISL_712478, EPI_ISL_712479, EPI_ISL_712480, EPI_ISL_712481, EPI_ISL_712482, EPI_ISL_712483, EPI_ISL_712484, EPI_ISL_712485, EPI_ISL_712486, EPI_ISL_712487, EPI_ISL_712488, EPI_ISL_712489, EPI_ISL_712490, EPI_ISL_712491, EPI_ISL_712492, EPI_ISL_712493, EPI_ISL_712494, EPI_ISL_712495, EPI_ISL_712496, EPI_ISL_712497, EPI_ISL_712498, EPI_ISL_712499, EPI_ISL_712500, EPI_ISL_712501, EPI_ISL_712502, EPI_ISL_712503, EPI_ISL_712504, EPI_ISL_712505, EPI_ISL_712506, EPI_ISL_712507, EPI_ISL_712508, EPI_ISL_712509, EPI_ISL_712510, EPI_ISL_712511, EPI_ISL_712512, EPI_ISL_712513, EPI_ISL_712514, EPI_ISL_712515, EPI_ISL_712516, EPI_ISL_712517, EPI_ISL_712518, EPI_ISL_712519, EPI_ISL_712520, EPI_ISL_712521, EPI_ISL_712522, EPI_ISL_712523, EPI_ISL_712524, EPI_ISL_712525, EPI_ISL_712526, EPI_ISL_712527, EPI_ISL_712528, EPI_ISL_712529, EPI_ISL_712530, EPI_ISL_747527, EPI_ISL_747528, EPI_ISL_747529, EPI_ISL_747533, EPI_ISL_747535, EPI_ISL_747536, EPI_ISL_747540, EPI_ISL_747541, EPI_ISL_747542, EPI_ISL_747543, EPI_ISL_747544, EPI_ISL_747545, EPI_ISL_747546, EPI_ISL_747547, EPI_ISL_747548, EPI_ISL_747549, EPI_ISL_747550, EPI_ISL_747551, EPI_ISL_747552, EPI_ISL_747553, EPI_ISL_747554, EPI_ISL_747555, EPI_ISL_747556, EPI_ISL_747557, EPI_ISL_747558, EPI_ISL_747559, EPI_ISL_747560, EPI_ISL_747561, EPI_ISL_747562, EPI_ISL_747563, EPI_ISL_747564, EPI_ISL_747565, EPI_ISL_747566, EPI_ISL_747567, EPI_ISL_747568, EPI_ISL_747569, EPI_ISL_747570, EPI_ISL_747571, EPI_ISL_747572, EPI_ISL_747573, EPI_ISL_747574, EPI_ISL_747575, EPI_ISL_747576, EPI_ISL_747589, EPI_ISL_747590, EPI_ISL_747591, EPI_ISL_747592, EPI_ISL_747593, EPI_ISL_747594, EPI_ISL_747595, EPI_ISL_747599, EPI_ISL_747601, EPI_ISL_747602, EPI_ISL_747607, EPI_ISL_747608, EPI_ISL_747610, EPI_ISL_747611, EPI_ISL_747612, EPI_ISL_747613, EPI_ISL_747614, EPI_ISL_747616, EPI_ISL_747617, EPI_ISL_747618, EPI_ISL_747619, EPI_ISL_747620, EPI_ISL_747621, EPI_ISL_747622, EPI_ISL_747623, EPI_ISL_747632, EPI_ISL_747636, EPI_ISL_747637, EPI_ISL_747638, EPI_ISL_747639, EPI_ISL_747640, EPI_ISL_747642, EPI_ISL_747643, EPI_ISL_747644, EPI_ISL_747645, EPI_ISL_747648, EPI_ISL_747652, EPI_ISL_747655, EPI_ISL_747656, EPI_ISL_747658, EPI_ISL_747659, EPI_ISL_747660, EPI_ISL_747661, EPI_ISL_747662, EPI_ISL_747663, EPI_ISL_747664, EPI_ISL_747665, EPI_ISL_747666, EPI_ISL_747667, EPI_ISL_747668, EPI_ISL_747669, EPI_ISL_747670, EPI_ISL_747671, EPI_ISL_747672, EPI_ISL_747673, EPI_ISL_747674, EPI_ISL_747681, EPI_ISL_747682, EPI_ISL_747683, EPI_ISL_747684, EPI_ISL_747685, EPI_ISL_747686, EPI_ISL_747687, EPI_ISL_747688, EPI_ISL_747689, EPI_ISL_747690, EPI_ISL_747691, EPI_ISL_747692, EPI_ISL_747693, EPI_ISL_747694, EPI_ISL_747695, EPI_ISL_747696, EPI_ISL_747697, EPI_ISL_747698, EPI_ISL_747699, EPI_ISL_747700, EPI_ISL_747701, EPI_ISL_747702, EPI_ISL_747703, EPI_ISL_747704, EPI_ISL_747705, EPI_ISL_747706, EPI_ISL_747707, EPI_ISL_747708, EPI_ISL_747721, EPI_ISL_747722, EPI_ISL_747723, EPI_ISL_747728, EPI_ISL_747729, EPI_ISL_747731, EPI_ISL_747732, EPI_ISL_747733, EPI_ISL_747734, EPI_ISL_747739, EPI_ISL_747740, EPI_ISL_747741, EPI_ISL_747742, EPI_ISL_747743, EPI_ISL_747745, EPI_ISL_747746, EPI_ISL_747748, EPI_ISL_747750, EPI_ISL_747752, EPI_ISL_747753, EPI_ISL_747754, EPI_ISL_747756, EPI_ISL_747757, EPI_ISL_747758, EPI_ISL_747759, EPI_ISL_747760, EPI_ISL_747761, EPI_ISL_747762, EPI_ISL_747763, EPI_ISL_747764, EPI_ISL_747765, EPI_ISL_747766, EPI_ISL_747767, EPI_ISL_747768, EPI_ISL_747769, EPI_ISL_747770, EPI_ISL_747771, EPI_ISL_747772, EPI_ISL_747773, EPI_ISL_747774, EPI_ISL_747775, EPI_ISL_747776, EPI_ISL_747777, EPI_ISL_747778, EPI_ISL_747779, EPI_ISL_747780, EPI_ISL_747781, EPI_ISL_747782, EPI_ISL_747783, EPI_ISL_747784, EPI_ISL_747785, EPI_ISL_747786, EPI_ISL_747787, EPI_ISL_747788, EPI_ISL_747789, EPI_ISL_747790, EPI_ISL_747791, EPI_ISL_747792, EPI_ISL_747793, EPI_ISL_747794, EPI_ISL_747795, EPI_ISL_747796, EPI_ISL_747797, EPI_ISL_747798, EPI_ISL_747799, EPI_ISL_747800, EPI_ISL_747801, EPI_ISL_747802, EPI_ISL_747803, EPI_ISL_747804, EPI_ISL_747805, EPI_ISL_747806, | | | |

We gratefully acknowledge the following Authors from the Originating laboratories responsible for obtaining the specimens, as well as the Submitting laboratories where the genome data were generated and shared via GISAID, on which this research is based.

All Submitters of data may be contacted directly via www.gisaid.org

Authors are sorted alphabetically.

| Accession ID | Originating Laboratory | Submitting Laboratory | Authors |
|---|---|---|---|
| EPI_ISL_1025345, EPI_ISL_1025346, EPI_ISL_1025347, EPI_ISL_1025348, EPI_ISL_1025349, EPI_ISL_1025350, EPI_ISL_1025351, EPI_ISL_1025352, EPI_ISL_1025353, EPI_ISL_1025354, EPI_ISL_1025355, EPI_ISL_1025356, EPI_ISL_1025357, EPI_ISL_1025358, EPI_ISL_1025359, EPI_ISL_1025360, EPI_ISL_1025361, EPI_ISL_1025362, EPI_ISL_1025363, EPI_ISL_1025364, EPI_ISL_1025365, EPI_ISL_1025366, EPI_ISL_1025367, EPI_ISL_1025368, EPI_ISL_1025369, EPI_ISL_1025370, EPI_ISL_1025371, EPI_ISL_1025372, EPI_ISL_1025373, EPI_ISL_1025374, EPI_ISL_1025375, EPI_ISL_1025376, EPI_ISL_1025377, EPI_ISL_1025378, EPI_ISL_1025379, EPI_ISL_1025380, EPI_ISL_1025381, EPI_ISL_1025382, EPI_ISL_1025383, EPI_ISL_1025384, EPI_ISL_1025385, EPI_ISL_1025386, EPI_ISL_1025387, EPI_ISL_1025388, EPI_ISL_1025389, EPI_ISL_1025390, EPI_ISL_1025391, EPI_ISL_1025392, EPI_ISL_1025393, EPI_ISL_1025394, EPI_ISL_1025395, EPI_ISL_1025396, EPI_ISL_1025397, EPI_ISL_1025398, EPI_ISL_1025399, EPI_ISL_1025400, EPI_ISL_1025401, EPI_ISL_1025402, EPI_ISL_1025403, EPI_ISL_1025404, EPI_ISL_1025405, EPI_ISL_1025406, EPI_ISL_1025407, EPI_ISL_1025408, EPI_ISL_1025409, EPI_ISL_1025410, EPI_ISL_1025411, EPI_ISL_1025412, EPI_ISL_1025413, EPI_ISL_1025414, EPI_ISL_1025415, EPI_ISL_1025416, EPI_ISL_1025417, EPI_ISL_1025418, EPI_ISL_1025419, EPI_ISL_1025420, EPI_ISL_1025421, EPI_ISL_1025422, EPI_ISL_1025423, EPI_ISL_1025424, EPI_ISL_1025425, EPI_ISL_1025426, EPI_ISL_1025427, EPI_ISL_1025428, EPI_ISL_1025429, EPI_ISL_1025430, EPI_ISL_1025431, EPI_ISL_1025432, EPI_ISL_1025433, EPI_ISL_1025434, EPI_ISL_1025435, EPI_ISL_1025436, EPI_ISL_1025437, EPI_ISL_1025438, EPI_ISL_1025439, EPI_ISL_1025440, EPI_ISL_1025441, EPI_ISL_1025442, EPI_ISL_1025443, EPI_ISL_1025444, EPI_ISL_1025445, EPI_ISL_1025446, EPI_ISL_1025447, EPI_ISL_1025448, EPI_ISL_1025449, EPI_ISL_1025450, EPI_ISL_1025451, EPI_ISL_1025452, EPI_ISL_1025453, EPI_ISL_1025454, EPI_ISL_1025455, EPI_ISL_1025456, EPI_ISL_1025457, EPI_ISL_1025458, EPI_ISL_1025459, EPI_ISL_1025460, EPI_ISL_1025461, EPI_ISL_1025462, EPI_ISL_1025463, EPI_ISL_1025464, EPI_ISL_1025465, EPI_ISL_1025466, EPI_ISL_1025467, EPI_ISL_1025468, EPI_ISL_1025469, EPI_ISL_1025470, EPI_ISL_1025471, EPI_ISL_1025472, EPI_ISL_1025473, EPI_ISL_1025474, EPI_ISL_1025475, EPI_ISL_1025476, EPI_ISL_1025477, EPI_ISL_1025478, EPI_ISL_1025479, EPI_ISL_1025480, EPI_ISL_1025481, EPI_ISL_1025482, EPI_ISL_1025483, EPI_ISL_1025484, EPI_ISL_1025485, EPI_ISL_1025486, EPI_ISL_1025487, EPI_ISL_1025488, EPI_ISL_1025489, EPI_ISL_1025490, EPI_ISL_1025491, EPI_ISL_1025492, EPI_ISL_1025493, EPI_ISL_1025494, EPI_ISL_1025495, EPI_ISL_1025496, EPI_ISL_1025497, EPI_ISL_1025498, EPI_ISL_1025499, EPI_ISL_1025500, EPI_ISL_1025501, EPI_ISL_1025502, EPI_ISL_1025503, EPI_ISL_1025504, EPI_ISL_1025505, EPI_ISL_1025506, EPI_ISL_1025507, EPI_ISL_1025508, EPI_ISL_1025509, EPI_ISL_1025510, EPI_ISL_1025511, EPI_ISL_1025512, EPI_ISL_1025513, EPI_ISL_1025514, EPI_ISL_1025515, EPI_ISL_1025516, EPI_ISL_1025517, EPI_ISL_1025518, EPI_ISL_1025519, EPI_ISL_1025520, EPI_ISL_1025521, EPI_ISL_1025522, EPI_ISL_1025523, EPI_ISL_1025524, EPI_ISL_1025525, EPI_ISL_1025526, EPI_ISL_1025527, EPI_ISL_1025528, EPI_ISL_1025529, EPI_ISL_1025530, EPI_ISL_1025531, EPI_ISL_1025532, EPI_ISL_1025533, EPI_ISL_1025534, EPI_ISL_1025535, EPI_ISL_1025536, EPI_ISL_1025537, EPI_ISL_1025538, EPI_ISL_1025539, EPI_ISL_1025540, EPI_ISL_1025541, EPI_ISL_1025542, EPI_ISL_1025543, EPI_ISL_1025544, EPI_ISL_1025545, EPI_ISL_1025546, EPI_ISL_1025547, EPI_ISL_1025548, EPI_ISL_1025549, EPI_ISL_1025550, EPI_ISL_1025551, EPI_ISL_1025552, EPI_ISL_1025553, EPI_ISL_1025554, EPI_ISL_1025555, EPI_ISL_1025556, EPI_ISL_1025557, EPI_ISL_1025558, EPI_ISL_1025559, EPI_ISL_1025560, EPI_ISL_1025561, EPI_ISL_1025562, EPI_ISL_1025563, EPI_ISL_1025564, EPI_ISL_1025565, EPI_ISL_1025566, EPI_ISL_1025567, EPI_ISL_1025568, EPI_ISL_1025569, EPI_ISL_1025570, EPI_ISL_1025571, EPI_ISL_1025572, EPI_ISL_1025573, EPI_ISL_1025574, EPI_ISL_1025575, EPI_ISL_1025576, EPI_ISL_1025577, EPI_ISL_1025578, EPI_ISL_1025579, EPI_ISL_1025580, EPI_ISL_1025581, EPI_ISL_1025582, EPI_ISL_1025583, EPI_ISL_1025584, EPI_ISL_1025585, EPI_ISL_1025586, EPI_ISL_1025587, EPI_ISL_1025588, EPI_ISL_1025589, EPI_ISL_1025590, EPI_ISL_1025591, EPI_ISL_1025592, EPI_ISL_1025593, EPI_ISL_1025594, EPI_ISL_1025595, EPI_ISL_1025596, EPI_ISL_1025597, EPI_ISL_1025598, EPI_ISL_1025599, EPI_ISL_1025600, EPI_ISL_1025601, EPI_ISL_1025602, EPI_ISL_1025603, EPI_ISL_1025604, EPI_ISL_1025605, EPI_ISL_1025606, EPI_ISL_1025607, EPI_ISL_1025608, EPI_ISL_1025609, EPI_ISL_1025610, EPI_ISL_1025611, EPI_ISL_1025612, EPI_ISL_1025613, EPI_ISL_1025614, EPI_ISL_1025615, EPI_ISL_1025616, EPI_ISL_1025617, EPI_ISL_1025618, EPI_ISL_1025619, EPI_ISL_1025620, EPI_ISL_1025621, EPI_ISL_1025622, EPI_ISL_1025623, EPI_ISL_1025624, EPI_ISL_1025625, EPI_ISL_1025626, EPI_ISL_1025627, EPI_ISL_1025628, EPI_ISL_1025629, EPI_ISL_1025630, EPI_ISL_1025631, EPI_ISL_1025632, EPI_ISL_1025633, EPI_ISL_1025634, EPI_ISL_1025635, EPI_ISL_1025636, EPI_ISL_1025637, EPI_ISL_1025638, EPI_ISL_1025639, EPI_ISL_1025640, EPI_ISL_1025641, EPI_ISL_1025642, EPI_ISL_1025643, EPI_ISL_1025644, EPI_ISL_1025645, EPI_ISL_1025646, EPI_ISL_1025647, EPI_ISL_1025648, EPI_ISL_1025649, EPI_ISL_1025650, EPI_ISL_1025651, EPI_ISL_1025652, EPI_ISL_1025653, EPI_ISL_1025654, EPI_ISL_1025655, EPI_ISL_1025656, EPI_ISL_1025657, EPI_ISL_1025658, EPI_ISL_1025659, EPI_ISL_1025660, EPI_ISL_1025661, EPI_ISL_1025662, EPI_ISL_1025663, EPI_ISL_1025664, EPI_ISL_1025665, EPI_ISL_1025666, EPI_ISL_1025667, EPI_ISL_1025668, EPI_ISL_1025669, EPI_ISL_1065105, EPI_ISL_1065111, EPI_ISL_1065156, EPI_ISL_1065207, EPI_ISL_1065219, EPI_ISL_1065229, EPI_ISL_1065232, EPI_ISL_1065240, EPI_ISL_1065265, EPI_ISL_1065293, EPI_ISL_1065300, EPI_ISL_1065306, EPI_ISL_1065317, EPI_ISL_1065330, EPI_ISL_1065341, EPI_ISL_1065382, EPI_ISL_1065383, EPI_ISL_1065386, EPI_ISL_1065417, EPI_ISL_1065425, EPI_ISL_1065479, EPI_ISL_1065488, EPI_ISL_1065502, EPI_ISL_1065514, EPI_ISL_1065516, EPI_ISL_1065535, EPI_ISL_1065542, EPI_ISL_1065613, EPI_ISL_1065615, EPI_ISL_1065622, EPI_ISL_1065652, EPI_ISL_1065681, EPI_ISL_1065697, EPI_ISL_1065698, EPI_ISL_1065738, EPI_ISL_1065741, EPI_ISL_1065751, EPI_ISL_1065754, EPI_ISL_1065769, EPI_ISL_1065783, EPI_ISL_1065804, EPI_ISL_1065809, EPI_ISL_1065828, EPI_ISL_1065830, EPI_ISL_1065841, EPI_ISL_1065866, EPI_ISL_1065878, EPI_ISL_1065906, EPI_ISL_1065915, EPI_ISL_1065919, EPI_ISL_1065931, EPI_ISL_1065934, EPI_ISL_1065935, EPI_ISL_1065949, EPI_ISL_1065952, EPI_ISL_1065972, EPI_ISL_1065999, EPI_ISL_1066024, EPI_ISL_1066026, EPI_ISL_1066033, EPI_ISL_1066049, EPI_ISL_1066061, EPI_ISL_1066069, EPI_ISL_1066071, EPI_ISL_1066074, EPI_ISL_1066078, EPI_ISL_1066080, EPI_ISL_1066089, EPI_ISL_1066090, EPI_ISL_1066157, EPI_ISL_1066164, EPI_ISL_1066180, EPI_ISL_1066190, EPI_ISL_1066202, EPI_ISL_1066210, EPI_ISL_1066239, EPI_ISL_1066242, EPI_ISL_1066243, EPI_ISL_1066250, EPI_ISL_1066256, EPI_ISL_1066266, EPI_ISL_1066279, EPI_ISL_1066314, EPI_ISL_1066333, EPI_ISL_1066338, EPI_ISL_1066344, EPI_ISL_1066373, EPI_ISL_1066382, EPI_ISL_1066388, EPI_ISL_1066406, EPI_ISL_1066429, EPI_ISL_1066431, EPI_ISL_1066433, EPI_ISL_1066436, EPI_ISL_1066448, EPI_ISL_1066456, EPI_ISL_1066459, EPI_ISL_1066461, EPI_ISL_1066467, EPI_ISL_1066478, EPI_ISL_1066480, EPI_ISL_1066492, EPI_ISL_1066512, EPI_ISL_1066528, EPI_ISL_1066533, EPI_ISL_1066546, EPI_ISL_1066571, EPI_ISL_1066583, EPI_ISL_1066609, EPI_ISL_1066629, EPI_ISL_1066631, EPI_ISL_1066633, EPI_ISL_1066653, EPI_ISL_1066663, EPI_ISL_1066676, EPI_ISL_1066712, EPI_ISL_1066716, EPI_ISL_1066730, EPI_ISL_1066740, EPI_ISL_1066751, EPI_ISL_1066772, EPI_ISL_1066800, EPI_ISL_1066803, EPI_ISL_1066807, EPI_ISL_1066821, EPI_ISL_1066840, EPI_ISL_1066843, EPI_ISL_1066848, EPI_ISL_1066851, EPI_ISL_1066854, EPI_ISL_1066874, EPI_ISL_1066896, EPI_ISL_1066898, EPI_ISL_1066901, EPI_ISL_1066918, EPI_ISL_1066929, EPI_ISL_1066930, EPI_ISL_1066942, EPI_ISL_1066959, EPI_ISL_1066978, EPI_ISL_1066992, EPI_ISL_1066998, EPI_ISL_1067010, EPI_ISL_1067030, EPI_ISL_1067057, EPI_ISL_1067058, EPI_ISL_1067063, EPI_ISL_1067088, EPI_ISL_1067095, EPI_ISL_1067103, EPI_ISL_1067115, EPI_ISL_1067117, EPI_ISL_1067119, EPI_ISL_1067128, EPI_ISL_1067135, EPI_ISL_1067136, EPI_ISL_1067138, EPI_ISL_1067139, EPI_ISL_1067159, EPI_ISL_1067162, EPI_ISL_1067170, EPI_ISL_1067175, EPI_ISL_1067179, EPI_ISL_1067187, EPI_ISL_1067190, EPI_ISL_1067192, EPI_ISL_1067194, EPI_ISL_1067195, EPI_ISL_1067201, EPI_ISL_1067221, EPI_ISL_1067230, EPI_ISL_1067243, EPI_ISL_1067248, EPI_ISL_1067252, EPI_ISL_1067271, EPI_ISL_1067276, EPI_ISL_1067278, EPI_ISL_1067335, EPI_ISL_1067348, EPI_ISL_1067363, EPI_ISL_1067372, EPI_ISL_1067375, EPI_ISL_1067394, EPI_ISL_1067397, EPI_ISL_1067398, EPI_ISL_1067431, EPI_ISL_1067432, EPI_ISL_1067434, EPI_ISL_1067442, EPI_ISL_1067453, EPI_ISL_1067458, EPI_ISL_1067459, EPI_ISL_1067481, EPI_ISL_1067485, EPI_ISL_1067492, EPI_ISL_1067503, EPI_ISL_1067510, EPI_ISL_1067514, EPI_ISL_1067526, EPI_ISL_1067550, EPI_ISL_1067552, EPI_ISL_1067562, EPI_ISL_1067566 | | | |
| see above | Department of Virus and Microbiological Special Diagnostics, Statens Serum Institut, Copenhagen, Denmark | Aalborg University | Danish Covid-19 Genome Consortium |
| EPI_ISL_793030, EPI_ISL_793031, EPI_ISL_793032, EPI_ISL_793033, EPI_ISL_793034, EPI_ISL_793035, EPI_ISL_793036, EPI_ISL_793037, EPI_ISL_793038, EPI_ISL_793039, EPI_ISL_793040, EPI_ISL_793041, EPI_ISL_793042, EPI_ISL_793043, EPI_ISL_793044, EPI_ISL_793045, EPI_ISL_793046, EPI_ISL_793047, EPI_ISL_793048, EPI_ISL_793049, EPI_ISL_793050, EPI_ISL_793116, EPI_ISL_793117, EPI_ISL_793120, EPI_ISL_793130, EPI_ISL_793134, EPI_ISL_793142, EPI_ISL_793145, EPI_ISL_793150, EPI_ISL_793151, EPI_ISL_793159, EPI_ISL_793163, EPI_ISL_793164, EPI_ISL_793171, EPI_ISL_793179, EPI_ISL_793180, EPI_ISL_793181, EPI_ISL_793182, EPI_ISL_793183, EPI_ISL_793184, EPI_ISL_793185, EPI_ISL_793186, EPI_ISL_793187, EPI_ISL_793188, EPI_ISL_793189, EPI_ISL_793190, EPI_ISL_793191, EPI_ISL_793192, EPI_ISL_793193, EPI_ISL_793205, EPI_ISL_793206, EPI_ISL_793207, EPI_ISL_793208, EPI_ISL_793209, EPI_ISL_793210, EPI_ISL_793211, EPI_ISL_793212, EPI_ISL_793213, EPI_ISL_793214, EPI_ISL_793215, EPI_ISL_793216, EPI_ISL_793217, EPI_ISL_793296, EPI_ISL_793299, EPI_ISL_793301, EPI_ISL_793316, EPI_ISL_793317, EPI_ISL_793318, EPI_ISL_793319, EPI_ISL_793320, EPI_ISL_793321, EPI_ISL_793322, EPI_ISL_793323, EPI_ISL_793324, EPI_ISL_793325, EPI_ISL_793326, EPI_ISL_793327, EPI_ISL_793328, EPI_ISL_793329, EPI_ISL_793330, EPI_ISL_793331, EPI_ISL_793371, EPI_ISL_793374, EPI_ISL_793394, EPI_ISL_793396, EPI_ISL_793399, EPI_ISL_793400, EPI_ISL_793401, EPI_ISL_793402, EPI_ISL_793403, EPI_ISL_793406, EPI_ISL_793417, EPI_ISL_793421, EPI_ISL_793422, EPI_ISL_793423, EPI_ISL_793424, EPI_ISL_793425, EPI_ISL_793426, EPI_ISL_793427, EPI_ISL_793428, EPI_ISL_793429, EPI_ISL_793430, EPI_ISL_793431, EPI_ISL_793432, EPI_ISL_793433, EPI_ISL_793434, EPI_ISL_793435, EPI_ISL_793436, EPI_ISL_793437, EPI_ISL_793438, EPI_ISL_793439, EPI_ISL_793440, EPI_ISL_793441, EPI_ISL_793442, EPI_ISL_793443, EPI_ISL_793444, EPI_ISL_793445, EPI_ISL_793446, EPI_ISL_793447, EPI_ISL_793448, EPI_ISL_793449, EPI_ISL_793450, EPI_ISL_793451, EPI_ISL_793452, EPI_ISL_793453, EPI_ISL_793454, EPI_ISL_793455, EPI_ISL_793456, EPI_ISL_793457, EPI_ISL_793458, EPI_ISL_793459, EPI_ISL_793460, EPI_ISL_793461, EPI_ISL_793462, EPI_ISL_793463, EPI_ISL_793464, EPI_ISL_793465, EPI_ISL_793466, EPI_ISL_793467, EPI_ISL_793468, EPI_ISL_793469, EPI_ISL_793470, EPI_ISL_793471, EPI_ISL_793472, EPI_ISL_793473, EPI_ISL_793474, EPI_ISL_793475, EPI_ISL_793476, EPI_ISL_793477, EPI_ISL_793478, EPI_ISL_793479, EPI_ISL_793480, EPI_ISL_793481, EPI_ISL_793482, EPI_ISL_793483, EPI_ISL_793484, EPI_ISL_793485, EPI_ISL_793486, EPI_ISL_793487, EPI_ISL_793488, EPI_ISL_793489, EPI_ISL_793490, EPI_ISL_793491, EPI_ISL_793492, EPI_ISL_793493, EPI_ISL_793494, EPI_ISL_793495, EPI_ISL_793496, EPI_ISL_793497, EPI_ISL_793498, EPI_ISL_793499, EPI_ISL_793500, EPI_ISL_793501, EPI_ISL_793502, EPI_ISL_793503, EPI_ISL_793504, EPI_ISL_793505, EPI_ISL_793506, EPI_ISL_793507, EPI_ISL_793508, EPI_ISL_793509, EPI_ISL_793510, EPI_ISL_793511, EPI_ISL_793512, EPI_ISL_793513, EPI_ISL_793514, EPI_ISL_793515, EPI_ISL_793516, EPI_ISL_793517, EPI_ISL_793518, EPI_ISL_793519, EPI_ISL_793520, EPI_ISL_793521, EPI_ISL_793522, EPI_ISL_793523, EPI_ISL_793524, EPI_ISL_793525, EPI_ISL_793526, EPI_ISL_793527, EPI_ISL_793528, EPI_ISL_793529, EPI_ISL_793530, EPI_ISL_793531, EPI_ISL_793532, EPI_ISL_793533, EPI_ISL_793534, EPI_ISL_793535, EPI_ISL_793536, EPI_ISL_793537, EPI_ISL_793538, EPI_ISL_793539, EPI_ISL_793540, EPI_ISL_793541, EPI_ISL_793542, EPI_ISL_793543, EPI_ISL_793544, EPI_ISL_793545, EPI_ISL_793546, EPI_ISL_793547, EPI_ISL_793548, EPI_ISL_793549, EPI_ISL_793550, EPI_ISL_793551, EPI_ISL_793552, EPI_ISL_793553, EPI_ISL_793554, EPI_ISL_793555, EPI_ISL_793556, EPI_ISL_793557, EPI_ISL_793558, EPI_ISL_793559, EPI_ISL_793560, EPI_ISL_793561, EPI_ISL_793562, EPI_ISL_793563, EPI_ISL_793564, EPI_ISL_793565, EPI_ISL_793566, EPI_ISL_793567, EPI_ISL_793568, EPI_ISL_793569, EPI_ISL_793570, EPI_ISL_793571, EPI_ISL_793572, EPI_ISL_793573, EPI_ISL_793574, EPI_ISL_793575, EPI_ISL_793576, EPI_ISL_793577, EPI_ISL_793578, EPI_ISL_793579, EPI_ISL_793580, EPI_ISL_793581, EPI_ISL_793582, EPI_ISL_793583, EPI_ISL_793584, EPI_ISL_793585, EPI_ISL_793586, EPI_ISL_793587, EPI_ISL_793588, EPI_ISL_793589, EPI_ISL_793590, EPI_ISL_793591, EPI_ISL_793592, EPI_ISL_793593, EPI_ISL_793594, EPI_ISL_793595, EPI_ISL_793596, EPI_ISL_793597, EPI_ISL_793598, EPI_ISL_793599, EPI_ISL_793600, EPI_ISL_793601, EPI_ISL_793602, EPI_ISL_793603, EPI_ISL_793604, EPI_ISL_793605, EPI_ISL_793606, EPI_ISL_793607, EPI_ISL_793608, EPI_ISL_793609, EPI_ISL_793610, EPI_ISL_793611, EPI_ISL_793612, EPI_ISL_793613, EPI_ISL_793614, EPI_ISL_793615, EPI_ISL_793616, EPI_ISL_793617, EPI_ISL_793618, EPI_ISL_793619, EPI_ISL_793620, EPI_ISL_793621, EPI_ISL_793622, EPI_ISL_793623, EPI_ISL_793624, EPI_ISL_793625, EPI_ISL_793626, EPI_ISL_793627, EPI_ISL_793628, EPI_ISL_793629, EPI_ISL_793630, EPI_ISL_793631, EPI_ISL_793632, EPI_ISL_793633, EPI_ISL_793634, EPI_ISL_793635, EPI_ISL_793636, EPI_ISL_793637, EPI_ISL_793638, EPI_ISL_793639, EPI_ISL_793640, EPI_ISL_793641, EPI_ISL_793642, EPI_ISL_793643, EPI_ISL_793644, EPI_ISL_793645, EPI_ISL_793646, EPI_ISL_793647, EPI_ISL_793648, EPI_ISL_793649, EPI_ISL_795011, EPI_ISL_795012, EPI_ISL_795013, EPI_ISL_795014, EPI_ISL_795015, EPI_ISL_795016, EPI_ISL_795017, EPI_ISL_795018, EPI_ISL_795019, EPI_ISL_795021, EPI_ISL_795022, EPI_ISL_795023, EPI_ISL_795024, EPI_ISL_795025, EPI_ISL_795026, EPI_ISL_795027, EPI_ISL_795028, EPI_ISL_795029, EPI_ISL_795030, EPI_ISL_795031, EPI_ISL_795032, EPI_ISL_795033, EPI_ISL_795034, EPI_ISL_795035, EPI_ISL_795036, EPI_ISL_795037, EPI_ISL_795038, EPI_ISL_795040, EPI_ISL_795041, EPI_ISL_795042, EPI_ISL_795043, EPI_ISL_795044, EPI_ISL_795045, EPI_ISL_795046, EPI_ISL_795047, EPI_ISL_795048, EPI_ISL_795049, EPI_ISL_795050, EPI_ISL_795051, EPI_ISL_795052, EPI_ISL_795053, EPI_ISL_795054, EPI_ISL_795055, EPI_ISL_795056, EPI_ISL_795057, EPI_ISL_795058, EPI_ISL_795059, EPI_ISL_795060, EPI_ISL_795061, EPI_ISL_795062, EPI_ISL_795063, EPI_ISL_795064, EPI_ISL_795065, EPI_ISL_795066, EPI_ISL_795067, EPI_ISL_795068, EPI_ISL_795069, EPI_ISL_795070, EPI_ISL_795071, EPI_ISL_795072, EPI_ISL_795073, EPI_ISL_795074, EPI_ISL_795075, EPI_ISL_795076, EPI_ISL_795077, EPI_ISL_795078, EPI_ISL_795079, EPI_ISL_795080, EPI_ISL_795081, EPI_ISL_795082, EPI_ISL_795083, EPI_ISL_795084, EPI_ISL_795085, EPI_ISL_795086, EPI_ISL_795087, EPI_ISL_795088, EPI_ISL_795089, EPI_ISL_795090, EPI_ISL_795091, EPI_ISL_795092, EPI_ISL_795093, EPI_ISL_795094, EPI_ISL_795095, EPI_ISL_795096, EPI_ISL_795097, EPI_ISL_795098, EPI_ISL_795099, EPI_ISL_795100, EPI_ISL_795101, EPI_ISL_795102, EPI_ISL_795103, EPI_ISL_795104, EPI_ISL_795105, EPI_ISL_795106, EPI_ISL_795107, EPI_ISL_795108, EPI_ISL_795109, EPI_ISL_795110, EPI_ISL_795111, EPI_ISL_795112, EPI_ISL_795113, EPI_ISL_795114, EPI_ISL_795115, EPI_ISL_795116, EPI_ISL_795117, EPI_ISL_795118, EPI_ISL_795119, EPI_ISL_795120, EPI_ISL_795121, EPI_ISL_795122, EPI_ISL_795123, EPI_ISL_795124, EPI_ISL_795125, EPI_ISL_795126, EPI_ISL_795127, EPI_ISL_795128, EPI_ISL_795129, EPI_ISL_795130, EPI_ISL_795131, EPI_ISL_795132, EPI_ISL_795133, EPI_ISL_795134, EPI_ISL_795135, EPI_ISL_795136, EPI_ISL_795137, EPI_ISL_795138, EPI_ISL_795139, EPI_ISL_795140, EPI_ISL_795141, EPI_ISL_795142, EPI_ISL_795143, EPI_ISL_795144, EPI_ISL_795145, EPI_ISL_795146, EPI_ISL_795147, EPI_ISL_795148, EPI_ISL_795154, EPI_ISL_795155, EPI_ISL_795156, EPI_ISL_795157, EPI_ISL_795158, EPI_ISL_795159, EPI_ISL_795160, EPI_ISL_795161, EPI_ISL_795162, EPI_ISL_795163, EPI_ISL_795164, EPI_ISL_795165, EPI_ISL_795166, EPI_ISL_795167, EPI_ISL_795168, EPI_ISL_795169, EPI_ISL_795171, EPI_ISL_795172, EPI_ISL_795173, EPI_ISL_795175, EPI_ISL_795176, EPI_ISL_795177, EPI_ISL_795178, EPI_ISL_795179, EPI_ISL_795180, EPI_ISL_795181, EPI_ISL_795182, EPI_ISL_795183, EPI_ISL_795184, EPI_ISL_795185, EPI_ISL_795186, EPI_ISL_795187, EPI_ISL_795188, EPI_ISL_795189, EPI_ISL_795190, EPI_ISL_795191, EPI_ISL_795192, EPI_ISL_795193, EPI_ISL_795194, EPI_ISL_795195, EPI_ISL_795196, EPI_ISL_795197, EPI_ISL_795198, EPI_ISL_795199, EPI_ISL_795200, EPI_ISL_795201, EPI_ISL_795202, EPI_ISL_795203, EPI_ISL_795204, EPI_ISL_795205, EPI_ISL_795206, EPI_ISL_795207, EPI_ISL_795208, EPI_ISL_795209, EPI_ISL_795210, EPI_ISL_795211, EPI_ISL_795212, EPI_ISL_795213, EPI_ISL_795214, EPI_ISL_795215, EPI_ISL_795216, EPI_ISL_795217, EPI_ISL_795218, EPI_ISL_795219, EPI_ISL_795220, EPI_ISL_795221, EPI_ISL_795222, EPI_ISL_795223, EPI_ISL_795224, EPI_ISL_795225, EPI_ISL_795226, EPI_ISL_795227, EPI_ISL_795228, EPI_ISL_795229, EPI_ISL_795230, EPI_ISL_795231, EPI_ISL_795232, EPI_ISL_795233, EPI_ISL_795234, EPI_ISL_795235, EPI_ISL_795236, EPI_ISL_795237, EPI_ISL_795238, EPI_ISL_795242, EPI_ISL_795244, EPI_ISL_795245, EPI_ISL_795246, EPI_ISL_795247, EPI_ISL_795248, EPI_ISL_795249, EPI_ISL_795250, EPI_ISL_795252, EPI_ISL_795253, EPI_ISL_795254, EPI_ISL_795255, EPI_ISL_795256, EPI_ISL_795257, EPI_ISL_795258, EPI_ISL_795259, EPI_ISL_795260, EPI_ISL_795261, EPI_ISL_795262, EPI_ISL_795263, EPI_ISL_795264, EPI_ISL_795265, EPI_ISL_795266, EPI_ISL_795267, EPI_ISL_795268, EPI_ISL_795269, EPI_ISL_795270, EPI_ISL_795271, EPI_ISL_795272, EPI_ISL_795273, EPI_ISL_795274, EPI_ISL_795275, EPI_ISL_795276, EPI_ISL_795277, EPI_ISL_795281, EPI_ISL_795282, EPI_ISL_795283, EPI_ISL_795284, EPI_ISL_795285, EPI_ISL_795286, EPI_ISL_795287, EPI_ISL_795288, EPI_ISL_795289, EPI_ISL_795290, EPI_ISL_795291, EPI_ISL_795292, EPI_ISL_795293, EPI_ISL_795294, EPI_ISL_795295, EPI_ISL_795296, EPI_ISL_795297, EPI_ISL_795298, EPI_ISL_795299, EPI_ISL_795300, EPI_ISL_795301, EPI_ISL_795302, EPI_ISL_795303, EPI_ISL_795304, EPI_ISL_795305, EPI_ISL_795306, EPI_ISL_795307, EPI_ISL_795308, EPI_ISL_795309, EPI_ISL_795310, EPI_ISL_795311, EPI_ISL_795312, EPI_ISL_795313, EPI_ISL_795314, EPI_ISL_795315, EPI_ISL_795316, EPI_ISL_795317, EPI_ISL_795318, EPI_ISL_795319, EPI_ISL_795320, EPI_ISL_795321, EPI_ISL_795322, EPI_ISL_795323, EPI_ISL_795324, EPI_ISL_795325, EPI_ISL_795326, EPI_ISL_795327, EPI_ISL_795328, EPI_ISL_795329, EPI_ISL_795330, EPI_ISL_795331, EPI_ISL_795332, EPI_ISL_795333, EPI_ISL_795334, EPI_ISL_795335, EPI_ISL_795336, EPI_ISL_795337, EPI_ISL_795338, EPI_ISL_795339, EPI_ISL_795340, EPI_ISL_795341, EPI_ISL_795342, EPI_ISL_795343, EPI_ISL_795344, EPI_ISL_795345, EPI_ISL_795346, EPI_ISL_795347, EPI_ISL_795348, EPI_ISL_795349, EPI_ISL_795350, EPI_ISL_795351, EPI_ISL_795352, EPI_ISL_795353, EPI_ISL_795354, EPI_ISL_795355, EPI_ISL_795356, EPI_ISL_795357, EPI_ISL_795358, EPI_ISL_795359, EPI_ISL_795360, EPI_ISL_795361, EPI_ISL_795368, EPI_ISL_795369, EPI_ISL_795370, EPI_ISL_795371, EPI_ISL_795372, EPI_ISL_795373, EPI_ISL_795374, EPI_ISL_795375, EPI_ISL_795376, EPI_ISL_795377, EPI_ISL_795378, EPI_ISL_795379, EPI_ISL_795380, EPI_ISL_795381, EPI_ISL_795382, EPI_ISL_795384, EPI_ISL_795385, EPI_ISL_795386, EPI_ISL_795387, EPI_ISL_795388, EPI_ISL_795389, EPI_ISL_795390, EPI_ISL_795391, EPI_ISL_795392, EPI_ISL_795393, EPI_ISL_795394, EPI_ISL_795395, EPI_ISL_795396, EPI_ISL_795397, EPI_ISL_795398, EPI_ISL_795399, EPI_ISL_795400, EPI_ISL_795401, EPI_ISL_795402, EPI_ISL_795403, EPI_ISL_795404, EPI_ISL_795405, EPI_ISL_795406, EPI_ISL_795407, EPI_ISL_795408, EPI_ISL_795409, EPI_ISL_795410, EPI_ISL_795411, EPI_ISL_795412, EPI_ISL_795414, EPI_ISL_795415, EPI_ISL_795416, EPI_ISL_795417, EPI_ISL_795418, EPI_ISL_795419, EPI_ISL_795420, EPI_ISL_795421, EPI_ISL_795422, EPI_ISL_795423, EPI_ISL_795424, EPI_ISL_795426, EPI_ISL_795427, EPI_ISL_795428, EPI_ISL_795429, EPI_ISL_795430, EPI_ISL_795431, EPI_ISL_795432, EPI_ISL_795433, EPI_ISL_795434, EPI_ISL_795435, | | | |

We gratefully acknowledge the following Authors from the Originating laboratories responsible for obtaining the specimens, as well as the Submitting laboratories where the genome data were generated and shared via GISAID, on which this research is based.

All Submitters of data may be contacted directly via www.gisaid.org

Authors are sorted alphabetically.

| Accession ID | Originating Laboratory | Submitting Laboratory | Authors |
|---|---|---|---|
| EPI_ISL_1024983, EPI_ISL_1024984, EPI_ISL_1024985, EPI_ISL_1024986, EPI_ISL_1024987, EPI_ISL_1024988, EPI_ISL_1024989, EPI_ISL_1024990, EPI_ISL_1024991, EPI_ISL_1024992, EPI_ISL_1024993, EPI_ISL_1024994, EPI_ISL_1024995, EPI_ISL_1024996, EPI_ISL_1024997, EPI_ISL_1024998, EPI_ISL_1024999, EPI_ISL_1025000, EPI_ISL_1025001, EPI_ISL_1025002, EPI_ISL_1025003, EPI_ISL_1025004, EPI_ISL_1025005, EPI_ISL_1025006, EPI_ISL_1025007, EPI_ISL_1025008, EPI_ISL_1025009, EPI_ISL_1025010, EPI_ISL_1025011, EPI_ISL_1025012, EPI_ISL_1025013, EPI_ISL_1025014, EPI_ISL_1025015, EPI_ISL_1025016, EPI_ISL_1025017, EPI_ISL_1025018, EPI_ISL_1025019, EPI_ISL_1025020, EPI_ISL_1025021, EPI_ISL_1025022, EPI_ISL_1025023, EPI_ISL_1025024, EPI_ISL_1025025, EPI_ISL_1025026, EPI_ISL_1025027, EPI_ISL_1025028, EPI_ISL_1025029, EPI_ISL_1025030, EPI_ISL_1025031, EPI_ISL_1025032, EPI_ISL_1025033, EPI_ISL_1025034, EPI_ISL_1025035, EPI_ISL_1025036, EPI_ISL_1025037, EPI_ISL_1025038, EPI_ISL_1025039, EPI_ISL_1025040, EPI_ISL_1025041, EPI_ISL_1025042, EPI_ISL_1025043, EPI_ISL_1025044, EPI_ISL_1025045, EPI_ISL_1025046, EPI_ISL_1025047, EPI_ISL_1025048, EPI_ISL_1025049, EPI_ISL_1025050, EPI_ISL_1025051, EPI_ISL_1025052, EPI_ISL_1025053, EPI_ISL_1025054, EPI_ISL_1025055, EPI_ISL_1025056, EPI_ISL_1025057, EPI_ISL_1025058, EPI_ISL_1025059, EPI_ISL_1025060, EPI_ISL_1025061, EPI_ISL_1025062, EPI_ISL_1025063, EPI_ISL_1025064, EPI_ISL_1025065, EPI_ISL_1025066, EPI_ISL_1025067, EPI_ISL_1025068, EPI_ISL_1025069, EPI_ISL_1025070, EPI_ISL_1025071, EPI_ISL_1025072, EPI_ISL_1025073, EPI_ISL_1025074, EPI_ISL_1025075, EPI_ISL_1025076, EPI_ISL_1025077, EPI_ISL_1025078, EPI_ISL_1025079, EPI_ISL_1025080, EPI_ISL_1025081, EPI_ISL_1025082, EPI_ISL_1025083, EPI_ISL_1025084, EPI_ISL_1025085, EPI_ISL_1025086, EPI_ISL_1025087, EPI_ISL_1025088, EPI_ISL_1025089, EPI_ISL_1025090, EPI_ISL_1025091, EPI_ISL_1025092, EPI_ISL_1025093, EPI_ISL_1025094, EPI_ISL_1025095, EPI_ISL_1025096, EPI_ISL_1025097, EPI_ISL_1025098, EPI_ISL_1025099, EPI_ISL_1025100, EPI_ISL_1025101, EPI_ISL_1025102, EPI_ISL_1025103, EPI_ISL_1025104, EPI_ISL_1025105, EPI_ISL_1025106, EPI_ISL_1025107, EPI_ISL_1025108, EPI_ISL_1025109, EPI_ISL_1025110, EPI_ISL_1025111, EPI_ISL_1025112, EPI_ISL_1025113, EPI_ISL_1025114, EPI_ISL_1025115, EPI_ISL_1025116, EPI_ISL_1025117, EPI_ISL_1025118, EPI_ISL_1025119, EPI_ISL_1025120, EPI_ISL_1025121, EPI_ISL_1025122, EPI_ISL_1025123, EPI_ISL_1025124, EPI_ISL_1025125, EPI_ISL_1025126, EPI_ISL_1025127, EPI_ISL_1025128, EPI_ISL_1025129, EPI_ISL_1025130, EPI_ISL_1025131, EPI_ISL_1025132, EPI_ISL_1025133, EPI_ISL_1025134, EPI_ISL_1025135, EPI_ISL_1025136, EPI_ISL_1025137, EPI_ISL_1025138, EPI_ISL_1025139, EPI_ISL_1025140, EPI_ISL_1025141, EPI_ISL_1025142, EPI_ISL_1025143, EPI_ISL_1025144, EPI_ISL_1025145, EPI_ISL_1025146, EPI_ISL_1025147, EPI_ISL_1025148, EPI_ISL_1025149, EPI_ISL_1025150, EPI_ISL_1025151, EPI_ISL_1025152, EPI_ISL_1025153, EPI_ISL_1025154, EPI_ISL_1025155, EPI_ISL_1025156, EPI_ISL_1025157, EPI_ISL_1025158, EPI_ISL_1025159, EPI_ISL_1025160, EPI_ISL_1025161, EPI_ISL_1025162, EPI_ISL_1025163, EPI_ISL_1025164, EPI_ISL_1025165, EPI_ISL_1025166, EPI_ISL_1025167, EPI_ISL_1025168, EPI_ISL_1025169, EPI_ISL_1025170, EPI_ISL_1025171, EPI_ISL_1025172, EPI_ISL_1025173, EPI_ISL_1025174, EPI_ISL_1025175, EPI_ISL_1025176, EPI_ISL_1025177, EPI_ISL_1025178, EPI_ISL_1025179, EPI_ISL_1025180, EPI_ISL_1025181, EPI_ISL_1025182, EPI_ISL_1025183, EPI_ISL_1025184, EPI_ISL_1025185, EPI_ISL_1025186, EPI_ISL_1025187, EPI_ISL_1025188, EPI_ISL_1025189, EPI_ISL_1025190, EPI_ISL_1025191, EPI_ISL_1025192, EPI_ISL_1025193, EPI_ISL_1025194, EPI_ISL_1025195, EPI_ISL_1025196, EPI_ISL_1025197, EPI_ISL_1025198, EPI_ISL_1025199, EPI_ISL_1025200, EPI_ISL_1025201, EPI_ISL_1025202, EPI_ISL_1025203, EPI_ISL_1025204, EPI_ISL_1025205, EPI_ISL_1025206, EPI_ISL_1025207, EPI_ISL_1025208, EPI_ISL_1025209, EPI_ISL_1025210, EPI_ISL_1025211, EPI_ISL_1025212, EPI_ISL_1025213, EPI_ISL_1025214, EPI_ISL_1025215, EPI_ISL_1025216, EPI_ISL_1025217, EPI_ISL_1025218, EPI_ISL_1025219, EPI_ISL_1025220, EPI_ISL_1025221, EPI_ISL_1025222, EPI_ISL_1025223, EPI_ISL_1025224, EPI_ISL_1025225, EPI_ISL_1025226, EPI_ISL_1025227, EPI_ISL_1025228, EPI_ISL_1025229, EPI_ISL_1025230, EPI_ISL_1025231, EPI_ISL_1025232, EPI_ISL_1025233, EPI_ISL_1025234, EPI_ISL_1025235, EPI_ISL_1025236, EPI_ISL_1025237, EPI_ISL_1025238, EPI_ISL_1025239, EPI_ISL_1025240, EPI_ISL_1025241, EPI_ISL_1025242, EPI_ISL_1025243, EPI_ISL_1025244, EPI_ISL_1025245, EPI_ISL_1025246, EPI_ISL_1025247, EPI_ISL_1025248, EPI_ISL_1025249, EPI_ISL_1025250, EPI_ISL_1025251, EPI_ISL_1025252, EPI_ISL_1025253, EPI_ISL_1025254, EPI_ISL_1025255, EPI_ISL_1025256, EPI_ISL_1025257, EPI_ISL_1025258, EPI_ISL_1025259, EPI_ISL_1025260, EPI_ISL_1025261, EPI_ISL_1025262, EPI_ISL_1025263, EPI_ISL_1025264, EPI_ISL_1025265, EPI_ISL_1025266, EPI_ISL_1025267, EPI_ISL_1025268, EPI_ISL_1025269, EPI_ISL_1025270, EPI_ISL_1025271, EPI_ISL_1025272, EPI_ISL_1025273, EPI_ISL_1025274, EPI_ISL_1025275, EPI_ISL_1025276, EPI_ISL_1025277, EPI_ISL_1025278, EPI_ISL_1025279, EPI_ISL_1025280, EPI_ISL_1025281, EPI_ISL_1025282, EPI_ISL_1025283, EPI_ISL_1025284, EPI_ISL_1025285, EPI_ISL_1025286, EPI_ISL_1025287, EPI_ISL_1025288, EPI_ISL_1025289, EPI_ISL_1025290, EPI_ISL_1025291, EPI_ISL_1025292, EPI_ISL_1025293, EPI_ISL_1025294, EPI_ISL_1025295, EPI_ISL_1025296, EPI_ISL_1025297, EPI_ISL_1025298, EPI_ISL_1025299, EPI_ISL_1025300, EPI_ISL_1025301, EPI_ISL_1025302, EPI_ISL_1025303, EPI_ISL_1025304, EPI_ISL_1025305, EPI_ISL_1025306, EPI_ISL_1025307, EPI_ISL_1025308, EPI_ISL_1025309, EPI_ISL_1025310, EPI_ISL_1025311, EPI_ISL_1025312, EPI_ISL_1025313, EPI_ISL_1025314, EPI_ISL_1025315, EPI_ISL_1025316, EPI_ISL_1025317, EPI_ISL_1025318, EPI_ISL_1025319, EPI_ISL_1025320, EPI_ISL_1025321, EPI_ISL_1025322, EPI_ISL_1025323, EPI_ISL_1025324, EPI_ISL_1025325, EPI_ISL_1025326, EPI_ISL_1025327, EPI_ISL_1025328, EPI_ISL_1025329, EPI_ISL_1025330, EPI_ISL_1025331, EPI_ISL_1025332, EPI_ISL_1025333, EPI_ISL_1025334, EPI_ISL_1025335, EPI_ISL_1025336, EPI_ISL_1025337, EPI_ISL_1025338, EPI_ISL_1025339, EPI_ISL_1025340, EPI_ISL_1025341, EPI_ISL_1025342, EPI_ISL_1025343, EPI_ISL_1025344, EPI_ISL_1065095, EPI_ISL_1065113, EPI_ISL_1065127, EPI_ISL_1065135, EPI_ISL_1065137, EPI_ISL_1065143, EPI_ISL_1065161, EPI_ISL_1065170, EPI_ISL_1065172, EPI_ISL_1065185, EPI_ISL_1065227, EPI_ISL_1065237, EPI_ISL_1065238, EPI_ISL_1065241, EPI_ISL_1065268, EPI_ISL_1065269, EPI_ISL_1065274, EPI_ISL_1065276, EPI_ISL_1065283, EPI_ISL_1065292, EPI_ISL_1065299, EPI_ISL_1065303, EPI_ISL_1065305, EPI_ISL_1065316, EPI_ISL_1065320, EPI_ISL_1065321, EPI_ISL_1065333, EPI_ISL_1065338, EPI_ISL_1065408, EPI_ISL_1065410, EPI_ISL_1065457, EPI_ISL_1065472, EPI_ISL_1065480, EPI_ISL_1065498, EPI_ISL_1065499, EPI_ISL_1065511, EPI_ISL_1065513, EPI_ISL_1065518, EPI_ISL_1065524, EPI_ISL_1065525, EPI_ISL_1065526, EPI_ISL_1065536, EPI_ISL_1065563, EPI_ISL_1065565, EPI_ISL_1065570, EPI_ISL_1065588, EPI_ISL_1065596, EPI_ISL_1065628, EPI_ISL_1065637, EPI_ISL_1065645, EPI_ISL_1065654, EPI_ISL_1065663, EPI_ISL_1065688, EPI_ISL_1065703, EPI_ISL_1065734, EPI_ISL_1065743, EPI_ISL_1065767, EPI_ISL_1065802, EPI_ISL_1065808, EPI_ISL_1065811, EPI_ISL_1065859, EPI_ISL_1065871, EPI_ISL_1065875, EPI_ISL_1065885, EPI_ISL_1065888, EPI_ISL_1065898, EPI_ISL_1065908, EPI_ISL_1065937, EPI_ISL_1065950, EPI_ISL_1065957, EPI_ISL_1065960, EPI_ISL_1065974, EPI_ISL_1066002, EPI_ISL_1066009, EPI_ISL_1066010, EPI_ISL_1066012, EPI_ISL_1066019, EPI_ISL_1066025, EPI_ISL_1066040, EPI_ISL_1066048, EPI_ISL_1066066, EPI_ISL_1066087, EPI_ISL_1066099, EPI_ISL_1066127, EPI_ISL_1066129, EPI_ISL_1066140, EPI_ISL_1066141, EPI_ISL_1066158, EPI_ISL_1066159, EPI_ISL_1066183, EPI_ISL_1066184, EPI_ISL_1066188, EPI_ISL_1066191, EPI_ISL_1066192, EPI_ISL_1066194, EPI_ISL_1066195, EPI_ISL_1066199, EPI_ISL_1066205, EPI_ISL_1066208, EPI_ISL_1066232, EPI_ISL_1066233, EPI_ISL_1066245, EPI_ISL_1066285, EPI_ISL_1066304, EPI_ISL_1066324, EPI_ISL_1066339, EPI_ISL_1066346, EPI_ISL_1066359, EPI_ISL_1066370, EPI_ISL_1066371, EPI_ISL_1066374, EPI_ISL_1066375, EPI_ISL_1066403, EPI_ISL_1066412, EPI_ISL_1066415, EPI_ISL_1066432, EPI_ISL_1066437, EPI_ISL_1066444, EPI_ISL_1066494, EPI_ISL_1066496, EPI_ISL_1066506, EPI_ISL_1066508, EPI_ISL_1066515, EPI_ISL_1066516, EPI_ISL_1066545, EPI_ISL_1066572, EPI_ISL_1066576, EPI_ISL_1066591, EPI_ISL_1066606, EPI_ISL_1066610, EPI_ISL_1066630, EPI_ISL_1066659, EPI_ISL_1066666, EPI_ISL_1066685, EPI_ISL_1066697, EPI_ISL_1066698, EPI_ISL_1066708, EPI_ISL_1066710, EPI_ISL_1066726, EPI_ISL_1066727, EPI_ISL_1066753, EPI_ISL_1066754, EPI_ISL_1066757, EPI_ISL_1066761, EPI_ISL_1066812, EPI_ISL_1066813, EPI_ISL_1066814, EPI_ISL_1066832, EPI_ISL_1066866, EPI_ISL_1066867, EPI_ISL_1066875, EPI_ISL_1066876, EPI_ISL_1066909, EPI_ISL_1066938, EPI_ISL_1066944, EPI_ISL_1066954, EPI_ISL_1066988, EPI_ISL_1066997, EPI_ISL_1067011, EPI_ISL_1067012, EPI_ISL_1067037, EPI_ISL_1067048, EPI_ISL_1067074, EPI_ISL_1067092, EPI_ISL_1067098, EPI_ISL_1067105, EPI_ISL_1067112, EPI_ISL_1067166, EPI_ISL_1067203, EPI_ISL_1067214, EPI_ISL_1067238, EPI_ISL_1067242, EPI_ISL_1067254, EPI_ISL_1067259, EPI_ISL_1067303, EPI_ISL_1067306, EPI_ISL_1067353, EPI_ISL_1067361, EPI_ISL_1067362, EPI_ISL_1067376, EPI_ISL_1067387, EPI_ISL_1067389, EPI_ISL_1067393, EPI_ISL_1067413, EPI_ISL_1067422, EPI_ISL_1067436, EPI_ISL_1067452, EPI_ISL_1067466, EPI_ISL_1067468, EPI_ISL_1067482, EPI_ISL_1067488, EPI_ISL_1067520, EPI_ISL_1067539 | | | |
| see above | Department of Virus and Microbiological Special Diagnostics, Statens Serum Institut, Copenhagen, Denmark | Aalborg University | Danish Covid-19 Genome Consortium |
| EPI_ISL_844543, EPI_ISL_844545, EPI_ISL_844552, EPI_ISL_844553, EPI_ISL_844556, EPI_ISL_844557, EPI_ISL_844558, EPI_ISL_844559, EPI_ISL_844560, EPI_ISL_844561, EPI_ISL_844562, EPI_ISL_844563, EPI_ISL_844564, EPI_ISL_844565, EPI_ISL_844566, EPI_ISL_844567, EPI_ISL_844568, EPI_ISL_844569, EPI_ISL_844570, EPI_ISL_844571, EPI_ISL_844572, EPI_ISL_844573, EPI_ISL_844574, EPI_ISL_844575, EPI_ISL_844576, EPI_ISL_844577, EPI_ISL_844635, EPI_ISL_844636, EPI_ISL_844637, EPI_ISL_844638, EPI_ISL_844639, EPI_ISL_844653, EPI_ISL_844654, EPI_ISL_844655, EPI_ISL_844666, EPI_ISL_844667, EPI_ISL_844669, EPI_ISL_844670, EPI_ISL_844676, EPI_ISL_844680, EPI_ISL_844681, EPI_ISL_844693, EPI_ISL_844694, EPI_ISL_844702, EPI_ISL_844706, EPI_ISL_844707, EPI_ISL_844708, EPI_ISL_844716, EPI_ISL_844717, EPI_ISL_844718, EPI_ISL_844730, EPI_ISL_844731, EPI_ISL_844732, EPI_ISL_844733, EPI_ISL_844734, EPI_ISL_844735, EPI_ISL_844736, EPI_ISL_844737, EPI_ISL_844738, EPI_ISL_844747, EPI_ISL_844758, EPI_ISL_844759, EPI_ISL_844763, EPI_ISL_844766, EPI_ISL_844767, EPI_ISL_844768, EPI_ISL_844788, EPI_ISL_844805, EPI_ISL_844807, EPI_ISL_844808, EPI_ISL_844818, EPI_ISL_844821, EPI_ISL_844824, EPI_ISL_844834, EPI_ISL_844836, EPI_ISL_844837, EPI_ISL_844842, EPI_ISL_844843, EPI_ISL_844844, EPI_ISL_844845, EPI_ISL_844846, EPI_ISL_844847, EPI_ISL_844848, EPI_ISL_844849, EPI_ISL_844860, EPI_ISL_844861, EPI_ISL_844862, EPI_ISL_844863, EPI_ISL_844864, EPI_ISL_844865, EPI_ISL_844911, EPI_ISL_844912, EPI_ISL_844931, EPI_ISL_844932, EPI_ISL_844948, EPI_ISL_844951, EPI_ISL_844953, EPI_ISL_844954, EPI_ISL_844955, EPI_ISL_844956, EPI_ISL_844957, EPI_ISL_844958, EPI_ISL_844978, EPI_ISL_844981, EPI_ISL_844988, EPI_ISL_844991, EPI_ISL_844992, EPI_ISL_844993, EPI_ISL_844998, EPI_ISL_844999, EPI_ISL_845000, EPI_ISL_845001, EPI_ISL_845002, EPI_ISL_845003, EPI_ISL_845004, EPI_ISL_845005, EPI_ISL_845006, EPI_ISL_845007, EPI_ISL_845008, EPI_ISL_845009, EPI_ISL_845010, EPI_ISL_845011, EPI_ISL_845012, EPI_ISL_845013, EPI_ISL_845014, EPI_ISL_845015, EPI_ISL_845016, EPI_ISL_845017, EPI_ISL_845018, EPI_ISL_845019, EPI_ISL_845020, EPI_ISL_845021, EPI_ISL_845022, EPI_ISL_845023, EPI_ISL_845024, EPI_ISL_845025, EPI_ISL_845026, EPI_ISL_845027, EPI_ISL_845028, EPI_ISL_845029, EPI_ISL_845030, EPI_ISL_845031, EPI_ISL_845032, EPI_ISL_845033, EPI_ISL_845034, EPI_ISL_845035, EPI_ISL_845036, EPI_ISL_845037, EPI_ISL_845038, EPI_ISL_845039, EPI_ISL_845040, EPI_ISL_845041, EPI_ISL_845042, EPI_ISL_845043, EPI_ISL_845044, EPI_ISL_845045, EPI_ISL_845046, EPI_ISL_845047, EPI_ISL_845048, EPI_ISL_845049, EPI_ISL_845050, EPI_ISL_845051, EPI_ISL_845052, EPI_ISL_845053, EPI_ISL_845054, EPI_ISL_845055, EPI_ISL_845056, EPI_ISL_845057, EPI_ISL_845058, EPI_ISL_845059, EPI_ISL_845060, EPI_ISL_845061, EPI_ISL_845062, EPI_ISL_845063, EPI_ISL_845064, EPI_ISL_845065, EPI_ISL_845066, EPI_ISL_845067, EPI_ISL_845068, EPI_ISL_845069, EPI_ISL_845070, EPI_ISL_845071, EPI_ISL_845072, EPI_ISL_845073, EPI_ISL_845074, EPI_ISL_845075, EPI_ISL_845076, EPI_ISL_845077, EPI_ISL_845078, EPI_ISL_845079, EPI_ISL_845080, EPI_ISL_845275, EPI_ISL_845276, EPI_ISL_845277, EPI_ISL_845278, EPI_ISL_845279, EPI_ISL_845280, EPI_ISL_845377, EPI_ISL_845378, EPI_ISL_845379, EPI_ISL_845380, EPI_ISL_845381, EPI_ISL_845382, EPI_ISL_845383, EPI_ISL_845384, EPI_ISL_845385, EPI_ISL_845386, EPI_ISL_845387, EPI_ISL_845388, EPI_ISL_845389, EPI_ISL_845390, EPI_ISL_845391, EPI_ISL_845392, EPI_ISL_845393, EPI_ISL_845394, EPI_ISL_845395, EPI_ISL_845396, EPI_ISL_845397, EPI_ISL_845398, EPI_ISL_845399, EPI_ISL_845498, EPI_ISL_845499, EPI_ISL_845500, EPI_ISL_846775, EPI_ISL_846776, EPI_ISL_846777, EPI_ISL_846778, EPI_ISL_846780, EPI_ISL_846782, EPI_ISL_846783, EPI_ISL_846784, EPI_ISL_846785, EPI_ISL_846786, EPI_ISL_846787, EPI_ISL_846788, EPI_ISL_846789, EPI_ISL_846790, EPI_ISL_846791, EPI_ISL_846792, EPI_ISL_846793, EPI_ISL_846794, EPI_ISL_846795, EPI_ISL_846796, EPI_ISL_846797, EPI_ISL_846798, EPI_ISL_846799, EPI_ISL_846800, EPI_ISL_846801, EPI_ISL_846802, EPI_ISL_846803, EPI_ISL_846804, EPI_ISL_846805, EPI_ISL_846806, EPI_ISL_846807, EPI_ISL_846808, EPI_ISL_846809, EPI_ISL_846810, EPI_ISL_846811, EPI_ISL_846821, EPI_ISL_846822, EPI_ISL_846823, EPI_ISL_846824, EPI_ISL_846825, EPI_ISL_846826, EPI_ISL_846827, EPI_ISL_846828, EPI_ISL_846829, EPI_ISL_846830, EPI_ISL_846831, EPI_ISL_846832, EPI_ISL_846833, EPI_ISL_846834, EPI_ISL_846835, EPI_ISL_846836, EPI_ISL_846837, EPI_ISL_846838, EPI_ISL_846839, EPI_ISL_846840, EPI_ISL_846841, EPI_ISL_846842, EPI_ISL_846843, EPI_ISL_846844, EPI_ISL_846845, EPI_ISL_846846, EPI_ISL_846847, EPI_ISL_846848, EPI_ISL_846849, EPI_ISL_846850, EPI_ISL_846854, EPI_ISL_846855, EPI_ISL_846856, EPI_ISL_846857, EPI_ISL_846858, EPI_ISL_846859, EPI_ISL_846862, EPI_ISL_846863, EPI_ISL_846864, EPI_ISL_846865, EPI_ISL_846867, EPI_ISL_846868, EPI_ISL_846869, EPI_ISL_846870, EPI_ISL_846871, EPI_ISL_846872, EPI_ISL_846873, EPI_ISL_846874, EPI_ISL_846875, EPI_ISL_846876, EPI_ISL_846878, EPI_ISL_846879, EPI_ISL_846880, EPI_ISL_846881, EPI_ISL_846882, EPI_ISL_846883, EPI_ISL_846885, EPI_ISL_846887, EPI_ISL_846890, EPI_ISL_846891, EPI_ISL_846892, EPI_ISL_846896, EPI_ISL_846897, EPI_ISL_846899, EPI_ISL_846900, EPI_ISL_846901, EPI_ISL_846902, EPI_ISL_846903, EPI_ISL_846905, EPI_ISL_846906, EPI_ISL_846907, EPI_ISL_846908, EPI_ISL_846909, EPI_ISL_846911, EPI_ISL_846912, EPI_ISL_846914, EPI_ISL_846915, EPI_ISL_846916, EPI_ISL_846917, EPI_ISL_846918, EPI_ISL_846919, EPI_ISL_846920, EPI_ISL_846921, EPI_ISL_846922, EPI_ISL_846923, EPI_ISL_846924, EPI_ISL_846925, EPI_ISL_846926, EPI_ISL_846927, EPI_ISL_846928, EPI_ISL_846929, EPI_ISL_846930, EPI_ISL_846931, EPI_ISL_846932, EPI_ISL_846933, EPI_ISL_846934, EPI_ISL_846935, EPI_ISL_846936, EPI_ISL_846937, EPI_ISL_846938, EPI_ISL_846939, EPI_ISL_846940, EPI_ISL_846941, EPI_ISL_846942, EPI_ISL_846943, EPI_ISL_846944, EPI_ISL_846945, EPI_ISL_846946, EPI_ISL_846947, EPI_ISL_846948, EPI_ISL_846949, EPI_ISL_846950, EPI_ISL_846951, EPI_ISL_846952, EPI_ISL_846953, EPI_ISL_846954, EPI_ISL_846955, EPI_ISL_846956, EPI_ISL_846957, EPI_ISL_846958, EPI_ISL_846959, EPI_ISL_846960, EPI_ISL_846961, EPI_ISL_846962, EPI_ISL_846963, EPI_ISL_846964, EPI_ISL_846965, EPI_ISL_846966, EPI_ISL_846967, EPI_ISL_846968, EPI_ISL_846969, EPI_ISL_846970, EPI_ISL_846971, EPI_ISL_846972, EPI_ISL_846973, EPI_ISL_846974, EPI_ISL_846975, EPI_ISL_846976, EPI_ISL_846977, EPI_ISL_846978, EPI_ISL_846979, EPI_ISL_846980, EPI_ISL_846981, EPI_ISL_846982, EPI_ISL_846983, EPI_ISL_846984, EPI_ISL_846985, EPI_ISL_846986, EPI_ISL_846987, EPI_ISL_846988, EPI_ISL_846989, EPI_ISL_846990, EPI_ISL_846991, EPI_ISL_846992, EPI_ISL_846993, EPI_ISL_846994, EPI_ISL_846995, EPI_ISL_846996, EPI_ISL_846997, EPI_ISL_846998, EPI_ISL_846999, EPI_ISL_847000, EPI_ISL_847001, EPI_ISL_847002, EPI_ISL_847003, EPI_ISL_847004, EPI_ISL_847005, EPI_ISL_847006, EPI_ISL_847007, EPI_ISL_847008, EPI_ISL_847009, EPI_ISL_847010, EPI_ISL_847011, EPI_ISL_847012, EPI_ISL_847013, EPI_ISL_847014, EPI_ISL_847015, EPI_ISL_847016, EPI_ISL_847017, EPI_ISL_847018, EPI_ISL_847019, EPI_ISL_847020, EPI_ISL_847021, EPI_ISL_847022, EPI_ISL_847023, EPI_ISL_847024, EPI_ISL_847025, EPI_ISL_847026, EPI_ISL_847027, EPI_ISL_847028, EPI_ISL_847029, EPI_ISL_847030, EPI_ISL_847031, EPI_ISL_847032, EPI_ISL_847033, EPI_ISL_847034, EPI_ISL_847035, EPI_ISL_847036, EPI_ISL_847037, EPI_ISL_847038, EPI_ISL_847039, EPI_ISL_847040, EPI_ISL_847041, EPI_ISL_847042, EPI_ISL_847043, EPI_ISL_847044, EPI_ISL_847045, EPI_ISL_847046, EPI_ISL_847047, EPI_ISL_847048, EPI_ISL_847049, EPI_ISL_847050, EPI_ISL_847051, EPI_ISL_847052, EPI_ISL_847053, EPI_ISL_847054, EPI_ISL_847055, EPI_ISL_847056, EPI_ISL_847057, EPI_ISL_847058, EPI_ISL_847059, EPI_ISL_847060, EPI_ISL_847061, EPI_ISL_847062, EPI_ISL_847063, EPI_ISL_847064, EPI_ISL_847065, EPI_ISL_847066, EPI_ISL_847067, EPI_ISL_847068, EPI_ISL_847069, EPI_ISL_847070, EPI_ISL_847071, EPI_ISL_847072, EPI_ISL_847073, EPI_ISL_847074, EPI_ISL_847075, EPI_ISL_847076, EPI_ISL_847077, EPI_ISL_847078, EPI_ISL_847079, EPI_ISL_847080, EPI_ISL_847081, EPI_ISL_847082, EPI_ISL_847083, EPI_ISL_847084, EPI_ISL_847085, EPI_ISL_847086, EPI_ISL_847087, EPI_ISL_847088, EPI_ISL_847089, EPI_ISL_847090, EPI_ISL_847091, EPI_ISL_847092, EPI_ISL_847093, EPI_ISL_847094, EPI_ISL_847095, EPI_ISL_847096, EPI_ISL_847097, EPI_ISL_847098, EPI_ISL_847099, EPI_ISL_847100, EPI_ISL_847101, EPI_ISL_847102, EPI_ISL_847103, EPI_ISL_847104, EPI_ISL_847105, EPI_ISL_847106, EPI_ISL_847107, EPI_ISL_847108, EPI_ISL_847109, EPI_ISL_847110, EPI_ISL_847111, EPI_ISL_847112, EPI_ISL_847113, EPI_ISL_847114, EPI_ISL_847115, EPI_ISL_847116, EPI_ISL_847117, EPI_ISL_847118, EPI_ISL_847119, EPI_ISL_847120, EPI_ISL_847121, EPI_ISL_847122, EPI_ISL_847123, EPI_ISL_847124, EPI_ISL_847180, EPI_ISL_847181, EPI_ISL_847182, EPI_ISL_847183, EPI_ISL_847184, EPI_ISL_847185, EPI_ISL_847186, EPI_ISL_847187, EPI_ISL_847188, EPI_ISL_847189, EPI_ISL_847190, EPI_ISL_847191, EPI_ISL_847192, EPI_ISL_847193, EPI_ISL_847194, EPI_ISL_847195, EPI_ISL_847196, EPI_ISL_847197, EPI_ISL_847198, EPI_ISL_847206, EPI_ISL_847207, EPI_ISL_847208, EPI_ISL_847209, EPI_ISL_847210, EPI_ISL_847211, EPI_ISL_847212, EPI_ISL_847213, EPI_ISL_847214, EPI_ISL_847215, EPI_ISL_847216, EPI_ISL_847217, EPI_ISL_847218, EPI_ISL_847219, EPI_ISL_847220, EPI_ISL_847221, EPI_ISL_847222, EPI_ISL_847223, EPI_ISL_847224, EPI_ISL_847225, EPI_ISL_847226, EPI_ISL_847227, EPI_ISL_847228, EPI_ISL_847229, EPI_ISL_847230, EPI_ISL_847231, EPI_ISL_847232, EPI_ISL_847238, EPI_ISL_847239, EPI_ISL_847240, EPI_ISL_847241, EPI_ISL_847242, EPI_ISL_847243, EPI_ISL_847250, EPI_ISL_847251, EPI_ISL_847257, EPI_ISL_847258, EPI_ISL_847259, EPI_ISL_847261, EPI_ISL_847265, EPI_ISL_847267, EPI_ISL_847268, EPI_ISL_847269, EPI_ISL_847271, EPI_ISL_847272, EPI_ISL_847273, EPI_ISL_847275, EPI_ISL_847276, EPI_ISL_847277, EPI_ISL_847278, EPI_ISL_847279, EPI_ISL_847281, EPI_ISL_847282, EPI_ISL_847283, EPI_ISL_847284, EPI_ISL_847285, EPI_ISL_847286, EPI_ISL_847287, EPI_ISL_847288, EPI_ISL_847289, EPI_ISL_847290, EPI_ISL_847291, EPI_ISL_847292, EPI_ISL_847293, EPI_ISL_847294, EPI_ISL_847295, EPI_ISL_847296, EPI_ISL_847297, EPI_ISL_847298, EPI_ISL_847299, EPI_ISL_847300, EPI_ISL_847301, EPI_ISL_847302, EPI_ISL_847303, EPI_ISL_847304, EPI_ISL_847305, EPI_ISL_847306, EPI_ISL_847307, EPI_ISL_847308, EPI_ISL_847309, EPI_ISL_847310, EPI_ISL_847311, EPI_ISL_847312, EPI_ISL_847313, EPI_ISL_847314, EPI_ISL_847315, EPI_ISL_847316, EPI_ISL_847317, EPI_ISL_847318, EPI_ISL_847319, EPI_ISL_847321, EPI_ISL_847322, EPI_ISL_847323, EPI_ISL_847324, EPI_ISL_847325, EPI_ISL_847326, EPI_ISL_847327, EPI_ISL_847328, EPI_ISL_847329, EPI_ISL_847330, EPI_ISL_847331, EPI_ISL_847333, EPI_ISL_847334, EPI_ISL_847336, EPI_ISL_847338, EPI_ISL_847339, EPI_ISL_847340, EPI_ISL_847341, EPI_ISL_847342, EPI_ISL_847343, EPI_ISL_847344, EPI_ISL_847345, EPI_ISL_847346, EPI_ISL_847347, EPI_ISL_847350, EPI_ISL_847354, EPI_ISL_847355, EPI_ISL_847356, EPI_ISL_847357, EPI_ISL_847359, EPI_ISL_847360, EPI_ISL_847361, EPI_ISL_847362, EPI_ISL_847363, EPI_ISL_847364, EPI_ISL_847365, EPI_ISL_847367, EPI_ISL_847371, EPI_ISL_847372, EPI_ISL_847373, EPI_ISL_847374, EPI_ISL_847375, EPI_ISL_847376, EPI_ISL_847378, EPI_ISL_847379, EPI_ISL_847380, EPI_ISL_847381, EPI_ISL_847382, EPI_ISL_847383, | | | |

We gratefully acknowledge the following Authors from the Originating laboratories responsible for obtaining the specimens, as well as the Submitting laboratories where the genome data were generated and shared via GISAID, on which this research is based.

All Submitters of data may be contacted directly via www.gisaid.org

Authors are sorted alphabetically.

| Accession ID | Originating Laboratory | Submitting Laboratory | Authors |
| --- | --- | --- | --- |
| EPI_ISL_1022420, EPI_ISL_1022421, EPI_ISL_1022422, EPI_ISL_1022423, EPI_ISL_1022424, EPI_ISL_1022425, EPI_ISL_1022426, EPI_ISL_1022427, EPI_ISL_1022428, EPI_ISL_1022429, EPI_ISL_1022430, EPI_ISL_1022431, EPI_ISL_1022432, EPI_ISL_1022433, EPI_ISL_1022434, EPI_ISL_1022435, EPI_ISL_1022436, EPI_ISL_1022437, EPI_ISL_1022438, EPI_ISL_1022439, EPI_ISL_1022440, EPI_ISL_1022441, EPI_ISL_1022442, EPI_ISL_1022443, EPI_ISL_1022444, EPI_ISL_1022445, EPI_ISL_1022446, EPI_ISL_1022447, EPI_ISL_1022448, EPI_ISL_1022449, EPI_ISL_1022450, EPI_ISL_1022451, EPI_ISL_1022452, EPI_ISL_1022453, EPI_ISL_1022454, EPI_ISL_1022455, EPI_ISL_1022456, EPI_ISL_1022457, EPI_ISL_1022458, EPI_ISL_1022459, EPI_ISL_1022460, EPI_ISL_1022461, EPI_ISL_1022462, EPI_ISL_1022463, EPI_ISL_1022464, EPI_ISL_1022465, EPI_ISL_1022466, EPI_ISL_1022467, EPI_ISL_1022468, EPI_ISL_1022469, EPI_ISL_1022470, EPI_ISL_1022471, EPI_ISL_1022472, EPI_ISL_1022473, EPI_ISL_1022474, EPI_ISL_1022475, EPI_ISL_1022476, EPI_ISL_1022477, EPI_ISL_1022478, EPI_ISL_1022479, EPI_ISL_1022480, EPI_ISL_1022481, EPI_ISL_1022482, EPI_ISL_1022483, EPI_ISL_1022484, EPI_ISL_1022485, EPI_ISL_1022486, EPI_ISL_1022487, EPI_ISL_1022488, EPI_ISL_1022489, EPI_ISL_1022490, EPI_ISL_1022491, EPI_ISL_1022492, EPI_ISL_1022493, EPI_ISL_1022494, EPI_ISL_1022495, EPI_ISL_1022496, EPI_ISL_1022497, EPI_ISL_1022498, EPI_ISL_1022499, EPI_ISL_1022500, EPI_ISL_1022501, EPI_ISL_1022502, EPI_ISL_1022503, EPI_ISL_1022504, EPI_ISL_1022505, EPI_ISL_1022506, EPI_ISL_1022507, EPI_ISL_1022508, EPI_ISL_1022509, EPI_ISL_1022510, EPI_ISL_1022511, EPI_ISL_1022512, EPI_ISL_1022513, EPI_ISL_1022514, EPI_ISL_1022515, EPI_ISL_1022516, EPI_ISL_1022517, EPI_ISL_1022518, EPI_ISL_1022519, EPI_ISL_1022520, EPI_ISL_1022521, EPI_ISL_1022522, EPI_ISL_1022523, EPI_ISL_1022524, EPI_ISL_1022525, EPI_ISL_1022526, EPI_ISL_1022527, EPI_ISL_1022528, EPI_ISL_1022529, EPI_ISL_1022530, EPI_ISL_1022531, EPI_ISL_1022532, EPI_ISL_1022533, EPI_ISL_1022534, EPI_ISL_1022535, EPI_ISL_1022536, EPI_ISL_1022537, EPI_ISL_1022538, EPI_ISL_1022539, EPI_ISL_1022540, EPI_ISL_1022541, EPI_ISL_1022542, EPI_ISL_1022543, EPI_ISL_1022544, EPI_ISL_1022545, EPI_ISL_1022546, EPI_ISL_1022547, EPI_ISL_1022548, EPI_ISL_1022549, EPI_ISL_1022550, EPI_ISL_1022551, EPI_ISL_1022552, EPI_ISL_1022553, EPI_ISL_1022554, EPI_ISL_1022555, EPI_ISL_1022556, EPI_ISL_1022557, EPI_ISL_1022558, EPI_ISL_1022559, EPI_ISL_1022560, EPI_ISL_1022561, EPI_ISL_1022562, EPI_ISL_1022563, EPI_ISL_1022564, EPI_ISL_1022565, EPI_ISL_1022566, EPI_ISL_1022567, EPI_ISL_1022568, EPI_ISL_1022569, EPI_ISL_1022570, EPI_ISL_1022571, EPI_ISL_1022572, EPI_ISL_1022573, EPI_ISL_1022574, EPI_ISL_1022575, EPI_ISL_1022576, EPI_ISL_1022577, EPI_ISL_1022578, EPI_ISL_1022579, EPI_ISL_1022580, EPI_ISL_1022581, EPI_ISL_1022582, EPI_ISL_1022583, EPI_ISL_1022584, EPI_ISL_1022585, EPI_ISL_1022586, EPI_ISL_1022587, EPI_ISL_1022588, EPI_ISL_1022589, EPI_ISL_1022590, EPI_ISL_1022591, EPI_ISL_1022592, EPI_ISL_1022593, EPI_ISL_1022594, EPI_ISL_1022595, EPI_ISL_1022596, EPI_ISL_1022597, EPI_ISL_1022598, EPI_ISL_1022599, EPI_ISL_1022600, EPI_ISL_1022601, EPI_ISL_1022602, EPI_ISL_1022603, EPI_ISL_1022604, EPI_ISL_1022605, EPI_ISL_1022606, EPI_ISL_1022607, EPI_ISL_1022608, EPI_ISL_1022609, EPI_ISL_1022610, EPI_ISL_1022611, EPI_ISL_1022612, EPI_ISL_1022613, EPI_ISL_1022614, EPI_ISL_1022615, EPI_ISL_1022616, EPI_ISL_1022617, EPI_ISL_1022618, EPI_ISL_1022619, EPI_ISL_1022620, EPI_ISL_1022621, EPI_ISL_1022622, EPI_ISL_1022623, EPI_ISL_1022624, EPI_ISL_1022625, EPI_ISL_1022626, EPI_ISL_1022627, EPI_ISL_1022628, EPI_ISL_1022629, EPI_ISL_1022630, EPI_ISL_1022631, EPI_ISL_1022632, EPI_ISL_1022633, EPI_ISL_1022634, EPI_ISL_1022635, EPI_ISL_1022636, EPI_ISL_1022637, EPI_ISL_1022638, EPI_ISL_1022639, EPI_ISL_1022640, EPI_ISL_1022641, EPI_ISL_1022642, EPI_ISL_1022643, EPI_ISL_1022644, EPI_ISL_1022645, EPI_ISL_1022646, EPI_ISL_1022647, EPI_ISL_1022648, EPI_ISL_1022649, EPI_ISL_1022650, EPI_ISL_1022651, EPI_ISL_1022652, EPI_ISL_1022653, EPI_ISL_1022654, EPI_ISL_1022655, EPI_ISL_1022656, EPI_ISL_1022657, EPI_ISL_1022658, EPI_ISL_1022659, EPI_ISL_1022660, EPI_ISL_1022661, EPI_ISL_1022662, EPI_ISL_1022663, EPI_ISL_1022664, EPI_ISL_1022665, EPI_ISL_1022666, EPI_ISL_1022667, EPI_ISL_1022668, EPI_ISL_1022669, EPI_ISL_1022670, EPI_ISL_1022671, EPI_ISL_1022672, EPI_ISL_1022673, EPI_ISL_1022674, EPI_ISL_1022675, EPI_ISL_1022676, EPI_ISL_1022677, EPI_ISL_1022678, EPI_ISL_1022679, EPI_ISL_1022680, EPI_ISL_1022681, EPI_ISL_1022682, EPI_ISL_1022683, EPI_ISL_1022684, EPI_ISL_1022685, EPI_ISL_1022686, EPI_ISL_1022687, EPI_ISL_1022688, EPI_ISL_1022689, EPI_ISL_1022690, EPI_ISL_1022691, EPI_ISL_1022692, EPI_ISL_1022693, EPI_ISL_1022694, EPI_ISL_1022695, EPI_ISL_1022696, EPI_ISL_1022697, EPI_ISL_1022698, EPI_ISL_1022699, EPI_ISL_1022700, EPI_ISL_1022701, EPI_ISL_1022702, EPI_ISL_1022703, EPI_ISL_1022704, EPI_ISL_1022705, EPI_ISL_1022706, EPI_ISL_1022707, EPI_ISL_1022708, EPI_ISL_1022709, EPI_ISL_1022710, EPI_ISL_1022711, EPI_ISL_1022712, EPI_ISL_1022713, EPI_ISL_1022714, EPI_ISL_1022715, EPI_ISL_1022716, EPI_ISL_1022717, EPI_ISL_1022718, EPI_ISL_1022719, EPI_ISL_1022720, EPI_ISL_1022721, EPI_ISL_1022722, EPI_ISL_1022723, EPI_ISL_1022724, EPI_ISL_1022725, EPI_ISL_1022726, EPI_ISL_1022727, EPI_ISL_1022728, EPI_ISL_1022729, EPI_ISL_1022730, EPI_ISL_1022731, EPI_ISL_1022732, EPI_ISL_1022733, EPI_ISL_1022734, EPI_ISL_1022735, EPI_ISL_1022736, EPI_ISL_1022737, EPI_ISL_1022738, EPI_ISL_1022739, EPI_ISL_1022740, EPI_ISL_1022741, EPI_ISL_1022742, EPI_ISL_1022743, EPI_ISL_1022744, EPI_ISL_1022745, EPI_ISL_1022746, EPI_ISL_1022747, EPI_ISL_1022748, EPI_ISL_1022749, EPI_ISL_1022750, EPI_ISL_1022751, EPI_ISL_1022752, EPI_ISL_1022753, EPI_ISL_1022754, EPI_ISL_1022755, EPI_ISL_1022756, EPI_ISL_1022757, EPI_ISL_1022758, EPI_ISL_1022759, EPI_ISL_1022760, EPI_ISL_1022761, EPI_ISL_1022762, EPI_ISL_1022763, EPI_ISL_1022764, EPI_ISL_1022765, EPI_ISL_1022766, EPI_ISL_1022767, EPI_ISL_1022768, EPI_ISL_1022769, EPI_ISL_1022770, EPI_ISL_1022771, EPI_ISL_1022772, EPI_ISL_1022773, EPI_ISL_1022774, EPI_ISL_1022775, EPI_ISL_1022776, EPI_ISL_1022777, EPI_ISL_1022778, EPI_ISL_1022779, EPI_ISL_1022780, EPI_ISL_1022781, EPI_ISL_1022782, EPI_ISL_1022783, EPI_ISL_1022784, EPI_ISL_1022785, EPI_ISL_1022786, EPI_ISL_1022787, EPI_ISL_1022788, EPI_ISL_1022789, EPI_ISL_1022790, EPI_ISL_1022791, EPI_ISL_1022792, EPI_ISL_1022793, EPI_ISL_1022794, EPI_ISL_1022795, EPI_ISL_1022796, EPI_ISL_1022797, EPI_ISL_1022798, EPI_ISL_1022799, EPI_ISL_1022800, EPI_ISL_1022801, EPI_ISL_1022802, EPI_ISL_1022803, EPI_ISL_1022804, EPI_ISL_1022805, EPI_ISL_1022806, EPI_ISL_1022807, EPI_ISL_1022808, EPI_ISL_1022809, EPI_ISL_1022810, EPI_ISL_1022811, EPI_ISL_1022812, EPI_ISL_1022813, EPI_ISL_1022814, EPI_ISL_1022815, EPI_ISL_1022816, EPI_ISL_1022817, EPI_ISL_1022818, EPI_ISL_1022819, EPI_ISL_1022820, EPI_ISL_1022821, EPI_ISL_1022822, EPI_ISL_1022823, EPI_ISL_1022824, EPI_ISL_1022825, EPI_ISL_1022826, EPI_ISL_1022827, EPI_ISL_1022828, EPI_ISL_1022829, EPI_ISL_1022830, EPI_ISL_1022831, EPI_ISL_1022832, EPI_ISL_1022833, EPI_ISL_1022834, EPI_ISL_1022835, EPI_ISL_1022836, EPI_ISL_1022837, EPI_ISL_1022838, EPI_ISL_1022839, EPI_ISL_1022840, EPI_ISL_1022841, EPI_ISL_1022842, EPI_ISL_1022843, EPI_ISL_1022844, EPI_ISL_1022845, EPI_ISL_1022846, EPI_ISL_1022847, EPI_ISL_1022848, EPI_ISL_1022849, EPI_ISL_1022850, EPI_ISL_1022851, EPI_ISL_1022852, EPI_ISL_1022853, EPI_ISL_1022854, EPI_ISL_1022855, EPI_ISL_1022856, EPI_ISL_1022857, EPI_ISL_1022858, EPI_ISL_1022859, EPI_ISL_1022860, EPI_ISL_1022861, EPI_ISL_1022862, EPI_ISL_1022863, EPI_ISL_1022864, EPI_ISL_1022865, EPI_ISL_1022866, EPI_ISL_1022867, EPI_ISL_1022868, EPI_ISL_1022869, EPI_ISL_1022870, EPI_ISL_1022871, EPI_ISL_1022872, EPI_ISL_1022873, EPI_ISL_1022874, EPI_ISL_1022875, EPI_ISL_1022876, EPI_ISL_1022877, EPI_ISL_1022878, EPI_ISL_1022879, EPI_ISL_1022880, EPI_ISL_1022881, EPI_ISL_1022882, EPI_ISL_1022883, EPI_ISL_1022884, EPI_ISL_1022885, EPI_ISL_1022886, EPI_ISL_1022887, EPI_ISL_1022888, EPI_ISL_1022889, EPI_ISL_1022890, EPI_ISL_1022891, EPI_ISL_1022892, EPI_ISL_1022893, EPI_ISL_1022894, EPI_ISL_1022895, EPI_ISL_1022896, EPI_ISL_1022897, EPI_ISL_1022898, EPI_ISL_1022899, EPI_ISL_1022900, EPI_ISL_1022901, EPI_ISL_1022902, EPI_ISL_1022903, EPI_ISL_1022904, EPI_ISL_1022905, EPI_ISL_1022906, EPI_ISL_1022907, EPI_ISL_1022908, EPI_ISL_1022909, EPI_ISL_1022910, EPI_ISL_1022911, EPI_ISL_1022912, EPI_ISL_1022913, EPI_ISL_1022914, EPI_ISL_1022915, EPI_ISL_1022916, EPI_ISL_1022917, EPI_ISL_1022918, EPI_ISL_1022919, EPI_ISL_1022920, EPI_ISL_1022921, EPI_ISL_1022922, EPI_ISL_1022923, EPI_ISL_1022924, EPI_ISL_1022925, EPI_ISL_1022926, EPI_ISL_1022927, EPI_ISL_1022928, EPI_ISL_1022929, EPI_ISL_1022930, EPI_ISL_1022931, EPI_ISL_1022932, EPI_ISL_1022933, EPI_ISL_1022934, EPI_ISL_1022935, EPI_ISL_1022936, EPI_ISL_1022937, EPI_ISL_1022938, EPI_ISL_1022939, EPI_ISL_1022940, EPI_ISL_1022941, EPI_ISL_1022942, EPI_ISL_1022943, EPI_ISL_1022944, EPI_ISL_1022945, EPI_ISL_1022946, EPI_ISL_1022947, EPI_ISL_1022948, EPI_ISL_1022949, EPI_ISL_1022950, EPI_ISL_1022951, EPI_ISL_1022952, EPI_ISL_1022953, EPI_ISL_1022954, EPI_ISL_1022955, EPI_ISL_1022956, EPI_ISL_1022957, EPI_ISL_1022958, EPI_ISL_1022959, EPI_ISL_1022960, EPI_ISL_1022961, EPI_ISL_1022962, EPI_ISL_1022963, EPI_ISL_1022964, EPI_ISL_1022965, EPI_ISL_1022966, EPI_ISL_1022967, EPI_ISL_1022968, EPI_ISL_1022969, EPI_ISL_1022970, EPI_ISL_1022971, EPI_ISL_1022972, EPI_ISL_1022973, EPI_ISL_1022974, EPI_ISL_1022975, EPI_ISL_1022976, EPI_ISL_1022977, EPI_ISL_1022978, EPI_ISL_1022979, EPI_ISL_1022980, EPI_ISL_1022981, EPI_ISL_1022982, EPI_ISL_1022983, EPI_ISL_1022984, EPI_ISL_1022985, EPI_ISL_1022986, EPI_ISL_1022987, EPI_ISL_1022988, EPI_ISL_1022989, EPI_ISL_1022990, EPI_ISL_1022991, EPI_ISL_1022992, EPI_ISL_1022993, EPI_ISL_1022994, EPI_ISL_1022995, EPI_ISL_1022996, EPI_ISL_1022997, EPI_ISL_1022998, EPI_ISL_1022999, EPI_ISL_1023000, EPI_ISL_1023001, EPI_ISL_1023002, EPI_ISL_1023003, EPI_ISL_1023004, EPI_ISL_1023005, EPI_ISL_1023006, EPI_ISL_1023007, EPI_ISL_1023008, EPI_ISL_1023009, EPI_ISL_1023010, EPI_ISL_1023011, EPI_ISL_1023012, EPI_ISL_1023013, EPI_ISL_1023014, EPI_ISL_1023015, EPI_ISL_1023016, EPI_ISL_1023017, EPI_ISL_1023018, EPI_ISL_1023019, EPI_ISL_1023020, EPI_ISL_1023021, EPI_ISL_1023022, EPI_ISL_1023023, EPI_ISL_1023024, EPI_ISL_1023025, EPI_ISL_1023026, EPI_ISL_1023027, EPI_ISL_1023028, EPI_ISL_1023029, EPI_ISL_1023030, EPI_ISL_1023031, EPI_ISL_1023032, EPI_ISL_1023033, EPI_ISL_1023034, EPI_ISL_1023035, EPI_ISL_1023036, EPI_ISL_1023037, EPI_ISL_1023038, EPI_ISL_1023039, EPI_ISL_1023040, EPI_ISL_1023041, EPI_ISL_1023042, EPI_ISL_1023043, EPI_ISL_1023044, EPI_ISL_1023045, EPI_ISL_1023046, EPI_ISL_1023047, EPI_ISL_1023048, EPI_ISL_1023049, EPI_ISL_1023050, EPI_ISL_1023051, EPI_ISL_1023052, EPI_ISL_1023053, EPI_ISL_1023054, EPI_ISL_1023055, EPI_ISL_1023056, EPI_ISL_1023057, EPI_ISL_1023058, EPI_ISL_1023059, EPI_ISL_1023060, EPI_ISL_1023061, EPI_ISL_1023062, EPI_ISL_1023063, EPI_ISL_1023064, EPI_ISL_1023065, EPI_ISL_1023066, EPI_ISL_1023067, EPI_ISL_1023068, EPI_ISL_1023069, EPI_ISL_1023070, EPI_ISL_1023071, EPI_ISL_1023072, EPI_ISL_1023073, EPI_ISL_1023074, EPI_ISL_1023075, EPI_ISL_1023076, EPI_ISL_1023077, EPI_ISL_1023078, EPI_ISL_1023079, EPI_ISL_1023080, EPI_ISL_1023081, EPI_ISL_1023082, EPI_ISL_1023083, EPI_ISL_1023084, EPI_ISL_1023085, EPI_ISL_1023086, EPI_ISL_1023087, EPI_ISL_1023088, EPI_ISL_1023089, EPI_ISL_1023090, EPI_ISL_1023091, EPI_ISL_1023092, EPI_ISL_1023093, EPI_ISL_1023094, EPI_ISL_1023095, EPI_ISL_1023096, EPI_ISL_1023097, EPI_ISL_1023098, EPI_ISL_1023099, EPI_ISL_1023100, EPI_ISL_1023101, EPI_ISL_1023102, EPI_ISL_1023103, EPI_ISL_1023104, EPI_ISL_1023105, EPI_ISL_1023106, EPI_ISL_1023107, EPI_ISL_1023108, EPI_ISL_1023109, EPI_ISL_1023110, EPI_ISL_1023111, EPI_ISL_1023112, EPI_ISL_1023113, EPI_ISL_1023114, EPI_ISL_1023115, EPI_ISL_1023116, EPI_ISL_1023117, EPI_ISL_1023118, EPI_ISL_1023119, EPI_ISL_1023120, EPI_ISL_1023121, EPI_ISL_1023122, EPI_ISL_1023123, EPI_ISL_1023124, EPI_ISL_1023125, EPI_ISL_1023126, EPI_ISL_1023127, EPI_ISL_1023128, EPI_ISL_1023129, EPI_ISL_1023130, EPI_ISL_1023131, EPI_ISL_1023132, EPI_ISL_1023133, EPI_ISL_1023134, EPI_ISL_1023135, EPI_ISL_1023136, EPI_ISL_1023137, EPI_ISL_1023138, EPI_ISL_1023139, EPI_ISL_1023140, EPI_ISL_1023141, EPI_ISL_1023142, EPI_ISL_1023143, EPI_ISL_1023144, EPI_ISL_1023145, EPI_ISL_1023146, EPI_ISL_1023147, EPI_ISL_1023148, EPI_ISL_1023149, EPI_ISL_1023150, EPI_ISL_1023151, EPI_ISL_1023152, EPI_ISL_1023153, EPI_ISL_1023154, EPI_ISL_1023155, EPI_ISL_1023156, EPI_ISL_1023157, EPI_ISL_1023158, EPI_ISL_1023159, EPI_ISL_1023160, EPI_ISL_1023161, EPI_ISL_1023162, EPI_ISL_1023163, EPI_ISL_1023164, EPI_ISL_1023165, EPI_ISL_1023166, EPI_ISL_1023167, EPI_ISL_1023168, EPI_ISL_1023169, EPI_ISL_1023170, EPI_ISL_1023171, EPI_ISL_1023172, EPI_ISL_1023173, EPI_ISL_1023174, EPI_ISL_1023175, EPI_ISL_1023176, EPI_ISL_1023177, EPI_ISL_1023178, EPI_ISL_1023179, EPI_ISL_1023180, EPI_ISL_1023181, EPI_ISL_1023182, EPI_ISL_1023183, EPI_ISL_1023184, EPI_ISL_1023185, EPI_ISL_1023186, EPI_ISL_1023187, EPI_ISL_1023188, EPI_ISL_1023189, EPI_ISL_1023190, EPI_ISL_1023191, EPI_ISL_1023192, EPI_ISL_1023193, EPI_ISL_1023194, EPI_ISL_1023195, EPI_ISL_1023196, EPI_ISL_1023197, EPI_ISL_1023198, EPI_ISL_1023199, EPI_ISL_1023200, EPI_ISL_1023201, EPI_ISL_1023202, EPI_ISL_1023203, EPI_ISL_1023204, EPI_ISL_1023205, EPI_ISL_1023206, EPI_ISL_1023207, EPI_ISL_1023208, EPI_ISL_1023209, EPI_ISL_1023210, EPI_ISL_1023211, EPI_ISL_1023212, EPI_ISL_1023213, EPI_ISL_1023214, EPI_ISL_1023215, EPI_ISL_1023216, EPI_ISL_1023217, EPI_ISL_1023218, EPI_ISL_1023219, EPI_ISL_1023220, EPI_ISL_1023221, EPI_ISL_1023222, EPI_ISL_1023223, EPI_ISL_1023224, EPI_ISL_1023225, EPI_ISL_1023226, EPI_ISL_1023227, EPI_ISL_1023228, EPI_ISL_1023229, EPI_ISL_1023230, EPI_ISL_1023231, EPI_ISL_1023232, EPI_ISL_1023233, EPI_ISL_1023234, EPI_ISL_1023235, EPI_ISL_1023236, EPI_ISL_1023237, EPI_ISL_1023238, EPI_ISL_1023239, EPI_ISL_1023240, EPI_ISL_1023241, EPI_ISL_1023242, EPI_ISL_1023243, EPI_ISL_1023244, EPI_ISL_1023245, EPI_ISL_1023246, EPI_ISL_1023247, EPI_ISL_1023248, EPI_ISL_1023249, EPI_ISL_1023250, EPI_ISL_1023251, EPI_ISL_1023252, EPI_ISL_1023253, EPI_ISL_1023254, EPI_ISL_1023255, EPI_ISL_1023256, EPI_ISL_1023257, EPI_ISL_1023258, EPI_ISL_1023259, EPI_ISL_1023260, EPI_ISL_1023261, EPI_ISL_1023262, EPI_ISL_1023263, EPI_ISL_1023264, EPI_ISL_1023265, EPI_ISL_1023266, EPI_ISL_1023267, EPI_ISL_1023268, EPI_ISL_1023269, EPI_ISL_1023270, EPI_ISL_1023271, EPI_ISL_1023272, EPI_ISL_1023273, EPI_ISL_1023274, EPI_ISL_1023275, EPI_ISL_1023276, EPI_ISL_1023277, EPI_ISL_1023278, EPI_ISL_1023279, EPI_ISL_1023280, EPI_ISL_1023281, EPI_ISL_1023282, EPI_ISL_1023283, EPI_ISL_1023284, EPI_ISL_1023285, EPI_ISL_1023286, EPI_ISL_1023287, EPI_ISL_1023288, EPI_ISL_1023289, EPI_ISL_1023290, EPI_ISL_1023291, EPI_ISL_1023292, EPI_ISL_1023293, EPI_ISL_1023294, EPI_ISL_1023295, EPI_ISL_1023296, EPI_ISL_1023297, EPI_ISL_1023298, EPI_ISL_1023299, EPI_ISL_1023300, EPI_ISL_1023301, EPI_ISL_1023302, EPI_ISL_1023303, EPI_ISL_1023304, EPI_ISL_1023305, EPI_ISL_1023306, EPI_ISL_1023307, EPI_ISL_1023308, EPI_ISL_1023309, EPI_ISL_1023310, EPI_ISL_1023311, EPI_ISL_1023312, EPI_ISL_1023313, EPI_ISL_1023314, EPI_ISL_1023315, EPI_ISL_1023316, EPI_ISL_1023317, EPI_ISL_1023318, EPI_ISL_1023319, EPI_ISL_1023320, EPI_ISL_1023321, EPI_ISL_1023322, EPI_ISL_1023323, EPI_ISL_1023324, EPI_ISL_1023325, EPI_ISL_1023326, EPI_ISL_1023327, EPI_ISL_1023328, EPI_ISL_1023329, EPI_ISL_1023330, EPI_ISL_1023331, EPI_ISL_1023332, EPI_ISL_1023333, EPI_ISL_1023334, EPI_ISL_1023335, EPI_ISL_1023336, EPI_ISL_1023337, EPI_ISL_1023338, EPI_ISL_1023339, EPI_ISL_1023340, EPI_ISL_1023341, EPI_ISL_1023342, EPI_ISL_1023343, EPI_ISL_1023344, EPI_ISL_1023345, EPI_ISL_1023346, EPI_ISL_1023347, EPI_ISL_1023348, EPI_ISL_1023349, EPI_ISL_1023350, EPI_ISL_1023351, EPI_ISL_1023352, EPI_ISL_1023353, EPI_ISL_1023354, EPI_ISL_1023355, EPI_ISL_1023356, EPI_ISL_1023357, EPI_ISL_1023358, EPI_ISL_1023359, EPI_ISL_1023360, EPI_ISL_1023361, EPI_ISL_1023362, EPI_ISL_1023363, EPI_ISL_1023364, EPI_ISL_1023365, EPI_ISL_1023366, EPI_ISL_1023367, EPI_ISL_1023368, EPI_ISL_1023369, EPI_ISL_1023370, EPI_ISL_1023371, EPI_ISL_1023372, EPI_ISL_1023373, EPI_ISL_1023374, EPI_ISL_1023375, EPI_ISL_1023376, EPI_ISL_1023377, EPI_ISL_1023378, EPI_ISL_1023379, EPI_ISL_1023380, EPI_ISL_1023381, EPI_ISL_1023382, EPI_ISL_1023383, EPI_ISL_1023384, EPI_ISL_1023385, EPI_ISL_1023386, EPI_ISL_1023387, EPI_ISL_1023388, EPI_ISL_1023389, EPI_ISL_1023390, EPI_ISL_1023391, EPI_ISL_1023392, EPI_ISL_1023393, EPI_ISL_1023394, EPI_ISL_1023395, EPI_ISL_1023396, EPI_ISL_1023397, EPI_ISL_1023398, EPI_ISL_1023399, EPI_ISL_1023400, EPI_ISL_1023401, EPI_ISL_1023402, EPI_ISL_1023403, EPI_ISL_1023404, EPI_ISL_1023405, EPI_ISL_1023406, EPI_ISL_1023407, EPI_ISL_1023408, EPI_ISL_1023409, EPI_ISL_1023410, EPI_ISL_1023411, EPI_ISL_1023412, EPI_ISL_1023413, EPI_ISL_1023414, EPI_ISL_1023415, EPI_ISL_1023416, EPI_ISL_1023417, EPI_ISL_1023418, EPI_ISL_1023419, EPI_ISL_1023420, EPI_ISL_1023421, EPI_ISL_1023422, EPI_ISL_1023423, EPI_ISL_1023424, EPI_ISL_1023425, EPI_ISL_1023426, EPI_ISL_1023427, EPI_ISL_1023428, EPI_ISL_1023429, EPI_ISL_1023430, EPI_ISL_1023431, EPI_ISL_1023432, EPI_ISL_1023433, EPI_ISL_1023434, EPI_ISL_1023435, EPI_ISL_1023436, EPI_ISL_1023437, EPI_ISL_1023438, EPI_ISL_1023439, EPI_ISL_1023440, EPI_ISL_1023441, EPI_ISL_1023442, EPI_ISL_1023443, EPI_ISL_1023444, EPI_ISL_1023445, EPI_ISL_1023446, EPI_ISL_1023447, EPI_ISL_1023449, EPI_ISL_1023451, EPI_ISL_1023453, EPI_ISL_1023455, EPI_ISL_1023458, EPI_ISL_1023460, EPI_ISL_1023462, EPI_ISL_1023464, EPI_ISL_1023466, EPI_ISL_1023469, EPI_ISL_1023471, EPI_ISL_1023472, EPI_ISL_1023473, EPI_ISL_1023474, EPI_ISL_1023475, EPI_ISL_1023476, EPI_ISL_1023477, EPI_ISL_1023478, EPI_ISL_1023479, EPI_ISL_1023480, EPI_ISL_1023481, EPI_ISL_1023482, EPI_ISL_1023483, EPI_ISL_1023484, EPI_ISL_1023485, EPI_ISL_1023486, EPI_ISL_1023487, EPI_ISL_1023488, EPI_ISL_1023489, EPI_ISL_1023490, EPI_ISL_1023491, EPI_ISL_1023492, EPI_ISL_1023493, EPI_ISL_1023494, EPI_ISL_1023495, EPI_ISL_1023496, EPI_ISL_1023497, EPI_ISL_1023498, EPI_ISL_1023499, EPI_ISL_1023500, EPI_ISL_1023501, EPI_ISL_1023502, EPI_ISL_1023503, EPI_ISL_1023504, EPI_ISL_1023505, EPI_ISL_1023506, EPI_ISL_1023507, EPI_ISL_1023508, EPI_ISL_1023509, EPI_ISL_1023510, EPI_ISL_1023511, EPI_ISL_1023512, EPI_ISL_1023513, EPI_ISL_1023514, EPI_ISL_1023515, EPI_ISL_1023516, EPI_ISL_1023517, EPI_ISL_1023518, EPI_ISL_1023519, EPI_ISL_1023520, EPI_ISL_1023521, EPI_ISL_1023522, EPI_ISL_1023523, EPI_ISL_1023525, EPI_ISL_1023526, EPI_ISL_1023527, EPI_ISL_1023528, EPI_ISL_1023529, EPI_ISL_1023530, EPI_ISL_1023531, EPI_ISL_1023532, EPI_ISL_1023533, EPI_ISL_1023534, EPI_ISL_1023535, EPI_ISL_1023536, EPI_ISL_1023537, EPI_ISL_1023538, EPI_ISL_1023539, EPI_ISL_1023540, EPI_ISL_1023541, EPI_ISL_1023542, EPI_ISL_1023543, EPI_ISL_1023544, EPI_ISL_1023545, EPI_ISL_1023546, EPI_ISL_1023547, EPI_ISL_1023548, EPI_ISL_1023549, EPI_ISL_1023550, EPI_ISL_1023551, EPI_ISL_1023552, EPI_ISL_1023553, EPI_ISL_1023554, EPI_ISL_1023555, EPI_ISL_1023556, EPI_ISL_1023557, EPI_ISL_1023558, EPI_ISL_1023559, EPI_ISL_1023560, EPI_ISL_1023561, EPI_ISL_1023562, EPI_ISL_1023563, EPI_ISL_1023564, EPI_ISL_1023565, EPI_ISL_1023566, EPI_ISL_1023567, EPI_ISL_1023568, EPI_ISL_1023569, EPI_ISL_1023570, EPI_ISL_1023571, EPI_ISL_1023572, EPI_ISL_1023573, EPI_ISL_1023574, EPI_ISL_1023575, EPI_ISL_1023576, EPI_ISL_1023577, EPI_ISL_1023578, EPI_ISL_1023579, EPI_ISL_1023580, EPI_ISL_1023581, EPI_ISL_1023583, EPI_ISL_1023585, EPI_ISL_1023587, EPI_ISL_1023589, EPI_ISL_1023591, EPI_ISL_1023593, EPI_ISL_1023596, EPI_ISL_1023598, EPI_ISL_1023600, EPI_ISL_1023602, EPI_ISL_1023604, EPI_ISL_1023607, EPI_ISL_1023609, EPI_ISL_1023611, EPI_ISL_1023613, EPI_ISL_1023616, EPI_ISL_1023618, EPI_ISL_1023619, EPI_ISL_1023620, EPI_ISL_1023621, EPI_ISL_1023622, EPI_ISL_1023623, EPI_ISL_1023624, EPI_ISL_1023625, EPI_ISL_1023626, EPI_ISL_1023627, EPI_ISL_1023628, EPI_ISL_1023629, EPI_ISL_1023630, EPI_ISL_1023631, EPI_ISL_1023632, EPI_ISL_1023633, EPI_ISL_1023634, EPI_ISL_1023635, EPI_ISL_1023636, EPI_ISL_1023637, EPI_ISL_1023638, EPI_ISL_1023639, EPI_ISL_1023640, EPI_ISL_1023641, EPI_ISL_1023642, EPI_ISL_1023643, EPI_ISL_1023644, EPI_ISL_1023645, EPI_ISL_1023646, EPI_ISL_1023647, EPI_ISL_1023648, EPI_ISL_1023649, EPI_ISL_1023650, EPI_ISL_1023651, EPI_ISL_1023652, EPI_ISL_1023653, EPI_ISL_1023654, EPI_ISL_1023655, EPI_ISL_1023656, EPI_ISL_1023657, EPI_ISL_1023658, EPI_ISL_1023659, EPI_ISL_1023660, EPI_ISL_1023661, EPI_ISL_1023662, EPI_ISL_1023663, EPI_ISL_1023664, EPI_ISL_1023665, EPI_ISL_1023666, EPI_ISL_1023667, EPI_ISL_1023668, EPI_ISL_1023669, EPI_ISL_1023670, EPI_ISL_1023672, EPI_ISL_1023674, EPI_ISL_1023676, EPI_ISL_1023678, EPI_ISL_1023680, EPI_ISL_1023682, EPI_ISL_1023684, EPI_ISL_1023687, EPI_ISL_1023689, EPI_ISL_1023691, EPI_ISL_1023694, EPI_ISL_1023696, EPI_ISL_1023698, EPI_ISL_1023700, EPI_ISL_1023702, EPI_ISL_1023705, EPI_ISL_1023707, EPI_ISL_1023709, EPI_ISL_1023711, EPI_ISL_1023713, EPI_ISL_1023715, EPI_ISL_1023717, EPI_ISL_1023719, EPI_ISL_1023722, EPI_ISL_1023724, EPI_ISL_1023726, EPI_ISL_1023728, EPI_ISL_1023730, EPI_ISL_1023731, EPI_ISL_1023732, EPI_ISL_1023733, EPI_ISL_1023734, EPI_ISL_1023735, EPI_ISL_1023736, EPI_ISL_1023737, EPI_ISL_1023738, EPI_ISL_1023739, EPI_ISL_1023740, EPI_ISL_1023741, EPI_ISL_1023742, EPI_ISL_1023743, EPI_ISL_1023744, EPI_ISL_1023745, EPI_ISL_1023746, EPI_ISL_1023747, EPI_ISL_1023748, EPI_ISL_1023749, EPI_ISL_1023750, EPI_ISL_1023751, EPI_ISL_1023752, EPI_ISL_1023753, EPI_ISL_1023754, EPI_ISL_1023755, EPI_ISL_1023756, EPI_ISL_1023757, EPI_ISL_1023758, EPI_ISL_1023759, EPI_ISL_1023760, | | | |

### Scotland

We gratefully acknowledge the following Authors from the Originating laboratories responsible for obtaining the specimens, as well as the Submitting laboratories where the genome data were generated and shared via GISAID, on which this research is based.

All Submitters of data may be contacted directly via www.gisaid.org

Authors are sorted alphabetically.

| Accession ID | Originating Laboratory | Submitting Laboratory | Authors |
|---|---|---|---|
| EPI_ISL_1104230, EPI_ISL_1104233 | Virology Department, Royal Infirmary of Edinburgh, NHS Lothian / School of Biological Sciences, University of Edinburgh | COVID-19 Genomics UK (COG-UK) Consortium | McHugh M, Dewar R, Cotton S, Rooke S, O'Toole Á, Scher E, Hill V, McCrone JT, Colquhoun R, Yu X, Jackson B, Rambaut A, Templeton K |
| EPI_ISL_1208395, EPI_ISL_1208396, EPI_ISL_1208397 | Lighthouse Lab in Glasgow | Wellcome Sanger Institute for the COVID-19 Genomics UK (COG-UK) Consortium | Harper VanSteenhouse, Yumi Kasai, David Gray, Carol Clugston, Anna Dominiczak and Alex Alderton, Roberto Amato, Jeffrey Barrett, Sonia Goncalves, Ewan Harrison, David K. Jackson, Ian Johnston, Dominic Kwiatkowski, Cordelia Langford, John Sillitoe on behalf of the Wellcome Sanger Institute COVID-19 Surveillance Team |
| EPI_ISL_1730120, EPI_ISL_1730121 | Virology Department, Royal Infirmary of Edinburgh, NHS Lothian / School of Biological Sciences, University of Edinburgh | COVID-19 Genomics UK (COG-UK) Consortium | McHugh M, Dewar R, Cotton S, Gallagher A, Maloney D, Rooke S, O'Toole Á, Scher E, Hill V, McCrone JT, Colquhoun R, Yu X, Jackson B, Rambaut A, Templeton K |
| EPI_ISL_413221 | West of Scotland Specialist Virology Centre, NHSGGC | MRC-University of Glasgow Centre for Virus Research | Emma Thomson, Antonia Ho; James Shephard, Shirin Ashraf; Kathy Smollett, Daniel Mair, Stephen Carmichael, Ana da Silva Filipe; Richard Orton, Josh Singer, David L Robertson; Andrew Rambaut; Alasdair MacLean, Rory Gunson. |
| EPI_ISL_414027 | West of Scotland Specialist Virology Centre, NHSGGC | MRC-University of Glasgow Centre for Virus Research | Emma Thomson, Antonia Ho; Kathy Smollett, Daniel Mair, Stephen Carmichael, Ana da Silva Filipe; Richard Orton, David L Robertson; Alasdair MacLean, Rory Gunson. |
| EPI_ISL_415629 | Virology Department, Royal Infirmary of Edinburgh, NHS Lothian | Virology Department, Royal Infirmary of Edinburgh, NHS Lothian | McHugh M, Dewar R, O'Toole Á, Rambaut A, Williams TC, Templeton K |
| EPI_ISL_415630, EPI_ISL_415631 | West of Scotland Specialist Virology Centre, NHSGGC | MRC-University of Glasgow Centre for Virus Research | Kathy Smollett, Daniel Mair, Stephen Carmichael, Ana da Silva Filipe; Richard Orton, David L Robertson; Alasdair MacLean, Rory Gunson; Natasha Jesudason, Kathy Li, Antonia Ho; Emma Thomson. |
| EPI_ISL_415640 | Virology Department, Royal Infirmary of Edinburgh, NHS Lothian | Virology Department, Royal Infirmary of Edinburgh, NHS Lothian | McHugh M, Dewar R, O'Toole Á, Rambaut A, Williams TC, Templeton K |
| EPI_ISL_425647, EPI_ISL_425648, EPI_ISL_425650, EPI_ISL_425651, EPI_ISL_425652, EPI_ISL_425653, EPI_ISL_425654, EPI_ISL_425656, EPI_ISL_425657, EPI_ISL_425658, EPI_ISL_425659, EPI_ISL_425660, EPI_ISL_425662, EPI_ISL_425663, EPI_ISL_425668, EPI_ISL_425669, EPI_ISL_425670, EPI_ISL_425671, EPI_ISL_425672, EPI_ISL_425674, EPI_ISL_425675, EPI_ISL_425676, EPI_ISL_425677, EPI_ISL_425678, EPI_ISL_425679, EPI_ISL_425682, EPI_ISL_425684, EPI_ISL_425685, EPI_ISL_425687, EPI_ISL_425688, EPI_ISL_425692, EPI_ISL_425693, EPI_ISL_425695, EPI_ISL_425696, EPI_ISL_425697, EPI_ISL_425698, EPI_ISL_425699, EPI_ISL_425700, EPI_ISL_425701, EPI_ISL_425702, EPI_ISL_425703, EPI_ISL_425704, EPI_ISL_425705, EPI_ISL_425707, EPI_ISL_425708, EPI_ISL_425709, EPI_ISL_425711, EPI_ISL_425713, EPI_ISL_425715, EPI_ISL_425716, EPI_ISL_425717, EPI_ISL_425720, EPI_ISL_425721, EPI_ISL_425722, EPI_ISL_425724, EPI_ISL_425725, EPI_ISL_425726, EPI_ISL_425727, EPI_ISL_425728, EPI_ISL_425729, EPI_ISL_425730, EPI_ISL_425731, EPI_ISL_425732, EPI_ISL_425733, EPI_ISL_425734, EPI_ISL_425735, EPI_ISL_425736, EPI_ISL_425737, EPI_ISL_425739, EPI_ISL_425740, EPI_ISL_425741, EPI_ISL_425742, EPI_ISL_425743, EPI_ISL_425744, EPI_ISL_425745, EPI_ISL_425746, EPI_ISL_425747, EPI_ISL_425748, EPI_ISL_425749, EPI_ISL_425750, EPI_ISL_425751, EPI_ISL_425752, EPI_ISL_425754, EPI_ISL_425755, EPI_ISL_425756, EPI_ISL_425757, EPI_ISL_425758, EPI_ISL_425759, EPI_ISL_425760, EPI_ISL_425761, EPI_ISL_425762, EPI_ISL_425763, EPI_ISL_425764, EPI_ISL_425765, EPI_ISL_425766, EPI_ISL_425767, EPI_ISL_425768, EPI_ISL_425769, EPI_ISL_425770, EPI_ISL_425772, EPI_ISL_425773, EPI_ISL_425774, EPI_ISL_425775, EPI_ISL_425776, EPI_ISL_425777, EPI_ISL_425778, EPI_ISL_425779, EPI_ISL_425780, EPI_ISL_425782, EPI_ISL_425783, EPI_ISL_425784, EPI_ISL_425785, EPI_ISL_425786, EPI_ISL_425787, EPI_ISL_425788, EPI_ISL_425789, EPI_ISL_425790, EPI_ISL_425791, EPI_ISL_425792, EPI_ISL_425793, EPI_ISL_425794, EPI_ISL_425795, EPI_ISL_425796, EPI_ISL_425797, EPI_ISL_425798, EPI_ISL_425799, EPI_ISL_425800, EPI_ISL_425801, EPI_ISL_425802, EPI_ISL_425803, EPI_ISL_425804, EPI_ISL_425805, EPI_ISL_425806, EPI_ISL_425807, EPI_ISL_425808, EPI_ISL_425809, EPI_ISL_425810, EPI_ISL_425811, EPI_ISL_425812, EPI_ISL_425813, EPI_ISL_425815, EPI_ISL_425816, EPI_ISL_425817, EPI_ISL_425818 | | | |
| see above | West of Scotland Specialist Virology Centre, NHSGGC / MRC-University of Glasgow Centre for Virus Research | COVID-19 Genomics UK (COG-UK) Consortium | Ana da Silva Filipe, Kathy Smollett, Stephen Carmichael, Natasha Johnson, Daniel Mair, Lily Tong, Jenna Nichols; Sarah McDonald; Richard Orton, Joseph Hughes, Sreenu Vattipally, David L Robertson; Kathy Li, Natasha Jesudason, Rajiv Shah, James Shepherd, Antonia Ho, Emma Thomson; Alasdair MacLean, Rory Gunson. |
| EPI_ISL_425819, EPI_ISL_425820, EPI_ISL_425821, EPI_ISL_425822, EPI_ISL_425823, EPI_ISL_425824, EPI_ISL_425825, EPI_ISL_425826, EPI_ISL_425827, EPI_ISL_425828, EPI_ISL_425829, EPI_ISL_425830, EPI_ISL_425834, EPI_ISL_425836, EPI_ISL_425839, EPI_ISL_425842, EPI_ISL_425843, EPI_ISL_425844, EPI_ISL_425845, EPI_ISL_425846, EPI_ISL_425847, EPI_ISL_425848, EPI_ISL_425849, EPI_ISL_425850, EPI_ISL_425851, EPI_ISL_425852, EPI_ISL_425855, EPI_ISL_425856, EPI_ISL_425860, EPI_ISL_425861, EPI_ISL_425862, EPI_ISL_425863, EPI_ISL_425864, EPI_ISL_425865, EPI_ISL_425866, EPI_ISL_425867, EPI_ISL_425868, EPI_ISL_425869, EPI_ISL_425870, EPI_ISL_425871, EPI_ISL_425872, EPI_ISL_425873, EPI_ISL_425874, EPI_ISL_425875, EPI_ISL_425876, EPI_ISL_425877, EPI_ISL_425878, EPI_ISL_425879, EPI_ISL_425880, EPI_ISL_425881, EPI_ISL_425882, EPI_ISL_425883, EPI_ISL_425884, EPI_ISL_425885, EPI_ISL_425886, EPI_ISL_425887, EPI_ISL_425888, EPI_ISL_425889, EPI_ISL_425890, EPI_ISL_425891, EPI_ISL_425892, EPI_ISL_425893, EPI_ISL_425894, EPI_ISL_425895, EPI_ISL_425896, EPI_ISL_425897, EPI_ISL_425898, EPI_ISL_425899, EPI_ISL_425900, EPI_ISL_425901, EPI_ISL_425902, EPI_ISL_425903, EPI_ISL_425904, EPI_ISL_425905, EPI_ISL_425906, EPI_ISL_425907, EPI_ISL_425908, EPI_ISL_425911, EPI_ISL_425912, EPI_ISL_425913, EPI_ISL_425915, EPI_ISL_425916, EPI_ISL_425917, EPI_ISL_425918, EPI_ISL_425919, EPI_ISL_425920, EPI_ISL_425921, EPI_ISL_425922, EPI_ISL_425923, EPI_ISL_425924, EPI_ISL_425925, EPI_ISL_425926, EPI_ISL_425927, EPI_ISL_425928, EPI_ISL_425929, EPI_ISL_425930, EPI_ISL_425931, EPI_ISL_425932, EPI_ISL_425933, EPI_ISL_425934, EPI_ISL_425935, EPI_ISL_425936, EPI_ISL_425937, EPI_ISL_425938, EPI_ISL_425939, EPI_ISL_425940, EPI_ISL_425941, EPI_ISL_425942, EPI_ISL_425943, EPI_ISL_425944, EPI_ISL_425945, EPI_ISL_425946, EPI_ISL_425947, EPI_ISL_425948, EPI_ISL_425949, EPI_ISL_425950, EPI_ISL_425951, EPI_ISL_425952, EPI_ISL_425953, EPI_ISL_425954, EPI_ISL_425955, EPI_ISL_425956, EPI_ISL_425958, EPI_ISL_425959, EPI_ISL_425961, EPI_ISL_425962, EPI_ISL_425963, EPI_ISL_425964, EPI_ISL_425965, EPI_ISL_425966, EPI_ISL_425967, EPI_ISL_425968, EPI_ISL_425969, EPI_ISL_425971, EPI_ISL_425972, EPI_ISL_425973, EPI_ISL_425975, EPI_ISL_425976, EPI_ISL_425978, EPI_ISL_425979, EPI_ISL_425980, EPI_ISL_425982, EPI_ISL_425984, EPI_ISL_425985, EPI_ISL_425986, EPI_ISL_425987, EPI_ISL_425988, EPI_ISL_425990, EPI_ISL_425991, EPI_ISL_425992, EPI_ISL_425993, EPI_ISL_425994, EPI_ISL_425995, EPI_ISL_425996, EPI_ISL_425997, EPI_ISL_425998, EPI_ISL_425999, EPI_ISL_426000, EPI_ISL_426001, EPI_ISL_426002, EPI_ISL_426003, EPI_ISL_426004, EPI_ISL_426005, EPI_ISL_426006, EPI_ISL_426007, EPI_ISL_426008, EPI_ISL_426009, EPI_ISL_426010, EPI_ISL_426011, EPI_ISL_426012, EPI_ISL_426013, EPI_ISL_426014, EPI_ISL_426015, EPI_ISL_426016, EPI_ISL_426017, EPI_ISL_426019, EPI_ISL_426020, EPI_ISL_426021, EPI_ISL_426022, EPI_ISL_426023, EPI_ISL_426024 | | | |
| see above | Virology Department, Royal Infirmary of Edinburgh, NHS Lothian / School of Biological Sciences, University of Edinburgh / Institute of Genetics and Molecular Medicine, University of Edinburgh | COVID-19 Genomics UK (COG-UK) Consortium | McHugh M, Dewar R, Rooke S, Gallagher M, Balcaza C, O'Toole A, Hill V, McCrone JT, Colquhoun R, Yu X, Jackson B, Scher E, Rambaut A, Williams TC, Templeton K |
| EPI_ISL_432868, EPI_ISL_432869, EPI_ISL_432870, EPI_ISL_432871, EPI_ISL_432872, EPI_ISL_432873, EPI_ISL_432874, EPI_ISL_432875, EPI_ISL_432876, EPI_ISL_432877, EPI_ISL_432878, EPI_ISL_432880, EPI_ISL_432881, EPI_ISL_432883, EPI_ISL_432885, EPI_ISL_432886, EPI_ISL_432888, EPI_ISL_432889, EPI_ISL_432890, EPI_ISL_432891, EPI_ISL_432892, EPI_ISL_432893, EPI_ISL_432894, EPI_ISL_432895, EPI_ISL_432897, EPI_ISL_432898, EPI_ISL_432899, EPI_ISL_433068, EPI_ISL_433069, EPI_ISL_433070, EPI_ISL_433071, EPI_ISL_433072, EPI_ISL_433073, EPI_ISL_433075, EPI_ISL_433076, EPI_ISL_433077, EPI_ISL_433078, EPI_ISL_433079, EPI_ISL_433080, EPI_ISL_433081, EPI_ISL_433082, EPI_ISL_433083, EPI_ISL_433084, EPI_ISL_433085, EPI_ISL_433086, EPI_ISL_433087, EPI_ISL_433088, EPI_ISL_433089, EPI_ISL_433090, EPI_ISL_433091, EPI_ISL_433092, EPI_ISL_433093, EPI_ISL_433094, EPI_ISL_433095, EPI_ISL_433096, EPI_ISL_433097, EPI_ISL_433098, EPI_ISL_433099, EPI_ISL_433100, EPI_ISL_433101, EPI_ISL_433102, EPI_ISL_433104, EPI_ISL_433105, EPI_ISL_433107, EPI_ISL_433108, EPI_ISL_433109, EPI_ISL_433110, EPI_ISL_433111, EPI_ISL_433112, EPI_ISL_433113, EPI_ISL_433114, EPI_ISL_433115, EPI_ISL_433116, EPI_ISL_433117, EPI_ISL_433118, EPI_ISL_433119, EPI_ISL_433120, EPI_ISL_433121, EPI_ISL_433122, EPI_ISL_433123, EPI_ISL_433124, EPI_ISL_433125, EPI_ISL_433126, EPI_ISL_433127, EPI_ISL_433129, EPI_ISL_433130, EPI_ISL_433131, EPI_ISL_433132, EPI_ISL_433133, EPI_ISL_433135, EPI_ISL_433136, EPI_ISL_433137, EPI_ISL_433139, EPI_ISL_433140, EPI_ISL_433141, EPI_ISL_433142, EPI_ISL_433143, EPI_ISL_433145, EPI_ISL_433147, EPI_ISL_433148, EPI_ISL_433149, EPI_ISL_433150, EPI_ISL_433153, EPI_ISL_433154, EPI_ISL_433155, EPI_ISL_433156, EPI_ISL_433162, EPI_ISL_433164, EPI_ISL_433165, EPI_ISL_433166, EPI_ISL_433167, EPI_ISL_433168, EPI_ISL_433169, EPI_ISL_433170, EPI_ISL_433171, EPI_ISL_433172, EPI_ISL_433173, EPI_ISL_433174, EPI_ISL_433175, EPI_ISL_433176, EPI_ISL_433177, EPI_ISL_433178, EPI_ISL_433180, EPI_ISL_433181, EPI_ISL_433182, EPI_ISL_433183, EPI_ISL_433184, EPI_ISL_433185, EPI_ISL_433186, EPI_ISL_433187, EPI_ISL_433188, EPI_ISL_433190, EPI_ISL_433192, EPI_ISL_433193, EPI_ISL_433194, EPI_ISL_433195, EPI_ISL_433196, EPI_ISL_433197, EPI_ISL_433198, EPI_ISL_433200, EPI_ISL_433201, EPI_ISL_433202, EPI_ISL_433203, EPI_ISL_433204, EPI_ISL_433205, EPI_ISL_433206, EPI_ISL_433207, EPI_ISL_433208, EPI_ISL_433210, EPI_ISL_433211, EPI_ISL_433213, EPI_ISL_433214, EPI_ISL_433215, EPI_ISL_433216, EPI_ISL_433217, EPI_ISL_433218, EPI_ISL_433220, EPI_ISL_433221, EPI_ISL_433222, EPI_ISL_433223, EPI_ISL_433224, EPI_ISL_433225, EPI_ISL_433226, EPI_ISL_433227, EPI_ISL_433228, EPI_ISL_433229, EPI_ISL_433233, EPI_ISL_433234, EPI_ISL_433236, EPI_ISL_433237, EPI_ISL_433238, EPI_ISL_433240, EPI_ISL_433241, EPI_ISL_433242, EPI_ISL_433243, EPI_ISL_433244, EPI_ISL_433245, EPI_ISL_433246, EPI_ISL_433247, EPI_ISL_433248, EPI_ISL_433249, EPI_ISL_433250, EPI_ISL_433253, EPI_ISL_433254, EPI_ISL_433255, EPI_ISL_433256, EPI_ISL_433257, EPI_ISL_433260, EPI_ISL_433261, EPI_ISL_433262, EPI_ISL_433263, EPI_ISL_433265, EPI_ISL_433266, EPI_ISL_433267 | | | |
| see above | Virology Department, Royal Infirmary of Edinburgh, NHS Lothian / School of Biological Sciences, University of Edinburgh / Institute of Genetics and Molecular Medicine, University of Edinburgh | COVID-19 Genomics UK (COG-UK) Consortium | McHugh M, Dewar R, Rooke S, Gallagher M, Balcaza C, O'Toole A, Hill V, McCrone JT, Colquhoun R, Yu X, Jackson B, Rambaut A, Williams TC, Templeton K |
| EPI_ISL_433270, EPI_ISL_433274, EPI_ISL_433276, EPI_ISL_433277, EPI_ISL_433278, EPI_ISL_433279, EPI_ISL_433280, EPI_ISL_433281, EPI_ISL_433282, EPI_ISL_433283, EPI_ISL_433284, EPI_ISL_433285, EPI_ISL_433286, EPI_ISL_433287, EPI_ISL_433288, EPI_ISL_433289, EPI_ISL_433291, EPI_ISL_433293, EPI_ISL_433294, EPI_ISL_433295, EPI_ISL_433298, EPI_ISL_433299, EPI_ISL_433302, EPI_ISL_433303, EPI_ISL_433304, EPI_ISL_433305, EPI_ISL_433307, EPI_ISL_433308, EPI_ISL_433313, EPI_ISL_433314, EPI_ISL_433316, EPI_ISL_433321, EPI_ISL_433323, EPI_ISL_433324, EPI_ISL_433326, EPI_ISL_433327, EPI_ISL_433329, EPI_ISL_433330, EPI_ISL_433333, EPI_ISL_433334, EPI_ISL_433335, EPI_ISL_433338, EPI_ISL_433339, EPI_ISL_433340, EPI_ISL_433341, EPI_ISL_433342, EPI_ISL_433344, EPI_ISL_433346, EPI_ISL_433347, EPI_ISL_433348, EPI_ISL_433350, EPI_ISL_433351, EPI_ISL_433353, EPI_ISL_433355, EPI_ISL_433356, EPI_ISL_433357, EPI_ISL_433358, EPI_ISL_433359, EPI_ISL_433360, EPI_ISL_433361, EPI_ISL_433362, EPI_ISL_433364, EPI_ISL_433366, EPI_ISL_433367, EPI_ISL_433368, EPI_ISL_433370, EPI_ISL_433371, EPI_ISL_433372 | | | |
| see above | West of Scotland Specialist Virology Centre, NHSGGC / MRC-University of Glasgow Centre for Virus Research | COVID-19 Genomics UK (COG-UK) Consortium | Ana da Silva Filipe, Natasha Johnson, Kathy Smollett, Daniel Mair, Stephen Carmichael, Lily Tong, Jenna Nichols, Elihu Aranday-Cortes, Kirstyn Brunker, Yasmin Parr, Kyriaki Nomikou; Sarah McDonald, Marc Niebel, Patawee Asamaphan; Richard Orton, Joseph Hughes, Sreenu Vattipally, David L Robertson; Alasdair MacLean, Rory Gunson; Kathy Li, Natasha Jesudason, Rajiv Shah, James Shepherd, Antonia Ho, Emma Thomson |

We gratefully acknowledge the following Authors from the Originating laboratories responsible for obtaining the specimens, as well as the Submitting laboratories where the genome data were generated and shared via GISAID, on which this research is based.

All Submitters of data may be contacted directly via www.gisaid.org

Authors are sorted alphabetically.

| Accession ID | Originating Laboratory | Submitting Laboratory | Authors |
|---|---|---|---|
| EPI_ISL_1019507, EPI_ISL_1019508, EPI_ISL_1019509, EPI_ISL_1019510, EPI_ISL_1019511, EPI_ISL_1019512, EPI_ISL_1019513, EPI_ISL_1019514, EPI_ISL_1019516, EPI_ISL_1019518, EPI_ISL_1019519, EPI_ISL_1019520, EPI_ISL_1019521, EPI_ISL_1019522, EPI_ISL_1019523, EPI_ISL_1019524, EPI_ISL_1019525, EPI_ISL_1019526, EPI_ISL_1019527, EPI_ISL_1019528, EPI_ISL_1019529, EPI_ISL_1019530, EPI_ISL_1019531, EPI_ISL_1019532, EPI_ISL_1019533, EPI_ISL_1019534, EPI_ISL_1019535, EPI_ISL_1019536, EPI_ISL_1019537, EPI_ISL_1019538, EPI_ISL_1019539, EPI_ISL_1019540, EPI_ISL_1019541, EPI_ISL_1019542, EPI_ISL_1019543, EPI_ISL_1019544, EPI_ISL_1019545, EPI_ISL_1019546, EPI_ISL_1019547, EPI_ISL_1019548, EPI_ISL_1019549, EPI_ISL_1019550, EPI_ISL_1019551, EPI_ISL_1019552, EPI_ISL_1019553, EPI_ISL_1019554, EPI_ISL_1019555, EPI_ISL_1019556, EPI_ISL_1019557, EPI_ISL_1019558, EPI_ISL_1019559, EPI_ISL_1019560, EPI_ISL_1019561, EPI_ISL_1019562, EPI_ISL_1019563, EPI_ISL_1019564, EPI_ISL_1019565, EPI_ISL_1019566, EPI_ISL_1019567, EPI_ISL_1019568, EPI_ISL_1019569, EPI_ISL_1019570, EPI_ISL_1019571, EPI_ISL_1019572, EPI_ISL_1019573, EPI_ISL_1019574, EPI_ISL_1019575, EPI_ISL_1019577, EPI_ISL_1019578, EPI_ISL_1019579, EPI_ISL_1019580, EPI_ISL_1019581, EPI_ISL_1019582, EPI_ISL_1019583, EPI_ISL_1019584, EPI_ISL_1019585 | | | |
| see above | Lighthouse Lab in Glasgow | Wellcome Sanger Institute for the COVID-19 Genomics UK (COG-UK) Consortium | Harper VanSteenhouse, Yumi Kasai, David Gray, Carol Clugston, Anna Dominiczak and Alex Alderton, Roberto Amato, Jeffrey Barrett, Sonia Goncalves, Ewan Harrison, David K. Jackson, Ian Johnston, Dominic Kwiatkowski, Cordelia Langford, John Sillitoe on behalf of the Wellcome Sanger Institute COVID-19 Surveillance Team |
| EPI_ISL_1019586 | Lighthouse Lab in Milton Keynes | Wellcome Sanger Institute for the COVID-19 Genomics UK (COG-UK) Consortium | The Lighthouse Lab in Milton Keynes and Alex Alderton, Roberto Amato, Jeffrey Barrett, Sonia Goncalves, Ewan Harrison, David K. Jackson, Ian Johnston, Dominic Kwiatkowski, Cordelia Langford, John Sillitoe on behalf of the Wellcome Sanger Institute COVID-19 Surveillance Team |
| EPI_ISL_1019587, EPI_ISL_1019588, EPI_ISL_1019589, EPI_ISL_1019590, EPI_ISL_1019592, EPI_ISL_1019593, EPI_ISL_1019595, EPI_ISL_1019596, EPI_ISL_1019597, EPI_ISL_1019598, EPI_ISL_1019600, EPI_ISL_1019601, EPI_ISL_1019603, EPI_ISL_1019604, EPI_ISL_1019605, EPI_ISL_1019606, EPI_ISL_1019607, EPI_ISL_1019608, EPI_ISL_1019609, EPI_ISL_1019611, EPI_ISL_1019612, EPI_ISL_1019613, EPI_ISL_1019614, EPI_ISL_1019615, EPI_ISL_1019616, EPI_ISL_1019617, EPI_ISL_1019618, EPI_ISL_1019619, EPI_ISL_1019620, EPI_ISL_1019621, EPI_ISL_1019622, EPI_ISL_1019623, EPI_ISL_1019624, EPI_ISL_1019625, EPI_ISL_1019626, EPI_ISL_1019627, EPI_ISL_1019628, EPI_ISL_1019629, EPI_ISL_1019630, EPI_ISL_1019631, EPI_ISL_1019632, EPI_ISL_1019633, EPI_ISL_1019634, EPI_ISL_1019635, EPI_ISL_1019636, EPI_ISL_1019638, EPI_ISL_1019639, EPI_ISL_1019641, EPI_ISL_1019643, EPI_ISL_1019644, EPI_ISL_1019646, EPI_ISL_1019648, EPI_ISL_1019649, EPI_ISL_1019651, EPI_ISL_1019652, EPI_ISL_1019654, EPI_ISL_1019656, EPI_ISL_1019657, EPI_ISL_1019659, EPI_ISL_1019661, EPI_ISL_1019666, EPI_ISL_1019667, EPI_ISL_1019669, EPI_ISL_1019670, EPI_ISL_1019673, EPI_ISL_1019675, EPI_ISL_1019677, EPI_ISL_1019679, EPI_ISL_1019680, EPI_ISL_1019682, EPI_ISL_1019684, EPI_ISL_1019685, EPI_ISL_1019687, EPI_ISL_1019689, EPI_ISL_1019690, EPI_ISL_1019692, EPI_ISL_1019694, EPI_ISL_1019695, EPI_ISL_1019697, EPI_ISL_1019698, EPI_ISL_1019701, EPI_ISL_1019703, EPI_ISL_1019704, EPI_ISL_1019706, EPI_ISL_1019708, EPI_ISL_1019709, EPI_ISL_1019711, EPI_ISL_1019714, EPI_ISL_1019717, EPI_ISL_1019719, EPI_ISL_1019721, EPI_ISL_1019723, EPI_ISL_1019727, EPI_ISL_1019728, EPI_ISL_1019730, EPI_ISL_1019733, EPI_ISL_1019735, EPI_ISL_1019736, EPI_ISL_1019738, EPI_ISL_1019740, EPI_ISL_1019741, EPI_ISL_1019743, EPI_ISL_1019744, EPI_ISL_1019746, EPI_ISL_1019747, EPI_ISL_1019749, EPI_ISL_1019751, EPI_ISL_1019752, EPI_ISL_1019754, EPI_ISL_1019755, EPI_ISL_1019757, EPI_ISL_1019760, EPI_ISL_1019761, EPI_ISL_1019765, EPI_ISL_1019766, EPI_ISL_1019769, EPI_ISL_1019773, EPI_ISL_1019774, EPI_ISL_1019776, EPI_ISL_1019777, EPI_ISL_1019781, EPI_ISL_1019782, EPI_ISL_1019784, EPI_ISL_1019786, EPI_ISL_1019789, EPI_ISL_1019791, EPI_ISL_1019792, EPI_ISL_1019794, EPI_ISL_1019796, EPI_ISL_1019797, EPI_ISL_1019802, EPI_ISL_1019804, EPI_ISL_1019806, EPI_ISL_1019809, EPI_ISL_1019811, EPI_ISL_1019812, EPI_ISL_1019814, EPI_ISL_1019816, EPI_ISL_1019817, EPI_ISL_1019819, EPI_ISL_1019821, EPI_ISL_1019824, EPI_ISL_1019826, EPI_ISL_1019829, EPI_ISL_1019831, EPI_ISL_1019832, EPI_ISL_1019835, EPI_ISL_1019839, EPI_ISL_1019840, EPI_ISL_1019844, EPI_ISL_1019845, EPI_ISL_1019848, EPI_ISL_1019850, EPI_ISL_1019854, EPI_ISL_1019857, EPI_ISL_1019859, EPI_ISL_1019860, EPI_ISL_1019862, EPI_ISL_1019864, EPI_ISL_1019868, EPI_ISL_1019870, EPI_ISL_1019871, EPI_ISL_1019873, EPI_ISL_1019876, EPI_ISL_1019878, EPI_ISL_1019879, EPI_ISL_1019881, EPI_ISL_1019883, EPI_ISL_1019884, EPI_ISL_1019886, EPI_ISL_1019887, EPI_ISL_1019889, EPI_ISL_1019891, EPI_ISL_1019892, EPI_ISL_1019894, EPI_ISL_1019895, EPI_ISL_1019896, EPI_ISL_1019898, EPI_ISL_1019899, EPI_ISL_1019900, EPI_ISL_1019902, EPI_ISL_1019904, EPI_ISL_1019905, EPI_ISL_1019907, EPI_ISL_1019909, EPI_ISL_1019910, EPI_ISL_1019913, EPI_ISL_1019914, EPI_ISL_1019915, EPI_ISL_1019917, EPI_ISL_1019918, EPI_ISL_1019919, EPI_ISL_1019921, EPI_ISL_1019922, EPI_ISL_1019923, EPI_ISL_1019924, EPI_ISL_1019925, EPI_ISL_1019927, EPI_ISL_1019928, EPI_ISL_1019929, EPI_ISL_1019930, EPI_ISL_1019931, EPI_ISL_1019932, EPI_ISL_1019934, EPI_ISL_1019935, EPI_ISL_1019937, EPI_ISL_1019938, EPI_ISL_1019939, EPI_ISL_1019940, EPI_ISL_1019941, EPI_ISL_1019942, EPI_ISL_1019943, EPI_ISL_1019944, EPI_ISL_1019946, EPI_ISL_1019947, EPI_ISL_1019948, EPI_ISL_1019949, EPI_ISL_1019950, EPI_ISL_1019951, EPI_ISL_1019953, EPI_ISL_1019954, EPI_ISL_1019955, EPI_ISL_1019956, EPI_ISL_1019957, EPI_ISL_1019958, EPI_ISL_1019959, EPI_ISL_1019961, EPI_ISL_1019964, EPI_ISL_1019965, EPI_ISL_1019967, EPI_ISL_1019969, EPI_ISL_1019970, EPI_ISL_1019971, EPI_ISL_1019972, EPI_ISL_1019973, EPI_ISL_1019975, EPI_ISL_1019976, EPI_ISL_1019978, EPI_ISL_1019980, EPI_ISL_1019981, EPI_ISL_1019982, EPI_ISL_1019983, EPI_ISL_1019984, EPI_ISL_1019985, EPI_ISL_1019986, EPI_ISL_1019987, EPI_ISL_1019988, EPI_ISL_1019989, EPI_ISL_1019990, EPI_ISL_1019991, EPI_ISL_1019992, EPI_ISL_1019993, EPI_ISL_1019994, EPI_ISL_1019995, EPI_ISL_1019996, EPI_ISL_1019997, EPI_ISL_1019998, EPI_ISL_1019999, EPI_ISL_1020001, EPI_ISL_1020003, EPI_ISL_1020004, EPI_ISL_1020005, EPI_ISL_1020006, EPI_ISL_1020007, EPI_ISL_1020008, EPI_ISL_1020009, EPI_ISL_1020010, EPI_ISL_1020011, EPI_ISL_1020012, EPI_ISL_1020015, EPI_ISL_1042390, EPI_ISL_1042572 | | | |
| see above | Lighthouse Lab in Glasgow | Wellcome Sanger Institute for the COVID-19 Genomics UK (COG-UK) Consortium | Harper VanSteenhouse, Yumi Kasai, David Gray, Carol Clugston, Anna Dominiczak and Alex Alderton, Roberto Amato, Jeffrey Barrett, Sonia Goncalves, Ewan Harrison, David K. Jackson, Ian Johnston, Dominic Kwiatkowski, Cordelia Langford, John Sillitoe on behalf of the Wellcome Sanger Institute COVID-19 Surveillance Team |
| EPI_ISL_1042822 | Lighthouse Lab in Milton Keynes | Wellcome Sanger Institute for the COVID-19 Genomics UK (COG-UK) Consortium | The Lighthouse Lab in Milton Keynes and Alex Alderton, Roberto Amato, Jeffrey Barrett, Sonia Goncalves, Ewan Harrison, David K. Jackson, Ian Johnston, Dominic Kwiatkowski, Cordelia Langford, John Sillitoe on behalf of the Wellcome Sanger Institute COVID-19 Surveillance Team |
| EPI_ISL_1043461 | Lighthouse Lab in Glasgow | Wellcome Sanger Institute for the COVID-19 Genomics UK (COG-UK) Consortium | Harper VanSteenhouse, Yumi Kasai, David Gray, Carol Clugston, Anna Dominiczak and Alex Alderton, Roberto Amato, Jeffrey Barrett, Sonia Goncalves, Ewan Harrison, David K. Jackson, Ian Johnston, Dominic Kwiatkowski, Cordelia Langford, John Sillitoe on behalf of the Wellcome Sanger Institute COVID-19 Surveillance Team |
| EPI_ISL_1047804, EPI_ISL_1047805, EPI_ISL_1047806, EPI_ISL_1047807, EPI_ISL_1047809, EPI_ISL_1047810, EPI_ISL_1047811, EPI_ISL_1047812, EPI_ISL_1047813, EPI_ISL_1047814, EPI_ISL_1047815, EPI_ISL_1047816, EPI_ISL_1047818, EPI_ISL_1047819, EPI_ISL_1047820, EPI_ISL_1047821, EPI_ISL_1047822, EPI_ISL_1047823, EPI_ISL_1047824, EPI_ISL_1047825, EPI_ISL_1047827, EPI_ISL_1047828, EPI_ISL_1047829, EPI_ISL_1047830, EPI_ISL_1047831, EPI_ISL_1047832, EPI_ISL_1047833, EPI_ISL_1047834, EPI_ISL_1047835, EPI_ISL_1047836, EPI_ISL_1047837, EPI_ISL_1047838, EPI_ISL_1047839, EPI_ISL_1047840, EPI_ISL_1047841, EPI_ISL_1047842, EPI_ISL_1047843, EPI_ISL_1047846, EPI_ISL_1047847, EPI_ISL_1047848, EPI_ISL_1047849, EPI_ISL_1047850, EPI_ISL_1047851, EPI_ISL_1047852, EPI_ISL_1047853, EPI_ISL_1047854 | | | |
| see above | West of Scotland Specialist Virology Centre, NHSGGC / MRC-University of Glasgow Centre for Virus Research | COVID-19 Genomics UK (COG-UK) Consortium | Ana da Silva Filipe, Natasha Johnson, Kathy Smollett, Daniel Mair, Stephen Carmichael, Alice Broos, Lily Tong, Jenna Nichols, Kyriaki Nomikou; Sarah McDonald; Richard Orton, Joseph Hughes, Sreenu Vattipally, David L Robertson; Alasdair MacLean, Rory Gunson; Sharif Shaaban, Matthew Holden; Rachel Blacow, Guy Mollett, Kathy Li, James Shepherd, Antonia Ho, Emma Thomson |
| EPI_ISL_1047857, EPI_ISL_1047858, EPI_ISL_1047859, EPI_ISL_1047860, EPI_ISL_1047861, EPI_ISL_1047862, EPI_ISL_1047863, EPI_ISL_1047864, EPI_ISL_1047865, EPI_ISL_1047866, EPI_ISL_1047867, EPI_ISL_1047868, EPI_ISL_1047869, EPI_ISL_1047871, EPI_ISL_1047872, EPI_ISL_1047873, EPI_ISL_1047874, EPI_ISL_1047875, EPI_ISL_1047876, EPI_ISL_1047878, EPI_ISL_1047879, EPI_ISL_1047880, EPI_ISL_1047881, EPI_ISL_1047882, EPI_ISL_1047883, EPI_ISL_1047884, EPI_ISL_1047888, EPI_ISL_1047889, EPI_ISL_1047890, EPI_ISL_1047891, EPI_ISL_1047892, EPI_ISL_1047893, EPI_ISL_1047894, EPI_ISL_1047895, EPI_ISL_1047899, EPI_ISL_1047900, EPI_ISL_1047901, EPI_ISL_1047902, EPI_ISL_1047903, EPI_ISL_1047904, EPI_ISL_1047905, EPI_ISL_1047906, EPI_ISL_1047907, EPI_ISL_1047908, EPI_ISL_1047909, EPI_ISL_1047910, EPI_ISL_1047912, EPI_ISL_1047913 | | | |
| see above | Virology Department, Royal Infirmary of Edinburgh, NHS Lothian / School of Biological Sciences, University of Edinburgh | COVID-19 Genomics UK (COG-UK) Consortium | McHugh M, Dewar R, Cotton S, Rooke S, O'Toole Á, Scher E, Hill V, McCrone JT, Colquhoun R, Yu X, Jackson B, Rambaut A, Templeton K |
| EPI_ISL_1050174, EPI_ISL_1050175, EPI_ISL_1050176, EPI_ISL_1050177, EPI_ISL_1050178, EPI_ISL_1050179, EPI_ISL_1050180, EPI_ISL_1050181, EPI_ISL_1050182, EPI_ISL_1050183, EPI_ISL_1050184, EPI_ISL_1050185, EPI_ISL_1050186, EPI_ISL_1050187, EPI_ISL_1050188, EPI_ISL_1050189, EPI_ISL_1050190, EPI_ISL_1050191, EPI_ISL_1050192, EPI_ISL_1050193, EPI_ISL_1050194, EPI_ISL_1050195, EPI_ISL_1050196, EPI_ISL_1050197, EPI_ISL_1050198, EPI_ISL_1050199, EPI_ISL_1050200, EPI_ISL_1050201, EPI_ISL_1050202, EPI_ISL_1050203, EPI_ISL_1050204, EPI_ISL_1050205, EPI_ISL_1050206, EPI_ISL_1050207, EPI_ISL_1050209, EPI_ISL_1050210, EPI_ISL_1050211, EPI_ISL_1050212, EPI_ISL_1050213, EPI_ISL_1050214, EPI_ISL_1050215, EPI_ISL_1050216, EPI_ISL_1050218, EPI_ISL_1050219, EPI_ISL_1050220, EPI_ISL_1050221, EPI_ISL_1050222 | | | |
| see above | West of Scotland Specialist Virology Centre, NHSGGC / MRC-University of Glasgow Centre for Virus Research | COVID-19 Genomics UK (COG-UK) Consortium | Ana da Silva Filipe, Natasha Johnson, Kathy Smollett, Daniel Mair, Stephen Carmichael, Alice Broos, Lily Tong, Jenna Nichols, Kyriaki Nomikou; Sarah McDonald; Richard Orton, Joseph Hughes, Sreenu Vattipally, David L Robertson; Alasdair MacLean, Rory Gunson; Sharif Shaaban, Matthew Holden; Rachel Blacow, Guy Mollett, Kathy Li, James Shepherd, Antonia Ho, Emma Thomson |
| EPI_ISL_1050223, EPI_ISL_1050225, EPI_ISL_1050226, EPI_ISL_1050227, EPI_ISL_1050228, EPI_ISL_1050229, EPI_ISL_1050230, EPI_ISL_1050231, EPI_ISL_1050232 | Lighthouse Lab in Glasgow / MRC-University of Glasgow Centre for Virus Research | COVID-19 Genomics UK (COG-UK) Consortium | Ana da Silva Filipe, Natasha Johnson, Kathy Smollett, Daniel Mair, Stephen Carmichael, Alice Broos, Lily Tong, Jenna Nichols, Kyriaki Nomikou; Sarah McDonald; Harper VanSteenhouse, Yumi Kasai, David Gray, Carol Clugston, Anna Dominiczak; Alasdair MacLean, Rory Gunson; Richard Orton, Joseph Hughes, Sreenu Vattipally, David L Robertson; Sharif Shaaban, Matthew Holden; Kathy Li, James Shepherd, Antonia Ho, Emma Thomson |
| EPI_ISL_1050234, EPI_ISL_1050235, EPI_ISL_1050236, EPI_ISL_1050237, EPI_ISL_1050238 | West of Scotland Specialist Virology Centre, NHSGGC / MRC-University of Glasgow Centre for Virus Research | COVID-19 Genomics UK (COG-UK) Consortium | Ana da Silva Filipe, Natasha Johnson, Kathy Smollett, Daniel Mair, Stephen Carmichael, Alice Broos, Lily Tong, Jenna Nichols, Kyriaki Nomikou; Sarah McDonald; Richard Orton, Joseph Hughes, Sreenu Vattipally, David L Robertson; Alasdair MacLean, Rory Gunson; Sharif Shaaban, Matthew Holden; Rachel Blacow, Guy Mollett, Kathy Li, James Shepherd, Antonia Ho, Emma Thomson |
| EPI_ISL_1050239 | Lighthouse Lab in Glasgow / MRC-University of Glasgow Centre for Virus Research | COVID-19 Genomics UK (COG-UK) Consortium | Ana da Silva Filipe, Natasha Johnson, Kathy Smollett, Daniel Mair, Stephen Carmichael, Alice Broos, Lily Tong, Jenna Nichols, Kyriaki Nomikou; Sarah McDonald; Harper VanSteenhouse, Yumi Kasai, David Gray, Carol Clugston, Anna Dominiczak; Alasdair MacLean, Rory Gunson; Richard Orton, Joseph Hughes, Sreenu Vattipally, David L Robertson; Sharif Shaaban, Matthew Holden; Kathy Li, James Shepherd, Antonia Ho, Emma Thomson |
| EPI_ISL_1050240, EPI_ISL_1050241 | West of Scotland Specialist Virology Centre, NHSGGC / MRC-University of Glasgow Centre for Virus Research | COVID-19 Genomics UK (COG-UK) Consortium | Ana da Silva Filipe, Natasha Johnson, Kathy Smollett, Daniel Mair, Stephen Carmichael, Alice Broos, Lily Tong, Jenna Nichols, Kyriaki Nomikou; Sarah McDonald; Richard Orton, Joseph Hughes, Sreenu Vattipally, David L Robertson; Alasdair MacLean, Rory Gunson; Sharif Shaaban, Matthew Holden; Rachel Blacow, Guy Mollett, Kathy Li, James Shepherd, Antonia Ho, Emma Thomson |
| EPI_ISL_1050243, EPI_ISL_1050244 | Virology Department, Royal Infirmary of Edinburgh, NHS | COVID-19 Genomics UK (COG-UK) Consortium | McHugh M, Dewar R, Cotton S, Rooke S, O'Toole Á, Scher E, Hill V, McCrone JT, Colquhoun R, Yu X, Jackson B, Rambaut A, Templeton K |

We gratefully acknowledge the following Authors from the Originating laboratories responsible for obtaining the specimens, as well as the Submitting laboratories where the genome data were generated and shared via GISAID, on which this research is based.

All Submitters of data may be contacted directly via www.gisaid.org

Authors are sorted alphabetically.

| Accession ID | Originating Laboratory | Submitting Laboratory | Authors |
|---|---|---|---|
| EPI_ISL_1186740, EPI_ISL_1190342, EPI_ISL_1205538, EPI_ISL_1207809, EPI_ISL_1207810, EPI_ISL_1207811, EPI_ISL_1207812, EPI_ISL_1207813, EPI_ISL_1207814, EPI_ISL_1207815, EPI_ISL_1207816, EPI_ISL_1207817, EPI_ISL_1207818, EPI_ISL_1207819, EPI_ISL_1207820, EPI_ISL_1207821, EPI_ISL_1207822, EPI_ISL_1207823, EPI_ISL_1207824, EPI_ISL_1207825, EPI_ISL_1207826, EPI_ISL_1207827, EPI_ISL_1207828, EPI_ISL_1207830, EPI_ISL_1207831, EPI_ISL_1207832, EPI_ISL_1207833, EPI_ISL_1207834, EPI_ISL_1207835, EPI_ISL_1207836, EPI_ISL_1207837, EPI_ISL_1207838, EPI_ISL_1207839, EPI_ISL_1207840, EPI_ISL_1207841, EPI_ISL_1207842, EPI_ISL_1207843, EPI_ISL_1207844, EPI_ISL_1207845, EPI_ISL_1207846, EPI_ISL_1207847, EPI_ISL_1207848, EPI_ISL_1207849, EPI_ISL_1207850, EPI_ISL_1207851, EPI_ISL_1207852, EPI_ISL_1207853, EPI_ISL_1207854, EPI_ISL_1207855, EPI_ISL_1207857, EPI_ISL_1207858, EPI_ISL_1207859, EPI_ISL_1207861, EPI_ISL_1207862, EPI_ISL_1207863, EPI_ISL_1207864, EPI_ISL_1207865, EPI_ISL_1207866, EPI_ISL_1207867, EPI_ISL_1207868, EPI_ISL_1207869, EPI_ISL_1207870, EPI_ISL_1207871, EPI_ISL_1207872, EPI_ISL_1207874, EPI_ISL_1207875, EPI_ISL_1207876, EPI_ISL_1207877, EPI_ISL_1207878, EPI_ISL_1207879, EPI_ISL_1207880, EPI_ISL_1207881, EPI_ISL_1207882, EPI_ISL_1207883, EPI_ISL_1207884, EPI_ISL_1207885, EPI_ISL_1207886, EPI_ISL_1207887, EPI_ISL_1207889, EPI_ISL_1207890, EPI_ISL_1207891, EPI_ISL_1207892, EPI_ISL_1207893, EPI_ISL_1207894, EPI_ISL_1207895, EPI_ISL_1207897, EPI_ISL_1207898, EPI_ISL_1207899, EPI_ISL_1207900, EPI_ISL_1207901, EPI_ISL_1207902, EPI_ISL_1207903, EPI_ISL_1207904, EPI_ISL_1207905, EPI_ISL_1207906, EPI_ISL_1207907, EPI_ISL_1207908, EPI_ISL_1207909, EPI_ISL_1207910, EPI_ISL_1208007, EPI_ISL_1208048, EPI_ISL_1208049, EPI_ISL_1208050, EPI_ISL_1208051, EPI_ISL_1208052, EPI_ISL_1208053, EPI_ISL_1208055, EPI_ISL_1208056, EPI_ISL_1208057, EPI_ISL_1208058, EPI_ISL_1208060, EPI_ISL_1208061, EPI_ISL_1208062, EPI_ISL_1208063, EPI_ISL_1208064, EPI_ISL_1208065, EPI_ISL_1208066, EPI_ISL_1208067, EPI_ISL_1208069, EPI_ISL_1208070, EPI_ISL_1208071, EPI_ISL_1208072, EPI_ISL_1208073, EPI_ISL_1208074, EPI_ISL_1208075, EPI_ISL_1208076, EPI_ISL_1208078, EPI_ISL_1208079, EPI_ISL_1208080, EPI_ISL_1208081, EPI_ISL_1208082, EPI_ISL_1208083, EPI_ISL_1208084, EPI_ISL_1208085, EPI_ISL_1208086, EPI_ISL_1208087, EPI_ISL_1208088, EPI_ISL_1208089, EPI_ISL_1208091, EPI_ISL_1208092, EPI_ISL_1208093, EPI_ISL_1208094, EPI_ISL_1208095, EPI_ISL_1208096, EPI_ISL_1208098, EPI_ISL_1208099, EPI_ISL_1208101, EPI_ISL_1208103, EPI_ISL_1208104, EPI_ISL_1208105, EPI_ISL_1208107, EPI_ISL_1208108, EPI_ISL_1208109, EPI_ISL_1208111, EPI_ISL_1208112, EPI_ISL_1208113, EPI_ISL_1208114, EPI_ISL_1208115, EPI_ISL_1208116, EPI_ISL_1208265, EPI_ISL_1208267, EPI_ISL_1208268, EPI_ISL_1208269, EPI_ISL_1208270, EPI_ISL_1208271, EPI_ISL_1208272, EPI_ISL_1208273, EPI_ISL_1208274, EPI_ISL_1208275, EPI_ISL_1208276, EPI_ISL_1208277, EPI_ISL_1208278, EPI_ISL_1208279, EPI_ISL_1208280, EPI_ISL_1208281, EPI_ISL_1208283, EPI_ISL_1208284, EPI_ISL_1208285, EPI_ISL_1208286, EPI_ISL_1208287, EPI_ISL_1208288, EPI_ISL_1208289, EPI_ISL_1208290, EPI_ISL_1208291, EPI_ISL_1208292, EPI_ISL_1208293, EPI_ISL_1208295, EPI_ISL_1208296, EPI_ISL_1208299, EPI_ISL_1208300, EPI_ISL_1208301, EPI_ISL_1208303, EPI_ISL_1208304, EPI_ISL_1208305, EPI_ISL_1208306, EPI_ISL_1208307, EPI_ISL_1208308, EPI_ISL_1208310, EPI_ISL_1208312, EPI_ISL_1208313, EPI_ISL_1208314, EPI_ISL_1208315, EPI_ISL_1208316, EPI_ISL_1208317, EPI_ISL_1208318, EPI_ISL_1208319, EPI_ISL_1208320, EPI_ISL_1208322, EPI_ISL_1208323, EPI_ISL_1208324, EPI_ISL_1208326, EPI_ISL_1208327, EPI_ISL_1208328, EPI_ISL_1208329, EPI_ISL_1208330, EPI_ISL_1208331, EPI_ISL_1208332, EPI_ISL_1208334, EPI_ISL_1208335, EPI_ISL_1208336, EPI_ISL_1208337, EPI_ISL_1208340, EPI_ISL_1208341, EPI_ISL_1208342, EPI_ISL_1208343, EPI_ISL_1208344, EPI_ISL_1208347, EPI_ISL_1208348, EPI_ISL_1208349, EPI_ISL_1208350, EPI_ISL_1208351, EPI_ISL_1208354, EPI_ISL_1222950, EPI_ISL_1222987, EPI_ISL_1243144, EPI_ISL_1244644, EPI_ISL_1245011, EPI_ISL_1245013, EPI_ISL_1245018, EPI_ISL_1245024, EPI_ISL_1245026, EPI_ISL_1245027, EPI_ISL_1245030, EPI_ISL_1245035, EPI_ISL_1245036, EPI_ISL_1245040, EPI_ISL_1245042, EPI_ISL_1245043, EPI_ISL_1245044, EPI_ISL_1245046, EPI_ISL_1245047, EPI_ISL_1245048, EPI_ISL_1245051, EPI_ISL_1245054, EPI_ISL_1245060, EPI_ISL_1245061, EPI_ISL_1245063, EPI_ISL_1245067, EPI_ISL_1245073, EPI_ISL_1245076, EPI_ISL_1245079, EPI_ISL_1245081, EPI_ISL_1245085, EPI_ISL_1245087, EPI_ISL_1245088, EPI_ISL_1245094, EPI_ISL_1245095, EPI_ISL_1245101, EPI_ISL_1245102, EPI_ISL_1245108, EPI_ISL_1245110, EPI_ISL_1245111, EPI_ISL_1245117, EPI_ISL_1245123, EPI_ISL_1245125, EPI_ISL_1245127, EPI_ISL_1245128, EPI_ISL_1245129, EPI_ISL_1245130, EPI_ISL_1245132, EPI_ISL_1245133, EPI_ISL_1245134, EPI_ISL_1245135, EPI_ISL_1245138, EPI_ISL_1245143, EPI_ISL_1245144, EPI_ISL_1245146, EPI_ISL_1245149, EPI_ISL_1245151, EPI_ISL_1245152, EPI_ISL_1245155, EPI_ISL_1245161, EPI_ISL_1245166, EPI_ISL_1245167, EPI_ISL_1245169, EPI_ISL_1245176, EPI_ISL_1245180, EPI_ISL_1245181, EPI_ISL_1245182, EPI_ISL_1245186, EPI_ISL_1245187, EPI_ISL_1245190, EPI_ISL_1245193, EPI_ISL_1245194, EPI_ISL_1245198, EPI_ISL_1245202, EPI_ISL_1245203, EPI_ISL_1245206, EPI_ISL_1245207, EPI_ISL_1245212, EPI_ISL_1245216, EPI_ISL_1245217, EPI_ISL_1245218, EPI_ISL_1245224, EPI_ISL_1245226, EPI_ISL_1245228, EPI_ISL_1245234, EPI_ISL_1245235, EPI_ISL_1245236, EPI_ISL_1245241, EPI_ISL_1245244, EPI_ISL_1245245, EPI_ISL_1245246, EPI_ISL_1245247, EPI_ISL_1245249, EPI_ISL_1245256, EPI_ISL_1245257, EPI_ISL_1245259, EPI_ISL_1245261, EPI_ISL_1245266, EPI_ISL_1245267, EPI_ISL_1245268, EPI_ISL_1245270, EPI_ISL_1245275, EPI_ISL_1245276, EPI_ISL_1245277, EPI_ISL_1245279, EPI_ISL_1245280, EPI_ISL_1245282, EPI_ISL_1245290, EPI_ISL_1245292, EPI_ISL_1245293, EPI_ISL_1245297, EPI_ISL_1245298, EPI_ISL_1245300, EPI_ISL_1245301, EPI_ISL_1245302, EPI_ISL_1245304, EPI_ISL_1245305, EPI_ISL_1245308, EPI_ISL_1245313, EPI_ISL_1245314, EPI_ISL_1245316, EPI_ISL_1245317, EPI_ISL_1245319, EPI_ISL_1245321, EPI_ISL_1245326, EPI_ISL_1245328, EPI_ISL_1245330, EPI_ISL_1245333, EPI_ISL_1245334, EPI_ISL_1245335, EPI_ISL_1245336, EPI_ISL_1245339, EPI_ISL_1245344, EPI_ISL_1245346, EPI_ISL_1245349, EPI_ISL_1245352, EPI_ISL_1245355, EPI_ISL_1245357 | | | |
| see above | Lighthouse Lab in Glasgow | Wellcome Sanger Institute for the COVID-19 Genomics UK (COG-UK) Consortium | Harper VanSteenhouse, Yumi Kasai, David Gray, Carol Clugston, Anna Dominiczak and Alex Alderton, Roberto Amato, Jeffrey Barrett, Sonia Goncalves, Ewan Harrison, David K. Jackson, Ian Johnston, Dominic Kwiatkowski, Cordelia Langford, John Sillitoe on behalf of the Wellcome Sanger Institute COVID-19 Surveillance Team |
| EPI_ISL_1245587 | Lighthouse Lab in Milton Keynes | Wellcome Sanger Institute for the COVID-19 Genomics UK (COG-UK) Consortium | The Lighthouse Lab in Milton Keynes and Alex Alderton, Roberto Amato, Jeffrey Barrett, Sonia Goncalves, Ewan Harrison, David K. Jackson, Ian Johnston, Dominic Kwiatkowski, Cordelia Langford, John Sillitoe on behalf of the Wellcome Sanger Institute COVID-19 Surveillance Team |
| EPI_ISL_1246289, EPI_ISL_1246290, EPI_ISL_1246291, EPI_ISL_1246292, EPI_ISL_1246293, EPI_ISL_1246294, EPI_ISL_1246295, EPI_ISL_1246296, EPI_ISL_1246297, EPI_ISL_1246298, EPI_ISL_1246299, EPI_ISL_1246300, EPI_ISL_1246301, EPI_ISL_1246302, EPI_ISL_1246303, EPI_ISL_1246304, EPI_ISL_1246305, EPI_ISL_1246306, EPI_ISL_1246307, EPI_ISL_1246308, EPI_ISL_1246309, EPI_ISL_1246310, EPI_ISL_1246311, EPI_ISL_1246312, EPI_ISL_1246313, EPI_ISL_1246314, EPI_ISL_1246315, EPI_ISL_1246316, EPI_ISL_1246317, EPI_ISL_1246318, EPI_ISL_1246319, EPI_ISL_1246320, EPI_ISL_1246321, EPI_ISL_1246322, EPI_ISL_1246323, EPI_ISL_1246325, EPI_ISL_1246326, EPI_ISL_1246327, EPI_ISL_1246328, EPI_ISL_1246329, EPI_ISL_1246330, EPI_ISL_1246331, EPI_ISL_1246332, EPI_ISL_1246333, EPI_ISL_1246334, EPI_ISL_1246335, EPI_ISL_1246336, EPI_ISL_1246337, EPI_ISL_1246338, EPI_ISL_1246339, EPI_ISL_1246340, EPI_ISL_1246341, EPI_ISL_1246342, EPI_ISL_1246343, EPI_ISL_1246344, EPI_ISL_1246345, EPI_ISL_1246346, EPI_ISL_1246347, EPI_ISL_1246348, EPI_ISL_1246349, EPI_ISL_1246351, EPI_ISL_1246352, EPI_ISL_1246353, EPI_ISL_1246354, EPI_ISL_1246355, EPI_ISL_1246356, EPI_ISL_1246357, EPI_ISL_1246358, EPI_ISL_1246359, EPI_ISL_1246361, EPI_ISL_1246362, EPI_ISL_1246363, EPI_ISL_1246364, EPI_ISL_1246365, EPI_ISL_1246366, EPI_ISL_1246367, EPI_ISL_1246368, EPI_ISL_1246369, EPI_ISL_1246370, EPI_ISL_1246371, EPI_ISL_1246372, EPI_ISL_1246373, EPI_ISL_1246374, EPI_ISL_1246375, EPI_ISL_1246376, EPI_ISL_1246377, EPI_ISL_1246378, EPI_ISL_1246379, EPI_ISL_1246380, EPI_ISL_1246381, EPI_ISL_1246382, EPI_ISL_1246384, EPI_ISL_1246385, EPI_ISL_1246386, EPI_ISL_1246387, EPI_ISL_1246388, EPI_ISL_1246389, EPI_ISL_1246391, EPI_ISL_1246392, EPI_ISL_1246393, EPI_ISL_1246394, EPI_ISL_1246395, EPI_ISL_1246396, EPI_ISL_1246397, EPI_ISL_1246398, EPI_ISL_1246399, EPI_ISL_1246400, EPI_ISL_1246401, EPI_ISL_1246402, EPI_ISL_1246403, EPI_ISL_1246404, EPI_ISL_1246406, EPI_ISL_1246407, EPI_ISL_1246408, EPI_ISL_1246409, EPI_ISL_1246410, EPI_ISL_1246411, EPI_ISL_1246413, EPI_ISL_1246414, EPI_ISL_1246415, EPI_ISL_1246416, EPI_ISL_1246418, EPI_ISL_1246420, EPI_ISL_1246422, EPI_ISL_1246423, EPI_ISL_1246424, EPI_ISL_1246425, EPI_ISL_1246426, EPI_ISL_1246427, EPI_ISL_1246428, EPI_ISL_1246429, EPI_ISL_1246430, EPI_ISL_1246431, EPI_ISL_1246432, EPI_ISL_1246433, EPI_ISL_1246434, EPI_ISL_1246435, EPI_ISL_1246436, EPI_ISL_1246437, EPI_ISL_1246438, EPI_ISL_1246439, EPI_ISL_1246440, EPI_ISL_1246441, EPI_ISL_1246442, EPI_ISL_1246443, EPI_ISL_1246444, EPI_ISL_1246445, EPI_ISL_1246446, EPI_ISL_1246447, EPI_ISL_1246449, EPI_ISL_1246451, EPI_ISL_1246452, EPI_ISL_1246453, EPI_ISL_1246455, EPI_ISL_1246456, EPI_ISL_1246457, EPI_ISL_1246458, EPI_ISL_1246459, EPI_ISL_1246460, EPI_ISL_1246461, EPI_ISL_1246462, EPI_ISL_1246463, EPI_ISL_1246464, EPI_ISL_1246465, EPI_ISL_1246467, EPI_ISL_1246468, EPI_ISL_1246469, EPI_ISL_1246471, EPI_ISL_1246472, EPI_ISL_1246473, EPI_ISL_1246474, EPI_ISL_1246475, EPI_ISL_1246476, EPI_ISL_1246477, EPI_ISL_1246478, EPI_ISL_1246479, EPI_ISL_1246480, EPI_ISL_1246481, EPI_ISL_1246482, EPI_ISL_1246483, EPI_ISL_1246484, EPI_ISL_1246485, EPI_ISL_1246487, EPI_ISL_1246488, EPI_ISL_1246489, EPI_ISL_1246490, EPI_ISL_1246491, EPI_ISL_1246492, EPI_ISL_1246493, EPI_ISL_1246494, EPI_ISL_1246495, EPI_ISL_1246496, EPI_ISL_1246497, EPI_ISL_1246498, EPI_ISL_1246499, EPI_ISL_1246500, EPI_ISL_1246501, EPI_ISL_1246502, EPI_ISL_1246505, EPI_ISL_1246506, EPI_ISL_1246507, EPI_ISL_1246508, EPI_ISL_1246509, EPI_ISL_1246510, EPI_ISL_1246512, EPI_ISL_1246513, EPI_ISL_1246515, EPI_ISL_1246516, EPI_ISL_1246517, EPI_ISL_1246518, EPI_ISL_1246519, EPI_ISL_1246520, EPI_ISL_1246521, EPI_ISL_1246522, EPI_ISL_1246523, EPI_ISL_1246524, EPI_ISL_1246525, EPI_ISL_1246527, EPI_ISL_1246528, EPI_ISL_1246529, EPI_ISL_1246530, EPI_ISL_1246531, EPI_ISL_1246532, EPI_ISL_1246534, EPI_ISL_1246535, EPI_ISL_1246536, EPI_ISL_1246537, EPI_ISL_1246538, EPI_ISL_1246539, EPI_ISL_1246540, EPI_ISL_1246541, EPI_ISL_1246542, EPI_ISL_1246543, EPI_ISL_1246544, EPI_ISL_1246545, EPI_ISL_1246546, EPI_ISL_1246547, EPI_ISL_1246548, EPI_ISL_1246549, EPI_ISL_1246550, EPI_ISL_1246551, EPI_ISL_1246552, EPI_ISL_1246553, EPI_ISL_1246554, EPI_ISL_1246556, EPI_ISL_1246557, EPI_ISL_1246558, EPI_ISL_1246559, EPI_ISL_1246560, EPI_ISL_1246561, EPI_ISL_1246562, EPI_ISL_1246563, EPI_ISL_1246564, EPI_ISL_1246565, EPI_ISL_1246566, EPI_ISL_1246567, EPI_ISL_1246568, EPI_ISL_1246569, EPI_ISL_1246570, EPI_ISL_1246571, EPI_ISL_1246572, EPI_ISL_1246573, EPI_ISL_1246574, EPI_ISL_1246575, EPI_ISL_1246576, EPI_ISL_1246577, EPI_ISL_1246578, EPI_ISL_1246579, EPI_ISL_1246580, EPI_ISL_1246581, EPI_ISL_1246582, EPI_ISL_1246583, EPI_ISL_1246584, EPI_ISL_1246585, EPI_ISL_1246586, EPI_ISL_1246587, EPI_ISL_1246588, EPI_ISL_1246589, EPI_ISL_1246590, EPI_ISL_1246591, EPI_ISL_1246592, EPI_ISL_1246593, EPI_ISL_1246594, EPI_ISL_1246595, EPI_ISL_1246596, EPI_ISL_1246597, EPI_ISL_1246598, EPI_ISL_1246599, EPI_ISL_1246600, EPI_ISL_1246601, EPI_ISL_1246602, EPI_ISL_1246603, EPI_ISL_1246604, EPI_ISL_1246605, EPI_ISL_1246606, EPI_ISL_1246608, EPI_ISL_1246609, EPI_ISL_1246610, EPI_ISL_1246611, EPI_ISL_1246612, EPI_ISL_1246613, EPI_ISL_1246614, EPI_ISL_1246615, EPI_ISL_1246616, EPI_ISL_1246617, EPI_ISL_1246618, EPI_ISL_1246619, EPI_ISL_1246620, EPI_ISL_1246621, EPI_ISL_1246622, EPI_ISL_1246623, EPI_ISL_1246624, EPI_ISL_1246625, EPI_ISL_1246626, EPI_ISL_1246627, EPI_ISL_1246628, EPI_ISL_1246629, EPI_ISL_1246630, EPI_ISL_1246631, EPI_ISL_1246632, EPI_ISL_1246633, EPI_ISL_1246634, EPI_ISL_1246636, EPI_ISL_1246637, EPI_ISL_1246638, EPI_ISL_1246639, EPI_ISL_1246640, EPI_ISL_1246641, EPI_ISL_1246642, EPI_ISL_1246643, EPI_ISL_1246644, EPI_ISL_1246645, EPI_ISL_1246646, EPI_ISL_1246647, EPI_ISL_1246648, EPI_ISL_1246649, EPI_ISL_1246650, EPI_ISL_1246651, EPI_ISL_1246652, EPI_ISL_1246653, EPI_ISL_1246654, EPI_ISL_1246655, EPI_ISL_1246656, EPI_ISL_1246657, EPI_ISL_1246658, EPI_ISL_1246659, EPI_ISL_1246660, EPI_ISL_1246661, EPI_ISL_1246662, EPI_ISL_1246663, EPI_ISL_1246664, EPI_ISL_1246665, EPI_ISL_1246666, EPI_ISL_1246667, EPI_ISL_1246668, EPI_ISL_1246669, EPI_ISL_1246670, EPI_ISL_1246671, EPI_ISL_1246672, EPI_ISL_1246673, EPI_ISL_1246674, EPI_ISL_1246675, EPI_ISL_1246676, EPI_ISL_1246677, EPI_ISL_1246678, EPI_ISL_1246679, EPI_ISL_1246680, EPI_ISL_1246681, EPI_ISL_1246682, EPI_ISL_1246683, EPI_ISL_1246684, EPI_ISL_1246685, EPI_ISL_1246686, EPI_ISL_1246687, EPI_ISL_1246688, EPI_ISL_1246689, EPI_ISL_1246690, EPI_ISL_1246691, EPI_ISL_1246693, EPI_ISL_1246694, EPI_ISL_1246695, EPI_ISL_1246696, EPI_ISL_1246698, EPI_ISL_1246699, EPI_ISL_1246700, EPI_ISL_1246701, EPI_ISL_1246702, EPI_ISL_1246703, EPI_ISL_1246704, EPI_ISL_1246705, EPI_ISL_1246706, EPI_ISL_1246707, EPI_ISL_1246708, EPI_ISL_1246709, EPI_ISL_1246710, EPI_ISL_1246711, EPI_ISL_1246712, EPI_ISL_1246713, EPI_ISL_1246714, EPI_ISL_1246715, EPI_ISL_1246716, EPI_ISL_1246717, EPI_ISL_1246719, EPI_ISL_1246720, EPI_ISL_1246721, EPI_ISL_1246723, EPI_ISL_1246724, EPI_ISL_1246725, EPI_ISL_1246726, EPI_ISL_1246727, EPI_ISL_1246728, EPI_ISL_1246729, EPI_ISL_1246730, EPI_ISL_1246731, EPI_ISL_1246732, EPI_ISL_1246733, EPI_ISL_1246734, EPI_ISL_1246735, EPI_ISL_1246736, EPI_ISL_1246737, EPI_ISL_1246738, EPI_ISL_1246739, EPI_ISL_1246740, EPI_ISL_1246741, EPI_ISL_1246742, EPI_ISL_1246743, EPI_ISL_1246744, EPI_ISL_1246745, EPI_ISL_1246746, EPI_ISL_1246748, EPI_ISL_1246749, EPI_ISL_1246750, EPI_ISL_1246751, EPI_ISL_1246752, EPI_ISL_1246753, EPI_ISL_1246754, EPI_ISL_1246755, EPI_ISL_1246756, EPI_ISL_1246757, EPI_ISL_1246758, EPI_ISL_1246759, EPI_ISL_1246760, EPI_ISL_1246761, EPI_ISL_1246762, EPI_ISL_1246763, EPI_ISL_1246765, EPI_ISL_1246766, EPI_ISL_1246767, EPI_ISL_1246768, EPI_ISL_1246769, EPI_ISL_1246770, EPI_ISL_1246771, EPI_ISL_1246772, EPI_ISL_1246773, EPI_ISL_1246774, EPI_ISL_1246776, EPI_ISL_1246777, EPI_ISL_1246778, EPI_ISL_1246779, EPI_ISL_1246780, EPI_ISL_1246781, EPI_ISL_1246782, EPI_ISL_1246783, EPI_ISL_1246784, EPI_ISL_1246785, EPI_ISL_1246786, EPI_ISL_1246787, EPI_ISL_1246788, EPI_ISL_1246789, EPI_ISL_1246790, EPI_ISL_1246791, EPI_ISL_1246792, EPI_ISL_1246793, EPI_ISL_1246794, EPI_ISL_1246795, EPI_ISL_1246796, EPI_ISL_1246797, EPI_ISL_1246798, EPI_ISL_1246799, EPI_ISL_1246800, EPI_ISL_1246801, EPI_ISL_1246802, EPI_ISL_1246803, EPI_ISL_1246804, EPI_ISL_1246805, EPI_ISL_1246806, EPI_ISL_1246807, EPI_ISL_1246808, EPI_ISL_1246809, EPI_ISL_1246810, EPI_ISL_1246811, EPI_ISL_1246812, EPI_ISL_1246813, EPI_ISL_1246814, EPI_ISL_1246815, EPI_ISL_1246816, EPI_ISL_1246817, EPI_ISL_1246818, EPI_ISL_1246819, EPI_ISL_1246820, EPI_ISL_1246821, EPI_ISL_1246822, EPI_ISL_1246823, EPI_ISL_1246824, EPI_ISL_1246825, EPI_ISL_1246826, EPI_ISL_1246827, EPI_ISL_1246828, EPI_ISL_1246829, EPI_ISL_1246830, EPI_ISL_1246831, EPI_ISL_1246832, EPI_ISL_1246833, EPI_ISL_1246834, EPI_ISL_1246835, EPI_ISL_1246836, EPI_ISL_1246837, EPI_ISL_1246838, EPI_ISL_1246840, EPI_ISL_1246841, EPI_ISL_1246842, EPI_ISL_1246843, EPI_ISL_1246844, EPI_ISL_1246845, EPI_ISL_1246846, EPI_ISL_1246847, EPI_ISL_1246848, EPI_ISL_1246849, EPI_ISL_1246850, EPI_ISL_1246851, EPI_ISL_1246852, EPI_ISL_1246853, EPI_ISL_1246854, EPI_ISL_1246855, EPI_ISL_1246875, EPI_ISL_1246876, EPI_ISL_1246877, EPI_ISL_1246878, EPI_ISL_1246879, EPI_ISL_1246880, EPI_ISL_1246881, EPI_ISL_1246882, EPI_ISL_1246884, EPI_ISL_1246885, EPI_ISL_1246886, EPI_ISL_1246887, EPI_ISL_1246889, EPI_ISL_1246890, EPI_ISL_1246891, EPI_ISL_1246892, EPI_ISL_1246893, EPI_ISL_1246894, EPI_ISL_1246895, EPI_ISL_1246896, EPI_ISL_1246898, EPI_ISL_1246899, EPI_ISL_1246901, EPI_ISL_1246902, EPI_ISL_1246903, EPI_ISL_1246904, EPI_ISL_1246905, EPI_ISL_1246906, EPI_ISL_1246907, EPI_ISL_1246908, EPI_ISL_1246909, EPI_ISL_1246910, EPI_ISL_1246911, EPI_ISL_1246912, EPI_ISL_1246913, EPI_ISL_1246914, EPI_ISL_1246915, EPI_ISL_1246916, EPI_ISL_1246918, EPI_ISL_1246919, EPI_ISL_1246920, EPI_ISL_1246921, EPI_ISL_1246922, EPI_ISL_1246923, EPI_ISL_1246924, EPI_ISL_1246925, EPI_ISL_1246926, EPI_ISL_1246927, EPI_ISL_1246928, EPI_ISL_1246929, EPI_ISL_1246931, EPI_ISL_1246932, EPI_ISL_1246933, EPI_ISL_1246934, EPI_ISL_1246935, EPI_ISL_1246936, EPI_ISL_1246937, EPI_ISL_1246939, EPI_ISL_1246940, EPI_ISL_1246941, EPI_ISL_1246942, EPI_ISL_1246943, EPI_ISL_1246944, EPI_ISL_1246945, EPI_ISL_1246946, EPI_ISL_1246947, EPI_ISL_1246949, EPI_ISL_1246950, EPI_ISL_1246951, EPI_ISL_1246952 | | | |
| see above | Lighthouse Lab in Glasgow | Wellcome Sanger Institute for the COVID-19 Genomics UK (COG-UK) Consortium | Harper VanSteenhouse, Yumi Kasai, David Gray, Carol Clugston, Anna Dominiczak and Alex Alderton, Roberto Amato, Jeffrey Barrett, Sonia Goncalves, Ewan Harrison, David K. Jackson, Ian Johnston, Dominic Kwiatkowski, Cordelia Langford, John Sillitoe on behalf of the Wellcome Sanger Institute COVID-19 Surveillance Team |
| EPI_ISL_1247640, EPI_ISL_1247685 | West of Scotland Specialist Virology Centre, NHSGGC / MRC-University of Glasgow Centre for Virus Research | COVID-19 Genomics UK (COG-UK) Consortium | Ana da Silva Filipe, Natasha Johnson, Kathy Smollett, Daniel Mair, Stephen Carmichael, Alice Broos, Lily Tong, Jenna Nichols, Kyriaki Nomikou; Sarah McDonald; Richard Orton, Joseph Hughes, Sreenu Vattipally, David L Robertson; Alasdair MacLean, Rory Gunson; Sharif Shaaban, Matthew Holden; Rachel Blacow, Guy Mollett, Kathy Li, James Shepherd, Antonia Ho, Emma Thomson |
| EPI_ISL_1250124, EPI_ISL_1250126, EPI_ISL_1250132, EPI_ISL_1250133, EPI_ISL_1250137, EPI_ISL_1250141, EPI_ISL_1250144, EPI_ISL_1250148, EPI_ISL_1250152, EPI_ISL_1250155, EPI_ISL_1250164, EPI_ISL_1250167, EPI_ISL_1250171, EPI_ISL_1250172, EPI_ISL_1250181, EPI_ISL_1250185, EPI_ISL_1250187, EPI_ISL_1250191, EPI_ISL_1250192, EPI_ISL_1250199, EPI_ISL_1250203, EPI_ISL_1250206, EPI_ISL_1250207, EPI_ISL_1250210, EPI_ISL_1250216, EPI_ISL_1250217, EPI_ISL_1250218, EPI_ISL_1250219, EPI_ISL_1250221 | | | |
| see above | Department of Pathology, University of Cambridge | COVID-19 Genomics UK (COG-UK) Consortium | Aminu S. Jahun, Yasmin Chaudhry, Iliana Georgana, Myra Hosmillo, Rhys Izuagbe, William L. Hamilton, Martin D. Curran, Surendra Parmar, Ian |

We gratefully acknowledge the following Authors from the Originating laboratories responsible for obtaining the specimens, as well as the Submitting laboratories where the genome data were generated and shared via GISAID, on which this research is based.

All Submitters of data may be contacted directly via www.gisaid.org

Authors are sorted alphabetically.

| Accession ID | Originating Laboratory | Submitting Laboratory | Authors |
|---|---|---|---|
| EPI_ISL_1506134, EPI_ISL_1506135, EPI_ISL_1506136, EPI_ISL_1506137, EPI_ISL_1506138, EPI_ISL_1506140, EPI_ISL_1506141, EPI_ISL_1506142, EPI_ISL_1506143, EPI_ISL_1506146, EPI_ISL_1506147, EPI_ISL_1506148, EPI_ISL_1506149, EPI_ISL_1506150, EPI_ISL_1506151, EPI_ISL_1506152, EPI_ISL_1506153, EPI_ISL_1506154, EPI_ISL_1506155, EPI_ISL_1506156, EPI_ISL_1506160, EPI_ISL_1506161, EPI_ISL_1506162, EPI_ISL_1506164, EPI_ISL_1506166, EPI_ISL_1506167, EPI_ISL_1506168, EPI_ISL_1506169, EPI_ISL_1506170 | | | |
| see above | Lighthouse Lab in Glasgow | Wellcome Sanger Institute for the COVID-19 Genomics UK (COG-UK) Consortium | Harper VanSteenhouse, Yumi Kasai, David Gray, Carol Clugston, Anna Dominiczak and Alex Alderton, Roberto Amato, Jeffrey Barrett, Sonia Goncalves, Ewan Harrison, David K. Jackson, Ian Johnston, Dominic Kwiatkowski, Cordelia Langford, John Sillitoe on behalf of the Wellcome Sanger Institute COVID-19 Surveillance Team |
| EPI_ISL_1506172 | Lighthouse Lab in Cambridge | Wellcome Sanger Institute for the COVID-19 Genomics UK (COG-UK) Consortium | Rob Howes, The Lighthouse Lab in Cambridge and Alex Alderton, Roberto Amato, Jeffrey Barrett, Sonia Goncalves, Ewan Harrison, David K. Jackson, Ian Johnston, Dominic Kwiatkowski, Cordelia Langford, John Sillitoe on behalf of the Wellcome Sanger Institute COVID-19 Surveillance Team |
| EPI_ISL_1506173, EPI_ISL_1506174, EPI_ISL_1506176, EPI_ISL_1506177, EPI_ISL_1506178, EPI_ISL_1506179, EPI_ISL_1506180, EPI_ISL_1506181, EPI_ISL_1506182, EPI_ISL_1506183, EPI_ISL_1506184, EPI_ISL_1506185, EPI_ISL_1506186, EPI_ISL_1506187, EPI_ISL_1506188, EPI_ISL_1506189, EPI_ISL_1506190, EPI_ISL_1506191, EPI_ISL_1506193, EPI_ISL_1506194, EPI_ISL_1506195, EPI_ISL_1506196, EPI_ISL_1506197, EPI_ISL_1506198, EPI_ISL_1506199, EPI_ISL_1506200, EPI_ISL_1506201, EPI_ISL_1506202, EPI_ISL_1506203, EPI_ISL_1506204, EPI_ISL_1506205, EPI_ISL_1506206, EPI_ISL_1506209, EPI_ISL_1506210, EPI_ISL_1506211, EPI_ISL_1506212, EPI_ISL_1506213, EPI_ISL_1506215, EPI_ISL_1506217, EPI_ISL_1506218, EPI_ISL_1506219, EPI_ISL_1506220, EPI_ISL_1506221, EPI_ISL_1506223, EPI_ISL_1506224, EPI_ISL_1506225, EPI_ISL_1506226, EPI_ISL_1506227, EPI_ISL_1506228, EPI_ISL_1506229, EPI_ISL_1506231, EPI_ISL_1506233, EPI_ISL_1506234, EPI_ISL_1506235, EPI_ISL_1506236, EPI_ISL_1506237, EPI_ISL_1506238, EPI_ISL_1506239, EPI_ISL_1506240, EPI_ISL_1506241, EPI_ISL_1506242, EPI_ISL_1506243, EPI_ISL_1506244, EPI_ISL_1506245, EPI_ISL_1506246, EPI_ISL_1506247, EPI_ISL_1506248, EPI_ISL_1506249, EPI_ISL_1506250, EPI_ISL_1506252, EPI_ISL_1506254, EPI_ISL_1506255, EPI_ISL_1506256, EPI_ISL_1506257, EPI_ISL_1506258, EPI_ISL_1506259, EPI_ISL_1506260, EPI_ISL_1506261, EPI_ISL_1506262, EPI_ISL_1506263, EPI_ISL_1506264, EPI_ISL_1506265, EPI_ISL_1506267, EPI_ISL_1506268, EPI_ISL_1506269, EPI_ISL_1506270, EPI_ISL_1506271, EPI_ISL_1506272, EPI_ISL_1506273, EPI_ISL_1506274, EPI_ISL_1506275, EPI_ISL_1506276, EPI_ISL_1506279, EPI_ISL_1506280, EPI_ISL_1506281, EPI_ISL_1506282, EPI_ISL_1506283, EPI_ISL_1506284, EPI_ISL_1506285, EPI_ISL_1506286, EPI_ISL_1506287, EPI_ISL_1506288, EPI_ISL_1506289, EPI_ISL_1506290, EPI_ISL_1506291, EPI_ISL_1506293, EPI_ISL_1506294, EPI_ISL_1506295, EPI_ISL_1506296, EPI_ISL_1506297, EPI_ISL_1506298, EPI_ISL_1506299, EPI_ISL_1506300, EPI_ISL_1506301, EPI_ISL_1506302, EPI_ISL_1506303, EPI_ISL_1506304, EPI_ISL_1506306, EPI_ISL_1506307, EPI_ISL_1506308, EPI_ISL_1506309, EPI_ISL_1506310, EPI_ISL_1506311, EPI_ISL_1506312, EPI_ISL_1506313, EPI_ISL_1506314, EPI_ISL_1506315, EPI_ISL_1506316, EPI_ISL_1506317, EPI_ISL_1506318, EPI_ISL_1506319, EPI_ISL_1506320, EPI_ISL_1506322, EPI_ISL_1506323, EPI_ISL_1506324, EPI_ISL_1506325, EPI_ISL_1506326, EPI_ISL_1506327, EPI_ISL_1506328, EPI_ISL_1506329, EPI_ISL_1506330, EPI_ISL_1506331, EPI_ISL_1506332, EPI_ISL_1506333, EPI_ISL_1506334, EPI_ISL_1506336, EPI_ISL_1506337, EPI_ISL_1506338, EPI_ISL_1506339, EPI_ISL_1506340, EPI_ISL_1506341, EPI_ISL_1506342, EPI_ISL_1506343, EPI_ISL_1506344, EPI_ISL_1506345, EPI_ISL_1506346, EPI_ISL_1506347, EPI_ISL_1506348, EPI_ISL_1506349, EPI_ISL_1506350, EPI_ISL_1506351, EPI_ISL_1506352, EPI_ISL_1506354, EPI_ISL_1506355, EPI_ISL_1506356, EPI_ISL_1506357, EPI_ISL_1506358, EPI_ISL_1506359, EPI_ISL_1506360, EPI_ISL_1506361, EPI_ISL_1506362, EPI_ISL_1506363, EPI_ISL_1506364, EPI_ISL_1506365, EPI_ISL_1506366, EPI_ISL_1506367, EPI_ISL_1506368, EPI_ISL_1506369, EPI_ISL_1506370, EPI_ISL_1506371, EPI_ISL_1506372, EPI_ISL_1506373, EPI_ISL_1506374, EPI_ISL_1506375, EPI_ISL_1506376, EPI_ISL_1506377, EPI_ISL_1506378, EPI_ISL_1506379, EPI_ISL_1506380, EPI_ISL_1506382, EPI_ISL_1506383, EPI_ISL_1506384, EPI_ISL_1506385, EPI_ISL_1506386, EPI_ISL_1506387, EPI_ISL_1506388, EPI_ISL_1506389, EPI_ISL_1506391, EPI_ISL_1506392, EPI_ISL_1506393, EPI_ISL_1506394, EPI_ISL_1506395, EPI_ISL_1506396, EPI_ISL_1506398, EPI_ISL_1506399, EPI_ISL_1506400, EPI_ISL_1506401, EPI_ISL_1506402, EPI_ISL_1506403, EPI_ISL_1506406, EPI_ISL_1506408, EPI_ISL_1506409, EPI_ISL_1506410, EPI_ISL_1506411, EPI_ISL_1506413, EPI_ISL_1506414, EPI_ISL_1506415, EPI_ISL_1506416, EPI_ISL_1506417, EPI_ISL_1506418, EPI_ISL_1506419, EPI_ISL_1506420, EPI_ISL_1506421, EPI_ISL_1506422, EPI_ISL_1506423, EPI_ISL_1506428, EPI_ISL_1506429, EPI_ISL_1506430, EPI_ISL_1506432, EPI_ISL_1506433, EPI_ISL_1506435, EPI_ISL_1506436, EPI_ISL_1506437, EPI_ISL_1506438, EPI_ISL_1506439, EPI_ISL_1506441, EPI_ISL_1506442, EPI_ISL_1506443, EPI_ISL_1506444, EPI_ISL_1506446, EPI_ISL_1506447, EPI_ISL_1506448, EPI_ISL_1506449, EPI_ISL_1506450, EPI_ISL_1506451, EPI_ISL_1506454, EPI_ISL_1506455, EPI_ISL_1506457, EPI_ISL_1506458, EPI_ISL_1506459, EPI_ISL_1506460, EPI_ISL_1506462, EPI_ISL_1506463, EPI_ISL_1506464, EPI_ISL_1506465, EPI_ISL_1506466, EPI_ISL_1506467, EPI_ISL_1506468, EPI_ISL_1506469, EPI_ISL_1506470, EPI_ISL_1506471, EPI_ISL_1506472, EPI_ISL_1506473, EPI_ISL_1506474, EPI_ISL_1506475, EPI_ISL_1506476, EPI_ISL_1506477, EPI_ISL_1506478, EPI_ISL_1506479, EPI_ISL_1506481, EPI_ISL_1506482, EPI_ISL_1506483, EPI_ISL_1506484, EPI_ISL_1506485, EPI_ISL_1506486, EPI_ISL_1506487, EPI_ISL_1506488, EPI_ISL_1506489, EPI_ISL_1506490, EPI_ISL_1506491, EPI_ISL_1506492, EPI_ISL_1506493, EPI_ISL_1506494, EPI_ISL_1506495, EPI_ISL_1506496, EPI_ISL_1506497, EPI_ISL_1506498, EPI_ISL_1506499, EPI_ISL_1506500, EPI_ISL_1506501, EPI_ISL_1506502, EPI_ISL_1506503, EPI_ISL_1506504, EPI_ISL_1506505, EPI_ISL_1506506, EPI_ISL_1506507, EPI_ISL_1506508, EPI_ISL_1506510, EPI_ISL_1506511, EPI_ISL_1506537, EPI_ISL_1506538, EPI_ISL_1506539, EPI_ISL_1506540, EPI_ISL_1506542, EPI_ISL_1506543, EPI_ISL_1506544, EPI_ISL_1506545, EPI_ISL_1506546, EPI_ISL_1506547, EPI_ISL_1506548, EPI_ISL_1506550, EPI_ISL_1506551, EPI_ISL_1506552, EPI_ISL_1506553, EPI_ISL_1506554, EPI_ISL_1506555, EPI_ISL_1506556, EPI_ISL_1506557, EPI_ISL_1506558, EPI_ISL_1506559, EPI_ISL_1506560, EPI_ISL_1506561, EPI_ISL_1506563, EPI_ISL_1506566, EPI_ISL_1506568, EPI_ISL_1506569, EPI_ISL_1506570, EPI_ISL_1506608, EPI_ISL_1506635, EPI_ISL_1506649, EPI_ISL_1506656, EPI_ISL_1506666, EPI_ISL_1506670, EPI_ISL_1506672, EPI_ISL_1506673, EPI_ISL_1506676, EPI_ISL_1506713, EPI_ISL_1506740, EPI_ISL_1506751, EPI_ISL_1507078, EPI_ISL_1507086, EPI_ISL_1507087, EPI_ISL_1507095, EPI_ISL_1507147, EPI_ISL_1507150, EPI_ISL_1507155, EPI_ISL_1507156, EPI_ISL_1507161, EPI_ISL_1507193, EPI_ISL_1507195, EPI_ISL_1507255, EPI_ISL_1507526, EPI_ISL_1537142, EPI_ISL_1537143, EPI_ISL_1537144, EPI_ISL_1537145, EPI_ISL_1537146, EPI_ISL_1537147, EPI_ISL_1537148, EPI_ISL_1537149, EPI_ISL_1537150, EPI_ISL_1537151, EPI_ISL_1537153, EPI_ISL_1537154, EPI_ISL_1537155, EPI_ISL_1537156, EPI_ISL_1537157, EPI_ISL_1537158, EPI_ISL_1537159, EPI_ISL_1537161, EPI_ISL_1537162, EPI_ISL_1537163, EPI_ISL_1537164, EPI_ISL_1537165, EPI_ISL_1537166, EPI_ISL_1537167, EPI_ISL_1537168, EPI_ISL_1537169, EPI_ISL_1537170, EPI_ISL_1537171, EPI_ISL_1537172, EPI_ISL_1537173, EPI_ISL_1537174, EPI_ISL_1537175, EPI_ISL_1537176, EPI_ISL_1537177, EPI_ISL_1537178, EPI_ISL_1537179, EPI_ISL_1537180, EPI_ISL_1537181, EPI_ISL_1537182, EPI_ISL_1537183, EPI_ISL_1537184, EPI_ISL_1537185, EPI_ISL_1537186, EPI_ISL_1537187, EPI_ISL_1537188, EPI_ISL_1537189, EPI_ISL_1537190, EPI_ISL_1537191, EPI_ISL_1537192, EPI_ISL_1537195, EPI_ISL_1537196, EPI_ISL_1537197, EPI_ISL_1537198, EPI_ISL_1537199, EPI_ISL_1537200, EPI_ISL_1537201, EPI_ISL_1537202, EPI_ISL_1537203, EPI_ISL_1537204, EPI_ISL_1537205, EPI_ISL_1537206, EPI_ISL_1537207, EPI_ISL_1537208, EPI_ISL_1537210, EPI_ISL_1537211, EPI_ISL_1537212, EPI_ISL_1537213, EPI_ISL_1537214, EPI_ISL_1537215, EPI_ISL_1537216, EPI_ISL_1537217, EPI_ISL_1537218, EPI_ISL_1537219, EPI_ISL_1537220, EPI_ISL_1537221, EPI_ISL_1537222, EPI_ISL_1537223, EPI_ISL_1537224, EPI_ISL_1537225, EPI_ISL_1537227, EPI_ISL_1537228, EPI_ISL_1537229, EPI_ISL_1537230, EPI_ISL_1537231, EPI_ISL_1537232, EPI_ISL_1537233, EPI_ISL_1537234, EPI_ISL_1537235, EPI_ISL_1537237, EPI_ISL_1537238, EPI_ISL_1537239, EPI_ISL_1537241, EPI_ISL_1537242, EPI_ISL_1537243, EPI_ISL_1537244, EPI_ISL_1537245, EPI_ISL_1537246, EPI_ISL_1537247, EPI_ISL_1537249, EPI_ISL_1537250, EPI_ISL_1537251, EPI_ISL_1537252, EPI_ISL_1537253, EPI_ISL_1537254, EPI_ISL_1537255, EPI_ISL_1537256, EPI_ISL_1537257, EPI_ISL_1537258, EPI_ISL_1537259, EPI_ISL_1537260, EPI_ISL_1537261, EPI_ISL_1537262, EPI_ISL_1537263, EPI_ISL_1537264, EPI_ISL_1537265, EPI_ISL_1537266, EPI_ISL_1537267, EPI_ISL_1537268, EPI_ISL_1537269, EPI_ISL_1537270, EPI_ISL_1537271, EPI_ISL_1537272, EPI_ISL_1537273, EPI_ISL_1537274, EPI_ISL_1537275, EPI_ISL_1537276, EPI_ISL_1537277, EPI_ISL_1537278, EPI_ISL_1537279, EPI_ISL_1537280, EPI_ISL_1537281, EPI_ISL_1537282, EPI_ISL_1537284, EPI_ISL_1537285, EPI_ISL_1537286, EPI_ISL_1537287, EPI_ISL_1537288, EPI_ISL_1537289, EPI_ISL_1537290, EPI_ISL_1537291, EPI_ISL_1537292, EPI_ISL_1537293, EPI_ISL_1537294, EPI_ISL_1537295, EPI_ISL_1537296, EPI_ISL_1537297, EPI_ISL_1537298, EPI_ISL_1537299, EPI_ISL_1537300, EPI_ISL_1537301, EPI_ISL_1537302, EPI_ISL_1537303, EPI_ISL_1537304, EPI_ISL_1537305, EPI_ISL_1537306, EPI_ISL_1537307, EPI_ISL_1537308, EPI_ISL_1537309, EPI_ISL_1537310, EPI_ISL_1537311, EPI_ISL_1537312, EPI_ISL_1537313, EPI_ISL_1537314, EPI_ISL_1537845, EPI_ISL_1537846, EPI_ISL_1537847, EPI_ISL_1537848, EPI_ISL_1537849, EPI_ISL_1537850, EPI_ISL_1537851, EPI_ISL_1537852, EPI_ISL_1537853, EPI_ISL_1537854, EPI_ISL_1537855, EPI_ISL_1537856, EPI_ISL_1537857, EPI_ISL_1537858, EPI_ISL_1537859, EPI_ISL_1537860, EPI_ISL_1537861, EPI_ISL_1537862, EPI_ISL_1537863, EPI_ISL_1537864, EPI_ISL_1537865, EPI_ISL_1537866, EPI_ISL_1537867, EPI_ISL_1537868, EPI_ISL_1537869, EPI_ISL_1537870, EPI_ISL_1537871, EPI_ISL_1537872, EPI_ISL_1537873, EPI_ISL_1537874, EPI_ISL_1537875, EPI_ISL_1537876, EPI_ISL_1537877, EPI_ISL_1537878, EPI_ISL_1537879, EPI_ISL_1537880, EPI_ISL_1537881, EPI_ISL_1537882, EPI_ISL_1537884, EPI_ISL_1537885, EPI_ISL_1537886, EPI_ISL_1537887, EPI_ISL_1537888, EPI_ISL_1537889, EPI_ISL_1537890, EPI_ISL_1537891, EPI_ISL_1537893, EPI_ISL_1537894, EPI_ISL_1537895, EPI_ISL_1537896, EPI_ISL_1537897, EPI_ISL_1537898, EPI_ISL_1537899, EPI_ISL_1537900, EPI_ISL_1537901, EPI_ISL_1537902, EPI_ISL_1537903, EPI_ISL_1537904, EPI_ISL_1537905, EPI_ISL_1537906, EPI_ISL_1537907, EPI_ISL_1537908, EPI_ISL_1537909, EPI_ISL_1537910, EPI_ISL_1537911, EPI_ISL_1537912, EPI_ISL_1537913, EPI_ISL_1537914, EPI_ISL_1537915, EPI_ISL_1537916, EPI_ISL_1537917, EPI_ISL_1537918, EPI_ISL_1537919, EPI_ISL_1537920, EPI_ISL_1537921, EPI_ISL_1537922, EPI_ISL_1537923, EPI_ISL_1537924, EPI_ISL_1537925, EPI_ISL_1537926, EPI_ISL_1537927, EPI_ISL_1537928, EPI_ISL_1537929, EPI_ISL_1537930, EPI_ISL_1537931, EPI_ISL_1537932, EPI_ISL_1537933, EPI_ISL_1537934, EPI_ISL_1537935, EPI_ISL_1537937, EPI_ISL_1537938, EPI_ISL_1537939, EPI_ISL_1537940, EPI_ISL_1537941, EPI_ISL_1537942, EPI_ISL_1537943, EPI_ISL_1537944, EPI_ISL_1537945, EPI_ISL_1537946, EPI_ISL_1537947, EPI_ISL_1537948, EPI_ISL_1537949, EPI_ISL_1537950, EPI_ISL_1537951, EPI_ISL_1537952, EPI_ISL_1537953, EPI_ISL_1537954, EPI_ISL_1537955, EPI_ISL_1537958, EPI_ISL_1537959, EPI_ISL_1537960, EPI_ISL_1537961, EPI_ISL_1537962, EPI_ISL_1537963, EPI_ISL_1537964, EPI_ISL_1537965, EPI_ISL_1537966, EPI_ISL_1537967, EPI_ISL_1537968, EPI_ISL_1537969, EPI_ISL_1537970, EPI_ISL_1537971, EPI_ISL_1537972, EPI_ISL_1537973, EPI_ISL_1537974, EPI_ISL_1537976, EPI_ISL_1537977, EPI_ISL_1537978, EPI_ISL_1537979, EPI_ISL_1537980, EPI_ISL_1537981, EPI_ISL_1537982, EPI_ISL_1537983, EPI_ISL_1537984, EPI_ISL_1537985, EPI_ISL_1537986, EPI_ISL_1537988, EPI_ISL_1537989, EPI_ISL_1537990, EPI_ISL_1537991, EPI_ISL_1537992, EPI_ISL_1537993, EPI_ISL_1537994, EPI_ISL_1537995, EPI_ISL_1537996, EPI_ISL_1537997, EPI_ISL_1537998, EPI_ISL_1537999, EPI_ISL_1538000, EPI_ISL_1538001, EPI_ISL_1538002, EPI_ISL_1538003, EPI_ISL_1538004, EPI_ISL_1538005, EPI_ISL_1538006, EPI_ISL_1538007, EPI_ISL_1538008, EPI_ISL_1538009, EPI_ISL_1538010, EPI_ISL_1538011, EPI_ISL_1538012, EPI_ISL_1538013, EPI_ISL_1538014, EPI_ISL_1538015, EPI_ISL_1538016, EPI_ISL_1538017, EPI_ISL_1538018, EPI_ISL_1538019, EPI_ISL_1538021, EPI_ISL_1538022, EPI_ISL_1538023, EPI_ISL_1538024, EPI_ISL_1538025, EPI_ISL_1538026, EPI_ISL_1538027, EPI_ISL_1538028, EPI_ISL_1538030, EPI_ISL_1538032, EPI_ISL_1538033, EPI_ISL_1538034, EPI_ISL_1538035, EPI_ISL_1538037, EPI_ISL_1538038, EPI_ISL_1538041, EPI_ISL_1538042, EPI_ISL_1538044, EPI_ISL_1538045, EPI_ISL_1538046, EPI_ISL_1538047, EPI_ISL_1538048, EPI_ISL_1538050, EPI_ISL_1538051, EPI_ISL_1538052, EPI_ISL_1538054, EPI_ISL_1538055, EPI_ISL_1538056 | | | |
| see above | Lighthouse Lab in Glasgow | Wellcome Sanger Institute for the COVID-19 Genomics UK (COG-UK) Consortium | Harper VanSteenhouse, Yumi Kasai, David Gray, Carol Clugston, Anna Dominiczak and Alex Alderton, Roberto Amato, Jeffrey Barrett, Sonia Goncalves, Ewan Harrison, David K. Jackson, Ian Johnston, Dominic Kwiatkowski, Cordelia Langford, John Sillitoe on behalf of the Wellcome Sanger Institute COVID-19 Surveillance Team |
| EPI_ISL_1538057 | Lighthouse Lab in Cambridge | Wellcome Sanger Institute for the COVID-19 Genomics UK (COG-UK) Consortium | Rob Howes, The Lighthouse Lab in Cambridge and Alex Alderton, Roberto Amato, Jeffrey Barrett, Sonia Goncalves, Ewan Harrison, David K. Jackson, Ian Johnston, Dominic Kwiatkowski, Cordelia Langford, John Sillitoe on behalf of the Wellcome Sanger Institute COVID-19 Surveillance Team |
| EPI_ISL_1538058, EPI_ISL_1538059, EPI_ISL_1538060, EPI_ISL_1538061, EPI_ISL_1538062 | Lighthouse Lab in Glasgow | Wellcome Sanger Institute for the COVID-19 Genomics UK (COG-UK) Consortium | Harper VanSteenhouse, Yumi Kasai, David Gray, Carol Clugston, Anna Dominiczak and Alex Alderton, Roberto Amato, Jeffrey Barrett, Sonia Goncalves, Ewan Harrison, David K. Jackson, Ian Johnston, Dominic Kwiatkowski, Cordelia Langford, John Sillitoe on behalf of the Wellcome Sanger Institute COVID-19 Surveillance Team |
| EPI_ISL_1538063 | Lighthouse Lab in Cambridge | Wellcome Sanger Institute for the COVID-19 Genomics UK (COG-UK) Consortium | Rob Howes, The Lighthouse Lab in Cambridge and Alex Alderton, Roberto Amato, Jeffrey Barrett, Sonia Goncalves, Ewan Harrison, David K. Jackson, Ian Johnston, Dominic Kwiatkowski, Cordelia Langford, John Sillitoe on behalf of the Wellcome Sanger Institute COVID-19 Surveillance Team |
| EPI_ISL_1538064, EPI_ISL_1538065, EPI_ISL_1538066 | Lighthouse Lab in Glasgow | Wellcome Sanger Institute for the COVID-19 Genomics UK (COG-UK) Consortium | Harper VanSteenhouse, Yumi Kasai, David Gray, Carol Clugston, Anna Dominiczak and Alex Alderton, Roberto Amato, Jeffrey Barrett, Sonia Goncalves, Ewan Harrison, David K. Jackson, Ian Johnston, Dominic Kwiatkowski, Cordelia Langford, John Sillitoe on behalf of the Wellcome Sanger Institute COVID-19 Surveillance Team |
| EPI_ISL_1538067 | Lighthouse Lab in Cambridge | Wellcome Sanger Institute for the COVID-19 Genomics UK (COG-UK) Consortium | Rob Howes, The Lighthouse Lab in Cambridge and Alex Alderton, Roberto Amato, Jeffrey Barrett, Sonia Goncalves, Ewan Harrison, David K. Jackson, Ian Johnston, Dominic Kwiatkowski, Cordelia Langford, John Sillitoe on behalf of the Wellcome Sanger Institute COVID-19 Surveillance Team |
| EPI_ISL_1538068 | Lighthouse Lab in Glasgow | Wellcome Sanger Institute for the COVID-19 Genomics UK (COG-UK) Consortium | Harper VanSteenhouse, Yumi Kasai, David Gray, Carol Clugston, Anna Dominiczak and Alex Alderton, Roberto Amato, Jeffrey Barrett, Sonia Goncalves, Ewan Harrison, David K. Jackson, Ian Johnston, Dominic Kwiatkowski, Cordelia Langford, John Sillitoe on behalf of the Wellcome Sanger Institute COVID-19 Surveillance Team |
| EPI_ISL_1538072 | Lighthouse Lab in Cambridge | Wellcome Sanger Institute for the COVID-19 Genomics UK (COG-UK) Consortium | Rob Howes, The Lighthouse Lab in Cambridge and Alex Alderton, Roberto Amato, Jeffrey Barrett, Sonia Goncalves, Ewan Harrison, David K. Jackson, Ian Johnston, Dominic Kwiatkowski, Cordelia Langford, John Sillitoe on behalf of the Wellcome Sanger Institute COVID-19 Surveillance Team |

### Switzerland

We gratefully acknowledge the following Authors from the Originating laboratories responsible for obtaining the specimens, as well as the Submitting laboratories where the genome data were generated and shared via GISAID, on which this research is based.

All Submitters of data may be contacted directly via www.gisaid.org

Authors are sorted alphabetically.

| Accession ID | Originating Laboratory | Submitting Laboratory | Authors |
|---|---|---|---|
| EPI_ISL_1002357 | Viollier AG | Department of Biosystems Science and Engineering, ETH Zürich | Chaoran Chen, Sarah Nadeau, Catharine Aquino, Ivan Topolsky, Philipp Jablonski, Lara Fuhrmann, David Dreifuss, Katharina Jahn, Andreia Cabral de Gouvea, Maria Domenica Moccia, Simon Grüter, Timothy Sykes, Lennart Opitz, Griffin White, Laura Neff, Doris Popovic, Andrea Patrignani, Jay Tracy, Ralph Schlapbach, Christiane Beckmann, Maurice Redondo, Olivier Kobel, Christoph Noppen, Sophie Seidel, Noemie Santamaria de Souza, Niko Beerenwinkel, Tanja Stadler |
| EPI_ISL_1004814, EPI_ISL_1004815, EPI_ISL_1004816, EPI_ISL_1004817, EPI_ISL_1004818, EPI_ISL_1004819, EPI_ISL_1004820, EPI_ISL_1004821, EPI_ISL_1004822, EPI_ISL_1004823, EPI_ISL_1004824, EPI_ISL_1004825, EPI_ISL_1004826, EPI_ISL_1004827, EPI_ISL_1004828, EPI_ISL_1004829, EPI_ISL_1004830, EPI_ISL_1004831, EPI_ISL_1004832, EPI_ISL_1004833, EPI_ISL_1004834, EPI_ISL_1004835, EPI_ISL_1004836, EPI_ISL_1004837, EPI_ISL_1004838, EPI_ISL_1004839, EPI_ISL_1004840, EPI_ISL_1004841, EPI_ISL_1004842, EPI_ISL_1004843, EPI_ISL_1004844, EPI_ISL_1004845, EPI_ISL_1004846, EPI_ISL_1004847, EPI_ISL_1004848, EPI_ISL_1004849, EPI_ISL_1004850, EPI_ISL_1004851, EPI_ISL_1004852, EPI_ISL_1004853, EPI_ISL_1004854, EPI_ISL_1004855, EPI_ISL_1004856, EPI_ISL_1004857, EPI_ISL_1004858, EPI_ISL_1004859, EPI_ISL_1004860, EPI_ISL_1004861, EPI_ISL_1004862, EPI_ISL_1004863, EPI_ISL_1004864, EPI_ISL_1004865, EPI_ISL_1004866, EPI_ISL_1004867, EPI_ISL_1004868, EPI_ISL_1004869, EPI_ISL_1004870, EPI_ISL_1004871, EPI_ISL_1004872, EPI_ISL_1004873, EPI_ISL_1004874, EPI_ISL_1004898, EPI_ISL_1004899, EPI_ISL_1004900, EPI_ISL_1004901, EPI_ISL_1004902, EPI_ISL_1004903, EPI_ISL_1004904, EPI_ISL_1004905, EPI_ISL_1004906, EPI_ISL_1004907, EPI_ISL_1004908, EPI_ISL_1004909, EPI_ISL_1004910, EPI_ISL_1004911, EPI_ISL_1004912, EPI_ISL_1004913, EPI_ISL_1004914, EPI_ISL_1004915, EPI_ISL_1004916, EPI_ISL_1004917, EPI_ISL_1004918, EPI_ISL_1004919, EPI_ISL_1004920, EPI_ISL_1004921, EPI_ISL_1004922, EPI_ISL_1004923, EPI_ISL_1004924, EPI_ISL_1004925, EPI_ISL_1004926, EPI_ISL_1004927, EPI_ISL_1004928, EPI_ISL_1004929, EPI_ISL_1004930, EPI_ISL_1004931, EPI_ISL_1004932, EPI_ISL_1004933, EPI_ISL_1004934, EPI_ISL_1004935, EPI_ISL_1004936, EPI_ISL_1004937, EPI_ISL_1004938, EPI_ISL_1004939, EPI_ISL_1004940, EPI_ISL_1004941, EPI_ISL_1004942, EPI_ISL_1004943, EPI_ISL_1004944, EPI_ISL_1004945, EPI_ISL_1004946, EPI_ISL_1004947, EPI_ISL_1004948, EPI_ISL_1004949, EPI_ISL_1004950, EPI_ISL_1004951, EPI_ISL_1004952, EPI_ISL_1004953, EPI_ISL_1004954, EPI_ISL_1004955, EPI_ISL_1004956, EPI_ISL_1004957, EPI_ISL_1004958, EPI_ISL_1004959, EPI_ISL_1004960, EPI_ISL_1004961, EPI_ISL_1004962, EPI_ISL_1004964, EPI_ISL_1004965, EPI_ISL_1004966, EPI_ISL_1004967, EPI_ISL_1004968, EPI_ISL_1004969, EPI_ISL_1004970, EPI_ISL_1004971, EPI_ISL_1004972, EPI_ISL_1004973, EPI_ISL_1004974, EPI_ISL_1004975 | | | |
| see above | Viollier AG | Department of Biosystems Science and Engineering, ETH Zürich | Chaoran Chen, Sarah Nadeau, Ivan Topolsky, Emmanouil Dermitzakis, Keith Harshman, Ioannis Xenarios, Henri Pegeot, Lorenzo Cerutti, Deborah Penet, Philipp Jablonski, Lara Fuhrmann, David Dreifuss, Katharina Jahn, Christiane Beckmann, Maurice Redondo, Olivier Kobel, Christoph Noppen, Sophie Seidel, Noemie Santamaria de Souza, Niko Beerenwinkel, Tanja Stadler |
| EPI_ISL_1005007, EPI_ISL_1005008, EPI_ISL_1005009, EPI_ISL_1005010, EPI_ISL_1005011, EPI_ISL_1005038, EPI_ISL_1005049, EPI_ISL_1005050, EPI_ISL_1005051, EPI_ISL_1005052, EPI_ISL_1005053, EPI_ISL_1005071, EPI_ISL_1005072, EPI_ISL_1005073, EPI_ISL_1005074, EPI_ISL_1005075, EPI_ISL_1005097, EPI_ISL_1005106, EPI_ISL_1005107, EPI_ISL_1005108, EPI_ISL_1005109, EPI_ISL_1005110, EPI_ISL_1005167, EPI_ISL_1005179, EPI_ISL_1005180, EPI_ISL_1005181, EPI_ISL_1005182, EPI_ISL_1005183, EPI_ISL_1005196, EPI_ISL_1005197, EPI_ISL_1005198, EPI_ISL_1005202, EPI_ISL_1005213, EPI_ISL_1005214, EPI_ISL_1005215, EPI_ISL_1005216, EPI_ISL_1005217 | | | |
| see above | Viollier AG | Department of Biosystems Science and Engineering, ETH Zürich | Chaoran Chen, Sarah Nadeau, Catharine Aquino, Ivan Topolsky, Philipp Jablonski, Lara Fuhrmann, David Dreifuss, Katharina Jahn, Andreia Cabral de Gouvea, Maria Domenica Moccia, Simon Grüter, Timothy Sykes, Lennart Opitz, Griffin White, Laura Neff, Doris Popovic, Andrea Patrignani, Jay Tracy, Ralph Schlapbach, Christiane Beckmann, Maurice Redondo, Olivier Kobel, Christoph Noppen, Sophie Seidel, Noemie Santamaria de Souza, Niko Beerenwinkel, Tanja Stadler |
| EPI_ISL_1007614, EPI_ISL_1007615 | CHUV | Laboratory of genomics and metagenomics, Institute of Microbiology, University Hospital Centre and University of Lausanne, Switzerland | Trestan Pillonel, Damien Jacot, Sébastien Aeby, Gilbert Greub, Claire Bertelli |
| EPI_ISL_1036105, EPI_ISL_1036106, EPI_ISL_1036107, EPI_ISL_1036108, EPI_ISL_1036109, EPI_ISL_1036110, EPI_ISL_1036111, EPI_ISL_1036112, EPI_ISL_1036114, EPI_ISL_1036115, EPI_ISL_1036117, EPI_ISL_1036118, EPI_ISL_1036119, EPI_ISL_1036120, EPI_ISL_1036121, EPI_ISL_1036122, EPI_ISL_1036123, EPI_ISL_1036124, EPI_ISL_1036125, EPI_ISL_1036128, EPI_ISL_1036129, EPI_ISL_1036131, EPI_ISL_1036132, EPI_ISL_1036133, EPI_ISL_1036134, EPI_ISL_1036136, EPI_ISL_1036138, EPI_ISL_1036139 | | | |
| see above | Labormedizinisches Zentrum Dr Risch | University Hospital Basel, Clinical Bacteriology | Tim Roloff, Madlen Stange, Helena MB Seth-Smith, Alfredo Mari, Karoline Leuzinger, Julia Bielicki, Nadia Wohlwend,Martin Risch, Lorenz Risch, Manuel Battegay, Hans Hirsch, Adrian Egli |
| EPI_ISL_1119024, EPI_ISL_1119025, EPI_ISL_1119026, EPI_ISL_1119027, EPI_ISL_1119028, EPI_ISL_1119029 | Viollier AG | Department of Biosystems Science and Engineering, ETH Zürich | Chaoran Chen, Sarah Nadeau, Catharine Aquino, Ivan Topolsky, Philipp Jablonski, Lara Fuhrmann, David Dreifuss, Katharina Jahn, Andreia Cabral de Gouvea, Maria Domenica Moccia, Simon Grüter, Timothy Sykes, Lennart Opitz, Griffin White, Laura Neff, Doris Popovic, Andrea Patrignani, Jay Tracy, Ralph Schlapbach, Christiane Beckmann, Maurice Redondo, Olivier Kobel, Christoph Noppen, Sophie Seidel, Noemie Santamaria de Souza, Niko Beerenwinkel, Tanja Stadler |
| EPI_ISL_1119041, EPI_ISL_1119042, EPI_ISL_1119043, EPI_ISL_1119044, EPI_ISL_1119045, EPI_ISL_1119046, EPI_ISL_1119047, EPI_ISL_1119048, EPI_ISL_1119049, EPI_ISL_1119050, EPI_ISL_1119202, EPI_ISL_1119222 | | | |
| see above | Viollier AG | Department of Biosystems Science and Engineering, ETH Zürich | Chaoran Chen, Sarah Nadeau, Ivan Topolsky, Emmanouil Dermitzakis, Keith Harshman, Ioannis Xenarios, Henri Pegeot, Lorenzo Cerutti, Deborah Penet, Philipp Jablonski, Lara Fuhrmann, David Dreifuss, Katharina Jahn, Christiane Beckmann, Maurice Redondo, Olivier Kobel, Christoph Noppen, Sophie Seidel, Noemie Santamaria de Souza, Niko Beerenwinkel, Tanja Stadler |
| EPI_ISL_1119234, EPI_ISL_1119235, EPI_ISL_1119236, EPI_ISL_1119237 | Viollier AG | Department of Biosystems Science and Engineering, ETH Zürich | Chaoran Chen, Sarah Nadeau, Catharine Aquino, Ivan Topolsky, Philipp Jablonski, Lara Fuhrmann, David Dreifuss, Katharina Jahn, Andreia Cabral de Gouvea, Maria Domenica Moccia, Simon Grüter, Timothy Sykes, Lennart Opitz, Griffin White, Laura Neff, Doris Popovic, Andrea Patrignani, Jay Tracy, Ralph Schlapbach, Christiane Beckmann, Maurice Redondo, Olivier Kobel, Christoph Noppen, Sophie Seidel, Noemie Santamaria de Souza, Niko Beerenwinkel, Tanja Stadler |
| EPI_ISL_1119238 | Viollier AG | Department of Biosystems Science and Engineering, ETH Zürich | Chaoran Chen, Sarah Nadeau, Ivan Topolsky, Emmanouil Dermitzakis, Keith Harshman, Ioannis Xenarios, Henri Pegeot, Lorenzo Cerutti, Deborah Penet, Philipp Jablonski, Lara Fuhrmann, David Dreifuss, Katharina Jahn, Christiane Beckmann, Maurice Redondo, Olivier Kobel, Christoph Noppen, Sophie Seidel, Noemie Santamaria de Souza, Niko Beerenwinkel, Tanja Stadler |
| EPI_ISL_1119241 | Viollier AG | Department of Biosystems Science and Engineering, ETH Zürich | Chaoran Chen, Sarah Nadeau, Catharine Aquino, Ivan Topolsky, Philipp Jablonski, Lara Fuhrmann, David Dreifuss, Katharina Jahn, Andreia Cabral de Gouvea, Maria Domenica Moccia, Simon Grüter, Timothy Sykes, Lennart Opitz, Griffin White, Laura Neff, Doris Popovic, Andrea Patrignani, Jay Tracy, Ralph Schlapbach, Christiane Beckmann, Maurice Redondo, Olivier Kobel, Christoph Noppen, Sophie Seidel, Noemie Santamaria de Souza, Niko Beerenwinkel, Tanja Stadler |
| EPI_ISL_1119244 | Viollier AG | Department of Biosystems Science and Engineering, ETH Zürich | Chaoran Chen, Sarah Nadeau, Ivan Topolsky, Emmanouil Dermitzakis, Keith Harshman, Ioannis Xenarios, Henri Pegeot, Lorenzo Cerutti, Deborah Penet, Philipp Jablonski, Lara Fuhrmann, David Dreifuss, Katharina Jahn, Christiane Beckmann, Maurice Redondo, Olivier Kobel, Christoph Noppen, Sophie Seidel, Noemie Santamaria de Souza, Niko Beerenwinkel, Tanja Stadler |
| EPI_ISL_1119340, EPI_ISL_1119341, EPI_ISL_1119342, EPI_ISL_1119343, EPI_ISL_1119348, EPI_ISL_1119349, EPI_ISL_1119350 | Viollier AG | Department of Biosystems Science and Engineering, ETH Zürich | Chaoran Chen, Sarah Nadeau, Catharine Aquino, Ivan Topolsky, Philipp Jablonski, Lara Fuhrmann, David Dreifuss, Katharina Jahn, Andreia Cabral de Gouvea, Maria Domenica Moccia, Simon Grüter, Timothy Sykes, Lennart Opitz, Griffin White, Laura Neff, Doris Popovic, Andrea Patrignani, Jay Tracy, Ralph Schlapbach, Christiane Beckmann, Maurice Redondo, Olivier Kobel, Christoph Noppen, Sophie Seidel, Noemie Santamaria de Souza, Niko Beerenwinkel, Tanja Stadler |
| EPI_ISL_1119351 | Viollier AG | Department of Biosystems Science and Engineering, ETH Zürich | Chaoran Chen, Sarah Nadeau, Ivan Topolsky, Emmanouil Dermitzakis, Keith Harshman, Ioannis Xenarios, Henri Pegeot, Lorenzo Cerutti, Deborah Penet, Philipp Jablonski, Lara Fuhrmann, David Dreifuss, Katharina Jahn, Christiane Beckmann, Maurice Redondo, Olivier Kobel, Christoph Noppen, Sophie Seidel, Noemie Santamaria de Souza, Niko Beerenwinkel, Tanja Stadler |
| EPI_ISL_1119367, EPI_ISL_1119369 | Viollier AG | Department of Biosystems Science and Engineering, ETH Zürich | Chaoran Chen, Sarah Nadeau, Catharine Aquino, Ivan Topolsky, Philipp Jablonski, Lara Fuhrmann, David Dreifuss, Katharina Jahn, Andreia Cabral de Gouvea, Maria Domenica Moccia, Simon Grüter, Timothy Sykes, Lennart Opitz, Griffin White, Laura Neff, Doris Popovic, Andrea Patrignani, Jay Tracy, Ralph Schlapbach, Christiane Beckmann, Maurice Redondo, Olivier Kobel, Christoph Noppen, Sophie Seidel, Noemie Santamaria de Souza, Niko Beerenwinkel, Tanja Stadler |
| EPI_ISL_1119370 | Viollier AG | Department of Biosystems Science and Engineering, ETH Zürich | Chaoran Chen, Sarah Nadeau, Ivan Topolsky, Emmanouil Dermitzakis, Keith Harshman, Ioannis Xenarios, Henri Pegeot, Lorenzo Cerutti, Deborah Penet, Philipp Jablonski, Lara Fuhrmann, David Dreifuss, Katharina Jahn, Christiane Beckmann, Maurice Redondo, Olivier Kobel, Christoph Noppen, Sophie Seidel, Noemie Santamaria de Souza, Niko Beerenwinkel, Tanja Stadler |

We gratefully acknowledge the following Authors from the Originating laboratories responsible for obtaining the specimens, as well as the Submitting laboratories where the genome data were generated and shared via GISAID, on which this research is based.

All Submitters of data may be contacted directly via www.gisaid.org

Authors are sorted alphabetically.

| Accession ID | Originating Laboratory | Submitting Laboratory | Authors |
|---|---|---|---|
| EPI_ISL_1001828 | Viollier AG | Department of Biosystems Science and Engineering, ETH Zürich | Chaoran Chen, Sarah Nadeau, Catharine Aquino, Ivan Topolsky, Philipp Jablonski, Lara Fuhrmann, David Dreifuss, Katharina Jahn, Andreia Cabral de Gouvea, Maria Domenica Moccia, Simon Grüter, Timothy Sykes, Lennart Opitz, Griffin White, Laura Neff, Doris Popovic, Andrea Patrignani, Jay Tracy, Ralph Schlapbach, Christiane Beckmann, Maurice Redondo, Olivier Kobel, Christoph Noppen, Sophie Seidel, Noemie Santamaria de Souza, Niko Beerenwinkel, Tanja Stadler |
| EPI_ISL_1001832 | Viollier AG | Department of Biosystems Science and Engineering, ETH Zürich | Christian Beisel, Sarah Nadeau, Chaoran Chen, Ivan Topolsky, Philipp Jablonski, Lara Fuhrmann, David Dreifuss, Katharina Jahn, Rebecca Denes, Ina Nissen, Natascha Santacroce, Elodie Burcklen, Christiane Beckmann, Maurice Redondo, Olivier Kobel, Christoph Noppen, Sophie Seidel, Noemie Santamaria de Souza, Niko Beerenwinkel, Tanja Stadler |
| EPI_ISL_1001833, EPI_ISL_1001834 | Viollier AG | Department of Biosystems Science and Engineering, ETH Zürich | Chaoran Chen, Sarah Nadeau, Catharine Aquino, Ivan Topolsky, Philipp Jablonski, Lara Fuhrmann, David Dreifuss, Katharina Jahn, Andreia Cabral de Gouvea, Maria Domenica Moccia, Simon Grüter, Timothy Sykes, Lennart Opitz, Griffin White, Laura Neff, Doris Popovic, Andrea Patrignani, Jay Tracy, Ralph Schlapbach, Christiane Beckmann, Maurice Redondo, Olivier Kobel, Christoph Noppen, Sophie Seidel, Noemie Santamaria de Souza, Niko Beerenwinkel, Tanja Stadler |
| EPI_ISL_1001835 | Viollier AG | Department of Biosystems Science and Engineering, ETH Zürich | Christian Beisel, Sarah Nadeau, Chaoran Chen, Ivan Topolsky, Philipp Jablonski, Lara Fuhrmann, David Dreifuss, Katharina Jahn, Rebecca Denes, Ina Nissen, Natascha Santacroce, Elodie Burcklen, Christiane Beckmann, Maurice Redondo, Olivier Kobel, Christoph Noppen, Sophie Seidel, Noemie Santamaria de Souza, Niko Beerenwinkel, Tanja Stadler |
| EPI_ISL_1001836, EPI_ISL_1001837, EPI_ISL_1001838, EPI_ISL_1001839, EPI_ISL_1001840, EPI_ISL_1001841, EPI_ISL_1001842, EPI_ISL_1001843, EPI_ISL_1001844, EPI_ISL_1001845, EPI_ISL_1001846, EPI_ISL_1001847, EPI_ISL_1001848, EPI_ISL_1001849, EPI_ISL_1001850 | | | |
| see above | Viollier AG | Department of Biosystems Science and Engineering, ETH Zürich | Chaoran Chen, Sarah Nadeau, Catharine Aquino, Ivan Topolsky, Philipp Jablonski, Lara Fuhrmann, David Dreifuss, Katharina Jahn, Andreia Cabral de Gouvea, Maria Domenica Moccia, Simon Grüter, Timothy Sykes, Lennart Opitz, Griffin White, Laura Neff, Doris Popovic, Andrea Patrignani, Jay Tracy, Ralph Schlapbach, Christiane Beckmann, Maurice Redondo, Olivier Kobel, Christoph Noppen, Sophie Seidel, Noemie Santamaria de Souza, Niko Beerenwinkel, Tanja Stadler |
| EPI_ISL_1001851, EPI_ISL_1001852 | Viollier AG | Department of Biosystems Science and Engineering, ETH Zürich | Christian Beisel, Sarah Nadeau, Chaoran Chen, Ivan Topolsky, Philipp Jablonski, Lara Fuhrmann, David Dreifuss, Katharina Jahn, Rebecca Denes, Ina Nissen, Natascha Santacroce, Elodie Burcklen, Christiane Beckmann, Maurice Redondo, Olivier Kobel, Christoph Noppen, Sophie Seidel, Noemie Santamaria de Souza, Niko Beerenwinkel, Tanja Stadler |
| EPI_ISL_1001853, EPI_ISL_1001854, EPI_ISL_1001856, EPI_ISL_1001857, EPI_ISL_1001858, EPI_ISL_1001859, EPI_ISL_1001860, EPI_ISL_1001861 | Viollier AG | Department of Biosystems Science and Engineering, ETH Zürich | Chaoran Chen, Sarah Nadeau, Catharine Aquino, Ivan Topolsky, Philipp Jablonski, Lara Fuhrmann, David Dreifuss, Katharina Jahn, Andreia Cabral de Gouvea, Maria Domenica Moccia, Simon Grüter, Timothy Sykes, Lennart Opitz, Griffin White, Laura Neff, Doris Popovic, Andrea Patrignani, Jay Tracy, Ralph Schlapbach, Christiane Beckmann, Maurice Redondo, Olivier Kobel, Christoph Noppen, Sophie Seidel, Noemie Santamaria de Souza, Niko Beerenwinkel, Tanja Stadler |
| EPI_ISL_1001862 | Viollier AG | Department of Biosystems Science and Engineering, ETH Zürich | Christian Beisel, Sarah Nadeau, Chaoran Chen, Ivan Topolsky, Philipp Jablonski, Lara Fuhrmann, David Dreifuss, Katharina Jahn, Rebecca Denes, Ina Nissen, Natascha Santacroce, Elodie Burcklen, Christiane Beckmann, Maurice Redondo, Olivier Kobel, Christoph Noppen, Sophie Seidel, Noemie Santamaria de Souza, Niko Beerenwinkel, Tanja Stadler |
| EPI_ISL_1001863, EPI_ISL_1001864 | Viollier AG | Department of Biosystems Science and Engineering, ETH Zürich | Chaoran Chen, Sarah Nadeau, Catharine Aquino, Ivan Topolsky, Philipp Jablonski, Lara Fuhrmann, David Dreifuss, Katharina Jahn, Andreia Cabral de Gouvea, Maria Domenica Moccia, Simon Grüter, Timothy Sykes, Lennart Opitz, Griffin White, Laura Neff, Doris Popovic, Andrea Patrignani, Jay Tracy, Ralph Schlapbach, Christiane Beckmann, Maurice Redondo, Olivier Kobel, Christoph Noppen, Sophie Seidel, Noemie Santamaria de Souza, Niko Beerenwinkel, Tanja Stadler |
| EPI_ISL_1001865 | Viollier AG | Department of Biosystems Science and Engineering, ETH Zürich | Christian Beisel, Sarah Nadeau, Chaoran Chen, Ivan Topolsky, Philipp Jablonski, Lara Fuhrmann, David Dreifuss, Katharina Jahn, Rebecca Denes, Ina Nissen, Natascha Santacroce, Elodie Burcklen, Christiane Beckmann, Maurice Redondo, Olivier Kobel, Christoph Noppen, Sophie Seidel, Noemie Santamaria de Souza, Niko Beerenwinkel, Tanja Stadler |
| EPI_ISL_1001866, EPI_ISL_1001867, EPI_ISL_1001868 | Viollier AG | Department of Biosystems Science and Engineering, ETH Zürich | Chaoran Chen, Sarah Nadeau, Catharine Aquino, Ivan Topolsky, Philipp Jablonski, Lara Fuhrmann, David Dreifuss, Katharina Jahn, Andreia Cabral de Gouvea, Maria Domenica Moccia, Simon Grüter, Timothy Sykes, Lennart Opitz, Griffin White, Laura Neff, Doris Popovic, Andrea Patrignani, Jay Tracy, Ralph Schlapbach, Christiane Beckmann, Maurice Redondo, Olivier Kobel, Christoph Noppen, Sophie Seidel, Noemie Santamaria de Souza, Niko Beerenwinkel, Tanja Stadler |
| EPI_ISL_1001869 | Viollier AG | Department of Biosystems Science and Engineering, ETH Zürich | Christian Beisel, Sarah Nadeau, Chaoran Chen, Ivan Topolsky, Philipp Jablonski, Lara Fuhrmann, David Dreifuss, Katharina Jahn, Rebecca Denes, Ina Nissen, Natascha Santacroce, Elodie Burcklen, Christiane Beckmann, Maurice Redondo, Olivier Kobel, Christoph Noppen, Sophie Seidel, Noemie Santamaria de Souza, Niko Beerenwinkel, Tanja Stadler |
| EPI_ISL_1001870, EPI_ISL_1001871, EPI_ISL_1001872, EPI_ISL_1001873, EPI_ISL_1001874, EPI_ISL_1001875, EPI_ISL_1001876, EPI_ISL_1001877, EPI_ISL_1001878, EPI_ISL_1001882, EPI_ISL_1001883, EPI_ISL_1001884, EPI_ISL_1001885, EPI_ISL_1001886, EPI_ISL_1001887, EPI_ISL_1001888, EPI_ISL_1001889, EPI_ISL_1001890, EPI_ISL_1001891, EPI_ISL_1001892, EPI_ISL_1002193, EPI_ISL_1002194, EPI_ISL_1002195, EPI_ISL_1002196, EPI_ISL_1002197, EPI_ISL_1002198, EPI_ISL_1002199 | | | |
| see above | Viollier AG | Department of Biosystems Science and Engineering, ETH Zürich | Chaoran Chen, Sarah Nadeau, Catharine Aquino, Ivan Topolsky, Philipp Jablonski, Lara Fuhrmann, David Dreifuss, Katharina Jahn, Andreia Cabral de Gouvea, Maria Domenica Moccia, Simon Grüter, Timothy Sykes, Lennart Opitz, Griffin White, Laura Neff, Doris Popovic, Andrea Patrignani, Jay Tracy, Ralph Schlapbach, Christiane Beckmann, Maurice Redondo, Olivier Kobel, Christoph Noppen, Sophie Seidel, Noemie Santamaria de Souza, Niko Beerenwinkel, Tanja Stadler |
| EPI_ISL_1002200 | Viollier AG | Department of Biosystems Science and Engineering, ETH Zürich | Christian Beisel, Sarah Nadeau, Chaoran Chen, Ivan Topolsky, Philipp Jablonski, Lara Fuhrmann, David Dreifuss, Katharina Jahn, Rebecca Denes, Ina Nissen, Natascha Santacroce, Elodie Burcklen, Christiane Beckmann, Maurice Redondo, Olivier Kobel, Christoph Noppen, Sophie Seidel, Noemie Santamaria de Souza, Niko Beerenwinkel, Tanja Stadler |
| EPI_ISL_1002201, EPI_ISL_1002202, EPI_ISL_1002203, EPI_ISL_1002204, EPI_ISL_1002205, EPI_ISL_1002206, EPI_ISL_1002207 | Viollier AG | Department of Biosystems Science and Engineering, ETH Zürich | Chaoran Chen, Sarah Nadeau, Catharine Aquino, Ivan Topolsky, Philipp Jablonski, Lara Fuhrmann, David Dreifuss, Katharina Jahn, Andreia Cabral de Gouvea, Maria Domenica Moccia, Simon Grüter, Timothy Sykes, Lennart Opitz, Griffin White, Laura Neff, Doris Popovic, Andrea Patrignani, Jay Tracy, Ralph Schlapbach, Christiane Beckmann, Maurice Redondo, Olivier Kobel, Christoph Noppen, Sophie Seidel, Noemie Santamaria de Souza, Niko Beerenwinkel, Tanja Stadler |
| EPI_ISL_1002208, EPI_ISL_1002211, EPI_ISL_1002212, EPI_ISL_1002213, EPI_ISL_1002214, EPI_ISL_1002215, EPI_ISL_1002216 | Viollier AG | Department of Biosystems Science and Engineering, ETH Zürich | Christian Beisel, Sarah Nadeau, Chaoran Chen, Ivan Topolsky, Philipp Jablonski, Lara Fuhrmann, David Dreifuss, Katharina Jahn, Rebecca Denes, Ina Nissen, Natascha Santacroce, Elodie Burcklen, Christiane Beckmann, Maurice Redondo, Olivier Kobel, Christoph Noppen, Sophie Seidel, Noemie Santamaria de Souza, Niko Beerenwinkel, Tanja Stadler |
| EPI_ISL_1002217, EPI_ISL_1002218, EPI_ISL_1002219, EPI_ISL_1002220, EPI_ISL_1002221, EPI_ISL_1002222, EPI_ISL_1002223, EPI_ISL_1002224, EPI_ISL_1002225, EPI_ISL_1002226, EPI_ISL_1002227, EPI_ISL_1002228, EPI_ISL_1002229, EPI_ISL_1002230, EPI_ISL_1002231, EPI_ISL_1002232, EPI_ISL_1002233, EPI_ISL_1002234, EPI_ISL_1002235, EPI_ISL_1002236, EPI_ISL_1002237, EPI_ISL_1002238, EPI_ISL_1002239, EPI_ISL_1002240, EPI_ISL_1002241, EPI_ISL_1002242, EPI_ISL_1002243, EPI_ISL_1002244, EPI_ISL_1002245, EPI_ISL_1002246 | | | |
| see above | Viollier AG | Department of Biosystems Science and Engineering, ETH Zürich | Chaoran Chen, Sarah Nadeau, Catharine Aquino, Ivan Topolsky, Philipp Jablonski, Lara Fuhrmann, David Dreifuss, Katharina Jahn, Andreia Cabral de Gouvea, Maria Domenica Moccia, Simon Grüter, Timothy Sykes, Lennart Opitz, Griffin White, Laura Neff, Doris Popovic, Andrea Patrignani, Jay Tracy, Ralph Schlapbach, Christiane Beckmann, Maurice Redondo, Olivier Kobel, Christoph Noppen, Sophie Seidel, Noemie Santamaria de Souza, Niko Beerenwinkel, Tanja Stadler |

We gratefully acknowledge the following Authors from the Originating laboratories responsible for obtaining the specimens, as well as the Submitting laboratories where the genome data were generated and shared via GISAID, on which this research is based.

All Submitters of data may be contacted directly via www.gisaid.org

Authors are sorted alphabetically.

| Accession ID | Originating Laboratory | Submitting Laboratory | Authors |
|---|---|---|---|
| EPI_ISL_1228828, EPI_ISL_1228829, EPI_ISL_1228830, EPI_ISL_1228831, EPI_ISL_1228832, EPI_ISL_1228833, EPI_ISL_1228834, EPI_ISL_1228835, EPI_ISL_1228836, EPI_ISL_1228837, EPI_ISL_1228854, EPI_ISL_1228855, EPI_ISL_1228856, EPI_ISL_1228857, EPI_ISL_1228858, EPI_ISL_1228859, EPI_ISL_1228860, EPI_ISL_1228861, EPI_ISL_1228862, EPI_ISL_1228863, EPI_ISL_1228864, EPI_ISL_1228866, EPI_ISL_1228867, EPI_ISL_1228868, EPI_ISL_1228870, EPI_ISL_1228871, EPI_ISL_1228872, EPI_ISL_1228873, EPI_ISL_1228874, EPI_ISL_1228875, EPI_ISL_1228876, EPI_ISL_1228877, EPI_ISL_1228878, EPI_ISL_1228879, EPI_ISL_1228880, EPI_ISL_1228881, EPI_ISL_1228882, EPI_ISL_1228884, EPI_ISL_1228885, EPI_ISL_1228886, EPI_ISL_1228887, EPI_ISL_1228888, EPI_ISL_1228889, EPI_ISL_1228890, EPI_ISL_1228891, EPI_ISL_1228892, EPI_ISL_1228893, EPI_ISL_1228894, EPI_ISL_1228895, EPI_ISL_1228896, EPI_ISL_1228898, EPI_ISL_1228899, EPI_ISL_1228900, EPI_ISL_1228901, EPI_ISL_1228902, EPI_ISL_1228903, EPI_ISL_1228904, EPI_ISL_1228906, EPI_ISL_1228907 | | | |
| see above | University Hospitals of Geneva, Laboratory of Virology | HUG, Laboratory of Virology and the Health2030 Genome Center | Samuel Cordey, Ana Rita Goncalves, Laurent Kaiser, Lorenzo Cerutti, Henri Pegeot, Melyssa Elies, Deborah Penet, Keith Harshman, Ioannis Xenarios, Emmanouil Dermitzakis |
| EPI_ISL_1259448, EPI_ISL_1259610, EPI_ISL_1259613, EPI_ISL_1259622, EPI_ISL_1259626, EPI_ISL_1259641, EPI_ISL_1259643, EPI_ISL_1259644, EPI_ISL_1259658, EPI_ISL_1259659, EPI_ISL_1259670, EPI_ISL_1259675, EPI_ISL_1259678, EPI_ISL_1259706, EPI_ISL_1259712, EPI_ISL_1259723, EPI_ISL_1259730, EPI_ISL_1259731, EPI_ISL_1259732, EPI_ISL_1259733, EPI_ISL_1259739, EPI_ISL_1259743, EPI_ISL_1259744, EPI_ISL_1259759, EPI_ISL_1260010, EPI_ISL_1260092, EPI_ISL_1260390, EPI_ISL_1260396, EPI_ISL_1260402, EPI_ISL_1260412, EPI_ISL_1260448 | | | |
| see above | Viollier AG | Department of Biosystems Science and Engineering, ETH Zürich | Chaoran Chen, Sarah Nadeau, Ivan Topolsky, Emmanouil Dermitzakis, Keith Harshman, Ioannis Xenarios, Henri Pegeot, Lorenzo Cerutti, Deborah Penet, Philipp Jablonski, Lara Fuhrmann, David Dreifuss, Katharina Jahn, Christiane Beckmann, Maurice Redondo, Olivier Kobel, Christoph Noppen, Sophie Seidel, Noemie Santamaria de Souza, Niko Beerenwinkel, Tanja Stadler |
| EPI_ISL_1295984, EPI_ISL_1295985, EPI_ISL_1295986, EPI_ISL_1295987, EPI_ISL_1295988, EPI_ISL_1295989, EPI_ISL_1295990, EPI_ISL_1295991, EPI_ISL_1295992, EPI_ISL_1295993, EPI_ISL_1295994, EPI_ISL_1295995, EPI_ISL_1295996, EPI_ISL_1295997, EPI_ISL_1295998, EPI_ISL_1295999, EPI_ISL_1296000, EPI_ISL_1296001, EPI_ISL_1296002, EPI_ISL_1296003, EPI_ISL_1296004, EPI_ISL_1296005, EPI_ISL_1296006, EPI_ISL_1296007, EPI_ISL_1296008, EPI_ISL_1296009, EPI_ISL_1296010, EPI_ISL_1296011, EPI_ISL_1296012, EPI_ISL_1296013, EPI_ISL_1296014, EPI_ISL_1296015, EPI_ISL_1296016, EPI_ISL_1296017, EPI_ISL_1296018, EPI_ISL_1296019, EPI_ISL_1296020, EPI_ISL_1296021, EPI_ISL_1296022, EPI_ISL_1296023, EPI_ISL_1296024, EPI_ISL_1296025, EPI_ISL_1296026, EPI_ISL_1296027, EPI_ISL_1296028, EPI_ISL_1296029, EPI_ISL_1296030, EPI_ISL_1296031, EPI_ISL_1296032, EPI_ISL_1296033, EPI_ISL_1296034, EPI_ISL_1296035, EPI_ISL_1296036, EPI_ISL_1296037, EPI_ISL_1296038, EPI_ISL_1296039, EPI_ISL_1296040, EPI_ISL_1296041, EPI_ISL_1296042, EPI_ISL_1296043, EPI_ISL_1296044, EPI_ISL_1296045, EPI_ISL_1296046, EPI_ISL_1296047, EPI_ISL_1296048, EPI_ISL_1296049, EPI_ISL_1296050, EPI_ISL_1296051, EPI_ISL_1296052, EPI_ISL_1296053, EPI_ISL_1296054, EPI_ISL_1296055, EPI_ISL_1296056, EPI_ISL_1296057, EPI_ISL_1296058, EPI_ISL_1296059, EPI_ISL_1296060, EPI_ISL_1296061, EPI_ISL_1296062, EPI_ISL_1296063, EPI_ISL_1296064, EPI_ISL_1296065, EPI_ISL_1296066, EPI_ISL_1296067, EPI_ISL_1296068, EPI_ISL_1296069, EPI_ISL_1296070, EPI_ISL_1296071, EPI_ISL_1296072, EPI_ISL_1296073, EPI_ISL_1296074, EPI_ISL_1296075, EPI_ISL_1296076, EPI_ISL_1296077, EPI_ISL_1296078, EPI_ISL_1296079, EPI_ISL_1296080, EPI_ISL_1296081, EPI_ISL_1296082, EPI_ISL_1296083, EPI_ISL_1296084, EPI_ISL_1296085, EPI_ISL_1296086, EPI_ISL_1296087, EPI_ISL_1296088, EPI_ISL_1296089, EPI_ISL_1296090, EPI_ISL_1296091, EPI_ISL_1296092, EPI_ISL_1296093, EPI_ISL_1296094, EPI_ISL_1296095, EPI_ISL_1296096, EPI_ISL_1296097, EPI_ISL_1296098, EPI_ISL_1296099, EPI_ISL_1296100, EPI_ISL_1296101, EPI_ISL_1296102, EPI_ISL_1296103, EPI_ISL_1296104, EPI_ISL_1296105, EPI_ISL_1296106, EPI_ISL_1296107, EPI_ISL_1296108, EPI_ISL_1296109, EPI_ISL_1296110, EPI_ISL_1296111, EPI_ISL_1296112, EPI_ISL_1296113, EPI_ISL_1296114, EPI_ISL_1296115, EPI_ISL_1296116, EPI_ISL_1296117, EPI_ISL_1296118, EPI_ISL_1296119, EPI_ISL_1296120, EPI_ISL_1296121, EPI_ISL_1296122, EPI_ISL_1296123, EPI_ISL_1296124, EPI_ISL_1296125, EPI_ISL_1296126, EPI_ISL_1296127, EPI_ISL_1296128, EPI_ISL_1296129, EPI_ISL_1296130, EPI_ISL_1296131, EPI_ISL_1296132, EPI_ISL_1296133, EPI_ISL_1296134, EPI_ISL_1296135, EPI_ISL_1296136, EPI_ISL_1296137, EPI_ISL_1296138, EPI_ISL_1296139, EPI_ISL_1296140, EPI_ISL_1296141, EPI_ISL_1296142, EPI_ISL_1296143, EPI_ISL_1296144, EPI_ISL_1296145, EPI_ISL_1296146, EPI_ISL_1296147, EPI_ISL_1296148, EPI_ISL_1296149, EPI_ISL_1296150, EPI_ISL_1296151, EPI_ISL_1296152, EPI_ISL_1296153, EPI_ISL_1296154, EPI_ISL_1296155, EPI_ISL_1296156, EPI_ISL_1296157, EPI_ISL_1296158, EPI_ISL_1296159, EPI_ISL_1296160, EPI_ISL_1296161, EPI_ISL_1296162, EPI_ISL_1296163, EPI_ISL_1296164, EPI_ISL_1296165, EPI_ISL_1296166, EPI_ISL_1296167, EPI_ISL_1296168, EPI_ISL_1296169, EPI_ISL_1296170, EPI_ISL_1296171, EPI_ISL_1296172, EPI_ISL_1296173, EPI_ISL_1296174, EPI_ISL_1296175, EPI_ISL_1296176, EPI_ISL_1296177, EPI_ISL_1296178, EPI_ISL_1296179, EPI_ISL_1296180, EPI_ISL_1296181, EPI_ISL_1296182, EPI_ISL_1296183, EPI_ISL_1296184, EPI_ISL_1296185, EPI_ISL_1296186, EPI_ISL_1296187, EPI_ISL_1296188, EPI_ISL_1296189, EPI_ISL_1296190, EPI_ISL_1296191, EPI_ISL_1296192, EPI_ISL_1296193, EPI_ISL_1296194, EPI_ISL_1296195, EPI_ISL_1296196, EPI_ISL_1296197, EPI_ISL_1296198, EPI_ISL_1296199, EPI_ISL_1296200, EPI_ISL_1296201, EPI_ISL_1296202, EPI_ISL_1296203, EPI_ISL_1296204, EPI_ISL_1296205, EPI_ISL_1296206, EPI_ISL_1296207, EPI_ISL_1296208, EPI_ISL_1296209, EPI_ISL_1296210, EPI_ISL_1296211, EPI_ISL_1296212, EPI_ISL_1296213, EPI_ISL_1296214, EPI_ISL_1296215, EPI_ISL_1296216, EPI_ISL_1296217, EPI_ISL_1296218, EPI_ISL_1296219, EPI_ISL_1296220, EPI_ISL_1296221, EPI_ISL_1296222, EPI_ISL_1296223, EPI_ISL_1296224, EPI_ISL_1296225, EPI_ISL_1296226, EPI_ISL_1296227, EPI_ISL_1296228, EPI_ISL_1296229, EPI_ISL_1296230, EPI_ISL_1296231, EPI_ISL_1296232, EPI_ISL_1296233 | | | |
| see above | University Hospitals of Geneva, Laboratory of Virology | HUG, Laboratory of Virology and the Health2030 Genome Center | Samuel Cordey, Ana Rita Goncalves, Laurent Kaiser, Lorenzo Cerutti, Henri Pegeot, Melyssa Elies, Deborah Penet, Keith Harshman, Ioannis Xenarios, Emmanouil Dermitzakis |
| EPI_ISL_1302990, EPI_ISL_1302992, EPI_ISL_1302993, EPI_ISL_1302994, EPI_ISL_1302995, EPI_ISL_1302996, EPI_ISL_1302997, EPI_ISL_1302998, EPI_ISL_1302999, EPI_ISL_1303000, EPI_ISL_1303001, EPI_ISL_1303002, EPI_ISL_1303003, EPI_ISL_1303004, EPI_ISL_1303005, EPI_ISL_1303006, EPI_ISL_1303007, EPI_ISL_1303008, EPI_ISL_1303009, EPI_ISL_1303010, EPI_ISL_1303011, EPI_ISL_1303012, EPI_ISL_1303013, EPI_ISL_1303014, EPI_ISL_1303015, EPI_ISL_1303016, EPI_ISL_1303017, EPI_ISL_1303018, EPI_ISL_1303019, EPI_ISL_1303020, EPI_ISL_1303021, EPI_ISL_1303022, EPI_ISL_1303023, EPI_ISL_1303024, EPI_ISL_1303025, EPI_ISL_1303026, EPI_ISL_1303027, EPI_ISL_1303028, EPI_ISL_1303029, EPI_ISL_1303030 | | | |
| see above | ADMED Microbiologie | Genomics and Transcriptomics, Philip Morris International | Reto Lienhard, Marie-Lise Tritten, Emmanuel Guedj, Nicolas Sierro, Rémi Dulize, David Bornand, Mehdi Auberson, Maxime Berthouzoz, Nikolai Ivanov, Manuel Peitsch |
| EPI_ISL_1360454 | Viollier AG | Department of Biosystems Science and Engineering, ETH Zürich | Christian Beisel, Sarah Nadeau, Chaoran Chen, Ivan Topolsky, Philipp Jablonski, Lara Fuhrmann, David Dreifuss, Katharina Jahn, Rebecca Denes, Mirjam Feldkamp, Ina Nissen, Natascha Santacroce, Elodie Burcklen, Christiane Beckmann, Maurice Redondo, Olivier Kobel, Christoph Noppen, Sophie Seidel, Noemie Santamaria de Souza, Niko Beerenwinkel, Tanja Stadler |
| EPI_ISL_1360456 | Viollier AG | Department of Biosystems Science and Engineering, ETH Zürich | Chaoran Chen, Sarah Nadeau, Catharine Aquino, Ivan Topolsky, Philipp Jablonski, Lara Fuhrmann, David Dreifuss, Katharina Jahn, Andreia Cabral de Gouvea, Maria Domenica Moccia, Simon Grüter, Timothy Sykes, Lennart Opitz, Griffin White, Laura Neff, Doris Popovic, Andrea Patrignani, Jay Tracy, Ralph Schlapbach, Christiane Beckmann, Maurice Redondo, Olivier Kobel, Christoph Noppen, Sophie Seidel, Noemie Santamaria de Souza, Niko Beerenwinkel, Tanja Stadler |
| EPI_ISL_1360458, EPI_ISL_1360461 | Viollier AG | Department of Biosystems Science and Engineering, ETH Zürich | Chaoran Chen, Sarah Nadeau, Ivan Topolsky, Emmanouil Dermitzakis, Keith Harshman, Ioannis Xenarios, Henri Pegeot, Lorenzo Cerutti, Deborah Penet, Philipp Jablonski, Lara Fuhrmann, David Dreifuss, Katharina Jahn, Christiane Beckmann, Maurice Redondo, Olivier Kobel, Christoph Noppen, Sophie Seidel, Noemie Santamaria de Souza, Niko Beerenwinkel, Tanja Stadler |
| EPI_ISL_1360465 | Viollier AG | Department of Biosystems Science and Engineering, ETH Zürich | Christian Beisel, Sarah Nadeau, Chaoran Chen, Ivan Topolsky, Philipp Jablonski, Lara Fuhrmann, David Dreifuss, Katharina Jahn, Rebecca Denes, Mirjam Feldkamp, Ina Nissen, Natascha Santacroce, Elodie Burcklen, Christiane Beckmann, Maurice Redondo, Olivier Kobel, Christoph Noppen, Sophie Seidel, Noemie Santamaria de Souza, Niko Beerenwinkel, Tanja Stadler |
| EPI_ISL_1360466 | Viollier AG | Department of Biosystems Science and Engineering, ETH Zürich | Chaoran Chen, Sarah Nadeau, Catharine Aquino, Ivan Topolsky, Philipp Jablonski, Lara Fuhrmann, David Dreifuss, Katharina Jahn, Andreia Cabral de Gouvea, Maria Domenica Moccia, Simon Grüter, Timothy Sykes, Lennart Opitz, Griffin White, Laura Neff, Doris Popovic, Andrea Patrignani, Jay Tracy, Ralph Schlapbach, Christiane Beckmann, Maurice Redondo, Olivier Kobel, Christoph Noppen, Sophie Seidel, Noemie Santamaria de Souza, Niko Beerenwinkel, Tanja Stadler |
| EPI_ISL_1360467, EPI_ISL_1360472 | Viollier AG | Department of Biosystems Science and Engineering, ETH Zürich | Christian Beisel, Sarah Nadeau, Chaoran Chen, Ivan Topolsky, Philipp Jablonski, Lara Fuhrmann, David Dreifuss, Katharina Jahn, Rebecca Denes, Mirjam Feldkamp, Ina Nissen, Natascha Santacroce, Elodie Burcklen, Christiane Beckmann, Maurice Redondo, Olivier Kobel, Christoph Noppen, Sophie Seidel, Noemie Santamaria de Souza, Niko Beerenwinkel, Tanja Stadler |
| EPI_ISL_1360473 | Viollier AG | Department of Biosystems Science and Engineering, ETH Zürich | Chaoran Chen, Sarah Nadeau, Catharine Aquino, Ivan Topolsky, Philipp Jablonski, Lara Fuhrmann, David Dreifuss, Katharina Jahn, Andreia Cabral de Gouvea, Maria Domenica Moccia, Simon Grüter, Timothy Sykes, Lennart Opitz, Griffin White, Laura Neff, Doris Popovic, Andrea Patrignani, Jay Tracy, Ralph Schlapbach, Christiane Beckmann, Maurice Redondo, Olivier Kobel, Christoph Noppen, Sophie Seidel, Noemie Santamaria de Souza, Niko Beerenwinkel, Tanja Stadler |
| EPI_ISL_1360474 | Viollier AG | Department of Biosystems Science and Engineering, ETH Zürich | Chaoran Chen, Sarah Nadeau, Ivan Topolsky, Emmanouil Dermitzakis, Keith Harshman, Ioannis Xenarios, Henri Pegeot, Lorenzo Cerutti, Deborah Penet, Philipp Jablonski, Lara Fuhrmann, David Dreifuss, Katharina Jahn, Christiane Beckmann, Maurice Redondo, Olivier Kobel, Christoph Noppen, Sophie Seidel, Noemie Santamaria de Souza, Niko Beerenwinkel, Tanja Stadler |
| EPI_ISL_1360477 | Viollier AG | Department of Biosystems Science and Engineering, ETH Zürich | Christian Beisel, Sarah Nadeau, Chaoran Chen, Ivan Topolsky, Philipp Jablonski, Lara Fuhrmann, David Dreifuss, Katharina Jahn, Rebecca Denes, Mirjam Feldkamp, Ina Nissen, Natascha Santacroce, Elodie Burcklen, Christiane Beckmann, Maurice Redondo, Olivier Kobel, Christoph Noppen, Sophie Seidel, Noemie Santamaria de Souza, Niko Beerenwinkel, Tanja Stadler |
| EPI_ISL_1360479 | Viollier AG | Department of Biosystems Science and Engineering, ETH Zürich | Chaoran Chen, Sarah Nadeau, Catharine Aquino, Ivan Topolsky, Philipp Jablonski, Lara Fuhrmann, David Dreifuss, Katharina Jahn, Andreia Cabral de Gouvea, Maria Domenica Moccia, Simon Grüter, Timothy Sykes, Lennart Opitz, Griffin White, Laura Neff, Doris Popovic, Andrea Patrignani, Jay Tracy, Ralph Schlapbach, Christiane Beckmann, Maurice Redondo, Olivier Kobel, Christoph Noppen, Sophie Seidel, Noemie Santamaria de Souza, Niko Beerenwinkel, Tanja Stadler |

### Victoria/AU

We gratefully acknowledge the following Authors from the Originating laboratories responsible for obtaining the specimens, as well as the Submitting laboratories where the genome data were generated and shared via GISAID, on which this research is based.

All Submitters of data may be contacted directly via www.gisaid.org

Authors are sorted alphabetically.

| Accession ID | Originating Laboratory | Submitting Laboratory | Authors |
|---|---|---|---|
| EPI_ISL_1033125, EPI_ISL_1033128, EPI_ISL_1033129, EPI_ISL_1033130, EPI_ISL_1033136, EPI_ISL_1033137, EPI_ISL_1033138, EPI_ISL_1033139, EPI_ISL_1055266, EPI_ISL_1055268, EPI_ISL_1055269, EPI_ISL_1055271, EPI_ISL_1055283, EPI_ISL_1055284, EPI_ISL_1055285, EPI_ISL_1055287, EPI_ISL_1055288, EPI_ISL_1055289, EPI_ISL_1055290, EPI_ISL_1055291, EPI_ISL_1055292, EPI_ISL_1055293, EPI_ISL_1055294, EPI_ISL_1055295, EPI_ISL_1055296, EPI_ISL_1055300 | | | |
| see above | Microbiological Diagnostic Unit - Public Health Laboratory (MDU-PHL) | MDU-PHL | Seemann T., Sait, M.L., Sherry, N.L. |
| EPI_ISL_1055301, EPI_ISL_1055302 | Victorian Infectious Diseases Reference Laboratory (VIDRL) | VIDRL and MDU-PHL | Caly L., Seemann T., Sait, M.L., Druce J., Sherry, N.L. |
| EPI_ISL_1055303, EPI_ISL_1055304, EPI_ISL_1055305, EPI_ISL_1055306, EPI_ISL_1055308, EPI_ISL_1055310, EPI_ISL_1055311, EPI_ISL_1055312, EPI_ISL_1055313, EPI_ISL_1055314, EPI_ISL_1055315, EPI_ISL_1055316, EPI_ISL_1055317, EPI_ISL_1055319, EPI_ISL_1055320, EPI_ISL_1055321, EPI_ISL_1055322, EPI_ISL_1055323, EPI_ISL_1055324, EPI_ISL_1055325, EPI_ISL_1055326, EPI_ISL_1055327, EPI_ISL_1055329, EPI_ISL_1055330, EPI_ISL_1055332, EPI_ISL_1055334, EPI_ISL_1055335, EPI_ISL_1055336, EPI_ISL_1055337, EPI_ISL_1055338, EPI_ISL_1055339 | | | |
| see above | Microbiological Diagnostic Unit - Public Health Laboratory (MDU-PHL) | MDU-PHL | Seemann T., Sait, M.L., Sherry, N.L. |
| EPI_ISL_1055340, EPI_ISL_1055341 | Victorian Infectious Diseases Reference Laboratory (VIDRL) | VIDRL and MDU-PHL | Caly L., Seemann T., Sait, M.L., Druce J., Sherry, N.L. |
| EPI_ISL_1055342, EPI_ISL_1055343, EPI_ISL_1055344, EPI_ISL_1055345, EPI_ISL_1055346, EPI_ISL_1055347 | Microbiological Diagnostic Unit - Public Health Laboratory (MDU-PHL) | MDU-PHL | Seemann T., Sait, M.L., Sherry, N.L. |
| EPI_ISL_1055348 | Victorian Infectious Diseases Reference Laboratory (VIDRL) | VIDRL and MDU-PHL | Caly L., Seemann T., Sait, M.L., Druce J., Sherry, N.L. |
| EPI_ISL_1055352, EPI_ISL_1055354, EPI_ISL_1055357, EPI_ISL_1061399, EPI_ISL_1061400, EPI_ISL_1061401 | Microbiological Diagnostic Unit - Public Health Laboratory (MDU-PHL) | MDU-PHL | Seemann T., Sait, M.L., Sherry, N.L. |
| EPI_ISL_406844 | Monash Medical Centre | Collaboration between the University of Melbourne at The Peter Doherty Institute for Infection and Immunity, and the Victorian Infectious Disease Reference Laboratory | Caly,L., Seemann,T., Schultz,M., Druce,J. and Taiaroa,G |
| EPI_ISL_416410, EPI_ISL_416411, EPI_ISL_416412, EPI_ISL_416413, EPI_ISL_416415 | Victorian Infectious Diseases Reference Laboratory (VIDRL) | Victorian Infectious Diseases Reference Laboratory and Microbiological Diagnostic Unit Public Health Laboratory, Doherty Institute | Caly L., Seemann T., Schultz M., Druce J., Taiaroa, G. |
| EPI_ISL_416514, EPI_ISL_416515, EPI_ISL_416516 | Victorian Infectious Diseases Reference Laboratory (VIDRL) | Victorian Infectious Diseases Reference Laboratory and Microbiological Diagnostic Unit Public Health Laboratory, Doherty Institute | Caly L., Seemann T., Schultz M., Taiaroa, G., Druce J. |
| EPI_ISL_419714, EPI_ISL_419715, EPI_ISL_419716, EPI_ISL_419717, EPI_ISL_419718, EPI_ISL_419719, EPI_ISL_419720, EPI_ISL_419721, EPI_ISL_419722, EPI_ISL_419723, EPI_ISL_419725, EPI_ISL_419726, EPI_ISL_419727, EPI_ISL_419728, EPI_ISL_419729, EPI_ISL_419730, EPI_ISL_419731, EPI_ISL_419732 | | | |
| see above | Microbiological Diagnostic Unit Public Health Laboratory | Microbiological Diagnostic Unit Public Health Laboratory | Seemann T., Schultz M., Sait, M., Sherry, N. |
| EPI_ISL_419733, EPI_ISL_419736, EPI_ISL_419737, EPI_ISL_419739, EPI_ISL_419740, EPI_ISL_419741, EPI_ISL_419742, EPI_ISL_419743, EPI_ISL_419744, EPI_ISL_419745, EPI_ISL_419746, EPI_ISL_419747, EPI_ISL_419748, EPI_ISL_419749, EPI_ISL_419750, EPI_ISL_419751, EPI_ISL_419752, EPI_ISL_419753, EPI_ISL_419754, EPI_ISL_419755, EPI_ISL_419756, EPI_ISL_419757, EPI_ISL_419758, EPI_ISL_419759, EPI_ISL_419760, EPI_ISL_419761, EPI_ISL_419762, EPI_ISL_419763, EPI_ISL_419764, EPI_ISL_419765, EPI_ISL_419766, EPI_ISL_419767, EPI_ISL_419768, EPI_ISL_419769, EPI_ISL_419770, EPI_ISL_419771, EPI_ISL_419772, EPI_ISL_419773, EPI_ISL_419774, EPI_ISL_419775, EPI_ISL_419776, EPI_ISL_419777, EPI_ISL_419778, EPI_ISL_419779, EPI_ISL_419780, EPI_ISL_419781, EPI_ISL_419782, EPI_ISL_419783, EPI_ISL_419784, EPI_ISL_419785, EPI_ISL_419786, EPI_ISL_419787, EPI_ISL_419788, EPI_ISL_419789, EPI_ISL_419790, EPI_ISL_419791, EPI_ISL_419792, EPI_ISL_419793, EPI_ISL_419794, EPI_ISL_419795, EPI_ISL_419796, EPI_ISL_419797, EPI_ISL_419798, EPI_ISL_419799, EPI_ISL_419800, EPI_ISL_419801, EPI_ISL_419802, EPI_ISL_419803, EPI_ISL_419804, EPI_ISL_419805, EPI_ISL_419806, EPI_ISL_419807, EPI_ISL_419808, EPI_ISL_419809, EPI_ISL_419810, EPI_ISL_419811, EPI_ISL_419812, EPI_ISL_419813, EPI_ISL_419814, EPI_ISL_419816, EPI_ISL_419817, EPI_ISL_419818, EPI_ISL_419819, EPI_ISL_419820, EPI_ISL_419821, EPI_ISL_419822 | | | |
| see above | Victorian Infectious Diseases Reference Laboratory (VIDRL) | Victorian Infectious Diseases Reference Laboratory and Microbiological Diagnostic Unit Public Health Laboratory, Doherty Institute | Caly L., Seemann T., Sait, M., Schultz M., Druce J., Sherry, N. |
| EPI_ISL_419823, EPI_ISL_419824, EPI_ISL_419825 | Microbiological Diagnostic Unit Public Health Laboratory | Microbiological Diagnostic Unit Public Health Laboratory | Seemann T., Schultz M., Sait, M., Sherry, N. |
| EPI_ISL_419826 | Victorian Infectious Diseases Reference Laboratory (VIDRL) | Victorian Infectious Diseases Reference Laboratory and Microbiological Diagnostic Unit Public Health Laboratory, Doherty Institute | Caly L., Seemann T., Sait, M., Schultz M., Druce J., Sherry, N. |
| EPI_ISL_419827, EPI_ISL_419828, EPI_ISL_419829, EPI_ISL_419830 | Microbiological Diagnostic Unit Public Health Laboratory | Microbiological Diagnostic Unit Public Health Laboratory | Seemann T., Schultz M., Sait, M., Sherry, N. |
| EPI_ISL_419834, EPI_ISL_419836, EPI_ISL_419837, EPI_ISL_419838, EPI_ISL_419839, EPI_ISL_419840, EPI_ISL_419841, EPI_ISL_419842, EPI_ISL_419843, EPI_ISL_419844, EPI_ISL_419845, EPI_ISL_419846, EPI_ISL_419847, EPI_ISL_419848, EPI_ISL_419849, EPI_ISL_419850, EPI_ISL_419851, EPI_ISL_419852, EPI_ISL_419853, EPI_ISL_419854, EPI_ISL_419855, EPI_ISL_419856, EPI_ISL_419857, EPI_ISL_419859, EPI_ISL_419860, EPI_ISL_419861, EPI_ISL_419862, EPI_ISL_419863, EPI_ISL_419864, EPI_ISL_419865, EPI_ISL_419866, EPI_ISL_419867, EPI_ISL_419868, EPI_ISL_419869, EPI_ISL_419870, EPI_ISL_419871, EPI_ISL_419872, EPI_ISL_419873, EPI_ISL_419874, EPI_ISL_419875, EPI_ISL_419876, EPI_ISL_419877, EPI_ISL_419878, EPI_ISL_419879, EPI_ISL_419880, EPI_ISL_419881, EPI_ISL_419882, EPI_ISL_419883, EPI_ISL_419884, EPI_ISL_419885, EPI_ISL_419886, EPI_ISL_419887, EPI_ISL_419888, EPI_ISL_419889, EPI_ISL_419890, EPI_ISL_419891, EPI_ISL_419892, EPI_ISL_419893, EPI_ISL_419894, EPI_ISL_419895, EPI_ISL_419896, EPI_ISL_419897, EPI_ISL_419898, EPI_ISL_419899, EPI_ISL_419900, EPI_ISL_419901, EPI_ISL_419902, EPI_ISL_419903, EPI_ISL_419904, EPI_ISL_419905, EPI_ISL_419906, EPI_ISL_419907, EPI_ISL_419908, EPI_ISL_419909, EPI_ISL_419910, EPI_ISL_419911, EPI_ISL_419912, EPI_ISL_419913, EPI_ISL_419914, EPI_ISL_419915, EPI_ISL_419916, EPI_ISL_419917, EPI_ISL_419918, EPI_ISL_419919, EPI_ISL_419920, EPI_ISL_419921, EPI_ISL_419922, EPI_ISL_419923, EPI_ISL_419924, EPI_ISL_419925, EPI_ISL_419926, EPI_ISL_419927, EPI_ISL_419928, EPI_ISL_419929, EPI_ISL_419931, EPI_ISL_419932, EPI_ISL_419933, EPI_ISL_419934, EPI_ISL_419935, EPI_ISL_419936, EPI_ISL_419937, EPI_ISL_419938, EPI_ISL_419939, EPI_ISL_419940, EPI_ISL_419941, EPI_ISL_419942, EPI_ISL_419943, EPI_ISL_419944, EPI_ISL_419945, EPI_ISL_419946, EPI_ISL_419947, EPI_ISL_419948, EPI_ISL_419949, EPI_ISL_419950, EPI_ISL_419951, EPI_ISL_419952, EPI_ISL_419953, EPI_ISL_419954, EPI_ISL_419955, EPI_ISL_419956, EPI_ISL_419957, EPI_ISL_419958, EPI_ISL_419959, EPI_ISL_419960, EPI_ISL_419961, EPI_ISL_419962, EPI_ISL_419963, EPI_ISL_419964, EPI_ISL_419965, EPI_ISL_419966, EPI_ISL_419967, EPI_ISL_419968, EPI_ISL_419969, EPI_ISL_419970, EPI_ISL_419971, EPI_ISL_419972, EPI_ISL_419973, EPI_ISL_419974, EPI_ISL_419975, EPI_ISL_419976, EPI_ISL_419977, EPI_ISL_419978, EPI_ISL_419979 | | | |
| see above | Victorian Infectious Diseases Reference Laboratory (VIDRL) | Victorian Infectious Diseases Reference Laboratory and Microbiological Diagnostic Unit Public Health Laboratory, Doherty Institute | Caly L., Seemann T., Sait, M., Schultz M., Druce J., Sherry, N. |
| EPI_ISL_419980 | Microbiological Diagnostic Unit Public Health Laboratory | Microbiological Diagnostic Unit Public Health Laboratory | Seemann T., Schultz M., Sait, M., Sherry, N. |
| EPI_ISL_419982, EPI_ISL_419983, EPI_ISL_419984, EPI_ISL_419985, EPI_ISL_419986, EPI_ISL_419987, EPI_ISL_419988, EPI_ISL_419989, EPI_ISL_419990, EPI_ISL_419991, EPI_ISL_419992, EPI_ISL_419993, EPI_ISL_419994, EPI_ISL_419995, EPI_ISL_419996, EPI_ISL_419997, EPI_ISL_419998 | | | |
| see above | Victorian Infectious Diseases Reference Laboratory (VIDRL) | Victorian Infectious Diseases Reference Laboratory and Microbiological Diagnostic Unit Public Health Laboratory, Doherty Institute | Caly L., Seemann T., Sait, M., Schultz M., Druce J., Sherry, N. |
| EPI_ISL_419999, EPI_ISL_420000, EPI_ISL_420001, EPI_ISL_420002, EPI_ISL_420003 | Microbiological Diagnostic Unit Public Health Laboratory | Microbiological Diagnostic Unit Public Health Laboratory | Seemann T., Schultz M., Sait, M., Sherry, N. |
| EPI_ISL_420004, EPI_ISL_420005 | Victorian Infectious Diseases Reference Laboratory (VIDRL) | Victorian Infectious Diseases Reference Laboratory and | Caly L., Seemann T., Sait, M., Schultz M., Druce J., Sherry, N. |

We gratefully acknowledge the following Authors from the Originating laboratories responsible for obtaining the specimens, as well as the Submitting laboratories where the genome data were generated and shared via GISAID, on which this research is based.

All Submitters of data may be contacted directly via www.gisaid.org

Authors are sorted alphabetically.

| Accession ID | Originating Laboratory | Submitting Laboratory | Authors |
|---|---|---|---|
| EPI_ISL_1033126, EPI_ISL_1033127, EPI_ISL_1033131, EPI_ISL_1033132, EPI_ISL_1033133, EPI_ISL_1033134, EPI_ISL_1033141, EPI_ISL_1033144, EPI_ISL_1033145, EPI_ISL_1033146, EPI_ISL_1033147, EPI_ISL_1033148, EPI_ISL_1033149, EPI_ISL_1033150, EPI_ISL_1033151, EPI_ISL_1033152 | | | |
| see above | Microbiological Diagnostic Unit - Public Health Laboratory (MDU-PHL) | MDU-PHL | Seemann T., Sait, M.L., Sherry, N.L. |
| EPI_ISL_1033154, EPI_ISL_1033155 | Victorian Infectious Diseases Reference Laboratory (VIDRL) | VIDRL and MDU-PHL | Caly L., Seemann T., Sait, M.L., Druce J., Sherry, N.L. |
| EPI_ISL_1033156, EPI_ISL_1033157, EPI_ISL_1055265, EPI_ISL_1055267, EPI_ISL_1055270, EPI_ISL_1055272, EPI_ISL_1055274, EPI_ISL_1055275, EPI_ISL_1055276, EPI_ISL_1055277 | Microbiological Diagnostic Unit - Public Health Laboratory (MDU-PHL) | MDU-PHL | Seemann T., Sait, M.L., Sherry, N.L. |
| EPI_ISL_1055278 | Victorian Infectious Diseases Reference Laboratory (VIDRL) | VIDRL and MDU-PHL | Caly L., Seemann T., Sait, M.L., Druce J., Sherry, N.L. |
| EPI_ISL_1055279, EPI_ISL_1055280, EPI_ISL_1055281, EPI_ISL_1055282, EPI_ISL_1055286, EPI_ISL_1055297, EPI_ISL_1055298, EPI_ISL_1055299, EPI_ISL_1055307, EPI_ISL_1055309, EPI_ISL_1055318, EPI_ISL_1055331, EPI_ISL_1055333, EPI_ISL_1055349, EPI_ISL_1055350, EPI_ISL_1055353, EPI_ISL_1055355, EPI_ISL_1055356 | | | |
| see above | Microbiological Diagnostic Unit - Public Health Laboratory (MDU-PHL) | MDU-PHL | Seemann T., Sait, M.L., Sherry, N.L. |
| EPI_ISL_1055358 | Victorian Infectious Diseases Reference Laboratory (VIDRL) | VIDRL and MDU-PHL | Caly L., Seemann T., Sait, M.L., Druce J., Sherry, N.L. |
| EPI_ISL_1055359, EPI_ISL_1055360, EPI_ISL_1061402, EPI_ISL_1061403, EPI_ISL_1249986, EPI_ISL_1249988, EPI_ISL_1249990 | Microbiological Diagnostic Unit - Public Health Laboratory (MDU-PHL) | MDU-PHL | Seemann T., Sait, M.L., Sherry, N.L. |
| EPI_ISL_1249993 | Victorian Infectious Diseases Reference Laboratory (VIDRL) | VIDRL and MDU-PHL | Caly L., Seemann T., Sait, M.L., Druce J., Sherry, N.L. |
| EPI_ISL_1249999, EPI_ISL_1250005 | Microbiological Diagnostic Unit - Public Health Laboratory (MDU-PHL) | MDU-PHL | Seemann T., Sait, M.L., Sherry, N.L. |
| EPI_ISL_562169, EPI_ISL_562172, EPI_ISL_562173, EPI_ISL_562174, EPI_ISL_562591, EPI_ISL_562592 | Microbiological Diagnostic Unit - Public Health Laboratory (MDU-PHL) | MDU-PHL | Seemann, T., Schultz M. B., Sait, M., Sherry, N. |
| EPI_ISL_562593 | Victorian Infectious Diseases Reference Laboratory (VIDRL) | VIDRL and MDU-PHL | Caly, L., Seemann, T., Sait, M., Schultz, M. B., Druce J., Sherry, N. |
| EPI_ISL_562594 | Microbiological Diagnostic Unit - Public Health Laboratory (MDU-PHL) | MDU-PHL | Seemann, T., Schultz M. B., Sait, M., Sherry, N. |
| EPI_ISL_562595 | Victorian Infectious Diseases Reference Laboratory (VIDRL) | VIDRL and MDU-PHL | Caly, L., Seemann, T., Sait, M., Schultz, M. B., Druce J., Sherry, N. |
| EPI_ISL_562596, EPI_ISL_562597, EPI_ISL_562598, EPI_ISL_562599, EPI_ISL_562600, EPI_ISL_562603, EPI_ISL_562604, EPI_ISL_562605, EPI_ISL_562609, EPI_ISL_562612, EPI_ISL_562613, EPI_ISL_562614, EPI_ISL_562615, EPI_ISL_562616, EPI_ISL_562617, EPI_ISL_562620, EPI_ISL_562621, EPI_ISL_562622, EPI_ISL_562623, EPI_ISL_562624, EPI_ISL_562625, EPI_ISL_562626, EPI_ISL_562627, EPI_ISL_562629, EPI_ISL_562631, EPI_ISL_562632, EPI_ISL_562633, EPI_ISL_562634, EPI_ISL_562635, EPI_ISL_562636, EPI_ISL_562637 | | | |
| see above | Microbiological Diagnostic Unit - Public Health Laboratory (MDU-PHL) | MDU-PHL | Seemann, T., Schultz M. B., Sait, M., Sherry, N. |
| EPI_ISL_562640 | Victorian Infectious Diseases Reference Laboratory (VIDRL) | VIDRL and MDU-PHL | Caly, L., Seemann, T., Sait, M., Schultz, M. B., Druce J., Sherry, N. |
| EPI_ISL_562642, EPI_ISL_562643, EPI_ISL_562644 | Microbiological Diagnostic Unit - Public Health Laboratory (MDU-PHL) | MDU-PHL | Seemann, T., Schultz M. B., Sait, M., Sherry, N. |
| EPI_ISL_562645, EPI_ISL_562646 | Victorian Infectious Diseases Reference Laboratory (VIDRL) | VIDRL and MDU-PHL | Caly, L., Seemann, T., Sait, M., Schultz, M. B., Druce J., Sherry, N. |
| EPI_ISL_562648, EPI_ISL_562649, EPI_ISL_562650, EPI_ISL_562651, EPI_ISL_562652, EPI_ISL_562653, EPI_ISL_562654, EPI_ISL_562655, EPI_ISL_562657, EPI_ISL_562661, EPI_ISL_562662, EPI_ISL_562663, EPI_ISL_562664, EPI_ISL_562666, EPI_ISL_562670, EPI_ISL_562673, EPI_ISL_562674, EPI_ISL_562675, EPI_ISL_562676, EPI_ISL_562677, EPI_ISL_562678, EPI_ISL_562679, EPI_ISL_562681, EPI_ISL_562682, EPI_ISL_562683, EPI_ISL_562684, EPI_ISL_562685, EPI_ISL_562686, EPI_ISL_562687, EPI_ISL_562688, EPI_ISL_562691, EPI_ISL_562692, EPI_ISL_562693, EPI_ISL_562694, EPI_ISL_562695, EPI_ISL_562698 | | | |
| see above | Microbiological Diagnostic Unit - Public Health Laboratory (MDU-PHL) | MDU-PHL | Seemann, T., Schultz M. B., Sait, M., Sherry, N. |
| EPI_ISL_562700 | Victorian Infectious Diseases Reference Laboratory (VIDRL) | VIDRL and MDU-PHL | Caly, L., Seemann, T., Sait, M., Schultz, M. B., Druce J., Sherry, N. |
| EPI_ISL_562701, EPI_ISL_562702, EPI_ISL_562706, EPI_ISL_562708, EPI_ISL_562709, EPI_ISL_562710, EPI_ISL_562711, EPI_ISL_562712, EPI_ISL_562713, EPI_ISL_562714, EPI_ISL_562715, EPI_ISL_562716, EPI_ISL_562717, EPI_ISL_562718, EPI_ISL_562719, EPI_ISL_562720, EPI_ISL_562721, EPI_ISL_562722, EPI_ISL_562723, EPI_ISL_562724, EPI_ISL_562725, EPI_ISL_562726, EPI_ISL_562727, EPI_ISL_562728, EPI_ISL_562729, EPI_ISL_562730, EPI_ISL_562731, EPI_ISL_562732, EPI_ISL_562733, EPI_ISL_562734, EPI_ISL_562735, EPI_ISL_562736, EPI_ISL_562737, EPI_ISL_562738, EPI_ISL_562739, EPI_ISL_562740, EPI_ISL_562741, EPI_ISL_562742, EPI_ISL_562743, EPI_ISL_562744, EPI_ISL_562745, EPI_ISL_562746, EPI_ISL_562747, EPI_ISL_562748, EPI_ISL_562749, EPI_ISL_562750, EPI_ISL_562751, EPI_ISL_562752, EPI_ISL_562753, EPI_ISL_562756, EPI_ISL_562761, EPI_ISL_562762, EPI_ISL_562763, EPI_ISL_562764, EPI_ISL_562765, EPI_ISL_562766, EPI_ISL_562767, EPI_ISL_562768, EPI_ISL_562769, EPI_ISL_562770, EPI_ISL_562771, EPI_ISL_562772, EPI_ISL_562773, EPI_ISL_562774, EPI_ISL_562775, EPI_ISL_562776, EPI_ISL_562777, EPI_ISL_562778, EPI_ISL_562779, EPI_ISL_562780, EPI_ISL_562781, EPI_ISL_562782, EPI_ISL_562783, EPI_ISL_562784, EPI_ISL_562786, EPI_ISL_562787, EPI_ISL_562788, EPI_ISL_562789 | | | |
| see above | Microbiological Diagnostic Unit - Public Health Laboratory (MDU-PHL) | MDU-PHL | Seemann, T., Schultz M. B., Sait, M., Sherry, N. |
| EPI_ISL_562791 | Victorian Infectious Diseases Reference Laboratory (VIDRL) | VIDRL and MDU-PHL | Caly, L., Seemann, T., Sait, M., Schultz, M. B., Druce J., Sherry, N. |
| EPI_ISL_562793, EPI_ISL_562794, EPI_ISL_562796, EPI_ISL_562797, EPI_ISL_562798, EPI_ISL_562799, EPI_ISL_562800 | Microbiological Diagnostic Unit - Public Health Laboratory (MDU-PHL) | MDU-PHL | Seemann, T., Schultz M. B., Sait, M., Sherry, N. |
| EPI_ISL_562803, EPI_ISL_562804, EPI_ISL_562805, EPI_ISL_562806, EPI_ISL_562807, EPI_ISL_562808, EPI_ISL_562809 | Victorian Infectious Diseases Reference Laboratory (VIDRL) | VIDRL and MDU-PHL | Caly, L., Seemann, T., Sait, M., Schultz, M. B., Druce J., Sherry, N. |
| EPI_ISL_562810, EPI_ISL_562811, EPI_ISL_562812, EPI_ISL_562813, EPI_ISL_562814, EPI_ISL_562815, EPI_ISL_562816, EPI_ISL_562817, EPI_ISL_562818, EPI_ISL_562819, EPI_ISL_562820, EPI_ISL_562821, EPI_ISL_562822, EPI_ISL_562823, EPI_ISL_562824, EPI_ISL_562825, EPI_ISL_562826, EPI_ISL_562827, EPI_ISL_562828, EPI_ISL_562829, EPI_ISL_562830, EPI_ISL_562831, EPI_ISL_562832, EPI_ISL_562833, EPI_ISL_562834, EPI_ISL_562835, EPI_ISL_562836, EPI_ISL_562837 | | | |
| see above | Microbiological Diagnostic Unit - Public Health Laboratory | MDU-PHL | Seemann, T., Schultz M. B., Sait, M., Sherry, N. |
